# Health outcomes in children and adolescents with overweight or obesity exposed to physical activity interventions: an umbrella review covering over 1,200 trials

**DOI:** 10.1101/2025.01.21.25320866

**Authors:** Fernanda Dias Massierer, Cíntia Ehlers Botton, Jessica Pietra da Silva Carvalho, Gisele Cassão, Angélica Trevisan De Nardi, Jayne Feter, Andresa Conrado Ignacio, Rodrigo Leal-Menezes, Nórton Luís Oliveira, Lucineia Orsolin Pfeifer, Leandro dos Santos, Lucas Porto Santos, Larissa Xavier Neves da Silva, Luciana dos Passos e Silva, Frederico Moraes Schwingel, Carolina Weingärtner Welter, Daniel Umpierre

## Abstract

**Importance:** Childhood and adolescent obesity are linked to health risks. Standardizing outcome measurements in physical activity interventions for this group can increase evidence comparability and foster knowledge translation.

**Objective:** The objective of our study is to summarize individual health outcomes reported in systematic reviews and classify individual outcomes under an existing taxonomy of health domains for the development of a Core Outcome Set.

**Data Sources:** A comprehensive search was conducted across seven databases (PubMed, EMBASE, ERIC, SportDiscus, Cochrane Database, PROSPERO and Google) from inception to January 2023.

**Study Selection:** Eligible studies were systematic reviews with or without meta-analyses involving children and adolescents aged 4 to 19 years, with overweight or obesity, reporting on health outcomes from interventions including physical activity.

**Data extraction and synthesis:** Data extraction was carried out by independent reviewers in pairs. It included publication year, country, study characteristics, intervention details, participants demographics and health outcomes. Outcomes were classified into taxonomy-based domains.

**Main outcomes and measures:** This review identified key outcomes related to body composition, lipid profile, blood pressure, and physical functioning. In body composition, the most frequent outcomes were BMI, body weight, and body fat. For lipid profile, the common outcomes included HDL, total cholesterol, and LDL. Blood pressure outcomes encompassed systolic and diastolic blood pressure. Physical functioning was primarily assessed by time spent in physical activity.

**Results:** From the 138 included reviews, we cumulatively extracted 845 outcomes, with the identification of 169 unique outcomes distributed along 16 domains. The highest cumulative frequencies was in General (with 255 outcomes), Physical Functioning (128) and Blood and Lymphatic System (118) domains. The most frequently reported unique outcomes were body mass index (BMI) (52.2% of the reviews), followed by body weight, body fat, high-density lipoprotein (HDL), and systolic blood pressure.

**Conclusions and relevance:** This study highlights the focus on body composition in reviews of physical activity interventions for children and adolescents with overweight or obesity, while other outcomes showed inconsistencies. Establishing a Core Outcome Set (COS) is key to standardizing outcomes and improving the effectiveness of interventions for better pediatric health.

**Key Points:** *Question:* What are the main health outcomes for children and adolescents with overweight and obesity exposed to physical activity interventions?

*Findings:* In this umbrella review, including 138 studies, the main outcomes mentioned were related to body composition, lipid profile and blood pressure, covering over 370,000 trial participants.

*Meaning:* The outcomes found will serve for the development of a Core Outcome Set (COS), guiding new clinical trials, reducing inconsistencies and heterogeneity among possible future outcomes.

## Introduction

Obesity in children and adolescents represents a global health issue ^1,2^, increasing the risk of cardiometabolic ^3,4^, physical and psychological conditions ^5–9^. The World Health Organization (WHO) recommends to this population at least 60 minutes of moderate to vigorous physical activity daily, including muscle and bone strengthening activities at least three times a week ^10^. Conversely, a global survey indicates that 4 out of 5 students aged 11 to 17 do not meet this guideline ^11,12^. Numerous physical activity interventions have been assessed in clinical trials, reinforcing the importance of behavioral strategies to mitigate obesity’s impact ^13,14^.

The high heterogeneity of outcomes used in physical activity programs can hinder comparative analysis and evidence synthesis among the existing interventions. A review of 137 studies was unable to provide a comprehensive synthesis due to the high variability in outcome measures, which hindered the interpretation of the interventions’ impact on the physical fitness of students ^15^. A solution to minimize this inconsistency is through a Core Outcome Set (COS), which is a standardized group of outcomes for a given intervention and population, therefore increasing comparability across clinical trials. This process involves the engagement of relevant stakeholders and a comprehensive approach to selecting important outcomes ^16–18^. Such need is exemplified by the highlight of critical outcomes for studies in children and adolescents aged 5 to 17 by the WHO^10^, emphasizing the importance of a coordinated selection process for future trials.

Therefore, we aimed to summarize and classify outcomes reported in systematic reviews, seeking to inform the development of a COS for physical activity interventions for children and adolescents. Our objectives were 1) to describe individual health outcomes reported in reviews; and 2) to classify the individual outcomes under an existing taxonomy of health domains ^19^.

## Methods

This study is designed as an umbrella review encompassing systematic reviews with or without meta-analyses. The present work was registered in the International Prospective Register of Systematic Reviews (PROSPERO database) (CRD42019120334). The protocol, following the Preferred Reporting Items for Systematic Reviews and Meta-Analysis Protocols – PRISMA-P ^20^, materials, and data are available at the Open Science Framework (https://osf.io/7vaw5/). The study reporting follows the PRISMA Statement for Reporting Systematic Reviews and Meta-Analyses ^21^.

That were three protocol deviations: 1) expanding the age range by one year at the low and high limits to avoid excluding studies that encompass the population of interest, given the variability in school-age representation across the studies; 2) restricting the population to children and adolescents with overweight or obesity to generate more specific evidence; 3) the exclusion of cognitive outcomes due to the consensus among the reviewers of the update not considering these outcomes as health outcomes.

## Eligibility Criteria

Inclusion criteria were: 1) reports written in English, Portuguese or Spanish; 2) systematic reviews with or without meta-analysis of intervention studies (randomized or nonrandomized, controlled or non-controlled); 3) samples including children and adolescents with overweight or obesity (from 4 to 19 years old), healthy or at risk/diagnosis of cardiometabolic diseases; 4) included health lifestyle interventions with physical activity component (e.g., exercise, leisure-time physical activity, counseling, parent participation) with a duration of at least four weeks; 5) report any health outcome measured in children or adolescents.

## Information Sources

A search was conducted across five databases for indexed full-text publications (PubMed, EMBASE, ERIC, SportDiscus, Cochrane Database). Google Scholar and PROSPERO were databases used to retrieve non-published or non-retrieved literature (grey literature). The search period was from inception to January 2023, except for PROSPERO registration records, in which we carried out searches up to February 2024. Literature search strategies were developed using Medical Subject Headings (MeSH) and keywords. The full-search strategies for all databases are available in the Supporting Information (eTable 1).

## Study Selection

At first, the entire review process was conducted by three pairs of independent reviewers (CEB, LPS, ADN, LOP, LS, LXNS). The authors’ pairs screened the titles and abstracts yielded by the search based on the eligibility criteria. In the second stage, four pairs of independent researchers (FDM, LXNS, TSA, JF, ACI, LPS, CWW) conducted the study selection. A pilot screening of 100 articles was carried out to standardize the eligibility criteria in these two moments. After that, the reviewer pairs independently retrieved and assessed full texts of potentially eligible studies. The same reviewer pairs screened the full-text reports and made individual decisions on eligibility. Any discordance was solved by consensus and, in case of discordances, a third reviewer’s opinion was requested (DU). The articles assessed at the full text level are available in the Supporting Information (eTable 2).

## Definition of Outcomes and Domains

The definition of domain, outcome and outcome measure was based on Sinha et al. ^22^. The domain is a relatively broad aspect (e.g., cardiac) of the effect of a condition (e.g., overweight/obesity) on a population (e.g., children/adolescents), within which an improvement may occur in response to an intervention (e.g., physical activity). In general, these domains may not be directly measurable themselves, so outcomes are selected to assess change within the domains. The outcome is a measurable variable (e.g., systolic blood pressure) within a domain. In contrast, the outcome measure is a scale, scoring system, questionnaire, or other tools (e.g., office blood pressure measurement) used for measuring an outcome.

The domains were pre-defined according to a taxonomy developed for outcomes in medical research ^19^. The taxonomy comprises 38 outcomes domains, according to five core areas: mortality; physiological or clinical (e.g., endocrine, cardiac, psychiatric, musculoskeletal); life impact (e.g., physical functioning, social functioning, global quality of life); resource use (e.g., economic, societal/carer burden); adverse events.

## Data Extraction Process

Initially, three pairs of authors (CEB, NLO, ATDN, LOP, GC, LXNS) independently pilot-tested the extraction form for three included papers to ensure consistency in the interpretation of items and to generate internal definitions and improvements to the form components. Data extraction from the included studies was carried out on a standardized coded sheet. Subsequently, during the update phase, five pairs of independent reviewers (FDM, LXNS, ACI, JPSC, RLM, TSL, FMS, YFS, CWW, LPS) conducted a pilot with 10 papers to ensure standardization and consistency in the interpretation of items. This process aimed to enhance the quality and reliability of data extraction, contributing to a more robust and comparable analysis of the included studies.

In this context, a total of 24 items were extracted, with key elements including: 1) year and country of publication; 2) the number of studies and participants included; 3) characteristics of interventions; 4) intervention settings; 5) participants characteristics (such as age, sex, presence of overweight/obesity or other cardiometabolic diseases); 6) health outcomes considered in the reviews; and 7) measuring instruments.

The outcomes were extracted as indicated in the included reviews. Due to inconsistent or generic definitions of outcomes across several studies, the more specific description of the outcomes was also extracted, typically found in the methods and outcome measure sections. For instance, while authors identified the primary outcome as “adiposity”, the assessed outcome was specifically “abdominal fat” measured by “densitometry”. In such cases, both the broader outcome information, as described by the authors in the introduction and objectives (“adiposity”), and the more specific description in the methods (“abdominal fat”) were extracted. Both descriptions of outcomes are listed for each review in the Supporting Information (eTable 3).

To clarify the scope of this umbrella review, we focused on the outcomes of interest identified in the included reviews, rather than encompassing all outcomes investigated by the primary studies. For example, a systematic review synthesizing the effects of physical activity on blood pressure might have included primary studies evaluating diverse outcomes such as cognitive function, body composition or academic performance. However, if the review did not explicitly consider these additional outcomes, they were not taken into account in the present umbrella review.

## Data Synthesis and Analysis

The extracted data were analyzed through descriptive statistics, treated into two different levels. Initially, the computation involved the number of reviews investigating a unique outcome. Additionally, the total number of outcomes observed across reviews was calculated by summing the number of reviews for each unique outcome. Subsequently, outcomes were grouped into domains according to the taxonomy developed for outcomes in medical research ^19^. The outcomes were allocated to the domain of greatest affinity, according to the concept and examples described by the Core Outcome Measures in Effectiveness Trials – COMET initiative ^18^. The percentage frequency of unique outcomes or domains was relative to the total number of outcomes observed.

## Results

### Search Results

After excluding 3,446 duplicates, 19,233 articles were reviewed at the first stage of eligibility criteria assessment, with 18,263 being excluded after screening titles and abstracts (Figure 1). Among 970 full texts assessed (eTable 2), 138 were eligible and included in the qualitative analysis. This subset comprised 49 systematic reviews and 89 systematic reviews with meta-analysis, involving 371,553 participants in 1,219 clinical trials (eTable 4).

**Figure 1.**
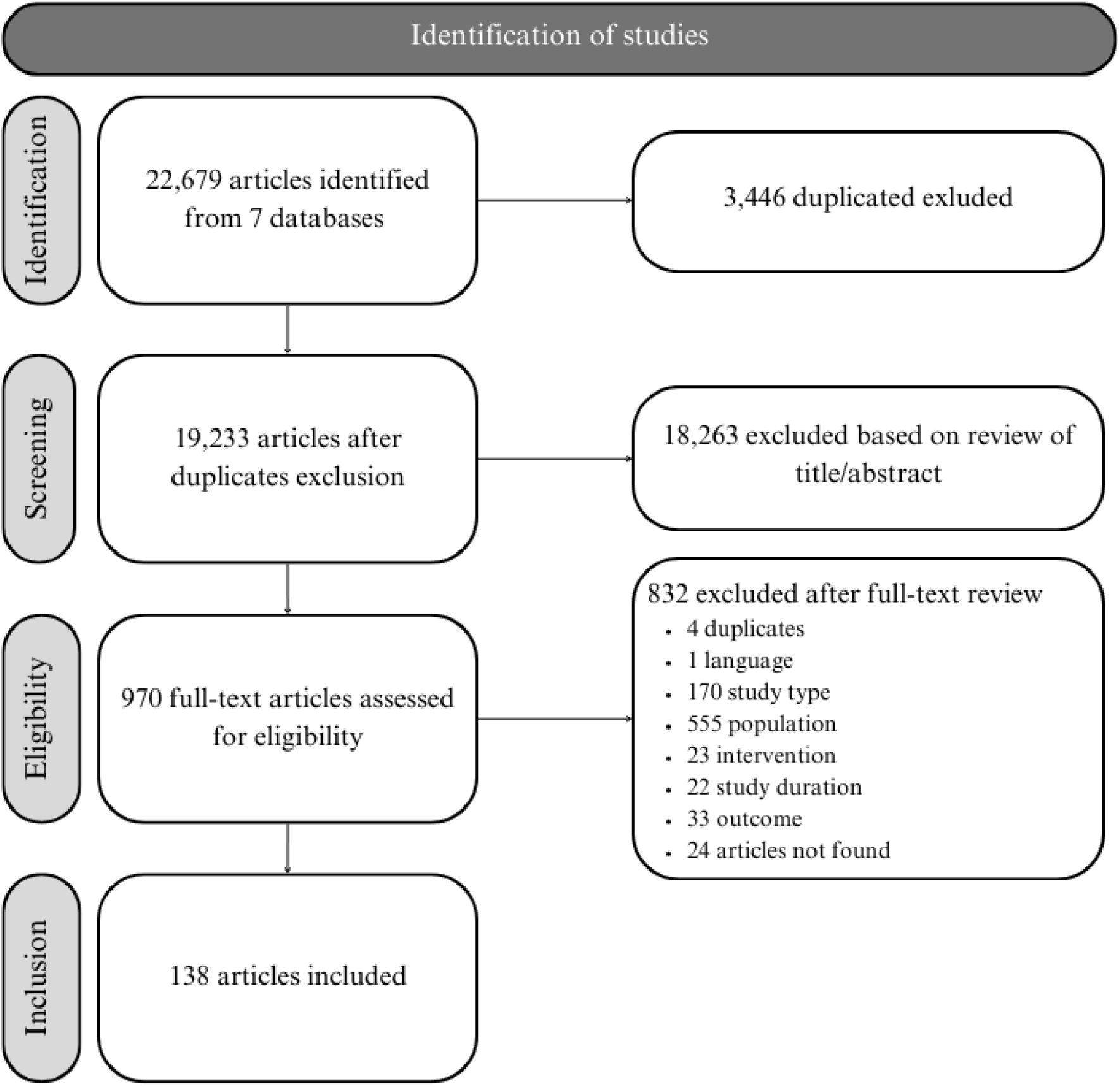
Identification and selection of articles.

## Study Characteristics

Participants were school-aged children and adolescents, aged four to 19 years old. Ten reviews included trials assessing only children, 17 focused solely on adolescents, 78 assessed both age groups, and 33 assessed children or adolescents with family involvement. All reviews summarized trials that included children or adolescents with overweight or obesity, whereas 48 (34.7%) reviews also included trials that recruited participants with normal weight. Moreover, all reviews summarized studies with participants of both sexes, with the exception of one review that considered only females. Trial interventions involved the delivery of physical activity, educational and behavioral approaches, or dietary approaches combined with physical activity. The interventions were carried out in schools, clinics, summer camps, communities, homes, hospitals, universities, primary and secondary health care. The characteristics of the reviews are available in the Supporting Information (eTable 5).

## Health Outcomes and Domains

From the 138 included reviews, we cumulatively extracted 845 outcomes. These outcomes often overlapped across the reviews and were subsequently grouped into 169 unique outcomes, which represent distinct conceptual categories. This classification was organized into 16 domains according to an established taxonomy (eTable 6).

The three most frequent domains were, respectively, ‘Endocrine’ (39 outcomes), ‘Physical Functioning’ (28 outcomes), and ‘Emotional Functioning/Wellbeing’ (18 outcomes). On the other hand, the less frequent domains were, respectively, ‘Perceived Health Status’, ‘Delivery of Care’ (each domain having 3 to 4 outcomes), and overall ‘Psychiatric’ and ‘Global Quality of Life’ (2 outcomes).

A total of 19 out of 169 outcomes were summarized in at least 10 reviews (Table 1). In this regard, body mass index (BMI) (n = 72; 52.2% of the reviews) was the outcome most largely used, followed by body weight (n = 39; 28.3%), body fat (n = 34; 24.6%) high-density lipoprotein (HDL cholesterol) (n = 31; 22.5%) and systolic blood pressure (n = 31; 22.5%). Although the ‘Endocrine’ domain showed the highest number of outcomes (39 outcomes), the ‘General’ domain included a variety of outcomes that together totalized a larger (n = 255) cumulative frequency.

**Table 1.** Frequencies of unique outcomes, domains, and subdomains (if any) for outcomes appearing in at least 10 systematic reviews.

| Outcomes | Frequency, N | Frequency, % | Domain (subdomain) |
| --- | --- | --- | --- |
| Body Mass Index (BMI) | 72 | 52.2 | General (body composition) |
| Body weight | 39 | 28.3 | General (body composition) |
| Body fat | 34 | 24.6 | General (body composition) |
| High-density lipoproteins (HDL) | 31 | 22.5 | Blood and Lymphatic System (lipid profile) |
| Systolic blood pressure (SBP) | 31 | 22.5 | Cardiac (blood pressure) |
| Body Fat-free mass | 30 | 21.7 | General (body composition) |
| Total Cholesterol | 28 | 20.3 | Blood and Lymphatic System (lipid profile) |
| Diastolic blood pressure (DBP) | 27 | 19.6 | Cardiac (blood pressure) |
| Low-density lipoproteins (LDL) | 26 | 18.8 | Blood and Lymphatic System (lipid profile) |
| Triglycerides | 25 | 18.1 | Blood and Lymphatic System (lipid profile) |
| Skinfold thickness | 17 | 12.3 | General (body composition) |
| Time spent in physical activity | 16 | 11.6 | Physical Functioning |
| Regional fat | 16 | 11.6 | General (body composition) |
| Waist Circumference | 14 | 10.1 | General (body composition) |
| Muscle strength | 13 | 9.4 | Physical Functioning |
| Physical activity level | 13 | 9.4 | Physical Functioning |
| Fasting blood glucose | 12 | 8.7 | Blood and Lymphatic System (lipid profile) |
| Sedentary behavior | 12 | 8.7 | Physical Functioning |

## Discussion

In this study, we systematically summarized outcomes assessed in systematic reviews of physical activity interventions in children and/or adolescents with overweight or obesity. Based on the 138 eligible reviews, we identified a total of 845 outcomes, comprising 169 unique ones, distributed into 16 domains. When analyzing the most relevant domain (General), outcomes related to body composition were cumulatively observed 255 times (30%) across the reviews. Meanwhile, the second most frequent domain (Physical Functioning) showed 128 outcomes (15%) related to physical function and fitness. Following this, the domains ‘Blood and Lymphatic System’, ‘Endocrine’ and ‘Cardiac’ were identified, representing 14%, 13.8% and 11% of the studies, respectively.

BMI is a simple, low-cost, and widely used anthropometric measure to detect overweight and obesity among several population subgroups. In our study, 1 out of 2 systematic reviews (n = 72; 52.2%) used BMI as an outcome in their syntheses. However, it is important to highlight its limitations to measure body fatness^23^. In children aged 6 to 14 years old, waist-to-height ratio could identify increased LDL cholesterol levels in children who would not be classified as overweight by the WHO BMI standard ^24,25^. Although both measures are useful in assessing overweight or obesity, we observed that only few reviews assessed waist-to-height ratio as an outcome, despite its diagnostic usefulness to characterize cardiometabolic risk factors in children ^26^.

We highlight that WHO guidelines on physical activity and sedentary behavior recently classified adiposity outcomes as critical for children and adolescents aged 5 to 17 ^10^. However, it is worth questioning whether body composition should be prioritized as the primary domain when selecting outcomes for physical activity programs for children with overweight or obesity. This consideration is particularly relevant given that the benefits of physical activity may extend beyond body composition, including improvements in cardiometabolic health, motor skills, and other physical and psychological domains. A broader perspective on outcome selection could provide a more comprehensive assessment of programs tailored for children or adolescents.

Although all interventions in this review included a physical activity component, only few studies reported outcomes related to either time spent in physical activity or sedentary behavior. Importantly, children and adolescents with obesity are 20 to 30% less physically active and present lower physical fitness ^27–29^ compared to their peers without obesity ^30–33^. Given the WHO’s emphasis on the role of physical activity in improving several health outcomes and reducing obesity risk ^34,35^, measures related to physical activity itself or sedentary behavior should be considered in future studies. Likewise, we emphasize that psychological and social outcomes were scarcely explored in the included systematic reviews, especially in the ‘Psychiatric’ and ‘Social Functioning’ domains. Therefore, outcomes related to depressive symptoms, anxiety, social acceptance, and body dissatisfaction could be further considered in clinical trials, since there is evidence showing their importance in children and adolescents with overweight or obesity ^8,9^.

Some studies have assessed whether obesity is associated with the cognitive function of children and adolescents ^7,36^. Lastly, current evidence has shown the importance of social support or parents’ engagement in physical activities for their children’s behavior, as well as parental involvement in weight control and engagement in lifestyle strategies ^37–39^. Although some studies have designed interventions with parental participation, only few assess outcomes related to parents’ or family aspects. Since, children and adolescents may rely on family support to engage and maintain health intervention, exploring parental and family outcomes, both related or not to physical activity, seems important in future research.

Building upon this need for standardized evaluations, domains are proposed by the COS developers, based on a summary of outcomes presented in the literature and consensus from stakeholders ^16^. Similar studies with the pediatric population have abandoned the use of taxonomy and classified their outcomes using their own system ^13,40^. To our knowledge, our study is a primary comprehensive synthesis of outcomes investigated in physical activity interventions for overweight/obese children and adolescents. The present review intends to inform a future COS, which requires other stages such as collecting outcomes from stakeholders, undertaking Delphi rounds, and convening a diverse panel to reach consensus in a COS ^41–43^. As outcomes may vary and be sensible to specific age ranges, it is reasonable that a COS development will need to consider children and adolescents separately. More importantly, the adoption of a COS would allow standardizing outcomes in future studies, providing a more consistent and comparable assessment of physical activity interventions for this population.

## Limitations

Some limitations should be considered in our study. Classifying outcomes into domains was challenging because the implemented taxonomy ^19^ was created for trials. Since we analyzed systematic reviews, which do not provide such detailed descriptions of outcomes, the classification was more imprecise than we anticipated. Similarly, it was created based on clinical conditions, disregarding outcomes that would be important for pediatric work, such as parental information. As a result, our description of outcomes is limited to broader terms. The wide age range considered in our meta-epidemiological assessment may require further data filtering if a future COS development occurs apart for children and adolescents.

## Conclusion

This umbrella review presents a high number of domains and outcomes used in studies with physical activity interventions for children and adolescents with overweight or obesity. While body composition outcomes were quite frequent, other outcomes related to physical activity, mental health, and social aspects were largely underexplored. This review informs the development of a future COS that may increase the standardization and comparability across trials of physical activity interventions for these populations.

## Data Availability

The data for this study are available on the Open Science Framework Platform (https://osf.io/7vaw5/)

## Author Contributions

Dr. Umpierre, Dr. Botton and Msc. Massierer had full access to all of the data in the study and take responsibility for the integrity of the data and the accuracy of the data analysis. *Concept and design:* Umpierre, Botton and Massierer.

*Study Selection and Data Extraction:* Massierer, Button, Carvalho, Cassão, De Nardi, Fetter, Ignacio, Leal-Menezes, Oliveira, Pfeifer, Santos, dos Santos, Silva, Passos, Schwingel, and Welter.

*Acquisition, analysis, or interpretation of data:* Umpierre, Massierer, Botton, Silva, and Oliveira.

*Drafting the manuscript:* Umpierre, Massierer, and Botton.

