## Supplementary Information for "Health outcomes in children and adolescents with overweight or obesity exposed to physical activity interventions: an umbrella review covering over 1,200 trials"

**eTable 1. Complete database search strategy.**

| Database | Search Strategy |
|---|---|
| Pubmed | #1 Child[Mesh] OR Child*[tiab] OR Youth[tiab] OR Adolescent[Mesh] OR Adolesc*[tiab] OR Pube*[tiab] OR Boys[tiab] OR Girls[tiab] OR School*[tiab] OR Pediatric*[tiab] OR Juvenile[tiab] OR Kid[tiab]<br><br>#2 Exercise Therapy[mesh] OR Exercise[mesh] OR Exercis*[tiab] OR Physical Activ*[tiab] OR Healthy Lifestyle[mesh] OR Health Behavior[mesh] OR Sports[mesh]<br><br>#3 MEDLINE[Title/Abstract] OR (systematic[Title/Abstract] AND review[Title/Abstract]) OR meta-analysis[Publication Type] OR (system*[Title/Abstract] AND review[Title/Abstract]) OR meta-an*[tiab]<br><br>#1 AND #2 AND #3 |
| Embase | #1 'child'/exp OR child*:ti,ab OR 'juvenile'/exp OR 'adolescent'/exp OR adolescen*:ti,ab OR puberty:ti,ab OR boy:ti,ab OR girl:ti,ab OR 'school'/exp OR pediatrics:ti,ab OR kid:ti,ab<br><br>#2 'exercise'/exp OR exercis*:ti,ab OR 'physical activity'/exp OR physical activit*:ti,ab OR 'healthy lifestyle'/exp OR healthy lifestyle:ti,ab OR 'health behavior'/exp OR health behavior:ti,ab OR 'sport'/exp OR sport:ti,ab<br><br>#3 'systematic review'/exp OR systematic review:ti,ab OR 'meta analysis'/exp OR meta anal*:ti,ab<br><br>#1 AND #2 AND #3 |
| Cochrane | #1 [Child] OR [Adolescent] OR [Puberty] OR [Schools] OR boys OR girls<br><br>#2 [Exercise] OR [Healthy Lifestyle] OR [Health Behavior] OR [Sports] OR Physical Activity<br><br>#3 [Meta-Analysis as Topic] OR Systematic Review<br><br>#1 AND #2 AND #3 |
| SPORTDiscus | #1 Children OR Youth OR Adolescent OR Puberty OR Boys OR Girls OR School OR Pediatrics OR Juvenile OR Kid<br><br>#2 Exercise Therapy OR Exercise OR Physical Activity OR Healthy Lifestyle OR Health Behavior OR Sports<br><br>#3 systematic review OR meta-analysis OR review<br><br>#1 AND #2 AND #3 |

| | |
|---|---|
| ERIC | #1 "Child" OR "Youth" OR "Adolescent" OR "Puberty" OR "Boys" OR "Girls" OR "School" OR "Pediatrics" OR "Juvenile" OR "Kid"<br><br>#2 "Exercise Therapy" OR "Exercise" OR "Physical Activity" OR "Healthy Lifestyle" OR "Health Behavior" OR "Sports"<br><br>#3 "systematic review" OR "meta-analysis"<br><br>#1 AND #2 AND #3 |
| PROSPERO | #1 Child [Mesh]/exp OR Adolescent [Mesh]/exp OR Pediatrics [Mesh]/exp OR Puberty:ti OR Youth:ti OR Boys:ti OR Girls:ti OR School:ti OR Juvenile:ti<br><br>#2 Exercise [Mesh]/exp OR Healthy Lifestyle [Mesh]/exp OR Health Behavior [Mesh]/exp OR Sport:ti OR Physical Activity:ti<br><br>#1 AND #2 |

### eTable 2. Articles assessed at the full text level.

| Title | Authors | Decision/Reason for exclusion |
|---|---|---|
| Diet, physical activity and behavioural interventions for the treatment of overweight or obese adolescents aged 12 to 17 years | Al-Khudairy et al., 2017 | INCLUDED |
| The Effect of Physical Activity Interventions on Children's Cognition and Metacognition: A Systematic Review and Meta-Analysis | Álvarez-Bueno et al., 2017 | INCLUDED |
| Weight loss interventions for overweight and obese adolescents: a systematic review | Boff et al,. 2017 | INCLUDED |
| Characteristics of physical activity interventions and effects on cardiorespiratory fitness in children aged 6-12 years-A systematic review | Braaksma et al., 2018 | INCLUDED |
| Diet and physical activity interventions to prevent or treat obesity in South Asian children and adults: a systematic review and meta-analysis | Brown et al., 2015 | INCLUDED |
| Efectos del entrenamiento de la aptitud muscular sobre la adiposidad corporal y el desempeño motriz en niños y jóvenes: un meta-análisis. / Effects of Resistance Training on body fat and motor performance skills in children and adolescents: A Meta-Analysis | Casas et al., 2018 | INCLUDED |
| Physical activity and cardiovascular risk factors in children: meta-analysis of randomized clinical trials | Cesa et al., 2014 | INCLUDED |
| The effect of resistance training interventions on weight status in youth: a meta-analysis | Collins et al., 2018 | INCLUDED |
| Serious games as an educational strategy to control childhood obesity: a systematic literature review1 | Dias et al., 2018 | INCLUDED |
| Exercise and Vascular Function in Child Obesity: A Meta-Analysis | Dias et al., 2015 | INCLUDED |
| School‑based physical activity programs for promoting physical activity and fitness in children and adolescents aged 6 to 18 | Dobbins et al., 2013 | INCLUDED |
| Effect of physical training on the blood pressure of adolescents with obesity | Farah et al., 2012 | INCLUDED |

| | | |
|---|---|---|
| Systematic review and meta-analysis of school-based obesity interventions in mainland China | Feng et al., 2017 | INCLUDED |
| Child and youth participatory interventions for addressing lifestyle-related childhood obesity: a systematic review | Frerichs et al., 2016 | INCLUDED |
| Exercise, adipokines and pediatric obesity: a meta-analysis of randomized controlled trials | García-Hermoso et al., 2017-a | INCLUDED |
| Is high-intensity interval training more effective on improving cardiometabolic risk and aerobic capacity than other forms of exercise in overweight and obese youth? A meta-analysis | García-Hermoso et al., 2016-a | INCLUDED |
| Effects of Exercise on Carotid Arterial Wall Thickness in Obese Pediatric Populations: A Meta-Analysis of Randomized Controlled Trials | García-Hermoso et al., 2017-b | INCLUDED |
| Concurrent aerobic plus resistance exercise versus aerobic exercise alone to improve health outcomes in paediatric obesity: a systematic review and meta-analysis | García-Hermoso et al., 2016-b | INCLUDED |
| Endocrinology and Adolescence: aerobic exercise reduces insulin resistance markers in obese youth: a | García-Hermoso et al., 2014 | INCLUDED |

| | | |
|---|---|---|
| meta-analysis of randomized controlled trials | | |
| Exercise-based interventions and C-reactive protein in overweight and obese youths: a meta-analysis of randomized controlled trials | García-Hermoso et al., 2016-c | INCLUDED |
| Effects of Aerobic Plus Resistance Exercise on Body Composition Related Variables in Pediatric Obesity: A Systematic Review and Meta-Analysis of Randomized Controlled Trials | García-Hermoso et al., 2015 | INCLUDED |
| A systematic review of the effectiveness of school-based obesity prevention programmes for First Nations, Inuit and Metis youth in Canada | Godin et al., 2015 | INCLUDED |
| The Effects of Exercise on Abdominal Fat and Liver Enzymes in Pediatric Obesity: A Systematic Review and Meta-Analysis | González-Ruiz et al., 2017 | INCLUDED |
| School-Based Interventions on Childhood Obesity. A Meta-Analysis | Gonzalez-Suarez et al., 2009 | INCLUDED |
| School-based physical activity and nutritional education interventions on body mass index: a meta-analysis of randomised community trials - project PANE | Guerra et al., 2014 | INCLUDED |

| | | |
|---|---|---|
| The effect of school-based physical activity interventions on body mass index: a meta-analysis of randomized trials | Guerra et al., 2013 | INCLUDED |
| Computer- and web-based interventions to increase preadolescent and adolescent physical activity: a systematic review | Hamel et al., 2011 | INCLUDED |
| Effect of school-based physical activity interventions on body mass index in children: a meta-analysis | Harris et al., 2009 | INCLUDED |
| Impact of dietary and exercise interventions on weight change and metabolic outcomes in obese children and adolescents: a systematic review and meta-analysis of randomized trials | Ho et al., 2013 | INCLUDED |
| Non-curricular approaches for increasing physical activity in youth: a review | Jago et al., 2004 | INCLUDED |
| Parent-only vs. parent-child (family-focused) approaches for weight loss in obese and overweight children: A systematic review and meta-analysis | Jull and Chen., 2013 | INCLUDED |
| Effects of aerobic exercise on non-high-density lipoprotein cholesterol in children and | Kelley and Kelley., 2008 | INCLUDED |

| | | |
|---|---|---|
| adolescents: a meta-analysis of randomized controlled trials | | |
| Sport-based physical activity intervention on body weight in children and adolescents: a meta-analysis | Kim et al., 2017 | INCLUDED |
| School-based obesity prevention programs: an evidence-based review | Kropski et al., 2008 | INCLUDED |
| The effects of aerobic physical activity on adiposity in school-aged children and youth: a systematic review of randomized controlled trials | Laframboise et al., 2011 | INCLUDED |
| Effectiveness of interventions to promote healthy weight in general populations of children and adults: a meta-analysis | Luckner et al., 2012 | INCLUDED |
| Family involvement in weight control, weight maintenance and weight-loss interventions: A systematic review of randomised trials | Mclean et al., 2003 | INCLUDED |
| Effectiveness of after-school interventions at increasing moderate-to-vigorous physical activity levels in 5- to 18-year olds: a systematic review and meta-analysis | Mears and Jago., 2016 | INCLUDED |
| [Intervention programs to promote physical activity in school children: systematic review] | Medina-Blanco et al., 2011 | INCLUDED |

| | | |
|---|---|---|
| The impact of long-term school-based physical activity interventions on body mass index of primary school children - a meta-analysis of randomized controlled trials | Mei et al., 2016 | INCLUDED |
| Is Physical Fitness Associated with Health in Overweight and Obese Youth? A Systematic Review | Millard-Staffor et al., 2013 | INCLUDED |
| Interventions for the treatment of obesity among children and adolescents in Latin America: a systematic review | Nagle et al., 2013 | INCLUDED |
| Effects of group sports on health-related physical fitness of overweight youth: A systematic review and meta-analysis | Oliveira et al., 2017 | INCLUDED |
| The effects of school-based lifestyle interventions on body mass index and blood pressure: a multivariate multilevel meta-analysis of randomized controlled trials | Oosterhoff et al., 2016 | INCLUDED |
| Metabolic effects of exercise on childhood obesity: a current view | Paes et al., 2015 | INCLUDED |
| Influence of Intensity of Physical Activity on Adiposity and Cardiorespiratory Fitness in 5-18 Year Olds | Parikh and Stratton et al., 2011 | INCLUDED |
| Efectos de los programas de intervención enfocados al tratamiento del sobrepeso/obesidad infantil y adolescenteEffects of intervention programs focused on the treatment of overweight/obese children and adolescents | Silva et al., 2014 | INCLUDED |
| Systematic review of physical activity and exercise interventions on body mass indices, subsequent physical activity and psychological symptoms in overweight and obese adolescents | Ruotsalainen et al., 2015 | INCLUDED |
| Improvement of aerobic fitness in obese children: a meta-analysis | Saavedra et al., 2011 | INCLUDED |
| Educational interventions in childhood obesity: a systematic review with meta-analysis of randomized clinical trials | Sbruzzi et al., 2013 | INCLUDED |
| What is the effect of resistance training on the strength, body composition and psychosocial status of overweight and obese children and adolescents? A Systematic review and meta-analysis | Schranz et al., 2013 | INCLUDED |
| A systematic review and meta-analysis of energy and macronutrient intake responses to physical activity | Schwartz et al., 2017 | INCLUDED |

| | | |
|---|---|---|
| interventions in children and adolescents with obesity | | |
| The Effectiveness of Interventions on Sustained Childhood Physical Activity: A Systematic Review and Meta-Analysis of Controlled Studies | Sims et al., 2015 | INCLUDED |
| Effects of Physical Exercise on Adiponectin, Leptin, and Inflammatory Markers in Childhood Obesity: Systematic Review and Meta-Analysis | Sirico et al., 2018 | INCLUDED |
| Effective Weight Loss for Children: A Meta-analysis of Intervention Studies 2002-2015 | Snethen et al., 2016 | INCLUDED |
| Treatment of Childhood Obesity: A Systematic Review | Staniford et al., 2011 | INCLUDED |
| Efficacy of Exercise Intervention for Weight Loss in Overweight and Obese Adolescents: Meta-Analysis and Implications | Stoner et al., 2016 | INCLUDED |
| Family-based childhood obesity interventions in the UK: a systematic review of published studies | Upton et al., 2014 | INCLUDED |
| The impact of school-based prevention of overweight on psychosocial well-being of children: Obesity Prevention | van Wijnen et al., 2009 | INCLUDED |

| | | |
|---|---|---|
| Physical activity in overweight and obese adolescents: systematic review of the effects on physical fitness components and cardiovascular risk factors | Vasconcellos et al., 2014 | INCLUDED |
| Effectiveness of school-based physical activity and nutrition interventions with direct parental involvement on children's BMI and energy balance-related behaviors - A systematic review | Verjans-Janssen et al., 2018 | INCLUDED |
| The Effect of Diet or Exercise on Visceral Adipose Tissue in Overweight Youth | Vissers et al., 2016 | INCLUDED |
| Effect of Physical Activity Interventions for Girls on Objectively Measured Outcomes: A Systematic Review of Randomized Controlled Trials | Voskuil et al., 2017 | INCLUDED |
| Effectiveness of weight management interventions in children: A targeted systematic review for the USPSTF | Whitlock et al., 2010 | INCLUDED |
| A Meta-Analysis of the Effects of Different Training Modalities on the Inflammatory Response in Adolescents with Obesity | Zhao et at., 2022-b | INCLUDED |
| Association between Exercise-Induced Changes in Cardiorespiratory Fitness and Adiposity among Overweight and Obese Youth: A | Garcia-Hermoso et al., 2020-b | INCLUDED |

| | | |
|---|---|---|
| Meta-Analysis and Meta-Regression Analysis | | |
| Association of Physical Education With Improvement of Health-Related Physical Fitness Outcomes and Fundamental Motor Skills Among Youths A Systematic Review and Meta-analysis | Garcia-Hermoso et al., 2020-a | INCLUDED |
| Beyond bariatric surgery and weight loss medicaments. A systematic review of the current practice in obesity rehabilitative inpatient programs in adults and pediatrics | Spadaccini et al., 2022 | INCLUDED |
| Characteristics of Multicomponent Interventions to Treat Childhood Overweight and Obesity in Extremely Cold Climates: A Systematic Review of a Randomized Controlled Trial | Albornoz-Guerrero et al., 2021 | INCLUDED |
| Does Physical Activity-Based Intervention Improve Systemic Proinflammatory Cytokine Levels in Overweight or Obese Children and Adolescents? Insights from a Meta-Analysis of Randomized Control Trials | Han et al., 2019 | INCLUDED |
| Effects of High-Intensity Interval Training and Moderate-Intensity Continuous Training on Cardiometabolic Risk Factors in Overweight and Obesity Children and | Cao et al., 2021 | INCLUDED |

| | | |
|---|---|---|
| Adolescents: A Meta-Analysis of Randomized Controlled Trials | | |
| Effect of Active Video Games on Healthy Children’s Fundamental Motor Skills and Physical Fitness: A Systematic Review | Liu, W et al., 2020 | INCLUDED |
| Effect of High-Intensity Interval Training versus Moderate-Intensity Continuous Training on Cardiorespiratory Fitness in Children and Adolescents: A Meta-Analysis | Cao et al., 2019 | INCLUDED |
| Effectiveness and process evaluation in obesity and type 2 diabetes prevention programs in children: a systematic review and meta-analysis | Seral-Cortes et al., 2021 | INCLUDED |
| Effectiveness of family-based eHealth interventions in cardiovascular disease risk reduction: A systematic review | Kemp et al., 2021 | INCLUDED |
| Effectiveness of interventions aiming at reducing sedentary behaviour in a non-surgical population with overweight or obesity: A systematic review and meta-analysis | Zabatiero et al., 2018 | INCLUDED |
| Effectiveness of lifestyle interventions for treatment of overweight/obesity among children in China: A systematic review and meta-analysis | Li et al., 2022 | INCLUDED |

| | | |
|---|---|---|
| EFFECTIVENESS OF OBESITY INTERVENTION PROGRAMS BASED ON GUIDELINES FOR ADOLESCENT STUDENTS: SYSTEMATIC REVIEW | Wolfa et al., 2019 | INCLUDED |
| Effectiveness of obesity interventions among South Korean children and adolescents and importance of the type of intervention component: a meta-analysis | Choe et al., 2022 | INCLUDED |
| Effects of aerobic exercise and resistance exercise on physical indexes and cardiovascular risk factors in obese and overweight school-age children: A systematic review and meta-analysis | Chen et al., 2021 | INCLUDED |
| Effects of different exercise types on visceral fat in young individuals with obesity aged 6–24years old: A systematic review and meta-analysis | Wang et al., 2022 | INCLUDED |
| Effects of exercise combined with diet intervention on body composition and serum biochemical markers in adolescents with obesity: a systematic review and meta-analysis | Zhao et al., 2022-a | INCLUDED |
| Effects of Exercise in Improving Cardiometabolic Risk Factors in Overweight Children: A Systematic Review and Meta-Analysis | Busnatu et al., 2022 | INCLUDED |
| Effects of Exercise Intervention on Health‑Related Physical Fitness and Blood Pressure in Preschool Children: A Systematic Review and Meta‑Analysis of Randomized Controlled Trials | García-Hermoso et al., 2020-c | INCLUDED |
| Effects of Exercise Intervention on Visceral Fat in Obese Children and Adolescents: Meta-analysis | Jung et al., 2018 | INCLUDED |
| Effects of physical training on anthropometrics, physical and physiological capacities in individuals with obesity: A systematic review | Zouhal et al., 2020 | INCLUDED |
| Effects of Physical Training on Heart Rate Variability in Children and Adolescents with Chronic Diseases: A Systematic Review and Meta-analysis | Estéves-González et al., 2022 | INCLUDED |
| Effects of Strength Training on Body Fat in Children and Adolescents with Overweight and Obesity: A Systematic Review with Meta-Analysis | Méndez-Hernández et al., 2022 | INCLUDED |
| Evaluation of Physical Activity and Lifestyle Interventions Focused on School Children with Obesity Using Accelerometry: A Systematic Review and Meta-Analysis | Jurado-Castro et al., 2020 | INCLUDED |

| | | |
|---|---|---|
| Exercise and adiposity in overweight and obese children and adolescents: a systematic review with network meta-analysis of randomised trials | Kelley et al., 2019 | INCLUDED |
| Exergames in Childhood Obesity Treatment: A Systematic Review | Valeriani et al., 2021 | INCLUDED |
| High Intensity Interval Training (HIIT) Improves Cardiorespiratory Fitness (CRF) in Healthy, Overweight and Obese Adolescents: A Systematic Review and Meta-Analysis of Controlled Studies | Martin-Smith et al., 2020 | INCLUDED |
| High-Intensity Interval Training and Cardiometabolic Risk Factors in Children: A Meta-analysis | Solera-Martinez et al., 2021 | INCLUDED |
| High-intensity interval training in overweight and obese children and adolescents: systematic review and meta-analysis | Thivel et al., 2018 | INCLUDED |
| Impact of Lifestyle Intervention Programs for Children and Adolescents with Overweight or Obesity on Body Weight and Selected Cardiometabolic Factors—A Systematic Review | Bondyra-Wisniewska et al., 2021 | INCLUDED |

| | | |
|---|---|---|
| Influence of Chronic Exposure to Exercise on Heart Rate Variability in Children and Adolescents Affected by Obesity: A Systematic Review and Meta-Analysis | Dias et al., 2021 | INCLUDED |
| Influences of exercise interventions on overweight and obesity in children and adolescents | Lee et al., 2020 | INCLUDED |
| Interventions to prevent or treat childhood obesity in Māori & Pacific Islanders: a systematic review | Littlewood et al., 2020 | INCLUDED |
| Mobile phone interventions to improve adolescents' physical health: A systematic review and meta‑analysis | Shin et al., 2019 | INCLUDED |
| Exercise and Cardiovascular Disease Risk Factors in Children and Adolescents With Obesity: A Systematic Review With Meta-Analysis of Randomized Controlled Trials | Kelley et al., 2021 | INCLUDED |
| Physiological Responses to Combat Sports in Metabolic Diseases: A Systematic Review | Eckstein et al., 2022 | INCLUDED |
| Recreational Soccer Training Effects on Pediatric Populations Physical Fitness and Health: A Systematic Review | Clemente et al., 2022 | INCLUDED |
| Resistance Training and Insulin Sensitivity in Youth: A Meta-analysis | Burns et al., 2019 | INCLUDED |

| | | |
|---|---|---|
| School-Based Exercise Programs for Promoting Cardiorespiratory Fitness in Overweight and Obese Children Aged 6 to 10 | Mijalkovic et al., 2022 | INCLUDED |
| School‑Based Intervention Programs for Preventing Obesity and Promoting Physical Activity and Fitness: A Systematic Review | Yuksel et al., 2020 | INCLUDED |
| Slow and Steady, or Hard and Fast? A Systematic Review and Meta-Analysis of Studies Comparing Body Composition Changes between Interval Training and Moderate Intensity Continuous Training | Steele et al., 2021 | INCLUDED |
| The Benefts of Resistance Training in Obese Adolescents: A Systematic Review and Meta-analysis | Ribeiro et al., 2022 | INCLUDED |
| The Psychological Effects of Exergames for Children and Adolescents with Obesity: A Systematic Review and Meta-Analysis | Andrade et al., 2019 | INCLUDED |
| The Effects of Different Exercise Modalities in the Treatment of Cardiometabolic Risk Factors in Obese Adolescents with Sedentary Behavior—A Systematic Review and Meta-Analysis of Randomized Controlled Trials | Li and Chen et al., 2021 | INCLUDED |

| | | |
|---|---|---|
| What change in body mass index is associated with improvement in percentage body fat in childhood obesity? A meta-regression | Birch et al., 2019 | INCLUDED |
| The Effect of Resistance Training Interventions on 'The Self' in Youth: a Systematic Review and Meta-analysis | Collins et al., 2019 | INCLUDED |
| The effectiveness of e-health interventions for the treatment of overweight or obesity in children and adolescents: systematic review and meta-analysis | Azevedo et al., 2023 | INCLUDED |
| The Effects of Football Practice on Nutritional Status and Body Composition in Children: A Systematic Review and Meta-Analysis | Hernandez-Martin et al., 2021 | INCLUDED |
| High-Intensity Interval Training Improves physical morphology, Cardiopulmonary Fitness and Metabolic Risk Indicators of Cardiovascular Disease in Children and Adolescents: A Systematic Review and Meta-Analysis | Menjie et al., 2022 | INCLUDED |
| Effect of Exercise on Concentration of High-Density Lipoprotein in Youth: A Systematic Review and Meta-Analysis. | Lopes et al., 2019 | INCLUDED |

| | | |
|---|---|---|
| Effects of supervised exercise training on lipid profile of children and adolescents: Systematic review, meta-analysis and meta-regression. | Costa et al., 2020 | INCLUDED |
| The Morphofunctional Adaptations of Resistance Exercise on Reducing Cardiovascular Risk Factors in Adolescents: A Systematic Review (2012 to 2018). | da Rosa-Santos et al., 2019 | INCLUDED |
| School-Based High-Intensity Interval Training Programs for Promoting Physical Activity and Fitness in Adolescents: A Systematic Review. | Bento et al., 2022 | INCLUDED |
| Feasibility of incorporating high-intensity interval training into physical education programs to improve body composition and cardiorespiratory capacity of overweight and obese children: A systematic review. | Delgado-Floody et al., 2019 | INCLUDED |
| Interventions for preventing obesity in children (Review) | Brown et al., 2019 | INCLUDED |
| School-Based High-Intensity Interval Training Programs for Promoting Physical Activity and Fitness in Adolescents: A Systematic Review | Bento et al., 2021 | INCLUDED |

| | | |
|---|---|---|
| Systematic Review: Frameworks Used in School-Based Interventions, the Impact on Hispanic Children's Obesity-Related Outcomes | Andrade et al., 2018 | INCLUDED |
| Interventions to Treat Obesity in Mexican Children and Adolescents: Systematic Review and Meta-Analysis. | Aceves-Martins et al., 2022 | INCLUDED |
| Physical activity interventions for the mental health of children: A systematic review | Hale et al., 2023 | INCLUDED |
| Physical Activity Interventions to Alleviate Depressive Symptoms in Children and | Recchia et al., 2023 | INCLUDED |
| Effect of exercise training on arterial stiffness in obese and overweight children: a meta-analysis. | Cheng et al., 2022 | INCLUDED |
| The effect and safety of high-intensity interval training in the treatment of adolescent obesity: a meta-analysis | Zhu et al., 2021 | INCLUDED |
| Football as a Health Promotion Strategy | Eberl et al., 2019 | INCLUDED |
| Effects and dose-response relationships of exercise intervention on weight loss in overweight and obese children: a meta-regression and system review | Xu et al., 2022 | INCLUDED |

| | | |
|---|---|---|
| The Effects of Park-Based Interventions on Health-Related Outcomes Among Youth: | Deshira et al., 2022 | INCLUDED |
| Effects of Physical Exercise on Cardiometabolic Biomarkers and Inflammatory | Hejazi et al., 2022 | INCLUDED |
| Diet, physical activity, and behavioural interventions for the treatment of overweight or obesity in preschool children up to the age of 6 years | Colquitt et al., 2016 | INCLUDED |
| Diet, physical activity and behavioural interventions for the treatment of overweight or obese children from the age of 6 to 11 years | Mead et al., 2017 | INCLUDED |
| RESPONSES OF PLASMA ADIPOKINES TO HIGH INTENSITY INTERVAL TRAINING: SYSTEMATIC REVIEW. / RESPUESTAS DE LAS ADIPOCINAS PLASMÁTICAS AL ENTRENAMIENTO POR INTERVALOS DE ALTA INTENSIDAD: REVISIÓN SISTEMÁTICA | Da Silva et al., 2020 | INCLUDED |
| The effects of active video games on health-related physical fitness and motor competence in overweight or obese youth a systematic review and meta analysis | Comeras-Chueca et al., 2021 | INCLUDED |

| | | |
|---|---|---|
| Comparative Effectiveness of High-Intensity Interval Training and Moderate-Intensity Continuous Training for Cardiometabolic Risk Factors and Cardiorespiratory Fitness in Childhood Obesity: A Meta-Analysis of Randomized Controlled Trials | Liu et al., 2020 | INCLUDED |
| Does process evaluation enhance school-based High-Intensity Interval Training (HIIT) Interventions? A systematic review and meta-analysis for Randomized Controlled Trials | Liu et al., 2024 | INCLUDED |
| Effects of dance on the physical activity level of children and adolescents - a systematic review | Dos Santos et al., 2021 | INCLUDED |
| Diet, physical activity and behavioural interventions for the treatment of overweight or obese adolescents aged 12 to 17 years | Lena Al‑Khudairy, Emma Loveman,Jill L Colquitt, Emma Mead, Rebecca E Johnson, Hannah Fraser, Joan Olajide, Marie Murphy, Rochelle Marian Velho, Claire O'Malley, Liane B Azevedo, Louisa J Ells, Maria‑Inti Metzendorf, Karen Rees | Duplicate |
| Efectos del entrenamiento de la aptitud muscular sobre la adiposidad corporal y el desempeño motriz en niños y jóvenes: un meta-análisis. / Effects of Resistance Training | Casas, Adrián; Naclerio, Fernando; Calvo, Xurxo Dopico; García, Christian | Duplicate |

| | | |
|---|---|---|
| on body fat and motor performance skills in children and adolescents: A Meta-Analysis. | | |
| Effects of physical activity programs on body composition, physical fitness and cardiometabolic parameters in pre-school children: a meta-analysis of randomized controlled trials | Antonio García-Hermoso, Alicia M. Alonso-Martinez, Robinson Ramírez-Vélez, Mikel Izquierdo | Duplicate |
| Family-based, eHealth interventions to reduce cardiovascular risk: a systematic review | Bridie J. Kemp, David R. Thompson, Chris J. Watson, Karen McGuigan, Jayne V. Woodside, Chantal F. Ski | Duplicate |
| What impact does physical activity have on obesity in 3- to 6-year-olds? | Ducret, S. and C. Feller | Ineligible language |
| After-school interventions to increase physical activity among youth | Pate, R. R. | Ineligible study type |
| Approaches to Promoting More Widespread Participation in Physical Activity | Owen, N. and T. Dwyer | Ineligible study type |
| Assessing and Increasing Physical Activity | Van Camp, C. M. and L. B. Hayes | Ineligible study type |
| Body composition changes after weight-loss interventions for overweight and obesity | Santarpia, L., F. Contaldo and F. Pasanisi | Ineligible study type |
| Childhood obesity: prevention and strategies of intervention. A systematic review of | Verrotti, A., L. Penta, L. Zenzeri, S. Agostinelli and P. De Feo | Ineligible study type |

| | | |
|---|---|---|
| school-based interventions in primary schools | | |
| Culturally Competent Interventions to Address Obesity Among African American and Latino Children and Youth | Suarez-Balcazar, Y., J. Friesema and V. Lukyanova | Ineligible study type |
| Effects of nutrition, exercise and lifestyle interventions on ghrelin in obese children and adolescents | Poggiogalle, E., G. Polidoro, M. Arca, A. Lenzi and L. M. Donini | Ineligible study type |
| Efficacy of interventions that use apps to improve diet, physical activity and sedentary behaviour: a systematic review | Schoeppe, S., S. Alley, W. Van Lippevelde, N. A. Bray, S. L. Williams, M. J. Duncan and C. Vandelanotte | Ineligible study type |
| Health Literacy in Adolescents and Young Adults: An Updated Review | Sansom-Daly, U. M., M. Lin, E. G. Robertson, C. E. Wakefield, B. C. McGill, A. Girgis and R. J. Cohn | Ineligible study type |
| Physical activity interventions in the prevention and treatment of paediatric obesity: systematic review and critical appraisal | Reilly, J. J. and Z. C. McDowell | Ineligible study type |
| School-based obesity interventions: a literature review | Shaya, F. T., D. Flores, C. M. Gbarayor and J. Wang | Ineligible study type |
| Systematic review and meta-analysis of the association between childhood overweight and obesity and primary school diet and physical activity policies | Williams, A. J., W. E. Henley, C. A. Williams, A. J. Hurst, S. Logan and K. M. Wyatt | Ineligible study type |

| | | |
|---|---|---|
| Systematic review of effectiveness of universal self-regulation-based interventions and their effects on distal health and social outcomes in children and adolescents: Review protocol | Pandey, A., D. Hale, A. L. Goddings, S. J. Blakemore and R. Viner | Ineligible study type |
| Systematic review of the effectiveness of interventions used in the management of obesity | O'Meara, S., A. M. Glenny, T. Sheldon, A. Melville and C. Wilson | Ineligible study type |
| What are the health benefits of active travel? A systematic review of trials and cohort studies | Saunders, L. E., J. M. Green, M. P. Petticrew, R. Steinbach and H. Roberts | Ineligible study type |
| 'Measuring' Physical Literacy and Related Constructs: A Systematic Review of Empirical Findings | Edwards LC, Bryant AS, Keegan RJ, Morgan K, Cooper SM, Jones AM. | Ineligible study type |
| Effectiveness of educational interventions conducted in latin america for the prevention of overweight and obesity in scholar children from 6-17 years old; a systematic review | Mancipe Navarrete, J. A., S. S. Garcia Villamil, J. E. Correa Bautista, J. F. Meneses-Echavez, E. Gonzalez-Jimenez and J. Schmidt Rio-Valle | Ineligible study type |
| A comparison of the associations between bone health and three diferent intensities of accelerometer‑derived habitual physical activity in children and adolescents: a systematic review | Gemma Brailey, Brad Metcalf,Rebecca Lear, Lisa Price Sean Cumming, Victoria Stiles. | Ineligible study type |

| | | |
|---|---|---|
| A meta-analysis of school-based obesity prevention programs demonstrates limited efficacy of decreasing childhood obesity | Hung, L. S., D. K. Tidwell, M. E. Hall, M. L. Lee, C. A. Briley and B. P. Hunt | Ineligible study type |
| A review of electronic interventions for prevention and treatment of overweight and obesity in young people | Nguyen, B., K. P. Kornman and L. A. Baur | Ineligible study type |
| A Review of Systematic Reviews Targeting the Prevention and Treatment of | Lisa K. Militello, Ph.D., M.P.H.a, | Ineligible study type |
| A systematic review and meta-analysis of school-based interventions with health education to reduce body mass index in adolescents aged 10 to 19 years | Chandni Maria Jacob, , Polly Louise Hardy-Johnson, Hazel M. Inskip, Taylor Morris, Camille M. Parsons, Millie Barrett, Mark Hanson, Kathryn Woods-Townsend and Janis Baird. | Ineligible study type |
| A SYSTEMATIC REVIEW AND META-ANALYSIS OF THE EFFECTS OF IMPACT EXERCISE ON BONE STRUCTURE ACROSS THE LIFESPAN | C.-A. Ng, A. Gandham, J. Mesinovic, P. Ebeling, D. Scott. | Ineligible study type |
| A systematic review investigating the effects of implementing game-based approaches in school-based Physical Education and Sport among Primary school children | N. Eather, P. Morgan, J. Smith, J. Yan | Ineligible study type |
| A Systematic Review of Children's Physical Activity Patterns: Concept, Operational | Thayse Natacha Gomes, Peter T. Katzmarzyk, Sara Pereira, | Ineligible study type |

| | | |
|---|---|---|
| Definitions, Instruments, Statistical Analyses, and Health Implications | Mabliny Thuany, Martyn Standage and José Maia | |
| A systematic review of interventions for promoting active transportation to school | Chillon, P., K. R. Evenson, A. Vaughn and D. S. Ward | Ineligible study type |
| A Systematic Review of Interventions to Enhance Healthy Lifestyle Behaviors in Adolescents Delivered via Mobile Phone Text Messaging | Lois J. Loescher, Stephen A. Rains, Sandra S. Kramer, Chelsie Akers,Renee Moussa | Ineligible study type |
| A systematic review of lifestyle patterns and their association with adiposity in children aged 5–12 years | Ninoshka J. D'Souza, Konsita Kuswara, Miaobing Zheng, Rebecca Leech, Katherine L. Downing, Sandrine Lioret, Karen J. Campbell, Kylie D. Hesketh. | Ineligible study type |
| Access to bike lanes and childhood obesity: A systematic review and meta-analysis | Xiongfeng Pan, Li Zhao, Jiayou Luo, Yinhao Li, Lin Zhang, Tong Wu, Melody Smith, Shaoqing Dai, Peng Jia. | Ineligible study type |
| Active Commuting and Physical Fitness: A Systematic Review | Duarte Henriques-Neto, Miguel Peralta,Susana Garradas, Andreia Pelegrini, André Araújo Pinto, Pedro António Sánchez-Miguel and Adilson Marques | Ineligible study type |
| Active school transport, physical activity levels and body weight of children and youth: a systematic review | Faulkner, G. E., R. N. Buliung, P. K. Flora and C. Fusco | Ineligible study type |
| Active video game on children and adolescents' physical activity and weight | Robin Sze-Tak Ho \| Eddie Ka-Yui Chan \| Kenneth Kang-Yue Liu \| | Ineligible study type |

| | | |
|---|---|---|
| An investigation of the relationship between new fasting hormone asprosin, obesity and acute-chronic exercise: current systematic review. | Halil İbrahim Ceylan & Özcan Saygın | Ineligible study type |
| An online family-based self-monitoring and goal-setting intervention to improve children's physical activity: the FRESH feasibility trial and three-arm pilot RCT | Esther MF van Sluijs, Helen E Brown, Emma Coombes, Claire Hughes, Andrew P Jones, Katie L Morton and Justin M Guagliano | Ineligible study type |
| ANÁLISIS DE LAS CARACTERÍSTICAS Y LOS LOGROS FÍSICO-MOTRICES DE LAS CLASES BASADAS EN ACTIVIDAD FÍSICA: UNA REVISIÓN SISTEMÁTICA. / ANALYSIS OF THE CHARACTERISTICS AND PHYSICAL-MOTOR ACHIEVEMENTS OF THE CLASSES BASED ON PHYSICAL ACTIVITY: A SYSTEMATIC REVIEW. | Zapatero-Ayuso, Jorge Agustín | Ineligible study type |
| Are field-based exergames useful in preventing childhood obesity? A systematic review | Gao, Z. and S. Chen | Ineligible study type |
| Are There Inter-Individual Differences in Fat Mass and Percent Body Fat as a Result of Aerobic Exercise Training in Overweight and Obese Children and Adolescents? A Meta-Analytic Perspective | George A. Kelley, DA, FACSM, Kristi S. Kelley, MEd, and Russell R. Pate, PhD, FACSM | Ineligible study type |

| | | |
|---|---|---|
| Association between cardiorespiratory fitness and depressive symptoms | Arthur Ney Alves Donato a | Ineligible study type |
| Association between physical activity, fitness and cardiovascular risk factors in children. Systematic review | Batalau, R., R. Goncalves, J. Carmo, J. Cruz, J. Leal and A. Palmeira | Ineligible study type |
| Association of blood pressure, obesity and physical activity with arterial stiffness in children: a systematic review and meta-analysis | Giulia Lona, Christoph Hauser, Sabrina Köchli, Denis Infanger, Katharina Endes, Arno Schmidt-Trucksäss and Henner Hanssen | Ineligible study type |
| Association of objectively measured physical activity and bone health in children and | V. L. Bland1 & M. Heatherington-Rauth2 & C. Howe3 & S. B. Going1 & J. W. Bea1,4 | Ineligible study type |
| Associations between accelerometry measured physical activity and sedentary time and the metabolic syndrome: A meta‑analysis of more than 6000 children and adolescents | Marius Renninger, Bjørge H. Hansen, Jostein Steene‑Johannessen, Susi Kriemler, Karsten Froberg, Kate Northstone, Luis Sardinha, Sigmund A. Anderssen, Lars B. Andersen, Ulf Ekelund, On behalf of the International Children's Accelerometry Database (ICAD) Collaborators | Ineligible study type |
| Associations between physical activity, sedentary behaviour and self-rated health among the general population of children and adolescents: a systematic review and meta-analysis | Tong Zhang, Guohua Lu and Xiu Yun Wu | Ineligible study type |

| | | |
|---|---|---|
| Associations of Objectively Measured Vigorous Physical Activity With Body Composition, Cardiorespiratory Fitness, and Cardiometabolic Health in Youth: A Review | Morgan H. Gralla et al | Ineligible study type |
| Before-school physical activity programs and youth physical activity, health and learning-related outcomes: a systematic review. | J. Woodforde, T. Alspo, J. Salmon, S. Gomersall, M. Stylianou | Ineligible study type |
| Behavioral interventions to prevent childhood obesity: A systematic review and metaanalyses of randomized trials | Kamath, C. C., K. S. Vickers, A. Ehrlich, L. McGovern, J. Johnson, V. Singhal, R. Paulo, A. Hettinger, P. J. Erwin and V. M. Montori | Ineligible study type |
| Biomechanical variations in children who are overweight and obese during high-impact activities: A systematic review and meta-analysis | Claire Spech, Matthew Paponetti, Cody Mansfield, Laura Schmitt, Matthews Briggs | Ineligible study type |
| Capacity-oriented approaches to developing childhood obesity interventions: a systematic review | Foster, B. A., E. Fu, N. Bendiks, C. S. Gaspard and M. Sharifi | Ineligible study type |
| Cardiometabolic risk and health behaviours in adolescents with normal-weight obesity: a systematic review | Bruna Clemente Cota, Lara Gomes Suhett, Nathália Nogueira Leite, Patrícia Feliciano Pereira, Sarah Aparecida Vieira Ribeiro and Sylvia do Carmo Castro Franceschini | Ineligible study type |
| Chances and Limitations of Video Games in the Fight | Mack, I., C. Bayer, N. Schaffeler, N. Reiband, E. | Ineligible study type |

| | | |
|---|---|---|
| against Childhood Obesity-A Systematic Review | Brolz, G. Zurstiege, F. Fernandez-Aranda, C. Gawrilow and S. Zipfel | |
| Childhood obesity: a global public health crisis | Karnik, S. and A. Kanekar | Ineligible study type |
| Combinations of physical activity, sedentary time, and sleep duration and their associations with depressive symptoms and other mental health problems in children and adolescents: a systematic review | Hugues Sampasa-Kanyinga, Ian Colman, Gary S. Goldfield, Ian Janssen, JianLi Wang, Irina Podinic, Mark S. Tremblay, Travis J. Saunders, Margaret Sampson and Jean-Philippe Chaput. | Ineligible study type |
| Combinations of Physical Activity, Sedentary Behavior, and Sleep Duration and Their Associations With Physical, Psychological, and Educational Outcomes in Children and Adolescents: A Systematic Review | Katrina Wilhite, Bridget Booker, Bo-Huei Huang, Devan Antczak, Lucy Corbett, Philip Parker, Michael Noetel, Chris Rissel, Chris Lonsdale, Borja del Pozo Cruz, and Taren Sanders | Ineligible study type |
| Community-based interventions to reduce overweight and obesity in China: a systematic review of the Chinese and English literature | Gao, Y., S. Griffiths and E. Y. Chan | Ineligible study type |
| Comparing interventions with network meta-analysis. | Bagg, Matthew K; Salanti, Georgia; McAuley, James H; | Ineligible study type |
| Comparison of accelerometer measured levels of physical activity and sedentary time between obese and non-obese children and adolescents: a systematic review | Elmesmari, R., A. Martin, J. J. Reilly and J. Y. Paton | Ineligible study type |

| | | |
|---|---|---|
| Contralateral Effects after Unilateral Strength Training: A Meta-Analysis Comparing Training Loads. | Fisher, James; Cirer-Sastre, Rafel; Beltrán-Garrido, Jose V.; Corbi, Francisco | Ineligible study type |
| Contribution of structured settings to time spent in physical activity and sedentary in youth: systematic review and meta-analysis | Rafael M. Tassitano et al. | Ineligible study type |
| Contribution of Walking to School to Individual and Population Moderate-Vigorous Intensity Physical Activity: Systematic Review and Meta-Analysis | Martin, A., J. Boyle, F. Corlett, P. Kelly and J. J. Reilly | Ineligible study type |
| Digital Interventions for Universal Health Promotion in Children and Adolescents: A Systematic Review | Christina Oh, Bianca Carducci, Tyler Vaivada, Zulfiqar A Bhutta. | Ineligible study type |
| Does organized sports participation in childhood and adolescence positively influence health? A review of reviews | Helga Birgit Bjørnarå, Thomas Westergren, Ellen Sejersted, Monica Klungland Torstveit, Bjørge Herman Hansen, Sveinung Berntsen, Elling Bere | Ineligible study type |
| Dose Response of Cardiorespiratory Fitness Interventions in Adolescents: A Systematic Review: | Zezhao Chen, Jingyuan Zhu, Weimo Zhu | Ineligible study type |
| E-&mHealth interventions targeting nutrition, physical activity, sedentary behavior, and/or obesity among children: A scoping review of systematic reviews and meta-analyses | Chelsea L. Kracht, Melinda Hutchesson, Mavra Ahmed, Andre Matthias Muller, Lee M.Ashton, Hannah M. Brown, Ann DeSmet, Carol A. Maher, Chelsea E. Mauch, Corneel | Ineligible study type |

| | | |
|---|---|---|
| | Vandelanotte, Zenong Yin, Megan Whatnall, Camille E. Short, Amanda E. Staiano | |
| Efects of Resistance Training on Physical Fitness in Healthy Children and Adolescents: An Umbrella Review | Melanie Lesinski,Michael Herz,Alina Schmelcher,Urs Granacher. | Ineligible study type |
| Effect of childhood obesity prevention programmes on blood lipids: a systematic review and meta-analysis | Cai, L., Y. Wu, L. J. Cheskin, R. F. Wilson and Y. Wang | Ineligible study type |
| Effect of childhood obesity prevention programs on blood pressure: a systematic review and meta-analysis | Cai, L., Y. Wu, R. F. Wilson, J. B. Segal, M. T. Kim and Y. Wang | Ineligible study type |
| EFFECT OF VIRTUAL REALITY EQUIPMENT BY ACTIVE VIDEO GAMING ON PHYSICAL ACTIVITY. | KURT, İREM; TANRIVERDI, MÜBERRA | Ineligible study type |
| Effectiveness of exercise intervention on improving fundamental movement skills and motor coordination in overweight/obese children and adolescents: A systematic review. | Han, Ahreum; Fu, Allan; Cobley, Stephen; Sanders, Ross H. | Ineligible study type |
| Effectiveness of family-based weight management interventions for children with overweight and obesity: an umbrella review | Li Kheng Chai, Clare Collins, Chris May, Katherine Brain, Denise Wong See, Tracy Burrows. | Ineligible study type |
| Effectiveness of lifestyle interventions in preventing harmful weight gain among | Nematullah Hayba1 \| Chris Rissel2 \| Margaret Allman Farinelli1 | Ineligible study type |

| | | |
|---|---|---|
| Effectiveness of lifestyle interventions in preventing harmful weight gain among adolescents: A systematic review of systematic reviews | Nematullah Hayba, Chris Rissel, Margaret Allman Farinelli | Ineligible study type |
| Effectiveness of obesity prevention and control policies and programs for children: A systematic review of natural experiment studies | Bramante, C., R. Thornton, W. Bennett, A. Zhang, R. F. Wilson and G. Eva Tsen | Ineligible study type |
| Effectiveness of parent-centred interventions for the prevention and treatment of childhood overweight and obesity in community settings: A systematic review | Collins, C. E., T. L. Burrows, J. Bray, R. Asher, M. Young and P. J. Morgan | Ineligible study type |
| Effects of Active Videogames on Physical Activity and Related Outcomes Among Healthy Children: A Systematic Review | Liang, Y. and P. W. Lau | Ineligible study type |
| Effects of childhood obesity prevention programs on blood pressure and lipids: A systematic review and meta-analysis | Cai, L., Y. Wu, R. Wilson, L. Cheskin, M. Kim and Y. Wang | Ineligible study type |
| Effects of core training on dynamic balance stability: A systematic review and meta- | Stephen Heung-Sang Wong | Ineligible study type |
| Effects of dancing on physical activity levels of children and adolescents: a systematic review | Gabriela Cristina dos Santos, J ́essica do Nascimento Queiroz, Alvaro ́ Reischak-Oliveira, Josianne Rodrigues-Krause | Ineligible study type |

| | | |
|---|---|---|
| Effects of physical activity on heart rate variability in children and adolescents: a systematic review and meta-analysis | Hao Chen, Jing Xu, Hao Xie, Yufen Huang, Xin Shen, Fei Xu | Ineligible study type |
| Effects of resistance training on metabolic fitness in children and adolescents: a systematic review | Benson, A. C., M. E. Torode and M. A. Fiatarone Singh | Ineligible study type |
| Effects of the FIFA 11+ program on performance, biomechanical measures, and physiological responses: A systematic review | Mojtaba Asgari, Bahareh Nazari, Mario Bizzini, Thomas Jaitner | Ineligible study type |
| Efficacy of exercise for treating overweight in children and adolescents: a systematic review | Atlantis, E., E. H. Barnes and M. A. Singh | Ineligible study type |
| European normative values for physical fitness in children and adolescents aged 9–17 years: results from 2 779 165 Eurofit performances representing 30 countries | Grant R Tomkinson, Kevin D Carver, Frazer Atkinson,Nathan D Daniell,Lucy K Lewis, John S Fitzgerald,Justin J Lang,Francisco B Ortega | Ineligible study type |
| Evidence Supporting the Essential Components of Physical Education as a Measure of Quality | Lynne Bryant, Sheri L. Burson, Jenna Fisher, Chad M. Killian, Shannon C. Mulhearn, Danielle Nesbitt, Christopher D. Pfledderer & Darla M. Castelli | Ineligible study type |
| Exercise Dose and Weight Loss in Adolescents with Overweight–Obesity: A Meta‑Regression | Lee Stoner, Michael W. Beets, Keith Brazendale, Justin B. Moore, R. Glenn Weaver | Ineligible study type |

| | | |
|---|---|---|
| Exercise-associated prevention of adult cardiovascular disease in children and adolescents: monocytes, molecular mechanisms, and a call for discovery | Dan M. Cooper and Shlomit Radom-Aizik | Ineligible study type |
| Exercise, health outcomes, and pædiatric obesity: A systematic review of meta-analyses | Antonio García-Hermosoa, Robinson Ramírez-Vélez b, Jose M. Saavedrac, | Ineligible study type |
| Exercise, health outcomes, and paediatric obesity: A systematic review of meta-analyses | Garcia-Hermoso, A., R. Ramirez-Velez and J. M. Saavedra | Ineligible study type |
| EXERGAMES AS A TOOL FOR THE ACQUISITION AND DEVELOPMENT OF MOTOR SKILLS AND ABILITIES: A SYSTEMATIC REVIEW | Medeiros, P., R. Capistrano, M. A. Zequinao, S. A. D. Silva, T. S. Beltrame and F. L. Cardoso | Ineligible study type |
| Exergaming as a strategic tool in the fight against childhood obesity: a systematic review | Lamboglia, C. M., V. T. da Silva, J. E. de Vasconcelos Filho, M. H. Pinheiro, M. C. Munguba, F. V. Silva Junior, F. A. de Paula and C. A. da Silva | Ineligible study type |
| Health-Related Physical Fitness and Activity in Homeschool: A Systematic Review With Implications for Return to Public School | Laura S. Kabiri, Ashley Messineo, Nikhil Gattu, Brian D.Ray, Nicholas K. Iammarino. | Ineligible study type |
| How Many US Children and Adolescents with Overweight and Obesity Could Improve Their Percent Body Fat by Exercising?: Meta-Analytic Based Estimates | and Susan Buchholz, PhD, RN, ANP-BC, FAANP, FAAN3 | Ineligible study type |

| | | |
|---|---|---|
| Impact of physical activity intervention programs on self-efficacy in youths: a systematic review | Cataldo, R., J. John, L. Chandran, S. Pati and A. L. Shroyer | Ineligible study type |
| Impact of recess interventions on children's physical activity--a meta-analysis | Erwin, H. E., M. Ickes, S. Ahn and A. Fedewa | Ineligible study type |
| In search of quality evidence for lifestyle management and glycemic control in children and adolescents with type 2 diabetes: A systematic review | Johnson, S. T., A. S. Newton, M. Chopra, J. Buckingham, T. T. Huang, P. W. Franks, M. M. Jetha and G. D. Ball | Ineligible study type |
| Increasing weight-bearing physical activity and calcium intake for bone mass growth in children and adolescents: A review of intervention trials | French, S. A., J. A. Fulkerson and M. Story | Ineligible study type |
| Iniciativas escolares y deportivas lideradas desde la Federation Internationale de Football Association (FIFA): revision sistematica | Correa, J. E., J. F. Meneses-Echavez, N. C. Barengo, G. Tovar, E. Ruiz-Castellanos, F. Lobelo and R. Ramirez-Velez | Ineligible study type |
| Inter-individual differences in body mass index were not observed as a result of aerobic exercise in children and adolescents with overweight and obesity | George A. Kelley, Kristi S. Kelley, Russell R. Pate | Ineligible study type |
| Interdisciplinarity as a strategy for the prevention of systemic arterial hypertension in children: a systematic review | Bernardi, L., M. C. Franca, A. M. Xavier and D. Novello | Ineligible study type |
| Interventions for treating obesity in children | Hiltje Oude Luttikhuis, Louise Baur, Hanneke Jansen, Vanessa A Shrewsbury, Claire O'Malley, Ronald P Stolk, and Carolyn D Summerbell | Ineligible study type |
| Interventions in outside-school hours childcare settings for promoting physical activity amongst schoolchildren aged 4 to 12 years (Protocol) | Virgara R, Maher C, Lewis LK, Phillips A, Wolfenden L, Okely A, Beets MW, Richardson M, Baldock K | Ineligible study type |
| Interventions to prevent global childhood overweight and obesity: a systematic review | Bleich, S. N., K. A. Vercammen, L. Y. Zatz, J. M. Frelier, C. B. Ebbeling and A. Peeters | Ineligible study type |
| Interventions to prevent obesity in children and adolescents: a systematic literature review | Flodmark, C. E., C. Marcus and M. Britton | Ineligible study type |
| Interventions with children and parents to improve physical activity and body mass index: a meta-analysis | Dellert, J. C. and P. Johnson | Ineligible study type |
| Is adherence to the Mediterranean Diet associated with healthy habits and physical fitness? A systematic review and meta-analysis including 565,421 youths | Antonio García-Hermoso, Yasmin Ezzatvar, José Francisco López-Gil, Robinson Ramírez-Vélez, Jordi Olloquequi, Mikel Izquierdo | Ineligible study type |
| Is device-measured vigorous-intensity physical activity associated with health-related outcomes in children and adolescents? A systematic review and meta-analysis | Antonio Garc´ıa-Hermoso , Yasmin Ezzatvar , Robinson Ram´ırez-Velez , Jordi Olloquequi , Mikel Izquierdo´ | Ineligible study type |

| | | |
|---|---|---|
| Jump rope training effects on health- and sport-related physical fitness in young participants: A systematic review with meta-analysis | Utkarsh Singh, Akhilesh Kumar Ramachandran, Rodrigo Ramirez-Campillo, Alejandro Perez-Castilla, José Afonso, Filipe Manuel Clemente & Jon Oliver | Ineligible study type |
| mHealth Interventions to Reduce Physical Inactivity and Sedentary Behavior in Children and Adolescents: Systematic Review and Meta-analysis of Randomized Controlled Trials | Hannes Baumann, Janis Fiedler, Kathrin Wunsch, Alexander Woll, Bettina Wollesen. | Ineligible study type |
| Motor competence and health related physical fitness in youth: A systematic review | Cattuzzo, M. T., R. Dos Santos Henrique, A. H. Re, I. S. de Oliveira, B. M. Melo, M. de Sousa Moura, R. C. de Araujo and D. Stodden | Ineligible study type |
| Moving beyond the stigma: systematic review of video games and their potential to combat obesity | Guy, S., A. Ratzki-Leewing and F. Gwadry-Sridhar | Ineligible study type |
| Neighbourhood speed limit and childhood obesity | Miyang Luo, Hanqi Li, Xiongfeng Pan, Teng Fei, Shaoqing Dai, Ge Qiu, Yuxuan Zou, Heleen Vos, Jiayou Luo, Peng Jia | Ineligible study type |
| Non-alcoholic fatty liver disease in children and adolescents: Efficacy of lifestyle change-a systematic review and meta-analysis | Melere, M., C. Ferreira, B. Horta, M. Epifanio, A. Mattos and M. Mouzaki | Ineligible study type |
| Non-pharmacological interventions to reduce the risk of diabetes in people with | Gillett, M., P. Royle, A. Snaith, G. Scotland, A. Poobalan, M. Imamura, C. Black, M. | Ineligible study type |

| | | |
|---|---|---|
| impaired glucose regulation: a systematic review and economic evaluation | Boroujerdi, S. Jick, L. Wyness, P. McNamee, A. Brennan and N. Waugh | |
| Nonphysical effects of exergames on child and adolescent well-being: a comprehensive systematic review | Joronen, K., A. Aikasalo and A. Suvitie | Ineligible study type |
| O EFEITO DA ATIVIDADE FÍSICA NA COMPOSIÇÃO CORPORAL E APTIDÃO CARDIORRESPIRATÓRIA: UMA REVISÃO SISTEMÁTICA. / The effect of physical activity on body composition and cardiorespiratory fitness: A systematic review | Tirone Rosário, Gustavo; Pedretti, Augusto; Teixeira e Seabra, André Filipe | Ineligible study type |
| Obesity in children | Arteburn, D. E. | Ineligible study type |
| Obesity in children | Canoy, D. and P. Bundred | Ineligible study type |
| Overview of meta-analysis on prevention and treatment of childhood obesity | Luciana Bahia, Camila Wohlgemuth Schaan, Karen Sparrenberger, Gabriela de Azevedo Abreu, Laura Augusta Barufaldi, Walmir Coutinhof e Beatriz D. Schaan | Ineligible study type |
| Parental influence on models of primary prevention of cardiovascular disease in children | Norton, D. E., E. S. Froelicher, C. M. Waters and V. Carrieri-Kohlman | Ineligible study type |
| Peer-assisted learning in school physical education, sport and physical activity programmes: a systematic review | Jenkinson, K. A., G. Naughton and A. C. Benson | Ineligible study type |

| | | |
|---|---|---|
| Physical Activity and Cognitive Functioning of Children: A Systematic Review | Bidzan-Bluma, I. and M. Lipowska | Ineligible study type |
| Physical activity and low back pain in children and adolescents: a systematic review | Agnieszka Kędra, Magdalena Plandowska, Przemysław Kędra, Dariusz Czaprowski | Ineligible study type |
| Physical Activity in Young Children: A Systematic Review of Parental Influences | Mitchell, J., H. Skouteris, M. McCabe, L. A. Ricciardelli, J. Milgrom, L. A. Baur, M. Fuller-Tyszkiewicz and G. Dwyer | Ineligible study type |
| Physical activity increases bone mass during growth | Karlsson, M. K., A. Nordqvist and C. Karlsson | Ineligible study type |
| Physical Activity Interventions for Neurocognitive and Academic Performance in Overweight and Obese Youth: A Systematic Review | Bustamante, E. E., C. F. Williams and C. L. Davis | Ineligible study type |
| Physical Activity, Academic Performance and Cognition in Children and Adolescents. A Systematic Review | Haapala, E. | Ineligible study type |
| Physical Activity, Fitness, Cognitive Function, and Academic Achievement in Children: A Systematic Review | Donnelly, J. E., C. H. Hillman, D. Castelli, J. L. Etnier, S. Lee, P. Tomporowski, K. Lambourne and A. N. Szabo-Reed | Ineligible study type |
| Physical education and sport in Brazil | Pignone, M. P., C. J. Phillips, C. M. Lannon, C. D. Mulrow, S. M. Teutsch, K. N. Lohr and B. L. Whitener | Ineligible study type |
| Physical fitness related to sportive performance in artistic gymnastics. / AptidÃ£o fÃsica relacionada ao desempenho esportivo na ginÃ¡stica artÃstica | dos Santos Lemos, C. F., D. R. dos Santos and N. M. Moura Soares | Ineligible study type |
| Prevention and treatment of childhood and adolescent obesity: a systematic review of meta‐analyses | Theodora Psaltopoulou, Stamatios Tzanninis, Ioannis Ntanasis‐Stathopoulos, George Panotopoulos, Myrto Kostopoulou, Ioannis‐Georgios Tzanninis, Anastasia Tsagianni, Theodoros N. Sergentanis | Ineligible study type |
| Psychological Wellbeing in Physical Education and School Sports: A Systematic Review | Javier Piñeiro-Cossio, Antonio Fernández-Martínez, Alberto Nuviala, Raquel Pérez-Ordás. | Ineligible study type |
| REBOUND EFFECT OF INTERVENTION PROGRAMS TO REDUCE OVERWEIGHT AND OBESITY IN CHILDREN AND ADOLESCENTS; SYSTEMATIC REVIEW | Aguilar Cordero, M. J., A. Ortegon Pinero, L. Baena Garcia, J. P. Noack Segovia, M. C. Levet Hernandez and A. M. Sanchez Lopez | Ineligible study type |
| Recent Trends in Sedentary Time: A Systematic Literature Review | Hui Fang, Yuan Jing, Jie Chen , Yanqi Wu and Yuehua Wan | Ineligible study type |
| Recreational football is medicine against non‐communicable diseases: A systematic review. | Sarmento, Hugo; Manuel Clemente, Filipe; Marques, Adilson; Milanovic, Zoran; David Harper, Liam; Figueiredo, António; | Ineligible study type |
| Regular Doses of Nature: The Efficacy of Green Exercise | Mike Rogerson, Carly Wood, Jules Pretty, Patrick | Ineligible study type |

| | | |
|---|---|---|
| Interventions for Mental Wellbeing | Schoenmakers, Dan Bloomfield and Jo Barton | |
| Relationships of physical activity and sedentary behaviour with the previous and subsequent nights' sleep in children and youth: A systematic review and meta-analysis | Wendy Yajun Huang, Robin Sze-Tak Ho, Mark S. Tremblay, Stephen Heung-Sang Wong | Ineligible study type |
| Republished research: effectiveness of intervention on physical activity of children: systematic review and meta-analysis of controlled trials with objectively measured outcomes (EarlyBird 54) | Metcalf, B., W. Henley and T. Wilkin | Ineligible study type |
| Review article: the management of paediatric nonalcoholic fatty liver disease | Mitchel, E. B. and J. E. Lavine | Ineligible study type |
| RevisiÃ³n sistemÃ¡tica sobre los estudios de intervenciÃ³n de actividad fÃsica para el tratamiento de la obesidad. / Systematic Review of Physical Activity Programs for the treatment of Obesity | Cuadri FernÃ¡ndez, J., I. Tornero QuiÃ±ones, Ã. n. Sierra Robles and J. s. M. SÃ¡ez Padilla | Ineligible study type |
| School and family-based interventions for promoting a healthy lifestyle among children and adolescents in Italy: a systematic review | Gorga, E., V. Regazzoni, S. Bansilal, V. Carubelli, E. Trichaki, M. Gavazzoni, C. Lombardi, R. Raddino and M. Metra | Ineligible study type |
| School-based interventions promoting both physical activity and healthy eating in | De Bourdeaudhuij, I., E. Van Cauwenberghe, H. Spittaels, J. M. Oppert, C. Rostami, J. Brug, | Ineligible study type |

| | | |
|---|---|---|
| Europe: a systematic review within the HOPE project | F. Van Lenthe, T. Lobstein and L. Maes | |
| Sedentary Time and Behavior during School: A Systematic Review and Meta-Analysis | Catherine A. Egan, Collin A. Webster, Michael W. Beets, R. Glenn Weaver, Laura Russ, Daniel Michael, Danielle Nesbitt & Karie L. Orendorff | Ineligible study type |
| Skeletal adaptations associated with pre-pubertal gymnastics participation as determined by DXA and pQCT: a systematic review and meta-analysis | Burt, L. A., D. A. Greene, G. Ducher and G. A. Naughton | Ineligible study type |
| Sleep Health Promotion Interventions and Their Effectiveness: An Umbrella Review | Nguyen, N.-M.; Dibley, M.J.; Tang, H.K.; Alam, A.; | Ineligible study type |
| Sleep health promotion interventions and their effectiveness: An umbrella review | Albakri, U.; Drotos, E.; Meertens, R.; | Ineligible study type |
| Soccer helps build strong bones during growth: a systematic review and meta-analysis | Lozano-Berges, G., A. Matute-Llorente, A. Gonzalez-Aguero, A. Gomez-Bruton, A. Gomez-Cabello, G. Vicente-Rodriguez and J. A. Casajus | Ineligible study type |
| Sport and dance interventions for healthy young people (15–24 years) to promote subjective well-being: a systematic review | Louise Mansfield,Tess Kay, Catherine Meads,Lily Grigsby-Duffy, Jack Lane,Alistair John,Norma Daykin,Paul Dolan,Stefano Testoni,Guy Julier,Annette | Ineligible study type |

| | | |
|---|---|---|
| | Payne,Alan Tomlinson,Christina Victor | |
| Systematic review of community-based childhood obesity prevention studies | Bleich, S. N., J. Segal, Y. Wu, R. Wilson and Y. Wang | Ineligible study type |
| Systematic Review of Natural Experiments for Childhood Obesity Prevention and Control | Carolyn T. Bramante, MD, MPH,1 Rachel L.J. Thornton, MD, PhD,2,3,4 Wendy L. Bennett, MD, MPH,1,4 Allen Zhang, BS,5 Renee F. Wilson, MS,5 Eric B. Bass, MD, MPH,1,5 Eva Tseng, MD, MPH1,4 | Ineligible study type |
| Systematic review of paediatric weight management interventions delivered in the home setting | Appelhans, B. M., O. A. Moss and L. A. Cerwinske | Ineligible study type |
| SYSTEMATIC REVIEW OF PHYSICAL ACTIVITY AND SEDENTARY BEHAVIOR INDICATORS IN SOUTH-AMERICAN PRESCHOOL CHILDREN | Paulo Henrique Guerraa, Valter Cordeiro Barbosa Filhob , Angélica Almeidaa, Letícia de Souza Silvaa, Marcelo Tedesco Vidal Pintoa, Renan Martinelli Leonela, Evelyn Helena Corgosinho Ribeiroc , Alex Antonio Florindoc | Ineligible study type |
| Systematic review of the health benefits of physical activity and fitness in school-aged children and youth | Janssen, I. and A. G. Leblanc | Ineligible study type |
| The Effect of Aerobic Exercise on Recovery in Adolescents who Report Emotional and Cognitive Symptoms after Sport-Related Concussion | Andrew Nowak, Haley Chizuk, Muhammad Subhan Zahid Nazir, Abigail E. Bisson, Christopher Stavisky, John Leddy, Jeffery Miecznikowski, | Ineligible study type |

| | | |
|---|---|---|
| | Mohammad Haider, Barry Willer | |
| The Effect of Diet, Exercise, and Lifestyle Intervention on Childhood Obesity: A Network Meta-Analysis | Ji-Hyun Bae, Ph.D, Professor, Hyorim Lee, Ph.D, Assistant Professor | Ineligible study type |
| The effect of high Intensity interval training versus moderate intensity continuous training on arterial stiffness and 24h blood pressure responses: A systematic review and meta-analysis. | Way, Kimberley L.; Sultana, Rachelle N.; Sabag, Angelo; Baker, Michael K.; Johnson, Nathan A. | Ineligible study type |
| The effect of influencing autonomy for obesity prevention: A review and meta-analysis of school based interventions | Haynes, E., P. Glasziou and D. Reidlinger | Ineligible study type |
| The effect of physical activity on anxiety in children and young people: a systematic review and meta-analysis | Tim Carter, Michaela Pascoe, Anastasios Bastounis, Ioannis D. Morres, Patrick Callaghan, Alexandra G. Parker | Ineligible study type |
| The Effect of Swimming During Childhood and Adolescence on Bone Mineral Density: A Systematic Review and Meta-Analysis | Gomez-Bruton, A., J. Montero-Marin, A. Gonzalez-Aguero, J. Garcia-Campayo, L. A. Moreno, J. A. Casajus and G. Vicente-Rodriguez | Ineligible study type |
| The effectiveness of intervention on the physical activity of children: Systematic review and meta-analysis of controlled trials with objectively-measured outcomes | Metcalf, B. S., W. Henley and T. J. Wilkin | Ineligible study type |

| | | |
|---|---|---|
| The Effects of Exercise on BDNF Levels in Adolescents: A Systematic Review with Meta-Analysis | Kesley Pablo Morais de Azevedo, Victor Hugo de Oliveira, Gidyenne Christine Bandeira Silva de Medeiros, Ádala Nayana de Sousa Mata, Daniel Ángel García, Daniel Guillén Martínez, José Carlos Leitão, Maria Irany Knackfuss and Grasiela Piuvezam | Ineligible study type |
| The effects of high-intensity interval training versus moderate-intensity continuous training on fat loss and cardiometabolic health in pediatric obesity | Jing-Xin Liu, MSa,b, Lin Zhu, PhDa, Jia-Min Deng, MSa | Ineligible study type |
| The Effects of Physical Activity and Physical Fitness on Children's Achievement and Cognitive Outcomes: A Meta-Analysis | Fedewa, A. L. and S. Ahn | Ineligible study type |
| The Effects of Resistance Training on Blood Pressure in Preadolescents and Adolescents: A Systematic Review and Meta-Analysis | Carles Miguel Guillem, Andrés Felipe Loaiza-Betancur, Tamara Rial Rebullido, Avery D. Faigenbaum and Iván Chulvi-Medrano | Ineligible study type |
| The impact of multicomponent weight management interventions on quality of life in adolescents affected by overweight or obesity: a meta-analysis of randomized controlled trials | M. Murray , J. L. Pearson, A. L. Dordevic and M. P. Bonham | Ineligible study type |
| The relationship between active travel to school and health-related fitness in children and adolescents: a systematic review | Lubans, D. R., C. A. Boreham, P. Kelly and C. E. Foster | Ineligible study type |
| The relationship between physical activity and sleep from mid adolescence to early adulthood. A systematic review of methodological approaches and meta-analysis | Lang, C., N. Kalak, S. Brand, E. Holsboer-Trachsler, U. Puhse and M. Gerber | Ineligible study type |
| The school environment and adolescent physical activity and sedentary behaviour: a mixed-studies systematic review | Morton, K. L., A. J. Atkin, K. Corder, M. Suhrcke and E. M. van Sluijs | Ineligible study type |
| Treatment of pediatric obesity: A systematic review and meta-analysis of randomized trials | McGovern, L., J. N. Johnson, R. Paulo, A. Hettinger, V. Singhal, C. Kamath, P. J. Erwin and V. M. Montori | Ineligible study type |
| Unravelling the association between accelerometer‑derived physical activity and adiposity among preschool children: A systematic review and meta‑analyses | Rikstje Wiersma, Barbara F. Haverkamp, Jasper H. van Beek, André M.J. Riemersma, H. Marike Boezen, Nynke Smidt, Eva Corpeleijn, Esther Hartman | Ineligible study type |
| Weight training in youth-growth, maturation, and safety: an evidence-based review | Malina, R. M. | Ineligible study type |
| What works in china? A literature review on effectiveness of current intervention for combating obesity among Chinese school-aged children | Li, X. and T. Hesketh | Ineligible study type |

| | | |
|---|---|---|
| Clinical interventions in overweight and obesity: a systematic literature review 2009-2014 | Rajmil, L., J. Bel, R. Clofent, C. Cabezas, C. Castell and M. Espallargues | Ineligible population |
| EFFECTS OF SCHOOL-BASED PHYSICAL ACTIVITY AND NUTRITION PROGRAMS IN SPANISH ADOLESCENTS: SYSTEMATIC REVIEW | Perez Lopez, I. J., P. Tercedor Sanchez and M. Delgado-Fernandez | Ineligible population |
| Physical activity and healthy eating in Brazilian students: a review of intervention programs | Souza, E. A., V. C. Barbosa Filho, J. A. Nogueira and M. R. Azevedo Junior | Ineligible population |
| A Meta-Analysis of Obesity Interventions Among U.S. Minority Children | Seo, D. C. and J. Sa | Ineligible population |
| A meta-analysis to determine strength training related dose-response relationships for lower-limb muscle power development in young athletes | Slimani, M., A. Paravlic and U. Granacher | Ineligible population |
| A Meta-Analytic Review of Obesity Prevention Programs for Children and Adolescents: The Skinny on Interventions that Work | Stice, E., H. Shaw and C. N. Marti | Ineligible population |
| A systematic review and meta-analysis of whole of community interventions to prevent excessive population weight gain | Wolfenden, L., R. Wyse, M. Nichols, S. Allender, L. Millar and P. McElduff | Ineligible population |

| | | |
|---|---|---|
| A Systematic Review of Digital Interventions for Improving the Diet and Physical Activity Behaviors of Adolescents | Rose, T., M. Barker, C. Maria Jacob, L. Morrison, W. Lawrence, S. Strommer, C. Vogel, K. Woods-Townsend, D. Farrell, H. Inskip and J. Baird | Ineligible population |
| A systematic review of home-based childhood obesity prevention studies | Showell, N. N., O. Fawole, J. Segal, R. F. Wilson, L. J. Cheskin, S. N. Bleich, Y. Wu, B. Lau and Y. Wang | Ineligible population |
| A systematic review of interventions to promote physical activity in the preschool setting | Temple, M. and J. C. Robinson | Ineligible population |
| A systematic review of lifestyle interventions for chronic diseases in rural communities | Smith, S. A. and B. Ansa | Ineligible population |
| A Systematic Review of Obesity Disparities Research | Pratt, C. A., C. M. Loria, S. S. Arteaga, H. L. Nicastro, M. Lopez-Class, J. M. de Jesus, P. Srinivas, C. Maric-Bilkan, L. Schwartz Longacre, J. E. A. Boyington, A. Wouhib and N. Gavini | Ineligible population |
| A Systematic Review of Obesity Prevention Intervention Studies among Immigrant Populations in the US | Tovar, A., A. M. N. Renzaho, A. D. Guerrero, N. Mena and G. X. Ayala | Ineligible population |
| A Systematic Review of Rural, Theory-based Physical Activity Interventions | Walsh, S. M., M. R. Meyer, A. Gamble, M. S. Patterson and J. B. Moore | Ineligible population |
| A systematic review of school-based physical activity interventions on children's wellbeing | Rafferty, R., G. Breslin, D. Brennan and D. Hassan | Ineligible population |

| | | |
|---|---|---|
| Balance training for neuromuscular control and performance enhancement: a systematic review | Zech, A., M. Hubscher, L. Vogt, W. Banzer, F. Hansel and K. Pfeifer | Ineligible population |
| Childhood obesity: a (re) programming disease? | Paes, S. T., C. F. Goncalves, M. M. Terra, T. S. Fontoura, M. O. Guerra, V. M. Peters, P. C. Mathias and A. E. Andreazzi | Ineligible population |
| Clinical outcomes to exercise training in type 1 diabetes: A systematic review and meta-analysis | Ostman, C., D. Jewiss, N. King and N. A. Smart | Ineligible population |
| Direct and Indirect Influence of Physical Education-Based Interventions on Physical Activity: A Review | Slingerland, M. and L. Borghouts | Ineligible population |
| Do active video games benefit the motor skill development of non-typically developing children and adolescents: A systematic review | Page, Z. E., S. Barrington, J. Edwards and L. M. Barnett | Ineligible population |
| Do exercises used in injury prevention programmes modify cutting task biomechanics? A systematic review with meta-analysis | Pappas, E., E. J. Nightingale, M. Simic, K. R. Ford, T. E. Hewett and G. D. Myer | Ineligible population |
| Does Exercise Influence Pediatric Bone? A Systematic Review | Specker, B., N. W. Thiex and R. G. Sudhagoni | Ineligible population |
| Dose-Response Relationship of Neuromuscular Training for Injury Prevention in Youth Athletes: A Meta-Analysis | Steib, S., A. L. Rahlf, K. Pfeifer and A. Zech | Ineligible population |
| Educational and behavioral interventions in childhood obesity: A systematic review with metanalysis of randomized clinical trials | Sbruzzi, G., B. Eibel, C. C. Cesa, R. A. Ribeiro, S. M. Barbiero, R. Petkowicz, W. B. De Souza, C. Martins, B. D. Schaan and L. Pellanda | Ineligible population |
| Effect of classroom-based physical activity interventions on academic and physical activity outcomes: a systematic review and meta-analysis | Watson, A., A. Timperio, H. Brown, K. Best and K. D. Hesketh | Ineligible population |
| Effective interventions in overweight or obese young children: systematic review and meta-analysis | van Hoek, E., E. J. Feskens, L. I. Bouwman and A. J. Janse | Ineligible population |
| Effectiveness of interventions to improve lifestyle behaviors among socially disadvantaged children in Europe | Wijtzes, A. I., V. M. van de Gaar, A. van Grieken, M. L. de Kroon, J. P. Mackenbach, F. J. van Lenthe, W. Jansen and H. Raat | Ineligible population |
| Effectiveness of interventions to promote physical activity in children and adolescents: systematic review of controlled trials | van Sluijs, E. M., A. M. McMinn and S. J. Griffin | Ineligible population |
| EFFECTIVENESS OF OBESITY INTERVENTION PROGRAMS BASED ON GUIDELINES FOR ADOLESCENT STUDENTS: SYSTEMATIC REVIEW | Wolf, V. L. W., J. E. Samur-San-Martin, S. F. Sousa, H. D. O. Santos, A. G. Folmann, R. R. Ribeiro and G. Guerra-Junior | Ineligible population |
| Effectiveness of pre-school- and school-based interventions to impact weight-related behaviours in African American | Robinson, L. E., E. K. Webster, M. C. Whitt-Glover, T. G. Ceaser and S. Alhassan | Ineligible population |

| | | |
|---|---|---|
| children and youth: a literature review | | |
| Effectiveness of preventive school-based obesity interventions in low- and middle-income countries: a systematic review | Verstraeten, R., D. Roberfroid, C. Lachat, J. L. Leroy, M. Holdsworth, L. Maes and P. W. Kolsteren | Ineligible population |
| Effectiveness of school-based physical activity programmes on cardiorespiratory fitness in children: a meta-analysis of randomised controlled trials | Pozuelo-Carrascosa, D. P., A. Garcia-Hermoso, C. Alvarez-Bueno, M. Sanchez-Lopez and V. Martinez-Vizcaino | Ineligible population |
| Effectiveness of Universal Self-regulation-Based Interventions in Children and Adolescents: A Systematic Review and Meta-analysis | Pandey, A., D. Hale, S. Das, A. L. Goddings, S. J. Blakemore and R. M. Viner | Ineligible population |
| Effects of exercise training on blood rheology: a meta-analysis | Romain, A. J., J. F. Brun, E. Varlet-Marie and E. Raynaud de Mauverger | Ineligible population |
| Effects of intervention programs on child and adolescent BMI: A meta-analysis study | Vasques, C., P. Magalhaes, A. Cortinhas, P. Mota, J. Leitao and V. P. Lopes | Ineligible population |
| Effects of Physical Activity Intervention in Youth: A Review | Ringuet, C. J. and S. G. Trost | Ineligible population |
| Effects of physical activity interventions on cognitive and academic performance in children and adolescents: a novel combination of a systematic review and | Singh, A. S., E. Saliasi, V. van den Berg, L. Uijtdewilligen, R. H. M. de Groot, J. Jolles, L. B. Andersen, R. Bailey, Y. K. Chang, A. Diamond, I. Ericsson, J. L. Etnier, A. L. Fedewa, C. H. Hillman, T. | Ineligible population |
| recommendations from an expert panel | McMorris, C. Pesce, U. Puhse, P. D. Tomporowski and M. J. M. Chinapaw | |
| Effects of Physical Activity on Motor Skills and Cognitive Development in Early Childhood: A Systematic Review | Zeng, N., M. Ayyub, H. Sun, X. Wen, P. Xiang and Z. Gao | Ineligible population |
| Effects of school-based interventions for direct delivery of physical activity on fitness and cardiometabolic markers in children and adolescents: a systematic review of randomized controlled trials | Sun, C., A. Pezic, G. Tikellis, A. L. Ponsonby, M. Wake, J. B. Carlin, V. Cleland and T. Dwyer | Ineligible population |
| Effects of strategies to promote children's physical activity on potential mediators | Salmon, J., H. Brown and C. Hume | Ineligible population |
| Evaluation of the effectiveness of neuromuscular training to reduce anterior cruciate ligament injury in female athletes: a critical review of relative risk reduction and numbers-needed-to-treat analyses | Sugimoto, D., G. D. Myer, J. M. McKeon and T. E. Hewett | Ineligible population |
| Evidence based physical activity for school-age youth | Strong, W. B., R. M. Malina, C. J. Blimkie, S. R. Daniels, R. K. Dishman, B. Gutin, A. C. Hergenroeder, A. Must, P. A. Nixon, J. M. Pivarnik, T. Rowland, S. Trost and F. Trudeau | Ineligible population |

| | | |
|---|---|---|
| Exercise and VO2 max in children: a meta-analysis | Payne, V. G. and J. R. Morrow, Jr. | Ineligible population |
| Exercise interventions for smoking cessation | Ussher, M. H., A. H. Taylor and G. E. J. Faulkner | Ineligible population |
| Exercise-based injury prevention in child and adolescent sport: a systematic review and meta-analysis | Rossler, R., L. Donath, E. Verhagen, A. Junge, T. Schweizer and O. Faude | Ineligible population |
| Exergaming and obesity in youth: current perspectives | Zeng, N. and Z. Gao | Ineligible population |
| Health game interventions to enhance physical activity self-efficacy of children: a quantitative systematic review | Pakarinen, A., H. Parisod, J. Smed and S. Salantera | Ineligible population |
| Health information technology in screening and treatment of child obesity: a systematic review | Smith, A. J., A. Skow, J. Bodurtha and S. Kinra | Ineligible population |
| Impact of mobile apps to combat obesity in children and adolescents: A systematic literature review | Quelly, S. B., A. E. Norris and J. L. DiPietro | Ineligible population |
| Influence of physical activity on bone strength in children and adolescents: a systematic review and narrative synthesis | Tan, V. P., H. M. Macdonald, S. Kim, L. Nettlefold, L. Gabel, M. C. Ashe and H. A. McKay | Ineligible population |
| Interventions for preventing ankle ligament injuries | Quinn, K., P. Parker, R. de Bie, B. Rowe and H. Handoll | Ineligible population |
| Interventions for preventing obesity in children | Summerbell, C. D., E. Waters, L. D. Edmunds, S. Kelly, T. Brown and K. J. Campbell | Ineligible population |

| | | |
|---|---|---|
| Interventions for preventing obesity in children | Waters, E., A. de Silva‐Sanigorski, B. J. Burford, T. Brown, K. J. Campbell, Y. Gao, R. Armstrong, L. Prosser and C. D. Summerbell | Ineligible population |
| Interventions for treating obesity in children | Oude Luttikhuis, H., L. Baur, H. Jansen, V. A. Shrewsbury, C. O'Malley, R. P. Stolk and C. D. Summerbell | Ineligible population |
| Interventions for treating obesity in children | Summerbell, C. D., V. Ashton, K. J. Campbell, L. Edmunds, S. Kelly and E. Waters | Ineligible population |
| Interventions to promote walking: systematic review | Ogilvie, D., C. E. Foster, H. Rothnie, N. Cavill, V. Hamilton, C. F. Fitzsimons and N. Mutrie | Ineligible population |
| Maintenance interventions for overweight or obesity in children: a systematic review and meta-analysis | van der Heijden, L. B., E. J. M. Feskens and A. J. Janse | Ineligible population |
| Muscular adaptations in low- versus high-load resistance training: A meta-analysis | Schoenfeld, B. J., J. M. Wilson, R. P. Lowery and J. W. Krieger | Ineligible population |
| Pedagogical Approaches to and Effects of Fundamental Movement Skill Interventions on Health Outcomes: A Systematic Review | Tompsett, C., R. Sanders, C. Taylor and S. Cobley | Ineligible population |
| Physical activity interventions in children and young people with Type 1 diabetes mellitus: a systematic review with meta-analysis | Quirk, H., H. Blake, R. Tennyson, T. L. Randell and C. Glazebrook | Ineligible population |

| | | |
|---|---|---|
| Physical exercise and executive functions in preadolescent children, adolescents and young adults: a meta-analysis | Verburgh, L., M. Konigs, E. J. Scherder and J. Oosterlaan | Ineligible population |
| Prevention and treatment of pediatric obesity using mobile and wireless technologies: a systematic review | Turner, T., D. Spruijt-Metz, C. K. Wen and M. D. Hingle | Ineligible population |
| Prevention of Anterior Cruciate Ligament Rupture in Female Athletes: A Systematic Review | Ramirez, R. N., K. Baldwin and C. C. Franklin | Ineligible population |
| Prevention of overweight and obesity in children and youth: a systematic review and meta-analysis | Peirson, L., D. Fitzpatrick-Lewis, K. Morrison, D. Ciliska, M. Kenny, M. Usman Ali and P. Raina | Ineligible population |
| Promoting cycling: a review of interventions | Vuori, I. | Ineligible population |
| RelaciÃ³n entre actividad fÃsica, procesos cognitivos y rendimiento acadÃ©mico de escolares: revisiÃ³n de la literatura actual. / Relation of physical activity, cognitive and academic performance in children: Review of current literature | Reloba, S., L. J. Chirosa and R. E. Reigal | Ineligible population |
| Resistance training in children and youth: a meta-analysis | Payne, V. G., J. R. Morrow, Jr., L. Johnson and S. N. Dalton | Ineligible population |
| Resistance Training in Youth Improves Athletic Performance: A Systematic Review | Vesci, A. S., K. A. Webster, M. Sich and L. N. Marinko | Ineligible population |
| Review of Physical Activity and the IGF Family | Orenstein, M. R. and C. M. Friedenreich | Ineligible population |
| Role of video games in improving health-related outcomes: a systematic review | Primack, B. A., M. V. Carroll, M. McNamara, M. L. Klem, B. King, M. Rich, C. W. Chan and S. Nayak | Ineligible population |
| School Physical Education: The Effectiveness of Health-Related Interventions and Recommendations for Health-Promotion Practice | Quitério, A. L. D. | Ineligible population |
| School-based Internet obesity prevention programs for adolescents: A systematic literature review | Whittemore, R., A. Chao, R. Popick and M. Grey | Ineligible population |
| School-based obesity prevention programs: a meta-analysis of randomized controlled trials | Sobol-Goldberg, S., J. Rabinowitz and R. Gross | Ineligible population |
| Specific exercise effects of preventive neuromuscular training intervention on anterior cruciate ligament injury risk reduction in young females: meta-analysis and subgroup analysis | Sugimoto, D., G. D. Myer, K. D. Foss and T. E. Hewett | Ineligible population |
| Strength of obesity prevention interventions in early care and education settings: A systematic review | Ward, D. S., E. Welker, A. Choate, K. E. Henderson, M. Lott, A. Tovar, A. Wilson and J. F. Sallis | Ineligible population |
| Study quality on groin injury management remains low: a systematic review on treatment of groin pain in athletes | Serner, A., C. H. van Eijck, B. R. Beumer, P. Holmich, A. Weir and R. J. de Vos | Ineligible population |

| | | |
|---|---|---|
| Systematic literature review of built environment effects on physical activity and active transport - an update and new findings on health equity | Smith, M., J. Hosking, A. Woodward, K. Witten, A. MacMillan, A. Field, P. Baas and H. Mackie | Ineligible population |
| Systematic Review and Meta-Analysis of Multi-Component Interventions Through Schools to Increase Physical Activity | Russ, L. B., C. A. Webster, M. W. Beets and D. S. Phillips | Ineligible population |
| Systematic review of interventions for promoting active school transport | Villa-Gonzalez, E., Y. Barranco-Ruiz, K. R. Evenson and P. Chillon | Ineligible population |
| Systematic review of interventions to increase physical activity and physical fitness in African-Americans | Whitt-Glover, M. C. and S. K. Kumanyika | Ineligible population |
| The effect of school recess interventions on physical activity : a systematic review | Parrish, A. M., A. D. Okely, R. M. Stanley and N. D. Ridgers | Ineligible population |
| The effectiveness of interventions to increase physical activity among adolescent girls: a meta-analysis | Pearson, N., R. Braithwaite and S. J. Biddle | Ineligible population |
| The effectiveness of school-based physical activity interventions for adolescent girls: A systematic review and meta-analysis | Owen, M. B., W. B. Curry, C. Kerner, L. Newson and S. J. Fairclough | Ineligible population |
| The effects of active video games on patients' rehabilitative outcomes: A meta-analysis | Pope, Z., N. Zeng and Z. Gao | Ineligible population |

| | | |
|---|---|---|
| The effects of physical activity interventions on psychosocial outcomes in adolescents: A meta-analytic review | Spruit, A., M. Assink, E. van Vugt, C. van der Put and G. J. Stams | Ineligible population |
| The effects of resistance training interventions on vertical jump performance in basketball players: a meta-analysis | Sperlich, P. F., M. Behringer and J. Mester | Ineligible population |
| The Efficacy of Injury Prevention Programs in Adolescent Team Sports: A Meta-analysis | Soomro, N., R. Sanders, D. Hackett, T. Hubka, S. Ebrahimi, J. Freeston and S. Cobley | Ineligible population |
| Treatment of adolescent overweight and obesity | Tsiros, M. D., N. Sinn, A. M. Coates, P. R. Howe and J. D. Buckley | Ineligible population |
| Treatment of clinical insulin resistance in children: a systematic review | Quinn, S. M., L. A. Baur, S. P. Garnett and C. T. Cowell | Ineligible population |
| Treatment of Obesity in Young People—a Systematic Review and Meta-analysis | Selvendran, S. S., N. C. Penney, N. Aggarwal, A. W. Darzi and S. Purkayastha | Ineligible population |
| Treatment of overweight and obesity in children and youth: a systematic review and meta-analysis | Peirson, L., D. Fitzpatrick-Lewis, K. Morrison, R. Warren, M. Usman Ali and P. Raina | Ineligible population |
| Using Active Video Games for Physical Activity Promotion: A Systematic Review of the Current State of Research | Peng, W., J. C. Crouse and J.-H. Lin | Ineligible population |
| What childhood obesity prevention programmes work? | Wang, Y., L. Cai, Y. Wu, R. F. Wilson, C. Weston, O. Fawole, S. N. Bleich, L. J. Cheskin, N. | Ineligible population |

| | | |
|---|---|---|
| A systematic review and meta-analysis | N. Showell, B. D. Lau, D. T. Chiu, A. Zhang and J. Segal | |
| Yoga in the schools: a systematic review of the literature | Serwacki, M. L. and C. Cook-Cottone | Ineligible population |
| Young People and Physical Activity: A Systematic Review Matching Their Views to Effective Interventions | Rees, R., J. Kavanagh, A. Harden, J. Shepherd, G. Brunton, S. Oliver and A. Oakley | Ineligible population |
| “Kids Get in Shape with Nature”: A Systematic Review Exploring the Impact of Green Spaces on Childhood Obesity | Julius Cesar Alejandre1 and Mary Lynch2 | Ineligible population |
| A Majority of Anterior Cruciate Ligament Injuries Can Be Prevented by Injury Prevention Programs: A Systematic Review of Randomized Controlled Trials and Cluster–Randomized Controlled Trials With Meta-analysis. | Huang, Yu-Lun; Jung, Jaehun; Mulligan, Colin M.S.; Oh, Jaekeun; Norcross, Marc F. | Ineligible population |
| A meta-analysis of active video games on health outcomes among children and adolescents | Gao, Z., S. Chen, D. Pasco and Z. Pope | Ineligible population |
| A meta-analysis on the effects of exercise training on the VO2max in children and adolescents. / Meta anÃ¡lisis de los efectos del entrenamiento en el VO2mÃ¡x en niÃ±os y adolescentes | Carazo-Vargas, P. and J. Moncada-JimÃ©nez | Ineligible population |
| A meta-analytic review of eHealth interventions for pediatric health promoting and maintaining behaviors | Cushing, C. C. and R. G. Steele | Ineligible population |
| A rapid review of the effect of The Daily Mile on children&#39 | Gavin Breslin, Medbh HillyardID, Noel Brick, Stephen Shannon, Brenda McKay-Redmond, Barbara McConnell | Ineligible population |
| A review of eHealth interventions for physical activity and dietary behavior change | Norman, G. J., M. F. Zabinski, M. A. Adams, D. E. Rosenberg, A. L. Yaroch and A. A. Atienza | Ineligible population |
| A Systematic Literature Review of Peer-led Strategies for Promoting Physical Activity Levels of Adolescents | Fiona McHale, Kwok Ng, Sarah Taylor, Enrique Bengoechea, Catherine Norton, Donal O’Shea, Catherine Woods. | Ineligible population |
| A Systematic Literature Review of Peer-led Strategies for Promoting Physical Activity Levels of Adolescents | Fiona McHale, Kwok Ng, Sarah Taylor, Enrique Bengoechea, Catherine Norton, Donal O'Shea, Catherine Woods | Ineligible population |
| A systematic review and meta-analysis of exercise interventions for youth mental health | Xiang Wang, Zhi-dong Cai, Wan-ting Jiang, Yan-yan Fang, Wen-xin Sun & Xing Wang | Ineligible population |
| A systematic review and meta-analysis of interval training versus moderate-intensity continuous training on body adiposity | Keating, S. E., N. A. Johnson, G. I. Mielke and J. S. Coombes | Ineligible population |
| A Systematic Review and Meta-Analysis of Intervention for Pediatric Obesity Using Mobile Technology | Lee, J., M. Piao, A. Byun and J. Kim | Ineligible population |

| | | |
|---|---|---|
| A systematic review and meta-analysis of interventions designed to increase moderate-to-vigorous physical activity in school physical education lessons | Lonsdale, C., R. R. Rosenkranz, L. R. Peralta, A. Bennie, P. Fahey and D. R. Lubans | Ineligible population |
| A systematic review and meta-analysis of the overall effects of school-based obesity prevention interventions and effect differences by intervention components | Zheng Liu , Han-Meng Xu, Li-Ming Wen, Yuan-Zhou Peng, Li-Zi Lin Shuang Zhou, Wen-Hao Li and, Hai-Jun Wang. | Ineligible population |
| A systematic review exploring body image programmes and interventions in physical education | Charlotte Kerner, Amy Prescott, Robyn Smith, Michael Owen | Ineligible population |
| A Systematic Review of Active Video Games on Youth’s Body Composition and Physical Activity | Zan Gao, Nan Zeng, Daniel J. McDonough, Xiwen Su | Ineligible population |
| A systematic review of cognitive assessment in physical activity research involving children and adolescents. | Wade, L.; Leahy, A.; Lubans, D.; Smith, J.; Duncan, M. | Ineligible population |
| A systematic review of controlled trials of interventions to prevent childhood obesity and overweight: a realistic synthesis of the evidence | Connelly, J. B., M. J. Duaso and G. Butler | Ineligible population |
| A systematic review of intervention effects on potential mediators of children's physical activity | Brown, H., C. Hume, N. Pearson and J. Salmon | Ineligible population |

| | | |
|---|---|---|
| A systematic review of interventions targeting children and young peopleâ€™s physical activity behaviour in the home | Amanda L. Seims ,Jennifer Hall,Daniel D. Bingham,Amy Creaser,Anastasia Christoforou,Sally Barber,Andy Daly-Smith | Ineligible population |
| A systematic review of interventions to improve the dietary intake, physical activity and weight status of children attending family day care services | Sze Lin Yoong, Melanie Lum, Jannah Jones, Erin Kerr, Maryann Falkiner, Tessa Delaney, Sam McCrabb, Li Kheng Chai, Kirsty Seward and Alice Grady. | Ineligible population |
| A systematic review of interventions to increase physical activity and reduce sedentary behaviour following bariatric surgery. | James, Jennifer D.; Hardeman, Wendy; Goodall, Mark; Eborall, Helen; Sprung, Victoria S.; Bonnett, Laura J.; Wilding, John P.H. | Ineligible population |
| A Systematic Review of Multi-Component Comprehensive School Physical Activity Program (CSPAP) Interventions | Ann Pulling Kuhn, PhD, MS1 , Peter Stoepker, PhD2 | Ineligible population |
| A systematic review of parent-based interventions to improve multiple lifestyle risk behaviours among adolescents and parents | Katrina E. Champion, Lauren A. Gardner, Karrah McCann, Emily Hunter, Belinda Parmenter, Tess Aitken, Cath Chapman, Bonnie Spring, Louise Thornton, Tim Slade, Maree Teesson, Nicola C. Newton | Ineligible population |
| A systematic review of primary healthcare provider education and training using the Chronic Care Model for childhood obesity | Jacobson, D. and B. Gance-Cleveland | Ineligible population |

| | | |
|---|---|---|
| A systematic review of school based interventions aimed at the promotion of physical activity and/or healthy dietary behaviours in adolescents | Gilmartin, A., F. Sniehotta, V. Araujo-Soares, J. Macdiarmid, A. Avenell and D. Jackson | Ineligible population |
| A systematic review of school-based intervention studies for the prevention or reduction of excess weight among Chinese children and adolescents | Li, M., S. Li, L. A. Baur and R. R. Huxley | Ineligible population |
| A Systematic Review of the Effectiveness of Physical Education and School Sport Interventions Targeting Physical Activity, Movement Skills and Enjoyment of Physical Activity | Dudley, D., A. Okely, P. Pearson and W. Cotton | Ineligible population |
| A Systematic Review of the Relationships Between Physical Activity and Sleep in Early Childhood | Christine W. St. Laurent1, Katrina Rodheim1, Rebecca M.C. Spencer1,2 | Ineligible population |
| A systematic review on the effects of resistance and plyometric training on physical fitness in youth- What do comparative studies tell us? | Matti Peitz, Michael Behringer, Urs GranacherI | Ineligible population |
| A systematic review on workplace interventions to manage chronic musculoskeletal conditions. | Skamagki, Glykeria; King, Andrew; Duncan, Michael; Wåhlin, Charlotte | Ineligible population |
| A systematic review to determine the effectiveness of interventions designed to prevent overweight and obesity in pre-adolescent girls | Kesten, J. M., P. L. Griffiths and N. Cameron | Ineligible population |
| A Systematised Review of Primary School Whole Class Child Obesity Interventions: Effectiveness, Characteristics, and Strategies | Brown, E. C., D. S. Buchan, J. S. Baker, F. B. Wyatt, D. S. Bocalini and L. Kilgore | Ineligible population |
| Active Breaks and Cognitive Performance in Pupils: A Systematic Review. | Carlos Pastor-Vicedo, Juan; Prieto-Ayuso, Alejandro; López Pérez, Sergio; Martínez-Martínez, Jesús | Ineligible population |
| Active School-Based Interventions to Interrupt Prolonged Sitting Improve Daily Physical Activity: A Systematic Review and Meta-Analysis | Marta Amor-Barbosa, Anna Ortega-Martínez, Andoni Carrasco-Uribarren, Maria Caridad Bagur-Calafat | Ineligible population |
| Active video games for youth: a systematic review | Barnett, A., E. Cerin and T. Baranowski | Ineligible population |
| Active Video Games in Schools and Effects on Physical Activity and Health: A Systematic Review | Norris, E., M. Hamer and E. Stamatakis | Ineligible population |
| Active video games to promote physical activity in children and youth: a systematic review | Biddiss, E. and J. Irwin | Ineligible population |
| Acute effect of breaking up prolonged sitting on cognition: a systematic review | Ting-Yu Chueh, Yung-Chih Chen, Tsung-Min Hung | Ineligible population |
| Acute effects of moderate aerobic exercise on specific aspects of executive function in | Ludyga, S., M. Gerber, S. Brand, E. Holsboer-Trachsler and U. Puhse | Ineligible population |

| | | |
|---|---|---|
| different age and fitness groups: A meta-analysis | | |
| Adoption, implementation and sustainability of school-based physical activity and sedentary behaviour interventions in real-world settings: a systematic review | Samuel Cassar, Jo Salmon, Anna Timperio, Patti-Jean Naylor, Femke van Nassau, Ana María Contardo Ayala and Harriet Koorts | Ineligible population |
| Aerobic fitness and its relationship to sport, exercise training and habitual physical activity during youth | Armstrong, N., G. Tomkinson and U. Ekelund | Ineligible population |
| After-school program impact on physical activity and fitness: a meta-analysis | Beets, M. W., A. Beighle, H. E. Erwin and J. L. Huberty | Ineligible population |
| Age-Related Variation in Male Youth Athletes' Countermovement Jump After Plyometric Training: A Meta-Analysis of Controlled Trials | Moran, J. J., G. R. Sandercock, R. Ramirez-Campillo, C. M. Meylan, J. A. Collison and D. A. Parry | Ineligible population |
| Are primary/elementary school-based interventions effective in preventing/ameliorating excess weight gain? A systematic review of systematic reviews | Joanna Goldthorpe, Tracy Epton, Chris Keyworth, Rachel Calam, Christopher J. Armitage | Ineligible population |
| Are There Benefits from Teaching Yoga at Schools? A Systematic Review of Randomized Control Trials of Yoga-Based Interventions | Ferreira-Vorkapic, C., J. M. Feitoza, M. Marchioro, J. Simões, E. Kozasa and S. Telles | Ineligible population |

| | | |
|---|---|---|
| Are work organization interventions effective in preventing or reducing work-related musculoskeletal disorders? A systematic review of the literature. | Stock, Susan R.; Nicolakakis, Nektaria; Vézina, Nicole; Vézina, Michel; Gilbert, Louis; Turcot, Alice; Sultan-Taïeb, Hélène; Kin, Kathryn Sinden, R.; Denis, Marie-Agnès; Delga, Céline; Beaucage, Clément; | Ineligible population |
| Associations Among K–12 Student Outcomes, National Standards, and Physical Education Curricular Models: A Systematic Review | Christopher D. Pfledderer & Timothy A. Brusseau | Ineligible population |
| Associations between meeting 24-hour movement guidelines and health in the early | Feng, J., Zheng, C., Sit, C. H-P., | Ineligible population |
| Associations between outdoor play features and childrenâ€™s behavior and health: a systematic review | Joana V. Pereira, Fabio Vila-Nova, Guida Veiga, Frederico Lopes, Rita Cordovil | Ineligible population |
| Beneficial impact of exercise on bone mass in individuals under calorie restriction: a systematic review and Meta-analysis of randomized clinical trials. | Yarizadeh, Habib; Asadi, Sara; Baharlooi, Hussein; Setayesh, Leila; Kakavandi, Nader Rahimi; Hambly, Catherine; Djafarian, Kurosh; Mirzaei, Khadijeh | Ineligible population |
| Biomarkers of Physiological Responses to Periods of Intensified, Non-Resistance-Based Exercise Training in Well-Trained Male Athletes: A Systematic Review and Meta-Analysis. | Greenham, Grace; Buckley, Jonathan D.; Garrett, Joel; Eston, Roger; Norton, Kevin | Ineligible population |

| | | |
|---|---|---|
| Blood Flow Restriction Training for Athletes: A Systematic Review. | Wortman, Ryan J.; Brown, Symone M.; Savage-Elliott, Ian; Finley, Zachary J.; Mulcahey, Mary K. | Ineligible population |
| BONE GEOMETRY AND PHYSICAL ACTIVITY IN CHILDREN AND ADOLESCENTS: SYSTEMATIC REVIEW | Krahenbuhl, T., R. F. Guimaraes, A. A. Barros Filho and E. M. Goncalves | Ineligible population |
| Can chronic stretching change the muscle‐tendon mechanical properties? A review. | Freitas, S. R.; Mendes, B.; Le Sant, G.; Andrade, R. J.; Nordez, A.; Milanovic, Z.; | Ineligible population |
| Can exercise improve self esteem in children and young people? A systematic review of randomised controlled trials | Ekeland, E., F. Heian and K. B. Hagen | Ineligible population |
| Can respiratory muscle training change the blood pressure levels in hypertension? A systematic review with meta‐analysis. | Silva, Claudio D.; Abreu, Raphael M.; Rehder‐Santos, Patrícia; De Noronha, Marcos; Catai, Aparecida M. | Ineligible population |
| Change in Central Cardiovascular Function in Response to Intense Interval Training: A Systematic Review and Meta-analysis | ASTORINO, TODD A.; CAUSER, EJAZ; HAZELL, TOM J.; ARHEN, BENJAMIN B.; GURD, BRENDON J. | Ineligible population |
| Childhood Obesity Evidence Base Project: A Systematic Review and Meta-Analysis of a New Taxonomy of Intervention Components to Improve Weight Status in Children 2–5 Years of Age, 2005–2019 | Lori A.J. Scott-Sheldon, Larry V. Hedges, Chris Cyr,Deborah Young-Hyman,Laura Kettel Khan, Mackenzie Magnus,Heather King, Sonia Arteaga, John Cawley, Christina D. Economos, Debra Haire-Joshu, Christine M. Hunter, Bruce Y. Lee, Shiriki K. | Ineligible population |
| | Kumanyika, Lorrene D. Ritchie,Thomas N. Robinson,Marlene B. Schwartz. | |
| Childhood Obesity Prevention in Africa: A Systematic Review of Intervention Effectiveness and Implementation | Sonja Klingberg, Catherine E. Draper, Lisa K. Micklesfield, Sara E. Benjamin-Neelon and Esther M. F. van Sluijs. | Ineligible population |
| Childhood obesity prevention interventions in childcare settings: systematic review of randomized and nonrandomized controlled trials | Zhou, Y. E., J. S. Emerson, R. S. Levine, C. J. Kihlberg and P. C. Hull | Ineligible population |
| Children's Physical Activity, Academic Performance, and Cognitive Functioning: A Systematic Review and Meta-Analysis | Vedrana Sember, Gregor Jurak, Marjeta Kovac, Shawnda A. Morrison and Gregor Starc | Ineligible population |
| Chronic Adaptations to Eccentric Training: A Systematic Review | Douglas, J., S. Pearson, A. Ross and M. McGuigan | Ineligible population |
| Chronic efects of high‐intensity functional training on motor function: a systematic review with multilevel meta‐analysis | Jan Wilke & Lisa Mohr | Ineligible population |
| Chronic Effects of Altering Resistance Training Set Configurations Using Cluster Sets: A Systematic Review and Meta-Analysis. | Davies, Timothy B.; Tran, Derek L.; Hogan, Clorinda M.; Haff, G. Gregory; Latella, Christopher | Ineligible population |
| Chronic Physiological Effects of Swim Training Interventionsin Non-Elite Swimmers: A Systematic Review and Meta-Analysis | Ian M. Lahart, George S. Metsios | Ineligible population |

| | | |
|---|---|---|
| Classroom-Based Physical Activity and Sedentary Behavior Interventions in Adolescents: A Systematic Review and Meta-Analysis | McMichan, L., A. M. Gibson and D. A. Rowe | Ineligible population |
| Clinical Applications of Yoga for the Pediatric Population: A Systematic Review | Birdee, G. S., G. Y. Yeh, P. M. Wayne, R. S. Phillips, R. B. Davis and P. Gardiner | Ineligible population |
| Clinical review: behavioral interventions to prevent childhood obesity: a systematic review and metaanalyses of randomized trials | Kamath, C. C., K. S. Vickers, A. Ehrlich, L. McGovern, J. Johnson, V. Singhal, R. Paulo, A. Hettinger, P. J. Erwin and V. M. Montori | Ineligible population |
| Clinical review: treatment of pediatric obesity: a systematic review and meta-analysis of randomized trials | McGovern, L., J. N. Johnson, R. Paulo, A. Hettinger, V. Singhal, C. Kamath, P. J. Erwin and V. M. Montori | Ineligible population |
| Combined Diet and Physical Activity Promotion Programs to Prevent Type 2 Diabetes Among Persons at Increased Risk: A Systematic Review for the Community Preventive Services Task Force | Balk, E. M., A. Earley, G. Raman, E. A. Avendano, A. G. Pittas and P. L. Remington | Ineligible population |
| Combined effects of physical activity and calcium on bone health in children and adolescents: a systematic review of randomized controlled trials | Xi Yang, Yi Zhai, Jian Zhang, Jing‑Yi Chen, Dan Liu, Wen‑Hua Zhao. | Ineligible population |
| Combined Home and School Obesity Prevention Interventions for Children: What Behavior Change Strategies and Intervention Characteristics Are Associated With Effectiveness? | Hendrie, G. A., E. Brindal, N. Corsini, C. Gardner, D. Baird and R. K. Golley | Ineligible population |
| Commercial Off-The-Shelf Video Games for Reducing Stress and Anxiety: Systematic Review | Federica Pallavicini; Alessandro Pepe; Fabrizia Mantovani. | Ineligible population |
| Comparative Effectiveness of Physical Activity Intervention Programs on Motor Skills in Children and Adolescents: A Systematic Review and Network Meta-Analysis | Mohamed A. Hassan, Wenxi Liu, Daniel J. McDonough, Xiwen Su and Zan Gao | Ineligible population |
| Comparative effectiveness of school-based interventions targeting physical activity, physical fitness or sedentary behaviour on obesity prevention in 6- to 12-year-old children: A systematic review and meta-analysis | Hrvoje Podnar1 \| Petra Juric1 \| Josip Karuc1 \| Marc Saez2,3 \| | Ineligible population |
| Comparing Active Pediatric Obesity Treatments Using Meta-Analysis | Gilles, A., M. Cassano, E. J. Shepherd, D. Higgins, J. E. Hecker and D. W. Nangle | Ineligible population |
| Comparing the effects of variable and traditional resistance training on maximal strength and muscle power in healthy adults: A systematic review and meta-analysis. | Andersen, Vidar; Prieske, Olaf; Stien, Nicolay; Cumming, Kristoffer; Solstad, Tom Erik Jorung; Paulsen, Gøran; van den Tillaar, Roland; Pedersen, Helene; Saeterbakken, Atle H. | Ineligible population |
| Comparison of the effects of velocity-based vs. traditional resistance training methods on adaptations in strength, power, | Orange, Samuel T.; Hritz, Adam; Pearson, Liam; Jeffries, Owen; Jones, Thomas W.; Steele, James; | Ineligible population |

| | | |
|---|---|---|
| and sprint speed: A systematic review, meta-analysis, and quality of evidence appraisal | | |
| Comparison of Weightlifting, Traditional Resistance Training and Plyometrics on Strength, Power and Speed: A Systematic Review with Meta-Analysis. | Morris, Stephanie J.; Oliver, Jon L.; Pedley, Jason S.; Haff, G. Gregory; Lloyd, Rhodri S. | Ineligible population |
| Considering sex/gender in interventions to promote children’s and adolescents’ leisure-time physical activity: a systematic review and meta-analysis | Carolin Schulze, Jens Bucksch, Yolanda Demetriou, Sandra Emmerling, Stephanie Linder, Anne K Reimers. | Ineligible population |
| Controlling childhood obesity: A systematic review on strategies and challenges | Kelishadi, R. and F. Azizi-Soleiman | Ineligible population |
| Converging Evidence Supporting the Cognitive Link between Exercise and Esport Performance: A Dual Systematic Review | Adam J. Toth, Niall Ramsbottom, Magdalena Kowal and Mark J. Campbell | Ineligible population |
| DE-PASS Best Evidence Statement (BESt) - modifiable determinants, their association with and effect on physical activity and sedentary behaviour in children and adolescents aged 5-19 years: systematic reviews and meta-analyses | Mohammed Khudair et al. | Ineligible population |
| Deoxygenation of inspiratory muscles during cycling, hyperpnoea and loaded breathing in health and disease: a systematic review. | Tanaka, Takako; Basoudan, Nada; Melo, Luana T.; Wickerson, Lisa; Brochard, Laurent J.; Goligher, Ewan C.; Reid, W. Darlene | Ineligible population |
| Design features associated with engagement in mHealth physical activity interventions among youth: A systematic review of qualitative and quantitative studies | Ayla Schwarz et al. | Ineligible population |
| Development of Maximal Dynamic Strength During Concurrent Resistance and Endurance Training in Untrained, Moderately Trained, and Trained Individuals: A Systematic Review and Meta-analysis. | Petré, Henrik; Hemmingsson, Erik; Rosdahl, Hans; Psilander, Niklas | Ineligible population |
| Diet, physical activity and behavioural interventions for the treatment of overweight or obese children from the age of 6 to 11 years | Mead, E., T. Brown, K. Rees, L. B. Azevedo, V. Whittaker, D. Jones, J. Olajide, G. M. Mainardi, E. Corpeleijn, C. O'Malley, E. Beardsmore, L. Al-Khudairy, L. Baur, M. I. Metzendorf, A. Demaio and L. J. Ells | Ineligible population |
| Digital exercise interventions for improving measures of central obesity: a systematic review | Marcel Ballin, Andreas Hult, Sabine Bjo ̈rk John Dinsmore, Peter Nordstro ̈m, Anna Nordstro ̈m. | Ineligible population |
| Digital technology use by and with young children: A systematic review for the Statement on Young Children and Digital Technologies | Ana Mantilla e Susan Edwards | Ineligible population |

| | | |
|---|---|---|
| Do Interventions to Increase Walking Work? A Systematic Review of Interventions in Children and Adolescents | Carlin, A., M. H. Murphy and A. M. Gallagher | Ineligible population |
| Do text messages about health and development in young children affect caregiver behaviour and child outcomes? A systematic review | Brittany Paige Richardson, Jeannie van der Linde, Bhavani Pillay, De wet Swanepoel | Ineligible population |
| Does Aerobic Training Promote the Same Skeletal Muscle Hypertrophy as Resistance Training? A Systematic Review and Meta-Analysis. | Grgic, Jozo; Mcllvenna, Luke C.; Fyfe, Jackson J.; Sabol, Filip; Bishop, David J.; Schoenfeld, Brad J.; Pedisic, Zeljko; | Ineligible population |
| Does Physical Activity Improve Cognition and Academic Performance in Children? A Systematic Review of Randomized Controlled Trials | Camila Ferreira Vorkapic, Heloisa Alves, Larissa Araujo, Claudio Joaquim Borba-Pinheiro, Renato Coelho, Eugenio Fonseca, Ana Oliveira, Estelio H.M. Dantas, | Ineligible population |
| Does physical fitness moderate the effect of physical activity on cardiometabolic health in children?: a systematic review | Anders Husøy et al. | Ineligible population |
| Does the gut microbiota contribute to the antiobesity effect of exercise? A systematic review and meta-analysis. | Zheng, Chen; Chen, Xiang‑Ke; Tian, Xiao Yu; Ma, Alvin Chun‑Hang; Wong, Stephen Heung‑Sang; Chen, Xiang-Ke; Ma, Alvin Chun-Hang; Wong, Stephen Heung-Sang | Ineligible population |
| Efectiveness of physical activity programs in enhancing sleep outcomes among adolescents: a systematic review | Bashir Bello,Jibril Mohammed,Ushotanefe Useh. | Ineligible population |

| | | |
|---|---|---|
| Efects of High-Intensity Interval Training in School on the Physical Performance and Health of Children and Adolescents: A Systematic Review with Meta-Analysis | Nikolai Bauer, Billy Sperlich, Hans‑Christer Holmberg and Florian A. Engel | Ineligible population |
| Efects of Resistance Training on Change‑of‑Direction Speed in Youth and Young Physically Active and Athletic Adults: A Systematic Review with Meta‑Analysis | Helmi Chaabene, Olaf Prieske,Jason Moran,Yassine Negra,Ahmed Attia,Urs Granacher | Ineligible population |
| Effect modification by cardiorespiratory fitness on the association between physical activity and cardiometabolic health in youth: A systematic review | Anders Husøy , Knut Eirik Dalene , Jostein Steene-Johannessen , SigmundAlfred Anderssen , Ulf Ekelund & Jakob Tarp | Ineligible population |
| Effect of Different Exercise Modalities on Oxidative Stress: A Systematic Review | Anand Thirupathi, Meizi Wang, Ji Kai Lin, Gusztáv Fekete, Bíró István, Julien S. Baker, and Yaodong Gu | Ineligible population |
| Effect of eccentric overload training on change of direction speed performance: A systematic review and meta-analysis. | Liu, Ruidong; Liu, Jianxiu; Clarke, Caitlin Vitosky; An, Ruopeng | Ineligible population |
| Effect of exercise on intima-media thickness in children and adolescents with obesity: A meta-analysis | Ngai, S. and R. Cheung | Ineligible population |
| Effect of Exercise on Risk Factors of Diabetic Foot Ulcers: | Fuyuan Liao; Ruopeng An; Fang Pu; Burns, Stephanie; Sa Shen; Yih-Kuen Jan; | Ineligible population |

| | | |
|---|---|---|
| A Systematic Review and Meta-Analysis. | | |
| Effect of Exercise Training on Physical Fitness Among Young Tennis Players: A Systematic Review | Wensheng Xiao, Soh Kim Geok, Xiaorong Bai, Te Bu, Mohd Rozilee Norjali Wazir, Othman Talib, Wenfang Liu and Chongjiang Zhan | Ineligible population |
| Effect of interventions based on regular physical activity on weight management in adolescents: a systematic review and a meta-analysis | Babak Moeini, Forouzan Rezapur-Shahkolai, Saeed Bashiria, Amin Doosti-Irani, Maryam Afshari and Azam Geravandi | Ineligible population |
| Effect of interventions on the body mass index of school-age students | Friedrich, R. R., I. Schuch and M. B. Wagner | Ineligible population |
| Effect of Mobile Health Technology on Weight Control in Adolescents and Preteens: A Systematic Review and Meta-Analysis | Jui-Mei Yien, Hsiu-Hung Wang, Ruey-Hsia Wang, Fan-Hao Chou, Kuo-Hsiung Chen and Fu-Sheng Tsai | Ineligible population |
| Effect of pedometer-based physical activity interventions: a meta-analysis | Kang, M., S. J. Marshall, T. V. Barreira and J. O. Lee | Ineligible population |
| Effect of Physical Education-based stretching programs on hamstring extensibility in high school students: A systematic review. / Efecto de los programas de estiramiento en Educación Física sobre la extensibilidad de la musculatura isquiosural en estudiantes de Educación | Alberto Becerra-Fernández, Carlos; Mayorga-Vega, Daniel; Merino-Marban, Rafael | Ineligible population |

| | | |
|---|---|---|
| Secundaria: Una revisión sistemática. | | |
| Effect of Pilates Intervention on Physical Function of Children and Youth: A Systematic Review. | Hornsby, Elizabeth; Johnston, Leanne M. | Ineligible population |
| Effect of Plyometric Training on Sport Performance in Adolescent Overhead Athletes: A Systematic Review | Leyla Eraslan, Birgit Castelein, Valentien Spanhove, Ceren Orhan, Irem Duzgun, and Ann Cools | Ineligible population |
| Effect of Resistance Training Methods and Intensity on the Adolescent Swimmer’s Performance: A Systematic Review | Wei Guo, Kim Geok Soh, Noor Syamilah Zakaria, Mohamad Taufik Hidayat Baharuldin and Yongqi Gao | Ineligible population |
| Effect of resistance training with and without caloric restriction on visceral fat: A systemic review and meta-analysis | Mousa Khalafi, Abbas Malandish,Sara K. Rosenkranz, Ali A. Ravasi | Ineligible population |
| Effect of sedentary behavior interventions on vascular function in adults: A systematic review and meta‐analysis. | Zheng, Chen; Zhang, Xiaoyuan; Sheridan, Sinead; Ho, Robin Sze‐Tak; Sit, Cindy Hui‐Ping; Huang, Yu; Wong, Stephen Heung‐Sang | Ineligible population |
| Effect of Strength Training on Biomechanical and Neuromuscular Variables in Distance Runners: A Systematic Review and Meta-Analysis. | Trowell, Danielle; Vicenzino, Bill; Saunders, Natalie; Fox, Aaron; Bonacci, Jason | Ineligible population |
| Effective behaviour change techniques in the prevention and management of childhood obesity | Martin, J., A. Chater and F. Lorencatto | Ineligible population |

| | | |
|---|---|---|
| Effective Interventions for Improving Functional Movement Screen Scores Among "High-Risk" Athletes: A Systematic Review. | Clark, Sean C.; Rowe, Nicholas D.; Adnan, Mohamed; Brown, Symone M.; Mulcahey, Mary K. | Ineligible population |
| Effective physical education and school sport: A systematic review of physical education and school sport interventions targeting physical activity, movement skills and enjoyment of physical activity | Dudley, D., A. Okely, P. Pearson and W. Cotton | Ineligible population |
| Effectiveness of Adult Health Promotion Interventions Delivered Through Professional Sport: Systematic Review and Meta-Analysis | George, Emma S.; El Masri, Aymen; Kwasnicka, Dominika; Romeo, Alannah; Cavallin, Sarah; Bennie, Andrew; Kolt, Gregory S.; Guagliano, Justin M. | Ineligible population |
| Effectiveness of centre-based childcare interventions in increasing child physical activity: a systematic review and meta-analysis for policymakers and practitioners | Finch, M., J. Jones, S. Yoong, J. Wiggers and L. Wolfenden | Ineligible population |
| Effectiveness of diet and physical activity interventions among Chinese-origin populations living in high income countries: a systematic review | Jeannette M. Beasley, Janelle M. Wagnild, Tessa M. Pollard, Timothy R. Roberts and Nasima Ahkter | Ineligible population |
| Effectiveness of early care and education center-based interventions for improving cardiovascular fitness in early | Jacob Szeszulskia, Elizabeth Lorenzo, Gabriel Q. Shaibi, Matthew P. Bumana, Sonia Vega-Lópeza,Steven P. Hookerd, Rebecca E. Lee | Ineligible population |
| childhood: A systematic review and meta-analysis | | |
| Effectiveness of educational and lifestyle interventions to prevent paediatric obesity: systematic review and meta-analyses of randomized and non-randomized controlled trials | Gori, D., F. Guaraldi, S. Cinocca, G. Moser, P. Rucci and M. P. Fantini | Ineligible population |
| Effectiveness of intervention on physical activity of children: systematic review and meta-analysis of controlled trials with objectively measured outcomes (EarlyBird 54) | Metcalf, B., W. Henley and T. Wilkin | Ineligible population |
| Effectiveness of intervention programs in schools to reduce health risk factors in adolescents: a systematic review. / Efetividade de programas de intervenÃ§Ã£o escolar para reduzir fatores de risco Ã saÃºde em adolescentes: uma revisÃ£o sistemÃ¡tica | GuimarÃ£es, R. d. F. t., R. D. Langer, G. Guerra-JÃºnior and E. M. GonÃ§alves | Ineligible population |
| Effectiveness of Lifestyle Interventions for Prevention of Harmful Weight Gain among Adolescents from Ethnic Minorities: A Systematic Review | Nematullah Hayba *, Samiha Elkheir, Jessica Hu and Margaret Allman-Farinelli | Ineligible population |
| Effectiveness of lifestyle interventions in child obesity: systematic review with meta-analysis | Ho, M., S. P. Garnett, L. Baur, T. Burrows, L. Stewart, M. Neve and C. Collins | Ineligible population |

| | | |
|---|---|---|
| Effectiveness of Mobile Apps to Promote Health and Manage Disease: Systematic Review and Meta-analysis of Randomized Controlled Trials | Sarah J Iribarren, Tokunbo O Akande, Kendra J Kamp, Dwight Barry,Yazan G Kader, Elizabeth Suelzer | Ineligible population |
| Effectiveness of Physical-Activity-Based Interventions Targeting Overweight and Obesity among University Students—A Systematic Review | Julia Pfisterer, Constantin Rausch, Doreen Wohlfarth, Philip Bachert, Darko Jekauc and Kathrin Wunsch | Ineligible population |
| Effectiveness of school-based eHealth interventions to prevent multiple lifestyle risk behaviours among adolescents: a systematic review and meta-analysis | Katrina E Champion, Belinda Parmenter, Cyanna McGowan, Bonnie Spring, Q Eileen Wafford, Lauren A Gardner, Louise Thornton, Nyanda McBride, Emma L Barrett, Maree Teesson, Nicola C Newton, and The Health Life team* | Ineligible population |
| Effectiveness of school-based interventions to prevent obesity among children aged 4 to 12 years old in middle-income countries: A systematic review and meta-analysis | Juhi Singhal, Clare Herd, Peymane Adab, Miranda Pallan | Ineligible population |
| Effectiveness of school-based physical activity and sedentary time interventions in the prevention of childhood obesity for children from the age of 6 to 12 years: a systematic review and meta-analysis | Hrvoje Podnar, Petra Jurić, Josip Karuc, Marc Saez, Maria A. Barceló, Ivan Radman, Gregor Starc, Gregor Jurak, Saša Đurić, Žan Luca Potočnik, Maroje Sorić | Ineligible population |

| | | |
|---|---|---|
| Effectiveness of shoulder injury prevention programs in an overhead athletic population: A systematic review | Wright AA, Ness BM, Donaldson M, Hegedus EJ, Salamh P, Cleland JA | Ineligible population |
| Effects and Dose–Response Relationship of Balance Training on Balance Performance in Youth: A Systematic Review and Meta-Analysis | Arnd Gebel, Melanie Lesinski, David G. Behm, Urs Granacher. | Ineligible population |
| Effects and dose–response relationship of high-intensity interval training on cardiorespiratory fitness in overweight and obese adults: a systematic review and meta-analysis. | Wang, Kangle; Zhu, Yuxin; Wong, Stephen Heung-Sang; Chen, Yajun; Siu, Parco Ming-Fai; Baker, Julien S.; Sun, Fenghua | Ineligible population |
| Effects of Accumulated Short Bouts of Exercise on Weight and Obesity Indices in Adults: A Meta-Analysis | Heontae Kim, PhD , Joel Reece, PhD, and Minsoo Kang, PhD | Ineligible population |
| Effects of active video games on children and adolescents: A systematic review with meta‑analysis | Crystian B. Oliveira, Rafael Z. Pinto, Bruna T. C. Saraiva, William R. Tebar, Leandro D. Delfino, Marcia R. Franco, Claudiele C. M. Silva, Diego G. D. Christofaro | Ineligible population |
| Effects of Acute and Chronic Exercises on Executive Function in Children and Adolescents: A Systemic Review and Meta-Analysis | Shijie Liu, Qian Yu, Zaimin Li, Paolo Marcello Cunha, Yanjie Zhang,Zhaowei Kong, Wang Lin, Sitong Chen and Yujun Cai | Ineligible population |

| | | |
|---|---|---|
| Effects of Acute Physical Exercise With Low and High Cognitive Demands on Executive Functions in Children: A Systematic Review | Linda Paschen, Tim Lehmann, Miriam Kehne, and Jochen Baumeister | Ineligible population |
| Effects of aerobic exercise on obese children with metabolic syndrome: a systematic | Youxiang Cao, Lin Zhu* and Jingxin Liu | Ineligible population |
| Effects of aerobic, resistance, and combined exercise training on insulin resistance markers in overweight or obese children and adolescents: A systematic review and meta-analysis | Marson, E. C., R. S. Delevatti, A. K. Prado, N. Netto and L. F. Kruel | Ineligible population |
| Effects of Aquatic Exercise on Muscle Strength in Young and Elderly Adults: A Systematic Review and Meta-Analysis of Randomized Trials. | Prado, Alexandre K.G.; Reichert, Thais; Conceição, Matheus O.; Delevatti, Rodrigo S.; Kanitz, Ana C.; Kruel, Luiz F.M.; | Ineligible population |
| Effects of Bilateral and Unilateral Resistance Training on Horizontally Orientated Movement Performance: A Systematic Review and Meta-analysis. | Moran, Jason; Ramirez-Campillo, Rodrigo; Liew, Bernard; Chaabene, Helmi; Behm, David G.; García-Hermoso, Antonio; Izquierdo, Mikel; Granacher, Urs | Ineligible population |
| Effects of chronic exercise interventions on executive function among children and adolescents: a systematic review with meta-analysis | Yue Xue,Yanxiang Yang, Tao Huang | Ineligible population |
| Effects of chronic exercise on the inhibitory control of children and adolescents: A | Sandra Amatriain-Fernández, Milagros Ezquerro | Ineligible population |

| | | |
|---|---|---|
| systematic review and meta-analysis | García-Noblejas, Henning Budde | |
| Effects of Classroom Active Desks on Children and Adolescents' Physical Activity, Sedentary Behavior, Academic Achievements and Overall Health: A Systematic Review | Terry Guirado, Camille Chambonnière, Jean-Philippe Chaput, Lore Metz, David Thivel, and Martine Duclos. | Ineligible population |
| EFFECTS OF COMBAT SPORTS ON BONE MASS: SYSTEMATIC REVIEW. / EFECTO DE LOS DEPORTES DE COMBATE SOBRE LA MASA ÓSEA: REVISIÓN SISTEMÁTICA. | Camila Justino de Oliveira Barbeta, Tathyane Krahenbühl, Ezequiel Moreira Gonçalves, Gil Guerra-Júnior | Ineligible population |
| Effects of community health worker-based interventions on physical activity levels in children: a systematic review | Paulo Henrique Guerra, Rafael Silvestre, Tiago Henrique Toldo de Mello, Ana Luiza Babo Sedlacek Carvalho, Filipe Ferreira da Costa, Alex Antonio Florindo | Ineligible population |
| Effects of CrossFit intervention on students' physical fitness in physical education: a systematic review and meta-analysis. | YANKUN HAN; BIN SYED ALI, SYED KAMARUZAMAN; LIFU JI | Ineligible population |
| Effects of different physical activities on brain-derived neurotrophic factor: A systematic review and bayesian network meta-analysis | Bojun Zhou, Zhisheng Wang, Lianghao Zhu, Gang Huang, Bing Li, Chaofan Chen, Junda Huang, Fuhai Ma and Timon Chengyi Liu | Ineligible population |
| Effects of exercise interventions on body image: a meta-analysis | Campbell, A. and H. A. Hausenblas | Ineligible population |

| | | |
|---|---|---|
| Effects of exercise motivations on body image and eating habits/behaviours: A systematic review | Inês Panão, Eliana V. Carraça | Ineligible population |
| Effects of exercise on resting blood pressure in obese children: a meta-analysis of randomized controlled trials | Garcia-Hermoso, A., J. M. Saavedra and Y. Escalante | Ineligible population |
| Effects of Exercise on the Resting Heart Rate: A Systematic Review and Meta-Analysis of Interventional Studies | Anne Kerstin Reimers ,Guido Knapp and Carl-Detlev Reimers | Ineligible population |
| Effects of Exercise Training on Resting Testosterone Concentrations in Insufficiently Active Men: A Systematic Review and Meta-Analysis | Nolan J. Potter, Grant R. Tomkinson,Trevor J. Dufner, Tanis J. Walch, James N. Roemmich, Patrick B. Wilson, and John S. Fitzgerald | Ineligible population |
| Effects of flywheel training on strength-related variables in female populations. A systematic review. | Raya-González, Javier; de Keijzer, Kevin L.; Bishop, Chris; Beato, Marco | Ineligible population |
| Effects of Heat Acclimation and Acclimatisation on Maximal Aerobic Capacity Compared to Exercise Alone in Both Thermoneutral and Hot Environments: A Meta-Analysis and Meta-Regression. | Waldron, Mark; Fowler, Rebecca; Heffernan, Shane; Tallent, Jamie; Kilduff, Liam; Jeffries, Owen | Ineligible population |
| Effects of high-intensity interval training in men soccer player's physical fitness: A systematic review with meta-analysis of | Filipe Manuel Clemente, Rodrigo Ramirez-Campillo, Fábio Yuzo Nakamura & Hugo Sarmento | Ineligible population |

| | | |
|---|---|---|
| randomized-controlled and non-controlled trials | | |
| EFFECTS OF HIGH-INTENSITY INTERVAL TRAINING ON OLYMPIC COMBAT SPORTS ATHLETES' PERFORMANCE AND PHYSIOLOGICAL ADAPTATION: A SYSTEMATIC REVIEW | FRANCHINI, EMERSON; CORMACK, STUART; TAKITO, MONICA Y. | Ineligible population |
| Effects of In-Classroom Physical Activity Breaks on Children's Academic Performance, Cognition, Health Behaviours and Health Outcomes: A Systematic Review and Meta-Analysis of Randomised Controlled Trials | D. L. I. H. K. Peiris, Yanping Duan, Corneel Vandelanotte, Wei Liang, Min Yang and Julien Steven Baker | Ineligible population |
| Effects of interrupting prolonged sitting on postprandial glycemia and insulin responses: A network meta-analysis. | Quan, Minghui; Xun, Pengcheng; Wu, Hua; Wang, Jing; Cheng, Wei; Cao, Meng; Zhou, Tang; Huang, Tao; Gao, Zan; Chen, Peijie | Ineligible population |
| Effects of isometric, eccentric, or heavy slow resistance exercises on pain and function in individuals with patellar tendinopathy: A systematic review. | Lim, Hui Yin; Wong, Shi Hui | Ineligible population |
| Effects of jump training on physical fitness and athletic performance in endurance runners: A meta-analysis: Jump training in endurance runners. | Ramirez-Campillo, Rodrigo; Andrade, David C.; García-Pinillos, Felipe; Negra, Yassine; Boullosa, Daniel; Moran, Jason | Ineligible population |

| | | |
|---|---|---|
| Effects of Lifestyle Modification Interventions to Prevent and Manage Child and Adolescent Obesity: A Systematic Review and Meta-Analysis | Rehana A. Salam, Zahra A. Padhani, Jai K. Das, Amina Y. Shaikh, Zahra Hoodbhoy, Sarah Masroor Jeelani, Zohra S. Lassi and Zulfiqar A. Bhutta | Ineligible population |
| Effects of Multicomponent Injury Prevention Programs on Children and Adolescents' Fundamental Movement Skills: A Systematic Review With Meta-Analyses | John A. Jimenez-Garcia, Matthew B. Miller, Richard G. DeMont. | Ineligible population |
| Effects of Open versus Closed Skill Exercise on Cognitive Function: A Systematic Review | Qian Gu, Liye Zou, Paul D. Loprinzi, Minghui Quan, Tao Huang | Ineligible population |
| Effects of physical education interventions on cognition and academic performance | Antonio García-Hermoso ,1,2 Robinson Ramírez-Vélez ,1 | Ineligible population |
| Effects of physical exercise on myelin sheath regeneration: A systematic review and meta-analysis. | Feter, N.; Freitas, M.P.; Gonzales, N.G.; Umpierre, D.; Cardoso, R.K.; Rombaldi, A.J. | Ineligible population |
| Effects of Plyometric Jump Training on Balance Performance in Healthy Participants: A Systematic Review With Meta-Analysis | Akhilesh Kumar Ramachandran, Utkarsh Singh, Rodrigo Ramirez-Campillo, Filipe Manuel Clemente, José Afonso, Urs Granacher. | Ineligible population |
| Effects of Plyometric Jump Training on Electromyographic Activity and Its Relationship to Strength and Jump Performance in Healthy Trained and Untrained Populations: A Systematic Review of Randomized Controlled Trials | Ramirez-Campillo R, Garcia-Pinillos F, Chaabene H, Moran J, Behm DG, Granacher U | Ineligible population |
| Effects of Plyometric Jump Training on Jump and Sprint Performance in Young Male Soccer Players: A Systematic Review and Meta-analysis. | Ramirez-Campillo, Rodrigo; Castillo, Daniel; Raya-González, Javier; Moran, Jason; de Villarreal, Eduardo Sáez; Lloyd, Rhodri S. | Ineligible population |
| Effects of Plyometric Jump Training on Repeated Sprint Ability in Athletes: A Systematic Review and Meta-Analysis. | Ramirez-Campillo, Rodrigo; Gentil, Paulo; Negra, Yassine; Grgic, Jozo; Girard, Olivier | Ineligible population |
| Effects of plyometric jump training on soccer player's balance: a systematic review and meta-analysis of randomized-controlled trials | Clemente, Filipe Manuel; Ramirez-Campillo, Rodrigo; Castillo, Daniel; Raya-González, Javier; Rico-González, Markel; Oliveira, Rafael; Rosemann, Thomas; Knechtle, Beat; | Ineligible population |
| Effects of Plyometric Jump Training on Vertical Jump Height of Volleyball Players: A Systematic Review with Meta-Analysis of Randomized-Controlled Trial. | Ramirez-Campillo, Rodrigo; Andrade, David C.; Nikolaidis, Pantelis T.; Moran, Jason; Clemente, Filipe Manuel; Chaabene, Helmi; Comfort, Paul | Ineligible population |
| Effects of plyometric training on skill and physical performance in healthy tennis players: A systematic review and meta-analysis | Nuannuan Deng, Kim Geok Soh, Dandan Huang, Borhannudin Abdullah, Shengyao Luo,Watnawat Rattanakoses | Ineligible population |
| Effects of Pokémon GO on Physical Activity and Psychological and Social | Jung Eun Lee, Nan Zeng, Yoonsin Oh, Daehyoung Lee and Zan Gao | Ineligible population |

| | | |
|---|---|---|
| Outcomes: A Systematic Review | | |
| Effects of Regular Physical Activity on the Immune System, Vaccination and Risk of Community-Acquired Infectious Disease in the General Population: Systematic Review and Meta-Analysis. | Chastin, Sebastien F. M.; Abaraogu, Ukachukwu; Bourgois, Jan G.; Dall, Philippa M.; Darnborough, Jennifer; Duncan, Elaine; Dumortier, Jasmien; Pavón, David Jiménez; McParland, Joanna; Roberts, Nicola J.; Hamer, Mark | Ineligible population |
| Effects of resistance training in children and adolescents: a meta-analysis | Behringer, M., A. Vom Heede, Z. Yue and J. Mester | Ineligible population |
| Effects of Resistance Training on Arterial Stiffness in Healthy People: A Systematic Review. | García-Mateo, Pablo; García-de-Alcaraz, Antonio; Rodríguez-Peréz, Manuel A.; Alcaraz-Ibáñez, Manuel | Ineligible population |
| Effects of Resistance Training Performed to Failure or Not to Failure on Muscle Strength, Hypertrophy, and Power Output: A Systematic Review With Meta-Analysis. | Vieira, Alexandra F.; Umpierre, Daniel; Teodoro, Juliana L.; Lisboa, Salime C.; Baroni, Bruno M.; Izquierdo, Mikel; Cadore, Eduardo L.; | Ineligible population |
| Effects of resistance training performed to repetition failure or non-failure on muscular strength and hypertrophy: A systematic review and meta-analysis. | Grgic, Jozo; Schoenfeld, Brad J.; Orazem, John; Sabol, Filip | Ineligible population |
| Effects of school-based before-school physical activity programmes on children's | James Woodforde ,1 | Ineligible population |

| | | |
|---|---|---|
| Effects of school-based neuromuscular training on fundamental movement skills and physical fitness in children: a systematic review | Junlei Lin, Ruofei Zhang, Jie Shen and Aiguo Zhou | Ineligible population |
| Effects of Strength Training on Blood Pressure and Heart Rate Variability—A Systematic Review | Marcelo Corso, Tiago C. de Figueiredo, Danilo Carvalho, Amanda F. Brown, Belmiro F. de Salles, Roberto Simão, Jeffrey M. Willardson, Ingrid Dias | Ineligible population |
| Effects of Strength Training on the Physiological Determinants of Middle- and Long-Distance Running Performance: A Systematic Review. | Blagrove, Richard C.; Howatson, Glyn; Hayes, Philip R. | Ineligible population |
| Effects of Strength Training Using Unstable Surfaces on Strength, Power and Balance Performance Across the Lifespan: A Systematic Review and Meta-analysis | Behm, D. G., T. Muehlbauer, A. Kibele and U. Granacher | Ineligible population |
| Effects of Taekwondo Training on Body Composition: A Systematic Review and Meta-Analysis | Seunghui Baek, Jong-Beom Park, Sang-Hwan Choi, Jae-Don Lee,Sang-Seok Nam. | Ineligible population |
| Effects of tapering on neuromuscular and metabolic fitness in team sports: a systematic review and meta-analysis. | VACHON, ADRIEN; BERRYMAN, NICOLAS; MUJIKA, IÑIGO; PAQUET, JEAN-BAPTISTE; ARVISAIS, DENIS; BOSQUET, LAURENT | Ineligible population |

| | | |
|---|---|---|
| Effects of Vertically and Horizontally Orientated Plyometric Training on Physical Performance: A Meta-analytical Comparison. | Moran, Jason; Ramirez-Campillo, Rodrigo; Liew, Bernard; Chaabene, Helmi; Behm, David G.; García-Hermoso, Antonio; Izquierdo, Mikel; Granacher, Urs | Ineligible population |
| Effects of Vest and Sled Resisted Sprint Training on Sprint Performance in Young Soccer Players: A Systematic Review and Meta-analysis. | Fernández-Galván, Luis Miguel; Casado, Arturo; García-Ramos, Amador; Haff, Guy Gregory; | Ineligible population |
| Effects of weight-bearing activities on bone mineral content and density in children and adolescents: a meta-analysis | Behringer, M., S. Gruetzner, M. McCourt and J. Mester | Ineligible population |
| Effects of weight-bearing exercise on bone health in girls: a meta-analysis | Ishikawa, S., Y. Kim, M. Kang and D. W. Morgan | Ineligible population |
| Effects of weight-neutral approaches compared with traditional weight-loss approaches on behavioral, physical, and psychological health outcomes: a systematic review and meta-analysis. | Dugmore, Jaslyn A; Winten, Copeland G; Niven, Hannah E; Bauer, Judy; | Ineligible population |
| Effects of Workplace-Based Physical Activity Interventions on Cardiorespiratory Fitness: A Systematic Review and Meta-Analysis of Controlled Trials. | Burn, Naomi L.; Weston, Matthew; Maguire, Neil; Atkinson, Greg; Weston, Kathryn L. | Ineligible population |
| Efficacy of dietary intervention or in combination with exercise on primary prevention of cardiovascular disease: A systematic review | Manuela Abbate, Laura Gallardo Alfaro, Maria del Mar Bibiloni, Josep A. Tur | Ineligible population |
| Efficacy of hamstring stretching programs in schoolchildren. A systematic review | Becerra Fernandez, C.-A. and R. Merino-Marban | Ineligible population |
| Efficacy of physical activity interventions on psychological outcomes in refugee, asylum seeker and migrant populations: A systematic review and meta-analysis. | Purgato, Marianna; Richards, Justin; Prina, Eleonora; Kip, Ahlke; Del Piccolo, Lidia; Michencigh, Giulia; Rimondini, Michela; Rudi, Doriana; Vitali, Francesca; Carta, Mauro Giovanni; Morina, Nexhmedin; Schena, Federico; Barbui, Corrado | Ineligible population |
| Efficacy of school-based interventions for improving muscular fitness outcomes in children: A systematic review and meta-analysis | Emilio Villa-González, Yaira Barranco-Ruiz, Antonio García-Hermoso & Avery | Ineligible population |
| Eficacy of School‐Based Interventions for Improving Muscular Fitness Outcomes in Adolescent Boys: A Systematic Review and Meta‐analysis | Ashley Cox, Stuart J. Fairclough, Maria‐Christina Kosteli, Robert J. Noonan | Ineligible population |
| Electronic media-based health interventions promoting behavior change in youth: a systematic review | Hieftje, K., E. J. Edelman, D. R. Camenga and L. E. Fiellin | Ineligible population |
| Engaging parents to increase youth physical activity a systematic review | O'Connor, T. M., R. Jago and T. Baranowski | Ineligible population |

| | | |
|---|---|---|
| Entrenamiento de fuerza con bandas elásticas en niños y adolescentes: una revisión sistemática. / Resistance training using elastic band in children and adolescents. A systematic review. | Lagares Vázquez, Belén; Antonio Rebollo, José | Ineligible population |
| Entrenamiento de la capacidad aerobia en prepúberes. Revisión sistemática. / Trainability of aerobic capacity in prepubescent: Systematic review. | Flores, L. A.; De León, L. G.; Jiménez-Ponce, B. P.; | Ineligible population |
| Environmental interventions to promote healthier eating and physical activity behaviours in institutions: a systematic review | Anneliese M Shaw, Stephen A Wootton, Joanne L Fallowfield, Adrian J Allsopp and Emma L Parsons | Ineligible population |
| Equity effects of children's physical activity interventions: a systematic scoping review | Love, R. E., J. Adams and E. M. F. van Sluijs | Ineligible population |
| Evaluating the Effectiveness of Gamification on Physical Activity: Systematic Review and Meta-analysis of Randomized Controlled Trials | Alexandre Mazeas, Martine Duclos, Bruno Pereira, Aïna Chalabaev. | Ineligible population |
| Evaluation of feasibility, effectiveness, and sustainability of school-based physical activity "active break" interventions in pre-adolescent and adolescent students: a systematic review | Alice Masini & Andrea Ceciliani & Laura Dallolio & Davide Gori & Sofia Marini | Ineligible population |
| Evaluation of school-based interventions including extracurricular activities and homeworks promoting healthy lifestyle (physical activity, healthy diet and sleep hygiene) in children and adolescents: feasibility, effectiveness and sustainability over time. A systematic review. | Alice Masini, Aurelia Salussolia, Alessandra Anastasia, Alberto Grao-Cruces, Giorgia Soldà, Giorgia Zanutto, Stephan Riegger, Raffaela Mulato, David Sánchez-Oliva, Andrea Ceciliani, Sofia Marini & Laura Dallolio | Ineligible population |
| Evaluation of school-based interventions of active breaks in primary schools: A systematic review and meta-analysis | Alice Masini, Sofia Marini, Davide Gori , Erica Leoni , Andrea Rochira, Laura Dallolio | Ineligible population |
| Exercise and BMI in Overweight and Obese Children and Adolescents: A Systematic Review and Trial Sequential Meta-Analysis | Kelley, G. A., K. S. Kelley and R. R. Pate | Ineligible population |
| Exercise and BMI z-score in Overweight and Obese Children and Adolescents: A Systematic Review and Network Meta-Analysis of Randomized Trials | Kelley, G. A., K. S. Kelley and R. R. Pate | Ineligible population |
| Exercise and insulin resistance in youth: a meta-analysis | Fedewa, M. V., N. H. Gist, E. M. Evans and R. K. Dishman | Ineligible population |
| Exercise and resting blood pressure in children and adolescents: A meta-analysis | Kelley, G. A. and K. S. Kelley | Ineligible population |
| Exercise attenuates bone mineral density loss during diet-induced weight loss in adults with overweight and | Mesinovic, Jakub; Jansons, Paul; Zengin, Ayse; de Courten, Barbora; Rodriguez, Alexander | Ineligible population |

| | | |
|---|---|---|
| obesity: A systematic review and meta-analysis. | J.; Daly, Robin M.; Ebeling, Peter R.; Scott, David; | |
| Exercise in school Physical Education increase bone mineral content and density: systematic review and meta-analysis | Júlio B. Mello, Augusto Pedretti, Antonio García-Hermoso, Clarice M. L. Martins, Anelise R. Gaya, Michael J. Duncan & Adroaldo C. A. Gaya | Ineligible population |
| Exercise programs may be effective in preventing a new episode of neck pain: a systematic review and meta-analysis. | De Campos, Tarcisio F; Maher, Chris G; Steffens, Daniel; Fuller, Joel T; Hancock, Mark J | Ineligible population |
| Exercise Timing in Type 2 Diabetes Mellitus: A Systematic Review. | TEO, SHAUN Y. M.; KANALEY, JILL A.; GUELFI, KYM J.; COOK, SUMMER B.; HEBERT, JEFFREY J.; FORREST, MITCHELL R. L.; FAIRCHILD, TIMOTHY J.; | Ineligible population |
| Exercise to improve pediatric bone and fat: a systematic review and meta-analysis | Nogueira, R. C., B. K. Weeks and B. R. Beck | Ineligible population |
| Exercise to improve self-esteem in children and young people | Ekeland, E., F. Heian, K. B. Hagen, J. Abbott and L. Nordheim | Ineligible population |
| Exercise-Based Strategies to Prevent Muscle Injury in Elite Footballers: A Systematic Review and Best Evidence Synthesis. | Fanchini, Maurizio; Steendahl, Ida Bo; Impellizzeri, Franco M.; Pruna, Ricard; Dupont, Gregory; Coutts, Aaron J.; Meyer, Tim; McCall, Alan | Ineligible population |
| Exergame-based exercise training for depressive symptoms in adults: A | Kexin Huang, Yawei Zhao, Rendong He, Tangsheng Zhong, Haiqi Yang, Yuanxin Chen, | Ineligible population |

| | | |
|---|---|---|
| systematic review and meta-analysis | Zixin Liu, Linya Ma, Yong Jia, Li Chen | |
| Exploring the Relationship Between Fundamental Motor Skill Interventions and Physical Activity Levels in Children: A Systematic Review and Meta-analysis | Engel, A. C., C. R. Broderick, N. van Doorn, L. L. Hardy and B. J. Parmenter | Ineligible population |
| Exploring the Relationship Between Fundamental Motor Skill Interventions and Physical Activity Levels in Children: A Systematic Review and Meta-analysis. | Engel, Alexander C.; Broderick, Carolyn R.; van Doorn, Nancy; Hardy, Louise L.; Parmenter, Belinda J. | Ineligible population |
| Family sports interventions for the treatment of obesity in childhood: a meta-analysis | Lihong Yang, Chao Liang, Yaona Yu, Qian Xiao, Maomao Xiao and Lixu Tang | Ineligible population |
| Family-based child weight management intervention in early childhood in low-income families: A systematic review | Kisook Kim and Yoonyoung Lee | Ineligible population |
| Family-based childhood obesity prevention interventions: a systematic review and quantitative content analysis | Ash, T., A. Agaronov, T. Young, A. Aftosmes-Tobio and K. K. Davison | Ineligible population |
| Family-based interventions for reducing sedentary time in youth: a systematic review of randomized controlled trials | Marsh, S., L. S. Foley, D. C. Wilks and R. Maddison | Ineligible population |
| Family-Based Interventions Targeting Childhood Obesity: A Meta-Analysis | Berge, J. M. and J. C. Everts | Ineligible population |

| | | |
|---|---|---|
| Family-Centered Interventions for Treatment and Prevention of Childhood Obesity in | Milicen C. Tamayo1,2 · Page D. Dobbs2,3 · Yair Pincu2,4 | Ineligible population |
| Football can tackle type 2 diabetes: a systematic review of the health effects of recreational football practice in individuals with prediabetes and type 2 diabetes. | Barbosa, Ana; Brito, João; Figueiredo, Pedro; Seabra, André; Mendes, Romeu | Ineligible population |
| Getting patients to exercise more: a systematic review of underserved populations | Carroll, J. K., K. Fiscella, R. M. Epstein, P. Jean-Pierre, C. Figueroa-Moseley, G. C. Williams, K. M. Mustian and G. R. Morrow | Ineligible population |
| Ghrelin Response to Acute and Chronic Exercise: Insights and Implications from a Systematic Review of the Literature | Ouerghi, Nejmeddine; Feki, Moncef; Bragazzi, Nicola Luigi; Knechtle, Beat; Hill, Lee; Nikolaidis, Pantelis T.; Bouassida, Anissa | Ineligible population |
| Golf and Physical Health: A Systematic Review | Sorbie, Graeme G.; Beaumont, Alexander J.; Williams, Ashley K.; Lavallee, David | Ineligible population |
| Healthier Minds in Fitter Bodies: A Systematic Review and Meta‑Analysis of the Association between Physical Fitness and Mental Health in Youth | Cristina Cadenas‑Sanchez, Alejandra Mena‑Molina, Lucia V. Torres‑Lopez, Jairo H. Migueles, María Rodriguez‑Ayllon, David R. Lubans, Francisco B. Ortega | Ineligible population |
| High-intensity interval exercise versus moderate-intensity continuous exercise on postprandial glucose and insulin responses: A systematic review and meta-analysis | Khalafi M, Mojtahedi S, Ostovar A, Rosenkranz SK, Korivi M | Ineligible population |

| | | |
|---|---|---|
| High-intensity interval training for improving health-related fitness in adolescents: a systematic review and meta-analysis | Costigan, S. A., N. Eather, R. C. Plotnikoff, D. R. Taaffe and D. R. Lubans | Ineligible population |
| High-Intensity Interval Training Performed by Young Athletes: A Systematic Review and Meta-Analysis | Engel, F. A., A. Ackermann, H. Chtourou and B. Sperlich | Ineligible population |
| High-Intensity Interval Training Performed by Young Athletes: A Systematic Review and Meta-Analysis | Florian Azad Engel, Alexander Ackermann, Hamdi Chtourou, Billy Sperlich. | Ineligible population |
| High-Intensity Interval Training upon Cognitive and Psychological Outcomes in Youth: A Systematic Review | Ana R. Alves, Renata Dias, Henrique P. Neiva, Daniel A. Marinho, Mário C. Marques, António C. Sousa, Vânia Loureiro and Nuno Loureiro | Ineligible population |
| HIIT, RESISTANCE TRAINING, AND RISK FACTORS IN ADOLESCENTS: A SYSTEMATIC REVIEW. / HIIT, ENTRENAMIENTO RESISTIDO Y FACTORES DE RIESGO EN ADOLESCENTES: UNA REVISIÓN SISTEMÁTICA | Waynne Ferreira de Faria, Filipe Rodrigues Mendonça, Rui Gonçalves Marques Elias, Raphael Gonçalves de Oliveira, Antonio Stabelini Neto | Ineligible population |
| Home- and Community-Based Interventions for Physical Activity and Early Child Development: A Systematic Review of Effective Strategies | Samantha Moss and Xiangli Gu | Ineligible population |

| | | |
|---|---|---|
| How effective are physical activity intervention programmes in children? Systematic review and metaanalysis of controlled trials with objectively-measured outcomes | Metcalf, B., W. Henley and T. Wilkin | Ineligible population |
| Hybridizing pedagogical models: A systematic review | Sixto González-Víllora, Carlos Evangelio, Jacob Sierra-Díaz, Javier Fernández-Río | Ineligible population |
| Impact of isolated aerobic exercise in obese adolescents: systematic review | Fragnani, S., A. GonzÃ¡les, R. Lemos and P. c. Haas | Ineligible population |
| Impact of non-diet approaches on attitudes, behaviors, and health outcomes: a systematic review | Clifford, D., A. Ozier, J. Bundros, J. Moore, A. Kreiser and M. N. Morris | Ineligible population |
| Impact of physical activity interventions on anthropometric outcomes: systematic review and meta-analysis | Conn, V. S., A. Hafdahl, L. J. Phillips, T. M. Ruppar and J. A. Chase | Ineligible population |
| Impact of physical activity/ structured exercise programs on bone health in pre-school children: a systematic review | Callum McCaskie et al. | Ineligible population |
| Impact of training modes on fitness and body composition in women with obesity: A systematic review and meta-analysis. | Davis, Mary E.; Blake, Catherine; Perrotta, Carla; Cunningham, Caitriona; O'Donoghue, Gráinne | Ineligible population |
| Impact of weight loss interventions on patient‑reported outcomes in overweight and obese adults | Jordan Martenstyn, Madeleine King, Claudia Rutherford | Ineligible population |

| | | |
|---|---|---|
| with type 2 diabetes: a systematic review | | |
| Improvement of the lipid profile with exercise in obese children: a systematic review | Escalante, Y., J. M. Saavedra, A. Garcia-Hermoso and A. M. Dominguez | Ineligible population |
| Improving cognition in school children and adolescents through exergames. A systematic review and practical guide | Sebastián López-Serrano, Alberto Ruiz-Ariza, Manuel De La Torre-Cruz, Emilio J. Martínez-López. | Ineligible population |
| Improving youth physical, mental and social health through physical activity: A | Paul N. Kemel \| Joanne E. Porter \| Nicole Coombs | Ineligible population |
| Increasing activity to reduce obesity in adolescent girls: a research review | Clemmens, D. and L. L. Hayman | Ineligible population |
| Individualization and effectiveness of mobile health interventions to reduce physical inactivity and sedentary behavior in healthy children and adolescents: A systematic review and meta-analysis | Hannes Baumann et al. | Ineligible population |
| Influence of chronic stretching on muscle performance: Systematic review | Medeiros, D. M. and C. S. Lima | Ineligible population |
| Influence of exclusive resistance training on body composition and cardiovascular risk factors in overweight or obese children: a systematic review | Dietz, P., S. Hoffmann, E. Lachtermann and P. Simon | Ineligible population |

| | | |
|---|---|---|
| Influence of resistance training load on measures of skeletal muscle hypertrophy and improvements in maximal strength and neuromuscular task performance: A systematic review and meta-analysis. | Refalo, Martin C.; Hamilton, D. Lee; Paval, D. Robert; Gallagher, Iain J.; Feros, Simon A.; Fyfe, Jackson J. | Ineligible population |
| Intervention strategies for enhancing movement competencies in youth athletes: A narrative systematic review. | Rogers, Simon A; Hassmén, Peter; Alcock, Alison; Gilleard, Wendy L; Warmenhoven, John S | Ineligible population |
| Interventions addressing general parenting to prevent or treat childhood obesity | Gerards, S. M., E. F. Sleddens, P. C. Dagnelie, N. K. de Vries and S. P. Kremers | Ineligible population |
| Interventions for increasing the moderate-vigorous intensity physical activity (MVPA) content of primary school physical education | Lan S. Wong, Ann-Marie Gibson, Abdulaziz Farooq, John J.Reilly | Ineligible population |
| Interventions for preventing obesity in childhood. A systematic review | Campbell, K., E. Waters, S. O'Meara and C. Summerbell | Ineligible population |
| Interventions for preventing obesity in children | Campbell, K., E. Waters, S. O'Meara, S. Kelly and C. Summerbell | Ineligible population |
| Interventions for preventing obesity in children | Spinola e Castro, A. M. | Ineligible population |
| Interventions for promoting physical activity among European teenagers: a systematic review | De Meester, F., F. J. van Lenthe, H. Spittaels, N. Lien and I. De Bourdeaudhuij | Ineligible population |
| Interventions on children's and adolescents' physical activity and sedentary behaviour: protocol for a systematic review from a sex/gender perspective | Carolin Schulze et al. | Ineligible population |
| Interventions targeting working memory in 4–11 year olds within their everyday contexts: A systematic review | Anita Rowea, Jill Titterington, Joni Holmes, Lucy Henry, Laurence Taggart | Ineligible population |
| Interventions that impact weight status in Hispanic preschool children | Nancy Innella PhD, RN, CNE \| Beth E. Jameson PhD, RN | Ineligible population |
| Interventions to Change School Recess Activity Levels in Children and Adolescents: A Systematic Review and Meta‑Analysis | Anne‑Maree Parrish, Kar Hau Chong, Abbe L. Moriarty, Marijka Batterham, Nicola D. Ridgers | Ineligible population |
| Interventions to Increase Moderate-to-Vigorous Physical Activity in Elementary School Physical Education Lessons: Systematic Review | LAN S. WONG, MEda ANN-MARIE GIBSON, PhDb ABDULAZIZ FAROOQ, MSc, MPHc JOHN J. REILLY, PhDa, | Ineligible population |
| Interventions to Increase Moderate-to-Vigorous Physical Activity in Elementary School Physical Education Lessons: Systematic Review | Lan S. Wong, Ann-Marie Gibson, Abdulaziz Farooq, John J.Reilly | Ineligible population |
| Interventions to prevent weight gain: a systematic review of psychological models and behaviour change methods | Hardeman, W., S. Griffin, M. Johnston, A. L. Kinmonth and N. J. Wareham | Ineligible population |
| Interventions to Promote Physical Activity among Young | Camacho-Minano, M. J., N. M. LaVoi and D. J. Barr-Anderson | Ineligible population |

| | | |
|---|---|---|
| and Adolescent Girls: A Systematic Review | | |
| Interventions to Promote Physical Activity and Healthy Digital Media Use in Children and Adolescents: A Systematic Review | Christina Oh, a Bianca Carducci,Tyler Vaivada, Zulfiqar A. Bhutta. | Ineligible population |
| Interventions to promote physical activity in young people conducted in the hours immediately after school: a systematic review | Atkin, A. J., T. Gorely, S. J. Biddle, N. Cavill and C. Foster | Ineligible population |
| Is ankle plantar flexor strength associated with balance in healthy adults? A systematic review and meta-analysis. | Hill, K.; Jaberzadeh, S.; Jansons, P.; Malliaras, P.; Perraton, L.; Soh, S.-E.; Tavakkoli Osko, S.; | Ineligible population |
| Is Cumulative Load Associated with Injuries in Youth Team Sport? A Systematic Review | Katie Snifen, Kemba Noel‐London, Melody Schaefer and Oluwatoyosi Owoeye. | Ineligible population |
| Is exercise a senolytic medicine? A systematic review | Xiang-Ke Chen, Zhen-Ni Yi, Gordon Tin-Chun Wong, Kazi Md. Mahmudul Hasan, Joseph Shiu-Kwong Kwan, Alvin Chun-Hang Ma, Raymond Chuen-Chung Chang | Ineligible population |
| Is inertial flywheel resistance training superior to gravity-dependent resistance training in improving muscle strength? A systematic review with meta-analyses. | Vicens-Bordas, J.; Esteve, E.; Fort-Vanmeerhaeghe, A.; Bandholm, T.; Thorborg, K. | Ineligible population |
| Is School Gardening Combined with Physical Activity Intervention Effective for Improving Childhood Obesity? A Systematic Review and Meta-Analysis | Yufei Qi, Sareena Hanim Hamzah, Erya Gu, Haonan Wang, Yue Xi, Minghui Sun, Siyu Rong and Qian Lin | Ineligible population |
| Isometric training and long‐term adaptations: Effects of muscle length, intensity, and intent: A systematic review. | Oranchuk, Dustin J.; Storey, Adam G.; Nelson, André R.; Cronin, John B. | Ineligible population |
| Lifestyle interventions for youth who are overweight: a meta-analytic review | Kitzmann, K. M., W. T. Dalton, 3rd, C. M. Stanley, B. M. Beech, T. P. Reeves, J. Buscemi, C. J. Egli, H. L. Gamble and E. L. Midgett | Ineligible population |
| Long-Term Dietary and Physical Activity Interventions in the School Setting and Their Effects on BMI in Children Aged 6–12 Years: Meta-Analysis of Randomized Controlled Clinical Trials | Purificación Cerrato-Carretero, Raúl Roncero-Martín, Juan D. Pedrera-Zamorano, Fidel López-Espuela, Luis M. Puerto-Parejo, Antonio Sánchez-Fernández, María Luz Canal-Macías, Jose M. Moran and Jesus M. Lavado-García | Ineligible population |
| Longitudinal changes in moderate‐to‐vigorous‐intensity physical activity in children and adolescents: A systematic review and meta‐analysis | Abdulaziz Farooq, Anne Martin, Xanne Janssen, Mathew G. Wilson, Ann‐Marie Gibson, Adrienne Hughes, John J. Reilly | Ineligible population |
| Magnitude of Muscle Strength and Mass Adaptations Between High-Load Resistance Training Versus Low-Load Resistance Training Associated with Blood-Flow Restriction: A | Lixandrão, Manoel; Ugrinowitsch, Carlos; Berton, Ricardo; Vechin, Felipe; Conceição, Miguel; Damas, Felipe; Roschel, Hamilton; Libardi, Cleiton | Ineligible population |

| | | |
|---|---|---|
| Systematic Review and Meta-Analysis. | | |
| Meta-analysis and systematic review of physical activity on neurodevelopment disorders, depression, and obesity among children and adolescents | Sanying Peng, Yuan Fang, Ahmad Tajuddin Othman and Jinghong Liang | Ineligible population |
| Meta-analysis of interventions to prevent overweight and obesity in children and adults | Luckner, H., C. A. Gericke and J. Moss | Ineligible population |
| Mobile Health Interventions for Improving Health Outcomes in Youth: A Meta-analysis | Fedele, D. A., C. C. Cushing, A. Fritz, C. M. Amaro and A. Ortega | Ineligible population |
| Moderators of environmental intervention effects on diet and activity in youth | Kremers, S. P., G. J. de Bruijn, M. Droomers, F. van Lenthe and J. Brug | Ineligible population |
| Moderators of Resistance Training Effects in Overweight and Obese Adults: A Systematic Review and Meta-analysis | LOPEZ, PEDRO; RADAELLI, RÉGIS; TAAFFE, DENNIS R.; GALVÃO, DANIEL A.; NEWTON, ROBERT U.; NONEMACHER, ELISA R.; WENDT, VICTÓRIA M.; BASSANESI, RENATA N.; TURELLA, DOUGLAS J. P.; RECH, ANDERSON | Ineligible population |
| Moderators of School-Based Physical Activity Interventions on Cardiorespiratory Endurance in Primary School-Aged Children: A Meta-Regression | Burns, R. D., T. A. Brusseau and Y. Fu | Ineligible population |
| Monitoring and adapting endurance training on the basis of heart rate variability monitored by wearable | Düking, Peter; Zinner, Christoph; Trabelsi, Khaled; Reed, Jennifer L.; Holmberg, | Ineligible population |
| technologies: A systematic review with meta-analysis. | Hans-Christer; Kunz, Philipp; Sperlich, Billy | |
| Motivational Interviewing for Parent-child Health Interventions: A Systematic Review and Meta-Analysis | Borrelli, B., E. M. Tooley and L. A. Scott-Sheldon | Ineligible population |
| Motivational Interviewing for Weight Management Among Women: a Meta-Analysis and Systematic Review of RCTs | Suire, Kameron B.; Kavookjian, Jan; Feiss, Robyn; Wadsworth, Danielle D | Ineligible population |
| Multidisciplinary Neuromuscular and Endurance Interventions on Youth Basketball Players: A Systematic Review with Meta-Analysis and Meta-Regression | Arnau Sacot, Víctor López-Ros, Anna Prats-Puig, Jesús Escosa, Jordi Barretina, Julio Calleja-González. | Ineligible population |
| Muscle Fiber Hypertrophy and Myonuclei Addition: A Systematic Review and Meta-analysis. | CONCEIÇÃO, MIGUEL S.; VECHIN, FELIPE C.; LIXANDRÃO, MANOEL; DAMAS, FELIPE; TRICOLI, VALMOR; ROSCHEL, HAMILTON; UGRINOWITSCH, CARLOS; LIBARDI, CLEITON A.; CAMERA, DONNY | Ineligible population |
| Muscle hypertrophy and strength gains after resistance training with different volume-matched loads: a systematic review and meta-analysis. | Carvalho, Leonardo; Junior, Roberto Moriggi; Barreira, Júlia; Schoenfeld, Brad J.; Orazem, John; Barroso, Renato; | Ineligible population |

| | | |
|---|---|---|
| Nature-Based Early Childhood Education and Children's Physical Activity, Sedentary Behavior, Motor Competence, and Other Physical Health | Johnstone, A.; McCrorie, P.; Cordovil, R.; Fjørtoft, I.; Iivonen, S.; Jidovtseff, B.; Lopes, F.; Reilly, J.J.; Thomson, H.; Wells, V.; Martin, A.; | Ineligible population |
| Neuromuscular Training and Motor Control in Youth Athletes: A Meta-Analysis | Mark David Williams, Rodrigo Ramirez-Campillo, Helmi Chaabene, and Jason Moran | Ineligible population |
| Nutrition, Physical Activity, and New Technology Programs on Obesity Prevention in Primary Education: A Systematic Review | Lorenzo Navidad, Rosario Padial-Ruz * and Mar Cepero González | Ineligible population |
| Parent participation in weight-related health interventions for children and adolescents: a systematic review and meta-analysis | Niemeier, B. S., J. M. Hektner and K. B. Enger | Ineligible population |
| Parent-only interventions for childhood overweight or obesity in children aged 5 to 11 years | Loveman, E., L. Al-Khudairy, R. E. Johnson, W. Robertson, J. L. Colquitt, E. L. Mead, L. J. Ells, M. I. Metzendorf and K. Rees | Ineligible population |
| Pediatric Obesity Prevention and Treatment Among Hispanics: A Systematic Review and Meta-Analysis | Sara M. St. George, Marissa A. Kobayashi, Blanca Noriega Esquives, Manuel Ocasio, Rachel G. Wagstaff, David Dorcius | Ineligible population |
| Pediatric Residency Obesity and Overweight Training Curricula: A Systematic Review | Molly Silber, Lindsay Weiss,Salma Sharaf, Yan Wang, Erin Hager, Rebecca Carter. | Ineligible population |
| Physical Activity and Cognitive Development: A Meta-Analysis | Jackson, W. M., N. Davis, S. A. Sands, R. A. Whittington and L. S. Sun | Ineligible population |
| Physical Activity and Health in Children under 6 Years of Age: A Systematic Review | Russell R. Pate, Charles Hillman, Kathleen Janz, Peter T. Katzmarzyk, Kenneth E. Powell, Andrea Torres, Melicia C. Whitt-Glover, and for the 2018 Physical Activity Guidelines Advisory Committee* | Ineligible population |
| Physical activity and health-related quality of life in children and adolescents: A systematic review and meta-analysis | Marker, A. M., R. G. Steele and A. E. Noser | Ineligible population |
| Physical activity and physical self-concept in youth: systematic review and meta-analysis | Babic, M. J., P. J. Morgan, R. C. Plotnikoff, C. Lonsdale, R. L. White and D. R. Lubans | Ineligible population |
| Physical activity and prospective associations with indicators of health and development in children aged <5 years: a systematic review | Sanne L. C. Veldman* , Mai J. M. Chin A Paw and Teatske M. Altenburg | Ineligible population |
| Physical activity and sleep are inconsistently related in healthy children: A systematic review and meta-analysis | Devan Antczak, Chris Lonsdale, Jane Lee, Toni Hilland, Mitch J. Duncan, Borja del Pozo Cruz, Ryan M. Hulteen, Philip D. Parker, Taren Sanders | Ineligible population |
| Physical Activity for the Treatment of Adolescent Depression: A Systematic Review and Meta-Analysis | Max Oberste, Marie Medele, Florian Javelle, Heidrun Lioba Wunram, Daniel Walter, Wilhelm Bloch, Stephan Bender, Oliver Fricke, Niklas Joisten, David Walzik, Nicola | Ineligible population |

| | | |
|---|---|---|
| | Großheinrich and Philipp Zimmer | |
| Physical activity interventions in early life aimed at reducing later risk of obesity and related non-communicable diseases: A rapid review of systematic reviews | John J. Reilly, Adrienne R. Hughes, Jennifer Gillespie, Stephen Malden, Anne Martin | Ineligible population |
| Physical activity interventions in Latin America: expanding and classifying the evidence | Hoehner, C. M., I. C. Ribeiro, D. C. Parra, R. S. Reis, M. R. Azevedo, A. A. Hino, J. Soares, P. C. Hallal, E. J. Simoes and R. C. Brownson | Ineligible population |
| Physical activity interventions in the school setting: A systematic review | Demetriou, Y. and O. HÃ¶ner | Ineligible population |
| Physical activity programmes to reduce overweight and obesity in children and adolescents; a systematic review | Aguilar Cordero, M. J., A. Ortegon Pinero, N. Mur Vilar, J. C. Sanchez Garcia, J. J. Garcia Verazaluce, I. Garcia Garcia and A. M. Sanchez Lopez | Ineligible population |
| Physical activity promotion in children and adolescents using peer support: Systematic review of the literature | Goncalves, R., R. Batalau, J. Carmo, J. Cruz, J. Leal and A. Palmeira | Ineligible population |
| Physical Activity–Based Interventions Using Electronic Feedback May Be Ineffective in Reducing Pain and Disability in Patients With Chronic Musculoskeletal Pain: A Systematic Review With Meta-Analysis. | Oliveira, Crystian B.; Franco, Márcia R.; Maher, Chris G.; Ferreira, Paulo H.; Morelhão, Priscila K.; Damato, Tatiana M.; Gobbi, Cynthia; Pinto, Rafael Z.; | Ineligible population |
| Physical activity, diet and other behavioural interventions for improving cognition and school achievement in children and adolescents with obesity or overweight | Martin, A., J. N. Booth, Y. Laird, J. Sproule, J. J. Reilly and D. H. Saunders | Ineligible population |
| Physically active lessons in schools and their impact on physical activity, educational, health and cognition outcomes: a systematic review and meta-analysis | Emma Norris,Tommy van Steen, Artur Direito, Emmanuel Stamatakis | Ineligible population |
| Plyometric exercise and bone health in children and adolescents: a systematic review | Gomez-Bruton, A., A. Matute-Llorente, A. Gonzalez-Aguero, J. A. Casajus and G. Vicente-Rodriguez | Ineligible population |
| Preventative interventions that target cardiovascular dysfunction in children and young people: a systematic review of their effectiveness and an investigation of sexual dimorphism | Sophie Edwards, Murray Foster, S. Faisal Ahmed and Angela K. Lucas-Herald | Ineligible population |
| Prevention of Type 2 Diabetes among Youth: A Systematic Review, Implications for the School Nurse | Brackney, D. E. and M. Cutshall | Ineligible population |
| Prevention of Type 2 Diabetes in U.S. Hispanic Youth: A Systematic Review of Lifestyle Interventions | McCurley, J. L., M. A. Crawford and L. C. Gallo | Ineligible population |
| Prevention of unhealthy weight in children by promoting physical activity using a | Kellou, N., F. Sandalinas, N. Copin and C. Simon | Ineligible population |

| | | |
|---|---|---|
| socio-ecological approach: what can we learn from intervention studies? | | |
| Programming Interval Training to Optimize Time‐Trial Performance: A Systematic Review and Meta‐Analysis | Michael A. Rosenblat, Edward Lin, Bruno R. da Costa, Scott G. Thomas | Ineligible population |
| Promoting health-related cardiorespiratory fitness in physical education: A systematic review | Miguel Peralta, Duarte Henriques-Neto, E´lvio Ru´bio Gouveia, Lui´s B. Sardinha, Adilson Marques | Ineligible population |
| Psychosocial and Physiological Health Outcomes of Green Exercise in Children and Adolescents—A Systematic Review | Carina Mnic, Susanne Weyland, Darko Jekauc and Jasper Schipperijn | Ineligible population |
| Quadriceps Strength and Volitional Activation After Anterior Cruciate Ligament Reconstruction: A Systematic Review and Meta-analysis. | Lisee, Caroline; Lepley, Adam S.; Birchmeier, Thomas; O'Hagan, Kaitlin; Kuenze, Christopher; | Ineligible population |
| Recreational Football and Bone Health: A Systematic Review and Meta-analysis | Milanović, Z.; Čović, N.; Helge, E.W.; Krustrup, P.; Mohr, M.; | Ineligible population |
| Recreos activos como estrategia de promoción de la actividad física: una revisión sistemática. / Active recess as a strategy to promote physical activity: a systematic review. | Pastor-Vicedo, Juan Carlos; Martínez-Martínez, Jesús; López-Polo, María; Prieto-Ayuso, Alejandro | Ineligible population |
| Reducing aggression with martial arts: A meta-analysis of child and youth studies | Harwood, A., M. Lavidor and Y. Rassovsky | Ineligible population |
| Resistance Training and Handball Players' Isokinetic, Isometric and Maximal Strength, Muscle Power and Throwing Ball Velocity: A Systematic Review and Meta-Analysis | Nicola Luigi Bragazzi, Mehdi Rouissi, Souhail Hermassi and Karim Chamari | Ineligible population |
| Resistance Training Effects on Metabolic Function Among Youth: A Systematic Review | Bea, J. W., R. M. Blew, C. Howe, M. Hetherington-Rauth and S. B. Going | Ineligible population |
| Resistance Training Load Effects on Muscle Hypertrophy and Strength Gain: Systematic Review and Network Meta-analysis. | LOPEZ, PEDRO; RADAELLI, RÉGIS; TAAFFE, DENNIS R.; NEWTON, ROBERT U.; GALVÃO, DANIEL A.; TRAJANO, GABRIEL S.; TEODORO, JULIANA L.; KRAEMER, WILLIAM J.; HÄKKINEN, KEIJO; PINTO, RONEI S.; | Ineligible population |
| Resistance training to improve power and sports performance in adolescent athletes: a systematic review and meta-analysis | Harries, S. K., D. R. Lubans and R. Callister | Ineligible population |
| Review of High-Intensity Interval Training for Cognitive and Mental Health in Youth | ANGUS A. LEAHY, MYRTO F. MAVILIDI, JORDAN J. SMITH, CHARLES H. HILLMAN, NARELLE EATHER, DANIEL BARKER and DAVID R. LUBANS | Ineligible population |
| Review: A systematic review of the impact of physical activity programmes on social and emotional well-being in at-risk youth | Lubans, D. R., R. C. Plotnikoff and N. J. Lubans | Ineligible population |

| | | |
|---|---|---|
| Revisión sistemática del entrenamiento de fuerza en futbolistas preadolescentes y adolescentes. / Systematic review of strength training in preadolescent and adolescent football players. | Martínez-Pérez, Pablo; Vaquero-Cristóbal, Raquel | Ineligible population |
| Revisión sistemática sobre los estudios de intervención de actividad física para el tratamiento de la obesidad. / Systematic Review of Physical Activity Programs for the treatment of Obesity. | Cuadri Fernández, Jorge; Tornero Quiñones, Inmaculada; Sierra Robles, Ángela; Sáez Padilla, Jesús Manuel; | Ineligible population |
| REVISIÓN SISTEMÁTICA SOBRE PROGRAMAS PARA LA PREVENCIÓN DE LESIONES DE LA EXTREMIDAD INFERIOR EN JÓVENES DEPORTISTAS. / A SYSTEMATIC REVIEW ABOUT LOWER EXTREMITY INJURIES PREVENTION PROGRAMS FOR YOUNG ATHLETES. | Robles-Palazón, F. J.; Cejudo, A.; Ayala, F.; Sainz de Baranda, P. | Ineligible population |
| Right heart exercise‐training‐adaptation and remodelling in endurance athletes | Valeria Conti, Filippo Migliorini, Marco Pilone, María I. Barriopedro, Juan José Ramos‐Álvarez, Francisco Javer Calderon Montero & Nicola Mafulli | Ineligible population |
| Role of Physical Activity and Sedentary Behavior in the Mental Health of Preschoolers, Children and Adolescents: A Systematic Review and Meta‐Analysis. | María Rodriguez‐Ayllon,Cristina Cadenas‐Sánchez, Fernando Estévez‐López, Nicolas E. Muñoz, Jose Mora‐Gonzalez,Jairo H. Migueles,Pablo Molina‐García, Hanna Henriksson,Alejandra Mena‐Molina, Vicente Martínez‐Vizcaíno, Andrés Catena, Marie Löf,Kirk I. Erickson, David R. Lubans, Francisco B. Ortega, Irene Esteban‐Cornejo. | Ineligible population |
| School-Based Exercise Programs and Cardiometabolic Risk Factors: A Meta-analysis | Diana P. Pozuelo-Carrascosa, MSci,a Iván Cavero-Redondo, PhD,a Ángel Herráiz-Adillo, MSci,a,b Ana Díez-Fernández, PhD, Mairena Sánchez-López, PhD,a,d Vicente Martínez-Vizcaíno, MD, PhDa, | Ineligible population |
| School-based high-intensity interval training programs in children and adolescents: A systematic review and meta-analysis | Stephanie L. Duncombe, Alan R. Barker, Bert Bond, Renae Earle, Jo Varley-Campbell, Dimitris Vlachopoulos, Jacqueline L. Walker, Kathryn L. Weston, Michalis Stylianou | Ineligible population |
| School-based high-intensity interval training programs in children and adolescents: A systematic review and meta-analysis | Stephanie L. Duncombe, Alan R. Barke,Bert Bond, Renae Earle, Jo Varley-Campbell,Dimitris Vlachopoulos,Jacqueline L. Walker, Kathryn L. Weston,Michalis Stylianou | Ineligible population |

| | | |
|---|---|---|
| School-based high-intensity interval training programs in children and adolescents: A systematic review and meta-analysis | Stephanie L. DuncombeID1,2*, Alan R. Barker2 | Ineligible population |
| School-based high-intensity interval training programs in children and adolescents: A Systematic Review and Meta-Analysis. | Barker, A.; Bond, B.; Duncombe, S.; Earle, R.; Stylianou, M.; Varley-Campbell, J.; Vlachopoulos, D.; Walker, J.; Weston, K.; | Ineligible population |
| School-based intervention practices to reduce body mass index: Meta-analysis of 57 randomized trials | Guerra, P. H., M. R. C. Nobre, J. A. A. Taddei and J. A. C. Silveira | Ineligible population |
| School-Based Interventions Targeting Nutrition and Physical Activity, and Body Weight Status of African Children: A Systematic Review | Theodosia Adom, Anniza De Villiers , Thandi Puoane and André Pascal Kengne | Ineligible population |
| School-based interventions to improve spinal health of children and adolescents: a systematic review | Yolandi Brink , Rentia Amelia Maart , and Quinette Abigail Louw | Ineligible population |
| School-based Interventions to Reduce Sedentary Behaviour in Children: A Systematic Review | Hegarty, L. M., J. L. Mair, K. Kirby, E. Murtagh and M. H. Murphy | Ineligible population |
| School-based obesity prevention interventions in Latin America: A systematic review | Rosemary Cosme Chavez , Eun Woo NamI, | Ineligible population |

| | | |
|---|---|---|
| School-based physical activity programs for promoting physical activity and fitness in children and adolescents aged 6 to 18 (Review) | Neil-Sztramko SE, Caldwell H, Dobbins M | Ineligible population |
| School-related physical activity interventions and mental health among children: a systematic review and meta-analysis | Susanne Andermo, Mats Hallgren, Thi-Thuy-Dung Nguyen, Sofie Jonsson, Solveig Petersen, Marita Friberg, Anja Romqvist, Brendon Stubbs and Liselotte Schäfer Elinder | Ineligible population |
| School-wide programs aimed at obesity among Latino youth in the United States: a review of the evidence | Holub, C. K., F. Lobelo, S. M. Mehta, L. M. Sanchez Romero, E. M. Arredondo and J. P. Elder | Ineligible population |
| School‑based Physical Activity Interventions in Children and Adolescents: A Systematic Review | Mikel Vaquero‑Solís, Damián Iglesias Gallego, Miguel Ángel Tapia‑Serrano, Juan J. Pulido and Pedro Antonio Sánchez‑Miguel | Ineligible population |
| Sedentary Time and Behavior during School: A Systematic Review and Meta-Analysis. | Egan, Catherine A.; Webster, Collin A.; Beets, Michael W.; Weaver, R. Glenn; Russ, Laura; Michael, Daniel; Nesbitt, Danielle; Orendorff, Karie L. | Ineligible population |
| Self-Monitoring via Digital Health in Weight Loss Interventions: A Systematic Review Among Adults with Overweight or Obesity. | Patel, Michele L.; Wakayama, Lindsay N.; Bennett, Gary G.; | Ineligible population |
| Self-report use-of-time tools for the assessment of physical activity and sedentary | Foley, L., R. Maddison, T. Olds and K. Ridley | Ineligible population |

| | | |
|---|---|---|
| behaviour in young people: systematic review | | |
| Shaping Pathways to Child Health: A Systematic Review of Street-Scale Interventions in City Streets | Adriana Ortegon-Sanchez, Laura Vaughan , Nicola Christie and Rosemary R. C. McEachan | Ineligible population |
| Single Leg Balance Training: A Systematic Review. | Marcori, Alexandre J; Monteiro, Pedro H M; Oliveira, Júlia A; Doumas, Michail; Teixeira, Luis A | Ineligible population |
| Skeletal muscle functional and structural adaptations after eccentric overload flywheel resistance training: a systematic review and meta-analysis. | Vicens-Bordas, J.; Esteve, E.; Fort-Vanmeerhaeghe, A.; Bandholm, T.; Thorborg, K. | Ineligible population |
| Skeletal Muscle Glycogen Content at Rest and During Endurance Exercise in Humans: A Meta-Analysis. | Areta, José L.; Hopkins, Will G. | Ineligible population |
| Slackline Training (Balancing Over Narrow Nylon Ribbons) and Balance Performance: A Meta-Analytical Review | Donath, L., R. Roth, L. Zahner and O. Faude | Ineligible population |
| Small-sided games in volleyball: A systematic review of the state of the art. | de Oliveira Castro, Henrique; Laporta, Lorenzo; Lima, Ricardo Franco; Clemente, Filipe Manuel; Afonso, José; da Silva Aguiar, Samuel; de Araújo Ribeiro, Alexandre Lima; De Conti Teixeira Costa, Gustavo | Ineligible population |
| Smartphone-Based Interventions for Physical Activity Promotion: Scoping | Alex Domin, Donna Spruijt-Metz, Daniel Theisen, Yacine Ouzzahra, Claus Vögele. | Ineligible population |

| | | |
|---|---|---|
| Review of the Evidence Over the Last 10 Years | | |
| Specific Strategies for Promotion of Physical Activity in Kids-Which Ones Work? A Systematic Review of the Literature | McDonald, S. M., M. N. Clennin and R. R. Pate | Ineligible population |
| Strategies for enhancing the implementation of school-based policies or practices targeting risk factors for chronic disease(Review) | Wolfenden L, Nathan NK, Sutherland R, Yoong SL, Hodder RK, Wyse RJ, Delaney T, Grady A,Fielding A, Tzelepis F, Clinton-McHarg T, Parmenter B, Butler P, Wiggers J, Bauman A, Milat A,Booth D, Williams CM | Ineligible population |
| Strategies to improve the implementation of healthy eating, physical activity and obesity prevention policies, practices or programmes within childcare services (Review) | Wolfenden L, Barnes C, Jones J, Finch M, Wyse RJ, Kingsland M, Tzelepis F, Grady A, Hodder RK, Booth D, Yoong SL | Ineligible population |
| Strength and conditioning practices for the optimisation of speed and accuracy in cricket fast bowlers: A systematic review and meta-analysis. | Ramachandran, Akhilesh Kumar; Singh, Utkarsh; Lathlean, Timothy JH | Ineligible population |
| Systematic review and meta-analysis investigating moderators of long-term effects of exercise on cognition in healthy individuals | Sebastian Ludyga, Markus Gerber, Uwe Pühse, Vera N. Looser and Keita Kamijo | Ineligible population |
| Systematic review and meta-analysis of health promotion interventions for | Cushing, C. C., E. E. Brannon, K. I. Suorsa and D. K. Wilson | Ineligible population |

| | | |
|---|---|---|
| children and adolescents using an ecological framework | | |
| Systematic review and meta-analysis of linear and undulating periodized resistance training programs on muscular strength | Harries, S. K., D. R. Lubans and R. Callister | Ineligible population |
| Systematic review and meta-analysis of school-based interventions to reduce body mass index | Lavelle, H. V., D. F. Mackay and J. P. Pell | Ineligible population |
| Systematic review and meta-analysis of the efects of exercise on depression in adolescents | Xiang Wang, Zhi-dong Cai, Wan-ting Jiang, Yan-yan Fang, Wen-xin Sun and Xing Wang. | Ineligible population |
| Systematic review of interventions in the childcare setting with direct parental involvement: effectiveness on child weight status and energy balance-related behaviours | I. van de Kolk* , S. R. B. Verjans-Janssen, J. S. Gubbels, S. P. J. Kremers and S. M. P. L. Gerards | Ineligible population |
| Systematic review of multicomponent interventions with overweight middle adolescents: implications for clinical practice and research | Kelly, S. A. and B. M. Melnyk | Ineligible population |
| Systematic Review of Physical Education-Based Physical Activity Interventions Among Elementary School Children | Errisuriz, V. L., N. M. Golaszewski, K. Born and J. B. Bartholomew | Ineligible population |
| Systematic review of recess interventions to increase physical activity | Ickes, M. J., H. Erwin and A. Beighle | Ineligible population |

| | | |
|---|---|---|
| Systematic review of school-based obesity interventions targeting African American and Hispanic children | Knowlden, A. P. and M. Sharma | Ineligible population |
| Systematic Review of Setting-Based Interventions for Preventing Childhood Obesity. | Khadijah Angawi and Anood Gaissi | Ineligible population |
| Systematic review of text messaging as an intervention for adolescent obesity | Keating, S. R. and M. K. McCurry | Ineligible population |
| Systematic review of the efficacy of the prescription of physical activity in obese child population | Hernandez, E. D., M. V. Valero Bernal and E. M. Mancera Soto | Ineligible population |
| Systematic review of the relationship between 20m shuttle run performance and health indicators among children and youth | Justin J. Lang, Kevin Belanger, Veronica Poitras, Ian Janssen, Grant Tomkinson | Ineligible population |
| Systematic Review: Nutrition and Physical Activity in the Management of Paediatric Nonalcoholic Fatty Liver Disease | Gibson, P. S., S. Lang, A. Dhawan, E. Fitzpatrick, M. L. Blumfield, H. Truby, K. H. Hart and J. B. Moore | Ineligible population |
| Targeted exercise against osteoporosis: A systematic review and meta-analysis for optimising bone strength throughout life | Nikander, R., H. Sievanen, A. Heinonen, R. M. Daly, K. Uusi-Rasi and P. Kannus | Ineligible population |
| Temporal Trends in the Standing Broad Jump Performance of United States Children and Adolescents | Bridget K. Pinoniemi, Grant R. Tomkinson, Tanis J. Walch, James N. Roemmich & John S. Fitzgerald | Ineligible population |

| | | |
|---|---|---|
| The Acute and Chronic Effects of Implementing Velocity Loss Thresholds During Resistance Training: A Systematic Review, Meta-Analysis, and Critical Evaluation of the Literature. | Jukic, Ivan; Castilla, Alejandro Pérez; Ramos, Amador García; Van Hooren, Bas; McGuigan, Michael R.; Helms, Eric R. | Ineligible population |
| The association of resistance training with mortality: A systematic review and meta-analysis] | Farzane Saeidifard, Jose R Medina-Inojosa, Colin P West,Thomas P Olson, Virend K Somers, Amanda R Bonikowske, Larry J Prokop, Manlio Vinciguerra and Francisco Lopez-Jimenez | Ineligible population |
| The Efects of Trunk Muscle Training on Physical Fitness and Sport-Specifc Performance in Young and Adult Athletes: A Systematic Review and Meta-Analysis | Atle H. Saeterbakken, Nicolay Stien, Vidar Andersen, Suzanne Scott, Kristoffer T. Cumming, David G. Behm, Urs Granacher, Olaf Prieske | Ineligible population |
| The effect of acute and chronic exercise on cognitive function and academic performance in adolescents: A systematic review | Li, J. W., H. O'Connor, N. O'Dwyer and R. Orr | Ineligible population |
| The effect of aquatic high-intensity interval training on aerobic performance, strength and body composition in a non-athletic population: systematic review and meta-analysis. | Depiazzi, Julie E.; Forbes, Rachel A.; Gibson, Noula; Smith, Nadine L.; Wilson, Andrew C.; Boyd, Roslyn N.; Hill, Kylie | Ineligible population |
| The effect of augmented feedback on the performance and learning of gross motor and sport-specific skills: A systematic review | Emma Louise Petancevski, Joshua Inns, Job Fransen, Franco Milko Impellizzeri | Ineligible population |
| The Effect of Classroom-Based Interventions on Sedentary Behavior and Spinal Health in Schoolchildren:Systematic Review | Dominic Fisher, MSc; Quinette Louw, PhD | Ineligible population |
| The effect of diet or exercise on ectopic adiposity in children and adolescents with obesity: a systematic review and meta-analysis | Hens, W., D. Vissers, D. Hansen, S. Peeters, J. Gielen, L. Van Gaal and J. Taeymans | Ineligible population |
| The effect of dietary (poly)phenols on exercise-induced physiological adaptations: A systematic review and meta-analysis of human intervention trials. | Martinez-Negrin, Guille; Acton, Jarred P.; Cocksedge, Stuart P.; Bailey, Stephen J.; Clifford, Tom | Ineligible population |
| The effect of exercise interventions on resting metabolic rate: A systematic review and meta-analysis. | MacKenzie-Shalders, Kristen; Kelly, Jaimon T.; So, Daniel; Coffey, Vernon G.; Byrne, Nuala M. | Ineligible population |
| The Effect of Intensity, Frequency, Duration and Volume of Physical Activity in Children and Adolescents on Skeletal Muscle Fitness: A Systematic Review and Meta-Analysis of Randomized Controlled Trials | Chunchun Wu , Yongjin Xu , Zhaojing Chen , Yinhang Cao , Kehong Yu and Cong Huang | Ineligible population |
| The effect of martial arts training on mental health outcomes: A systematic review and meta-analysis. | Moore, Brian; Dudley, Dean; Woodcock, Stuart | Ineligible population |

| | | |
|---|---|---|
| The Effect of Nordic Hamstring Exercise Intervention Volume on Eccentric Strength and Muscle Architecture Adaptations: A Systematic Review and Meta-analyses. | Cuthbert, Matthew; Ripley, Nicholas; McMahon, John J.; Evans, Martin; Haff, G. Gregory; Comfort, Paul | Ineligible population |
| The effect of physical training on heart rate variability in healthy children: a systematic review with meta-analysis | da Silva, C. C., L. M. Pereira, J. R. Cardoso, J. P. Moore and F. Y. Nakamura | Ineligible population |
| The effect of resistance training interventions on fundamental movement skills in youth: a meta-analysis | Helen Collins, Josephine N. Booth, Audrey Duncan and Samantha Fawkner | Ineligible population |
| The Effect of Resistance Training in Healthy Adults on Body Fat Percentage, Fat Mass and Visceral Fat: A Systematic Review and Meta-Analysis. | Wewege, Michael A.; Desai, Imtiaz; Honey, Cameron; Coorie, Brandon; Jones, Matthew D.; Clifford, Briana K.; Leake, Hayley B.; Hagstrom, Amanda D.; | Ineligible population |
| The Effect of Resistance Training in Women on Dynamic Strength and Muscular Hypertrophy: A Systematic Review with Meta-analysis. | Hagstrom, Amanda D.; Marshall, Paul W.; Halaki, Mark; Hackett, Daniel A. | Ineligible population |
| The Effectiveness of Aquatic Plyometric Training in Improving Strength, Jumping, and Sprinting: A Systematic Review. | Heywood, Sophie E.; Mentiplay, Benjamin F.; Rahmann, Ann E.; McClelland, Jodie A.; Geigle, Paula R.; Bower, Kelly J.; Clark, Ross A. | Ineligible population |
| The Effectiveness of Classical Ballet Training on Health-Related Outcomes: A Systematic Review | Letton ME, Thom JM, Ward RE | Ineligible population |

| | | |
|---|---|---|
| The effectiveness of exercise interventions to prevent sports injuries: a systematic review and meta-analysis of randomised controlled trials | Lauersen, J. B., D. M. Bertelsen and L. B. Andersen | Ineligible population |
| The Effectiveness of Gait Retraining on Running Kinematics, Kinetics, Performance, Pain, and Injury in Distance Runners: A Systematic Review With Meta-analysis. | DOYLE, EOIN; DOYLI, TIM L. A.; BONACCI, JASON; FULLER, JOEL T | Ineligible population |
| The effectiveness of incidental physical activity interventions compared to other interventions in the management of people with low back pain: A systematic review and meta-analysis of randomised controlled trials. | Alzahrani, Hosam; Mackey, Martin; Stamatakis, Emmanuel; Pinheiro, Marina B.; Wicks, Manuela; Shirley, Debra; | Ineligible population |
| The effectiveness of intervention on the physical activity of children: Systematic review and metaanalysis of controlled trials with objectivelymeasured outcomes | Metcalf, B., W. Henley and T. Wilkin | Ineligible population |
| The effectiveness of interventions to increase physical activity among young girls: a meta-analysis | Biddle, S. J., R. Braithwaite and N. Pearson | Ineligible population |
| The effectiveness of interventions to increase physical activity. A systematic review | Kahn, E. B., L. T. Ramsey, R. C. Brownson, G. W. Heath, E. H. Howze, K. E. Powell, E. J. Stone, M. W. Rajab and P. Corso | Ineligible population |

| | | |
|---|---|---|
| The Effectiveness of Interventions to Reduce Sedentary Time in Different Target Groups and Settings in Germany: Systematic Review, Meta-Analysis and Recommendations on Interventions | Nida Mugler, Hansjörg Baurecht , Kevin Lam, Michael Leitzmann and Carmen Jochem | Ineligible population |
| The effectiveness of low intensity exercise and blood flow restriction without exercise on exercise induced muscle damage: A systematic review. | Ma, Fenghao; Li, Yingqi; Yang, Jinchao; Li, Xidian; Zeng, Na; Martin, RobRoy L. | Ineligible population |
| The Effectiveness of Physical Activity-Promoting Web- and Mobile-Based Distance Weight Loss Interventions on Body Composition in Rehabilitation Settings: Systematic Review, Meta-analysis, and Meta-Regression Analysis | Heli Lahtio, MSc, Aki Rintala, PhD, Jaakko Immonen, MSc, and Tuulikki Sjögren, PhD | Ineligible population |
| The Effectiveness of Sedentary Behaviour Reduction Workplace Interventions on Cardiometabolic Risk Markers: A Systematic Review. | Brierley, Marsha L.; Chater, Angel M.; Smith, Lindsey R.; Bailey, Daniel P. | Ineligible population |
| The Effectiveness of Wearable Devices as Physical Activity Interventions for Preventing and Treating Obesity in Children and Adolescents: Systematic Review and Meta-analysis | Wentao Wang, MEd, Jing Cheng, MEd, Weijun Song, MEd, and Yi Shen, MEd | Ineligible population |

| | | |
|---|---|---|
| The Effects of Acute Exposure to Prolonged Sitting, with and Without Interruption, on Peripheral Blood Pressure Among Adults: A Systematic Review and Meta-Analysis | Paterson, Craig; Fryer, Simon; Stone, Keeron; Zieff, Gabriel; Turner, Louise; Stoner, Lee | Ineligible population |
| The Effects of Concurrent Strength and Endurance Training on Physical Fitness and Athletic Performance in Youth: A Systematic Review and Meta-Analysis | Gabler, M., O. Prieske, T. Hortobagyi and U. Granacher | Ineligible population |
| The Effects of Exclusive Walking on Lipids and Lipoproteins in Women with Overweight and Obesity: A Systematic Review and Meta-Analysis. | Ballard, Anjulyn M.; Davis, Ashlee; Wong, Brett; Lyn, Rodney; Thompson, Walter R. | Ineligible population |
| The Effects of Exercise Dosage on Neck-Related Pain and Disability: A Systematic Review With Meta-analysis | WILHELM, MARK P.; DONALDSON, MEGAN; GRISWOLD, DAVID; LEARMAN, KENNETH E.; GARCIA, ALESSANDRA N.; LEARMAN, SHANE M.; CLELAND, JOSHUA A. | Ineligible population |
| The effects of exercise interventions on physical function tests and glycemic control in adults with type 2 diabetes: A systematic review. | Janssen, Sarah M.; Connelly, Denise M. | Ineligible population |
| The effects of exercise on resting blood pressure in children and adolescents: a meta-analysis of randomized controlled trials | Kelley, G. A., K. S. Kelley and Z. V. Tran | Ineligible population |

| | | |
|---|---|---|
| The Effects of Green Exercise on Physical and Mental Wellbeing: A Systematic Review | Ian Lahart, Patricia Darcy, Christopher Gidlow and Giovanna Calogiuri | Ineligible population |
| The Effects of Physical Activity and Diet Interventions on Body Mass Index in Latin American Children and Adolescents: A Systematic Review and Meta-Analysis | Andrés Godoy-Cumillaf , Paola Fuentes-Merino, Armando Díaz-González, Judith Jiménez-Díaz, Vicente Martínez-Vizcaíno, Celia Álvarez-Bueno and Iván Cavero-Redondo | Ineligible population |
| The Effects of Physical Activity on Positive Emotions in Children and Adolescents: A Systematic Review and Meta-Analysis | Jiayu Li, Zan Huang, Wenna Si and Tianyi Shao | Ineligible population |
| The Effects of Physical Education on Motor Competence in Children and Adolescents: A Systematic Review and Meta-Analysis | Håvard Lorås | Ineligible population |
| The effects of plyometric jump training on physical fitness attributes in basketball players: A meta-analysis. | Ramirez-Campillo, Rodrigo; García-Hermoso, Antonio; Moran, Jason; Chaabene, Helmi; Negra, Yassine; Scanlan, Aaron T. | Ineligible population |
| The Effects of Tai Chi and Qigong Exercise on Psychological Status in Adolescents: A Systematic Review and Meta-Analysis | Xuan Liu, Ru Li, Jiabao Cui , Fang Liu, Lee Smith, Xiaorong Chen and Debao Zhang | Ineligible population |
| The Effects of Thai Yoga on Physical Fitness: A | Kongkaew, C., P. Lertsinthai, K. Jampachaisri, P. Mongkhon, P. | Ineligible population |
| Meta-Analysis of Randomized Control Trials | Meesomperm, K. Kornkaew and P. Malaiwong | |
| The impact of child and adolescent obesity treatment interventions on physical activity: a systematic review | Cliff, D. P., A. D. Okely, P. J. Morgan, R. A. Jones and J. R. Steele | Ineligible population |
| The impact of exercise training versus caloric restriction on inflammation markers: a systemic review and meta-analysis. | Khalafi, Mousa; Symonds, Michael E.; Akbari, Amir | Ineligible population |
| The impact of high‑intensity interval training on inflammatory markers in metabolic disorders: A meta‑analysis. | Khalafi, Mousa; Symonds, Michael E. | Ineligible population |
| The impact of isometric handgrip exercise and training on health‑related factors: A review | Yamada, Yujiro; Spitz, Robert W.; Wong, Vickie; Bell, Zachary W.; Song, Jun Seob; Abe, Takashi; Loenneke, Jeremy P. | Ineligible population |
| The Impact of Sex on Left Ventricular Cardiac Adaptations to Endurance Training: a Systematic Review and Meta-analysis. | Diaz-Canestro, Candela; Montero, David | Ineligible population |
| The impacts of unstructured nature play on health in early childhood development: A systematic review | Kylie A. Dankiw, Margarita D. Tsiros, Katherine L. Baldock, Saravana Kumar | Ineligible population |
| The influence of dosing on effect size of exercise therapy for musculoskeletal foot and | Young, Jodi L.; Rhon, Daniel I.; de Zoete, Rutger M. J.; Cleland, Joshua A.; Snodgrass, Suzanne J. | Ineligible population |

| | | |
|---|---|---|
| ankle disorders: a systematic review. | | |
| The long-term effects of primary school-based obesity prevention interventions in children: A systematic review and meta-analysis | Michel S. Smit, Mirte Boelens, Famke J. M. Mölenberg, Hein Raat, Wilma Jansen | Ineligible population |
| The Natural History of Full-Thickness Rotator Cuff Tears in Randomized Controlled Trials: A Systematic Review and Meta-analysis. | Khatri, Chetan; Ahmed, Imran; Parsons, Helen; Smith, Nicholas A.; Lawrence, Thomas M.; Modi, Chetan S.; Drew, Stephen J.; Bhabra, Gev; Parsons, Nicholas R.; Underwood, Martin; Metcalfe, Andrew J.; | Ineligible population |
| The prospective association between objectively measured sedentary time, moderate-to-vigorous physical activity and cardiometabolic risk factors in youth: a systematic review and meta-analysis | T. Skrede, J. Steene-Johannessen, S. A. Anderssen, G. K. Resaland1 and U. Ekelund | Ineligible population |
| The Relationship Between Motor Competence and Physical Fitness from Early Childhood to Early Adulthood: A Meta‑Analysis | Till Utesch, Farid Bardid, Dirk Büsch, Bernd Strauss | Ineligible population |
| The Relationship Between Non-elite Sporting Activity and Calcaneal Bone Density in Adolescents and Young Adults: A Narrative Systematic Review | Hansa Patel, Luke Sammut , Hayley Denison, Paul Teesdale-Spittle and Elaine Dennison | Ineligible population |
| The Training of Medium- to Long-Distance Sprint | Nicholson, Ben; Dinsdale, Alex; Jones, Ben; Till, Kevin; | Ineligible population |

| | | |
|---|---|---|
| Performance in Football Code Athletes: A Systematic Review and Meta-analysis. | | |
| The treatment and prevention of obesity: a systematic review of the literature | Glenny, A. M., S. O'Meara, A. Melville, T. A. Sheldon and C. Wilson | Ineligible population |
| The WHO Health Promoting School framework for improving the health and well-being of students and their academic achievement | Langford, R., C. P. Bonell, H. E. Jones, T. Pouliou, S. M. Murphy, E. Waters, K. A. Komro, L. F. Gibbs, D. Magnus and R. Campbell | Ineligible population |
| Through the Looking Glass: A Systematic Review of Longitudinal Evidence, Providing New Insight for Motor Competence and Health | Lisa M. Barnett, E. Kipling Webster, Ryan M. Hulteen, An De Meester,Nadia C. Valentini, Matthieu Lenoir, Caterina Pesce,Nancy Getchell,Vitor P. Lopes,Leah E. Robinson, Ali Brian, Luis P. Rodrigues. | Ineligible population |
| Time Trends in Physical Activity Using Wearable Devices: A Systematic Review and Meta-analysis of Studies from 1995 to 2017 | SCOTT A. CONGER, LINDSAY P. TOTH, CHANNIE CRETSINGER, ANDERS RAUSTORP, JOSEF MITÁŠ, SHIGERU INOUE and DAVID R. BASSETT | Ineligible population |
| Toll like receptor expression induced by exercise in obesity and metabolic syndrome: A systematic review. | Rada, Isabel; Deldicque, Louise; Francaux, Marc; Zbinden-Foncea, Hermann | Ineligible population |
| Treatment Interventions for Early Childhood Obesity: A Systematic Review | Foster, B. A., J. Farragher, P. Parker and E. T. Sosa | Ineligible population |

| | | |
|---|---|---|
| Types of Interventions Targeting Dietary, Physical Activity, and Weight-Related Outcomes among University Students: A Systematic Review of Systematic Reviews | Katerina Belogianni and Christine Baldwin | Ineligible population |
| Understanding the Benefits of Brief Classroom-Based Physical Activity Interventions on Primary School-Aged Children's Enjoyment and Subjective Wellbeing: A Systematic Review | Nicole Papadopoulos, Ana Mantilla, Katherine Bussey, Chloe Emonson, Lisa Olive, Jane McGillvray, Caterina Pesce, Samantha Lewis, Nicole Rinehart. | Ineligible population |
| Universal and selective interventions to promote good mental health in young people: Systematic review and meta-analysis | Gonzalo Salazar de Pabloa, Andrea De Michelia,Dorien H. Niemane, Christoph U. Correllf, Lars Vedel Kessing, Andrea Pfennig, Andreas Bechdolfl, Stefan Borgwardt, Celso Arangoc, Therese van Amelsvoort,Eduard Vietas, Marco Solmia, Dominic Oliver,Ana Catalana, Valeria Verdinoa, Lucia Di Maggioa,Ilaria Bonoldiw, Julio Vaquerizo-Serrano, Ottone Baccaredda Boyw, Umberto Provenzania, Francesca Ruzzit, Federica Caloriot, Guido Nosariz, Benedetto Di Marcot, Irene Famularot, Silvia Moltenit, Eleonora Filosit, Martina Mensit,aa, Umberto Balottint, Pierluigi Politit, Jae Il Shinx, Paolo Fusar-Polia | Ineligible population |
| Use of Mobile Phone App Interventions to Promote Weight Loss: Meta-Analysis | Md Mohaimenul Islam, MSc, Tahmina Nasrin Poly, MSc, Bruno Andres Walther, PhD, and Yu-Chuan (Jack) Li, | Ineligible population |
| Wearable activity trackers for promoting physical activity: A systematic meta-analytic review | Caining Li, Xiaoyu Chen, Xinhua Bi | Ineligible population |
| Weight loss in children and adolescents | Muhlig, Y., M. Wabitsch, A. Moss and J. Hebebrand | Ineligible population |
| Weight-bearing exercise and bone mineral accrual in children and adolescents: a review of controlled trials | Hind, K. and M. Burrows | Ineligible population |
| Weight-related child behavioral interventions in Brazil: a systematic review | Marshall, S. J., E. J. Simoes, C. M. Eisenberg, C. K. Holub, E. M. Arredondo, S. Barquera and J. P. Elder | Ineligible population |
| What is the impact of structural and cultural factors and interventions within educational settings on promoting positive mental health and preventing poor mental health: a systematic review | David Troy, Joanna Anderson, Patricia E. Jessiman, Patricia N. Albers, Joanna G. Williams, Seamus Sheard, Emma Geijer‑Simpson, Liam Spencer, Eileen Kaner, Mark Limmer, Russell Viner and Judi Kidger | Ineligible population |
| Which type of sedentary behaviour intervention is more effective at reducing body mass index in children? A meta-analytic review | Liao, Y., J. Liao, C. P. Durand and G. F. Dunton | Ineligible population |
| Yoga as an Intervention for the Reduction of Symptoms of Anxiety and Depression in Children and Adolescents: A Systematic Review | Aurora James-Palmer, Ellen Z. Anderson, Lori Zucker, Yana Kofman and Jean-Francois Daneault | Ineligible population |

| | | |
|---|---|---|
| Components of primary care interventions to treat childhood overweight and obesity: a systematic review of effect | Sargent, G. M., L. S. Pilotto and L. A. Baur | Ineligible intervention |
| School‑based education programmes for the prevention of unintentional injuries in children and young people | Orton, E., J. Whitehead, J. Mhizha‑Murira, M. Clarkson, M. C. Watson, C. A. Mulvaney, J. U. L. Staniforth, M. Bhuchar and D. Kendrick | Ineligible intervention |
| Systematic review and meta-analysis of interventions targeting sleep and their impact on child body mass index, diet, and physical activity | Yoong, S. L., L. K. Chai, C. M. Williams, J. Wiggers, M. Finch and L. Wolfenden | Ineligible intervention |
| A meta-analysis of motivational interviewing interventions for pediatric health behavior change | Gayes, L. A. and R. G. Steele | Ineligible intervention |
| Characteristics of physical activity interventions and effects on cardiorespiratory fitness in children aged 6-12 years-A systematic review. | Braaksma, Petra; Stuive, Ilse; Garst, Rianne (M.E.); Wesselink, Carlijn F.; van der Sluis, Corry K.; Dekker, Rienk; Schoemaker, Marina M. | Ineligible intervention |
| Classroom Standing Desks and Sedentary Behavior: A Systematic Review | Minges, K. E., A. M. Chao, M. L. Irwin, N. Owen, C. Park, R. Whittemore and J. Salmon | Ineligible intervention |
| Digital Interventions to Improve Health Literacy Among Parents of Children Aged 0 to 12 Years With a Health Condition: Systematic Review | Evalotte Mörelius, Suzanne Robinson, Diana Arabiat, Lisa Whitehead. | Ineligible intervention |
| Effectiveness of intervention strategies exclusively targeting reductions in children's sedentary time: a systematic review of the literature | Altenburg, T. M., J. Kist-van Holthe and M. J. Chinapaw | Ineligible intervention |
| Effects of high-intensity interval training on executive functions in children and adolescents: A Systematic Review and Meta-analysis | Reyes-Amigo, Tomás; Bezerra, Andrea; Gomez-Mazorra, Mabel; Boppre, Giorjines; Martins, Clarice; Carrasco-Beltran, Hernaldo; Cordero-Roldan, Evelyn; Mota, Jorge | Ineligible intervention |
| Efficacy of Clinic-Based Telehealth vs. Face-to-Face Interventions for Obesity Treatment in Children and Adolescents in the United States and Canada: A Systematic Review | Andrea Whitley, MS, RDN, CSP, LD* and Najat Yahia, PhD, RDN, LD | Ineligible intervention |
| Identifying efective intervention strategies to reduce children's screen time: a systematic review and meta-analysis | Alexis Jones, Bridget Armstrong, R. Glenn Weaver, Hannah Parker, Lauren von Klinggraef and M. W. Beets | Ineligible intervention |
| Meta-analysis of lifestyle modification interventions addressing overweight and | David M. Janicke1 | Ineligible intervention |
| Motivational interviewing for treating overweight and obese youth: A systematic review | Vallabhan, M.K.; Jimenez, E.Y.; Kong, A.S.; | Ineligible intervention |
| Physical Activity Interventions Among American Indian and Alaska Native Persons: A | Molly C. Kokenge, MSN, RN, CEN, CNL1,2, Todd M. Ruppar, PhD, RN, FAHA, FAAN1, | Ineligible intervention |

| | | |
|---|---|---|
| Preventing and treating childhood overweight and obesity in children up to 5 years old: A systematic review by intervention setting | Angela C. Flynn1,2 \| Fatma Suleiman2 \| Hazel Windsor‐Aubrey2 \| | Ineligible intervention |
| Primary care prevention of cardiovascular risk behaviors in adolescents: A systematic review | Herv´e Tissot a, | Ineligible intervention |
| Research gaps in physical activity, sedentary behavior, and obesity among children in Qatar: A systematic review | Al Hammadi, E. M., S. Sayegh, F. Al Mudahka, I. Ibrahim and M. G. Al Kuwari | Ineligible intervention |
| Screening for Hypertension in Children and Adolescents: Systematic Review for the U.S. Preventive Services Task Force | Gerald Gartlehner, Emily B. Vander Schaaf, Colin Orr, Sara M. Kennedy, Rachel Clark, Meera Viswanathan | Ineligible intervention |
| Street connectivity, physical activity, and childhood obesity: A systematic review and meta‐analysis | Peng Jia, Yuxuan Zou, Zhifeng Wu, Dong Zhang, Tong Wu, Melody Smith, Qian Xiao | Ineligible intervention |
| The effectiveness of nurse-led interventions to prevent childhood and adolescent overweight and obesity: A systematic review of randomised trials | Lisa Whitehead, Istvan Kabdebo, Melissa Dunham, Robyn Quinn, Jennifer Hummelshoj, Cobie George, Elizabeth Denney-Wilson | Ineligible intervention |
| The Impact of Schoolyard Greening on Children's Physical Activity and Socioemotional Health: A Systematic Review of Experimental Studies | Jean C. Bikomeye, Joanna Balza and Kirsten M. Beyer | Ineligible intervention |
| The Physiological and Psychological Benefits of Dance and its Effects on Children and Adolescents: A Systematic Review | Dan Tao, Yang Gao, Alistair Cole, Julien S. Baker, Yaodong Gu, Rashmi Supriya, Tomas K. Tong, Qiuli Hu and Roger Awan-Scully | Ineligible intervention |
| Very-low-energy diets and morbidity: a systematic review of longer-term evidence | Mulholland, Y., E. Nicokavoura, J. Broom and C. Rolland | Ineligible intervention |
| High-intensity interval training in overweight and obese children and adolescents: systematic review and meta-analysis | Thivel, D., J. Masurier, G. Baquet, B. W. Timmons, B. Pereira, S. Berthoin, M. Duclos and J. Aucouturier | Ineligible study duration |
| What works in school-based energy balance behaviour interventions and what does not? A systematic review of mediating mechanisms | van Stralen, M. M., M. Yildirim, S. J. te Velde, J. Brug, W. van Mechelen and M. J. Chinapaw | Ineligible study duration |
| A Meta-Analysis of High-Intensity Interval Training on Glycolipid Metabolism in Children With Metabolic Disorders | Meng Cao, Shu Li, Yucheng Tang and Yu Zou | Ineligible study duration |
| A Meta-Analytic Review of Obesity Prevention in the Schools: 1997-2008 | Cook-Cottone, C., C. M. Casey, T. H. Feeley and J. Baran | Ineligible study duration |
| A Sex/Gender Perspective on Interventions to Reduce Sedentary Behaviour in Girls and Boys: Results of the genEffects Systematic Review | Catherina Vondung,Yolanda Demetriou, Anne K. Reimers, Annegret Schlund, and Jens Bucksch | Ineligible study duration |
| A systematic review of studies using pedometers to promote physical activity among youth | Lubans, D. R., P. J. Morgan and C. Tudor-Locke | Ineligible study duration |

| | | |
|---|---|---|
| A systematic review of the implementation of obesity prevention interventions in early childcare and education settings using the RE-AIM framework | Jennifer C. Sanchez-Flack,Annie Herman,Joanna Buscemi, Angela Kong, Alexis Bains, Marian L. Fitzgibbon | Ineligible study duration |
| Best practice dietetic management of overweight and obese children and adolescents: A 2010 update of a systematic review | Ho, M., M. E. Jensen, T. Burrows, M. Neve, S. P. Garnett, L. Baur, L. Stewart and C. Collins | Ineligible study duration |
| Can active video games be part of the solution to promote physical activity in youth? A systematic review | Carmo, J. L. F. and A. L. Palmeira | Ineligible study duration |
| Differences in Physical Fitness and Body Composition Between Active and Sedentary | Saša Đuric4 \| Žan Luca Potocˇnik4 \| Maroje Soric1,4 | Ineligible study duration |
| Effect of Active Lessons on Physical Activity, Academic, and Health Outcomes: A Systematic Review | Martin, R. and E. M. Murtagh | Ineligible study duration |
| Effectiveness of interventions on physical activity in overweight or obese children: a systematic review and meta-analysis including studies with objectively measured outcomes | Nooijen, C. F., M. R. Galanti, K. Engstrom, J. Moller and Y. Forsell | Ineligible study duration |
| Face-to-face physical activity incorporated into dietary intervention for overweight/obesity in children | Jing Hong Liang, Yu Zhao, Yi Can Chen, Nan Jiang, Shu Xin Zhang, Shan Huang, Aerziguli Kakaer and Ya Jun Chen | Ineligible study duration |

| | | |
|---|---|---|
| and adolescents: a Bayesian network meta-analysis | | |
| Factors that alter body fat, body mass, and fat-free mass in pediatric obesity | LeMura, L. M. and M. T. Maziekas | Ineligible study duration |
| Family-based interventions to increase physical activity in children: a systematic review, meta-analysis and realist synthesis | Brown, H. E., A. J. Atkin, J. Panter, G. Wong, M. J. Chinapaw and E. M. van Sluijs | Ineligible study duration |
| Family-focused physical activity, diet and obesity interventions in African-American girls: a systematic review | Barr-Anderson, D. J., A. W. Adams-Wynn, K. I. DiSantis and S. Kumanyika | Ineligible study duration |
| High-Intensity Interval Training Interventions in Children and Adolescents: A Systematic Review | Eddolls, W. T. B., M. A. McNarry, G. Stratton, C. O. N. Winn and K. A. Mackintosh | Ineligible study duration |
| Physical activity and exercise interventions for childhood obesity: A systematic review with meta-analysis of randomized clinical trials | Cesa, C. C., G. Sbruzzi, B. Eibel, S. M. Barbiero, R. D. O. Petkowicz, R. D. V. Marques, N. B. Machado, W. B. De Souza, B. D. Schaan and L. C. Pellanda | Ineligible study duration |
| Physical Activity for Cognitive and Mental Health in Youth: A Systematic Review of Mechanisms | Lubans, D., J. Richards, C. Hillman, G. Faulkner, M. Beauchamp, M. Nilsson, P. Kelly, J. Smith, L. Raine and S. Biddle | Ineligible study duration |

| | | |
|---|---|---|
| School-related sedentary behaviours and indicators of health and well-being among children and youth: a systematic review | Nicholas Kuzik, Bruno G. G. da Costa, Yeongho Hwang, Simone J. J. M. Verswijveren, Scott Rollo,Mark S. Tremblay, Stacey Bélanger, Valerie Carson, Melanie Davis, Susan Hornby, Wendy Yajun Huang, Barbi Law, Jo Salmon, Jennifer R. Tomasone, Lucy‑Joy Wachira, Katrien Wijndaele and Travis J. Saunders | Ineligible study duration |
| Systematic review of acute physically active learning and classroom movement breaks on children's physical activity, cognition, academic performance and classroom behaviour: understanding critical design features | Daly-Smith, A. J., S. Zwolinsky, J. McKenna, P. D. Tomporowski, M. A. Defeyter and A. Manley | Ineligible study duration |
| The effectiveness and promising strategies of obesity prevention and treatment programmes among adolescents from disadvantaged backgrounds: a systematic review | Kornet-van der Aa, D. A., T. M. Altenburg, C. H. van Randeraad-van der Zee and M. J. Chinapaw | Ineligible study duration |
| A systematic review of school-based interventions to prevent risk factors associated with noncommunicable diseases | Saraf, D. S., B. Nongkynrih, C. S. Pandav, S. K. Gupta, B. Shah, S. K. Kapoor and A. Krishnan | Ineligible outcome |
| Exercise targeted at the LIPOXmax for overweight or obesity: A meta-analysis | Romain, A. J., C. Fedou, J. F. Brun, A. Avignon and J. Mercie | Ineligible outcome |
| Obesity prevention interventions for middle school-age children of ethnic minority: a review of the literature | Stevens, C. J. | Ineligible outcome |
| Physical activity interventions in Hispanic American girls and women | Sharma, M. | Ineligible outcome |
| Strategies for enhancing the implementation of school-based policies or practices targeting risk factors for chronic disease | Wolfenden, L., N. K. Nathan, R. Sutherland, S. L. Yoong, R. K. Hodder, R. J. Wyse, T. Delaney, A. Grady, A. Fielding, F. Tzelepis, T. Clinton-McHarg, B. Parmenter, P. Butler, J. Wiggers, A. Bauman, A. Milat, D. Booth and C. M. Williams | Ineligible outcome |
| A prática da capoeira no ambiente escolar para a formação integral do aluno: uma revisão sistemática. / The practice of capoeira in the school environment for student integral formation: A systematic review. | Juliana Terra Ribeiro, Luis Felipe Milano Teixeira, Fabrício Teixeira Garramona | Ineligible outcome |
| A Systematic Review and Meta-Analysis of the Effects of Outdoor Education Programs on Adolescents' Self-Efficacy | Fang, Bin-Bin; Lu, Frank J. H.; Gill, Diane L.; Liu, Sean H.; Chyi, Theresa; Chen, Bin | Ineligible outcome |
| A systematic review exploring body image programmes and interventions in physical education | Kerner, Charlotte; Prescott, Amy; Smith, Robyn; Owen, Michael | Ineligible outcome |

| | | |
|---|---|---|
| A systematic review of physical activity promotion in children and adolescents by community health workers | Paulo Henrique Guerra, Rafael Silvestre, Tiago Henrique Toldo de Mello, Ana Luiza Babo Sedlacek Carvalho, Filipe Ferreira da Costa, Alex Antonio Florindo | Ineligible outcome |
| Classroom-based physical activity and sedentary 2 behavior interventions in adolescents: a systematic review and meta-analysis. Journal of Physical 3 Activity and Health. | McMichan, L., Gibson, A-M., & Rowe, D. A. | Ineligible outcome |
| Do behaviour change techniques promote physical activity in preschool children? A systematic review | Mosfer A. Al-walah, Michael Donnelly, Conor Cunningham & Neil Heron | Ineligible outcome |
| Effect of classroom-based physical activity interventions on attention and on-task behavior in schoolchildren: A systematic review | Stefanie Ruhland, Klaus W. Lange | Ineligible outcome |
| Effect of Warm-up on Fitness Performance of Schoolchildren. A Systematic Review | Fernandez-Agullo, Ruben; Merino-Marban, Rafael | Ineligible outcome |
| Effectiveness of active school transport interventions: a systematic review and update | Larouche, R., G. Mammen, D. A. Rowe and G. Faulkner | Ineligible outcome |
| Effectiveness of school-based interventions promoting physical activity and reducing sedentary time among children: A meta-analysis of | J. Rodrigo-Sanjoaquín | Ineligible outcome |
| accelerometerâ€assessed controlled trials. | | |
| Effects of physical activity interventions on cognitive performance of overweight or | Maria A. Barceló2,3 \| Ivan Radman1 \| Gregor Starc4 \| Gregor Jurak4 \| | Ineligible outcome |
| Effects of physical education interventions on academic performance and cognition in children and adolescents | Antonio García-Hermoso et al. | Ineligible outcome |
| Effects of school nurse-led interventions in promoting physical activity and reducing sedentary behaviours in children and adolescents: a systematic review with meta-analysis | Yari Longobucco et al. | Ineligible outcome |
| Fundamental movement skill interventions in youth: a systematic review and meta-analysis | Morgan, P. J., L. M. Barnett, D. P. Cliff, A. D. Okely, H. A. Scott, K. E. Cohen and D. R. Lubans | Ineligible outcome |
| Getting the fundamentals of movement: a meta-analysis of the effectiveness of motor skill interventions in children | Logan, S. W., L. E. Robinson, A. E. Wilson and W. A. Lucas | Ineligible outcome |
| Impact of Physical Activity-Based Interventions on Children and Adolescents' Physical Self-Concept: A Meta-Analysis | David Zamorano-García, Álvaro Infantes-Paniagua, Ricardo Cuevas-Campos & Juan Gregorio Fernández-Bustos | Ineligible outcome |
| It's Not Just What You Do but the Way You Do It: A Systematic Review of Process Evaluation of Interventions to | Ma, Jiani; Lander, Natalie; Eyre, Emma L. J.; Barnett, Lisa M.; Essiet, Inimfon A.; Duncan, Michael J. | Ineligible outcome |

| | | |
|---|---|---|
| Improve Gross Motor Competence. | | |
| Mental health and socioeconomic status impact adherence to youth activity and dietary programs: a meta-analysis | Mark E. Lemstra, Marla Rogers | Ineligible outcome |
| Neurobiological evidence of physical activity interventions on mental health outcomes and cognition in youth: a systematic review of randomised controlled trials | Heinze K, Cumming J, Dosanjh A, Palin S, Poulton S, Bagshaw AP, Broome MR | Ineligible outcome |
| Physical activity interventions targeting perceived body image among adolescent girls | CHIA-LIANG DAI; SHARMA, MANOJ; CHING-CHEN CHEN; LOCKLEY, HALEY | Ineligible outcome |
| Physical Activity, Fitness, School Readiness, and Cognition in Early Childhood: A Systematic Review | Christine W. St. Laurent,Sarah Burkart, Chloe Andre, Rebecca M.C. Spencer | Ineligible outcome |
| Play Smart, Be Smart? Effect of Cognitively Engaging Physical Activity Interventions on Executive Function among Children 4~12 Years Old: A Systematic Review and Meta-Analysis | Wenjing Song, Leyi Feng, Junwei Wang, Feifei Ma, Jiebo Chen, Sha Qu and Dongmei Luo | Ineligible outcome |
| School-Based Physical Activity Interventions in Prevocational Adolescents: A Systematic Review and Meta-Analyses | Joannis H. van de Kop, M.Sc., Willem G. van Kernebeek, M.Sc., Rene H. J. Otten, M.Sc., Huub M. Toussaint, Ph.D., and Arnoud P. Verhoeff, Prof | Ineligible outcome |
| School-based physical activity interventions in rural and urban/suburban communities: A systematic review and meta-analysis | Christopher D. Pfledderer, Ryan D. Burns, Wonwoo Byun, Russell L. Carson, Gregory J. Welk, Timothy A. Brusseau | Ineligible outcome |
| Social influence in childhood obesity interventions: a systematic review | Jalali, M. S., Z. Sharafi-Avarzaman, H. Rahmandad and A. S. Ammerman | Ineligible outcome |
| The effect of social media interventions on physical activity and dietary behaviours in young people and adults: a systematic review | Victoria A. Goodyear* , Grace Wood, Bethany Skinner and Janice L. Thompson | Ineligible outcome |
| The effects of the Nordic hamstring exercise on sprint performance and eccentric knee flexor strength: A systematic review and meta-analysis of intervention studies among team sport players. | Bautista, Iker J.; Vicente-Mampel, Juan; Baraja-Vegas, Luis; Segarra, Víctor; Martín, Fernando; Van Hooren, Bas | Ineligible outcome |
| What Is Known about School-Based Interventions for Health Promotion and Their Impact in Developing Countries? A Scoping Review of the Literature | Mukamana, O. and M. Johri | Ineligible outcome |
| Assessing the cost effectiveness of public health interventions to prevent obesity: A systematic review of the effectiveness of 16 obesity prevention interventions | Paech, D., K. Coleman, P. J. Mernagh and A. R. Weston | Article not found |

| | | |
|---|---|---|
| Effective behavioural components in child and adolescent weight management programmes | Sahota, P., J. Wordley and J. Woodward | Article not found |
| Effectiveness of universal self-regulation-based interventions to improve self-regulation, and effects on distant health and social outcomes in children and adolescents: A systematic review and meta-analysis | Pandey, A., D. Hale, S. Das, A. L. Goddings, S. J. Blakemore and R. Viner | Article not found |
| Effects of school-based interventions on body mass index in children and adolescents: A systematic overview and meta-analysis | Park, J. J., R. Bakhet, D. Karl, F. Yuan and E. Lonn | Article not found |
| Quantitative analysis of single- vs. multiple-set programs in resistance training | Wolfe, B. L., L. M. LeMura and P. J. Cole | Article not found |
| A Meta-analysis Of The Effect Of School-based Physical Activity Intervention On Students' Mvpa | Yanfei Sun, Zhaohua Zhang, Liu Zhang, Hongjuan Li | Article not found |
| Activity- and Occupation-Based Interventions to Support Mental Health, Positive Behavior, and Social Participation for Children and Youth: A Systematic Review | Susan M Cahill, Brad E Egan, Joanna Seber | Article not found |
| Adherence to mind-body therapies: A systematic review and meta-analysis | M. Shaw, M. Park, R.R. Bannuru, C. Wang. | Article not found |

| | | |
|---|---|---|
| Does exercise beneficially affect sex hormones when added to hypo-caloric diets in adults with overweight or obesity? A systematic review and meta-analysis of controlled clinical trials | Sahar Mohseni-Takalloo | Article not found |
| Does Regular Exercise Impact the Lung Function of Healthy Children and Adolescents? A Systematic Review and Meta-Analysis | Balbinot F, Claudino FCA, Lucas PK, Martins APD, Wendland EM, Gerbase MW | Article not found |
| Effect of high-intensity interval training on cardiometabolic risk factors in childhood obesity: a meta-analysis | Zhu L, Liu J, Yu Y, Tian Z | Article not found |
| Effect Of School-based Physical Activity Programs On Hamstring Flexibility: A Meta-analysis | Sonia Montero-Briceño, Isaura M. Castillo-Hernández | Article not found |
| Effectiveness of peer-led programs for overweight and obesity in children: systematic review and meta-analysis | Ngoc-Minh Nguyen | Article not found |
| Effects Of Classroom Active Breaks On Academic-related Outcomes In Children: A Systematic Review And Meta-analysis | NR | Article not found |
| Effects of Different Movement Programs on Motor Competence: A Systematic Review With Meta-Analysis. | Jiménez-Díaz J, Chaves-Castro K, Salazar W | Article not found |

| | | |
|---|---|---|
| Effects of Strength Training on Blood Pressure and Heart Rate Variability—A Systematic Review. | Corso, Marcelo \| de Figueiredo, Tiago C \| Carvalho, Danilo \| Brown, Amanda F \| de Salles, Belmiro F \| Simao, Roberto \| Willardson, Jeffrey M \| Dias, Ingrid | Article not found |
| Empowerment Approaches in Childhood Weight Management: A Systematic Review | Earle R, Littlewood R, Nalatu S, Walker J. | Article not found |
| Impact of dietary and exercise interventions on anthropometric and metabolic outcomes in obese children and adolescents: A systematic review and metaanalysis of randomised controlled trials | Ho, M., S. Garnett, L. Baur, T. Burrow, L. Stewart and C. Collins | Article not found |
| Interventions for Increasing Physical Activity in Low-Income, Ethnic Minority Children and Youths: Meta Analysis | Kyungun Kim, Seung Ho Chang, Jihyun Lee, Sukho Lee | Article not found |
| Physical Exercise and Brain-Derived Neurotrophic Factor Concentration in Children and Adolescents: A Systematic Review With Meta-Analysis | Francisco José de Menezes-Junior, Íncare C Jesus, Caroline Brand, Jorge Mota, Neiva Leite | Article not found |
| Responses of cardiorespiratory variables in crossover interventions of land running and deep-water running | Assunção, Y.; Santos, R.; Lima, G.; Gomes, I.; Silva, D.; Fortunato, I.; Matias, E.; Brito, A.; | Article not found |
| THE EFFECT OF PLYOMETRIC TRAINING | NR | Article not found |
| ON RUNNING ECONOMY IN LONG DISTANCE RUNNERS | | |
| Treating the ticking time bomb: Childhood obesity | Ells, L.J.; Rees, K.; Brown, T.; Mead, E.; McGeechan, G.; Farpour-Lambert, N.; Al-Kudairy, L.; Clements, H.; Loveman, E.; Baur, L.; Rayco-Solon, P.; Demaio, A.; Azevedo, L.; | Article not found |
| Treatments for childhood and adolescent obesity | Haddock, C. K., W. R. Shadish, R. C. Klesges and R. J. Stein | Article not found |

**eTable 3. Total outcomes.**

| Study (Year) | Domain | Generic outcome (as stated by the authors) | Outcome | Outcome measure |
|---|---|---|---|---|
| Aceves-Martins et al., 2022 | General | Anthropometry | Body mass index (BMI) | NR |
| Al-Khudairy et al., 2017 | Delivery of Care | Participants' views of the intervention | Participants' views of the intervention | Defined as documented accounts from participant feedback and measured at baseline and at least at six months (e.g. levels of satisfaction) |
| | Emotional Functioning/Wellbeing | Self-esteem | Self-esteem | Evaluated by a validated instrument such as Rosenberg Self-Esteem Scale and measured at baseline and at least at six months |
| | General | Anthropometric measures other than BMI | Body fat mass (% and kg) | Measured by bioelectrical impedance analysis or DXA or MRI scan |
| | | Changes in measured BMI or body weight | Body mass index (BMI) | KG/m² or BMI z score |
| | | Changes in measured BMI or body weight | Body weight | NR |
| | | Anthropometric measures other than BMI | Lean body mass | Measured by bioelectrical impedance analysis |
| | | Anthropometric measures other than BMI | Skinfold thickness | NR |
| | | Anthropometric measures other than BMI | Waist circumference | NR |
| | | Anthropometric measures other than BMI | Waist-to-hip ratio | NR |
| | Global Quality of Life | Health-related quality of life | Quality of life | Health-related quality of life: Paediatric Quality of Life Inventory |

| Study (Year) | Domain | Generic outcome (as stated by the authors) | Outcome | Outcome measure |
|---|---|---|---|---|
| | Metabolism and Nutrition | Behavior change of diet | Dietary behavior | Defined as validated measures of diet measured by either objective (e.g. Mandometer) or validated self-reported questionnaires |
| | | Behavior change of diet | Dietary intake | Defined as validated measures of diet as food records |
| | Physical Functioning | Behavior change of physical activity | Physical activity behavior | Defined as validated measures as self-reported measures or objective measures (e.g hours spent viewing television) |
| | | Behavior change of physical activity | Physical activity level | Assessed using objective tools or validated selfreported tools |
| | Social Functioning | Parenting skill and relationships | Parenting skill and relationships | Evaluated by a validated instrument and measured at baseline and at least at six months (e.g. parent-adolescent communication) |
| Albornoz-Guerrero et al., 2021 | Cardiac | | Blood Pressure | manual blood pressure cuff, automatic sphygmomanometer |
| | Emotional Functioning/Wellbeing | | Mood | the scale of intinsic versus extrinsic orientation in the classroom |
| | | | Self-perception | self-perceptions profile questionnaire for children, the child and youth physical self-perception profile questionnaire |
| | | | Self-steem | self-perceptions profile questionnaire for children, the child and youth physical self-perception profile questionnaire |
| | General | | Body composition | bio impedance, anthropometry, DXA, caliper, three-component model of body composition |
| | | | Body fat | bio impedance, anthropometry, DXA, caliper, three-component model of body composition |
| | | | Body mass index (BMI) | kg/m2 |
| | | | Hip circumference | bio impedance, anthropometry, DXA, caliper, three-component model of body composition |

| Study (Year) | Domain | Generic outcome (as stated by the authors) | Outcome | Outcome measure |
|---|---|---|---|---|
| | | | skinfold | bio impedance, anthropometry, DXA, caliper, three-component model of body composition |
| | | | strenght and difficulties percieved by parents | the questionnaire of streghts and difficulties of the child resported by the parents |
| | | | Waist Circunference | bio impedance, anthropometry, DXA, caliper, three-component model of body composition |
| | | | waist-to-heigh ratio | bio impedance, anthropometry, DXA, caliper, three-component model of body composition |
| | | | weight | bio impedance, anthropometry, DXA, caliper, three-component model of body composition |
| | Global Quality of Life | | Health-related QOL | pediatric QOL questionnaire, the child health questionnaire, the QOL questionnaire for children between 7 and 13 year old |
| | Metabolism and Nutrition | | Eating behaviour | food frequency questionnaire, 3-day diet recall form, 24h-dietary recall, Block Kids Questionnaire, Dutch eating behaviour questionnaire for children, The Family Eating and Activiy Habits questionnaire, Day in life questionnaire, 2-day nutritional dietary record. |
| | | | Food intake | food frequency questionnaire, 3-day diet recall form, 24h-dietary recall, Block Kids Questionnaire, Dutch eating behaviour questionnaire for children, The Family Eating and Activiy Habits questionnaire, Day in life questionnaire, 2-day nutritional dietary record. |
| | Physical Functioning | | Aerobic fitness | accelerometry, pedometer, Physical Activity Questionnaire for Children, Leisure Score Index, walking test, handgrip strenght test, ergospirometry test, jumping test |

| Study (Year) | Domain | Generic outcome (as stated by the authors) | Outcome | Outcome measure |
|---|---|---|---|---|
| | | | Muscle strength | bio impedance, anthropometry, DXA, caliper, three-component model of body composition |
| | | | physical activity level | accelerometry, pedometer, Physical Activity Questionnaire for Children, Leisure Score Index, walking test, handgrip strenght test, ergospirometry test, jumping test |
| | | | sedentary behaviour | accelerometry, pedometer, Physical Activity Questionnaire for Children, Leisure Score Index, walking test, handgrip strenght test, ergospirometry test, jumping test |
| | Psychiatric | | Anxiety | the multidimensional anxiety scale for children |
| | | | Depression | children's depression scale |
| Álvarez-Bueno et al., 2017 | Cognitive Functioning | Performance in core executive functions | Cognitive flexibility | Performance in core executive working cognitive flexibility |
| | | Metacognition | Metacognition | metacognition: including higher-level executive functions and cognitive life skills |
| | | Performance in non-executive cognitive function | Non-executive cognitive function | Performance in non-executive cognitive function (NR which non-executive functions) |
| | | Performance in core executive functions | Selective attention-inhibition | Performance in core executive working selective attention-inhibition |
| | | Performance in core executive functions | Working memory | Performance in core executive working memorie |
| Andrade et al., 2018 | Blood and Lymphatic System | biochemical measurements | Cholesterol | NR |
| | | biochemical measurements | Triglycerides | NR |
| | Cardiac | biochemical measurements | Blood pressure | NR |
| | General | anthropometric measurements | Body mass index (BMI) | NR |

| **Study (Year)** | **Domain** | **Generic outcome (as stated by the authors)** | **Outcome** | **Outcome measure** |
|---|---|---|---|---|
| | | anthropometric measurements | waist circumference | NR |
| Andrade et al., 2019 | Emotional Functioning/Wellbeing | Psychological effects | Athletic competence | scales, questionnaires / survey. |
| | | Psychological effects | Behavioral conduct | scales, questionnaires / survey. |
| | | Psychological effects | Intrinsic motivation | scales, questionnaires / survey. |
| | | Psychological effects | Liking to enjoy the exergame | scales, questionnaires / survey. |
| | | Psychological effects | Perceived competence | scales, questionnaires / survey. |
| | | Psychological effects | Physical competence | scales, questionnaires / survey. |
| | | Psychological effects | Positive expectations | scales, questionnaires / survey. |
| | | Psychological effects | Psychological attraction to exergame | scales, questionnaires / survey. |
| | | Psychological effects | Satisfaction | scales, questionnaires / survey. |
| | | Psychological effects | Self-concept (self-steem) | scales, questionnaires / survey. |
| | | Psychological effects | Self-efficacy | scales, questionnaires / survey. |
| | | Psychological effects | Self-worth (self-steem) | scales, questionnaires / survey. |
| | Psychiatric | Psychological effects | Anxiety | scales, questionnaires / survey. |
| | | Psychological effects | Depression | scales, questionnaires / survey. |
| | Social Functioning | Psychological effects | Externnalizing composite | scales, questionnaires / survey. |
| | | Psychological effects | Interpersonal relations | scales, questionnaires / survey. |
| | | Psychological effects | Peer support | scales, questionnaires / survey. |
| | | Psychological effects | Relationship with parents | scales, questionnaires / survey. |

| Study (Year) | Domain | Generic outcome (as stated by the authors) | Outcome | Outcome measure |
|---|---|---|---|---|
| | | Psychological effects | Social acceptance | scales, questionnaires / survey. |
| | | Psychological effects | Social stress | scales, questionnaires / survey. |
| Azevedo et al., 2023 | General | BMI and BMI z-score | Body mass index (BMI) | objectively measured (not-self-reported) weight and height |
| | | BMI and BMI z-score | Body mass index (BMI) | objectively measured (not-self-reported) weight and height |
| Bento et al., 2021 | General | body composition | body fat % | NR |
| | | body composition | body fat mass | NR |
| | | body composition | body mass | NR |
| | | body composition | Body mass index (BMI) | NR |
| | | body composition | Lean body mass | NR |
| | | body composition | sum of skinfolds, | NR |
| | | body composition | waist circumference | NR |
| | Physical Functioning | physical fitness | Cardiorespiratory fitness | NR |
| | | physical fitness | Countermovement jump | NR |
| | | physical fitness | Funcional obstacle performance | NR |
| | | physical fitness | Lower-body muscular strength | NR |
| | | physical fitness | Maximal aerobic speed | NR |
| | | physical fitness | squat jump | NR |
| | | physical fitness | VO2 | NR |
| Bento et al., 2022 | Emotional Functioning/Wellbeing | Physical Fitness | Benefits | 8-item questionnaire |
| | | Physical Fitness | Competence support | 8-item questionnaire |
| | | Physical Fitness | Effort | 8-item questionnaire |

| Study (Year) | Domain | Generic outcome (as stated by the authors) | Outcome | Outcome measure |
|---|---|---|---|---|
| | | Physical Fitness | Enjoyment | 8-item questionnaire |
| | | Physical Fitness | Fun | 8-item questionnaire |
| | | Physical Fitness | Importance | 8-item questionnaire |
| | | Physical Fitness | Intrinsic motivation | 8-item questionnaire |
| | | Physical Fitness | Pleasure | 8-item questionnaire |
| | | Physical Fitness | Restlessness | 8-item questionnaire |
| | | Physical Fitness | Satisfaction related to participation in PA | 8-item questionnaire |
| | General | Physical Fitness | Body fat | Body composition: waist circumference; body mass; body fat; lean body mass; sum of skinfolds; Power, assessed through jump tests; |
| | | Physical Fitness | Body mass | Body composition: waist circumference; body mass; body fat; lean body mass; sum of skinfolds; Power, assessed through jump tests; |
| | | Physical Fitness | Body mass index (BMI) | Body composition: waist circumference; body mass; body fat; lean body mass; sum of skinfolds; Power, assessed through jump tests; |
| | | Physical Fitness | Body mass index (BMI) | Body composition: waist circumference; body mass; body fat; lean body mass; sum of skinfolds; Power, assessed through jump tests; |
| | | Physical Fitness | Lean body mass | Body composition: waist circumference; body mass; body fat; lean body mass; sum of skinfolds; Power, assessed through jump tests; |
| | | Physical Fitness | Sum of skinfolds | Body composition: waist circumference; body mass; body fat; lean body mass; sum of skinfolds; Power, assessed through jump tests; |

| Study (Year) | Domain | Generic outcome (as stated by the authors) | Outcome | Outcome measure |
|---|---|---|---|---|
| | Physical Functioning | Physical Fitness | Agility | Body composition: waist circumference; body mass; body fat; lean body mass; sum of skinfolds; Power, assessed through jump tests; |
| | | Physical Fitness | Power | Body composition: waist circumference; body mass; body fat; lean body mass; sum of skinfolds; Power, assessed through jump tests; |
| | | Physical Fitness | Running speed | Body composition: waist circumference; body mass; body fat; lean body mass; sum of skinfolds; Power, assessed through jump tests; |
| | | Physical Fitness | Upper strength | Body composition: waist circumference; body mass; body fat; lean body mass; sum of skinfolds; Power, assessed through jump tests; |
| | | Physical Fitness | Moderate Physical Activity | assessed PA subjectively through questionnaires, whereas in the other two studies, it was objectively measured using accelerometers. |
| | | Physical Fitness | Reducing sedentady activity | assessed PA subjectively through questionnaires, whereas in the other two studies, it was objectively measured using accelerometers. |
| | | Physical Fitness | Total Physical Activity | assessed PA subjectively through questionnaires, whereas in the other two studies, it was objectively measured using accelerometers. |
| | | Physical Fitness | Vigorous Physical Activity | assessed PA subjectively through questionnaires, whereas in the other two studies, it was objectively measured using accelerometers. |
| Birch et al., 2019 | Blood and Lymphatic System | Lipid Profile | Lipid profile | NR |
| | Cardiac | Blood Pressure | Blood Pressure | NR |

| Study (Year) | Domain | Generic outcome (as stated by the authors) | Outcome | Outcome measure |
|---|---|---|---|---|
| | Endocrine | Glucose, Inflammation, Insulin sensitivity/resistance, liver function | Alanine aminotransferase (ALT) | NR |
| | | Glucose, Inflammation, Insulin sensitivity/resistance, liver function | C-reactive protein | NR |
| | | Glucose, Inflammation, Insulin sensitivity/resistance, liver function | Glucose | NR |
| | | Glucose, Inflammation, Insulin sensitivity/resistance, liver function | HOMA-index (Homeostatic Model Assessment for Insulin Resistance) | NR |
| | | Glucose, Inflammation, Insulin sensitivity/resistance, liver function | IL-6 | NR |
| | | Glucose, Inflammation, Insulin sensitivity/resistance, liver function | Inflammation | NR |
| | | Glucose, Inflammation, Insulin sensitivity/resistance, liver function | Insulin | NR |
| | | Glucose, Inflammation, Insulin sensitivity/resistance, liver function | Insulin sensitivity/resistance | NR |
| | | Glucose, Inflammation, Insulin sensitivity/resistance, liver function | liver function | NR |

| Study (Year) | Domain | Generic outcome (as stated by the authors) | Outcome | Outcome measure |
|---|---|---|---|---|
| | General | BMI measures and adiposity measures | Body fat-SDS | NR |
| | | BMI measures and adiposity measures | Body mass | NR |
| | | BMI measures and adiposity measures | Body mass index (BMI) | NR |
| | | BMI measures and adiposity measures | Fat | NR |
| | | BMI measures and adiposity measures | Fat mass | NR |
| | | BMI measures and adiposity measures | Fat-free mass | NR |
| | | BMI measures and adiposity measures | Percentace body fat | NR |
| | | BMI measures and adiposity measures | Waist circumference | NR |
| | | BMI measures and adiposity measures | Waist circumference-SDS | NR |
| Boff et al., 2017 | General | Weight loss | Body mass index (BMI) | BMI through Z-scores or BMI or BMI percentile |
| | | Weight loss | Body weight | NR |
| Bondyra-Wisniewska et al., 2021 | Blood and Lymphatic System | Lipid Parameters | High density lipoprotein (HDL) | NR |
| | | Lipid Parameters | Low density lipoprotein (LDL) | NR |
| | | Lipid Parameters | Total cholesterol | NR |

| Study (Year) | Domain | Generic outcome (as stated by the authors) | Outcome | Outcome measure |
|---|---|---|---|---|
| | | Lipid Parameters | Triglycerides | NR |
| | Cardiac | Blood Pressure | Diastolic blood pressure | NR |
| | | Blood Pressure | Systolic blood pressure | NR |
| | General | Anthropometric | Body mass index (BMI) | NR |
| | | Anthropometric | zBMI | NR |
| Braaksmaa et al., 2018 | Physical Functioning | Cardiorespiratory Fitness | Cardiorespiratory fitness | Measured by different tests as Eurofit endurance shuttle run test or 20 m shuttle run test or maximal graded cycle ergometer test or maximal graded treadmill test or 6 minute walk test or PACER (progressive aerobic cardiovascular endurance run) or YAPHV (years of age to peak height velocity (skeletal maturity)) or VO2peak or yo-yo intermittent recovery level 1 or yo–yo intermittent endurance test |
| Brown et al., 2015 | General | Obesity outcomes | Body mass index (BMI) | Body mass index (kg/m²) or body mass index z-score (zBMI) |
| | | Obesity outcomes | Body weight | NR |
| | | Obesity outcomes | Waist circumference | NR |
| Brown et al., 2019 | General | obesity | Per cent fat content | NR |
| | | obesity | Ponderal index | NR |
| | | obesity | Prevalence of overweight and obesity | NR |
| | | obesity | Skin-fold thickness | NR |
| | | obesity | Weight and height | NR |
| | | obesity | zBMI score/BMI | Weight and height |
| Burns et al., 2019 | Endocrine | Insuline sensitivity | Fasting glucose | NR |

| Study (Year) | Domain | Generic outcome (as stated by the authors) | Outcome | Outcome measure |
|---|---|---|---|---|
| | | Insuline sensitivity | Fasting insulin | NR |
| Busnatu et al., 2022 | Blood and Lymphatic System | Cardiometabolic Factors | Glucose | NR |
| | | Cardiometabolic Factors | High density lipoprotein (HDL) | NR |
| | | Cardiometabolic factors | Low density lipoprotein (LDL) | NR |
| | Cardiac | Cardiometabolic Factors | Diastolic blood pressure | NR |
| | | Cardiometabolic Factors | Systolic blood pressure | NR |
| | General | Cardiometabolic Factors | Body mass index (BMI) | NR |
| | | Cardiometabolic Factors | Body weight | NR |
| | | Cardiometabolic Factors | Waist circumference | NR |
| Cao et al., 2019 | Physical Functioning | Cardiorespiratory Fitness | VO2max | yoyo test; 20 meters shuttle run test |
| Cao et al., 2021 | Blood and Lymphatic System | Cardiometabolic risk factors | High density lipoprotein (HDL) | NR |
| | | Cardiometabolic risk factors | Low density lipoprotein (LDL) | NR |
| | | Cardiometabolic risk factors | Total cholesterol | NR |
| | | Cardiometabolic risk factors | Triglycerides | NR |
| | Cardiac | Cardiometabolic risk factors | Diastolic blood pressure | NR |
| | | Cardiometabolic risk factors | Systolic blood pressure | NR |
| | Endocrine | Cardiometabolic risk factors | Glucose | NR |
| | | Cardiometabolic risk factors | HOMA-index (Homeostatic Model | NR |

| Study (Year) | Domain | Generic outcome (as stated by the authors) | Outcome | Outcome measure |
|---|---|---|---|---|
| | | | Assessment for Insulin Resistance) | |
| | | Cardiometabolic risk factors | Insulin | NR |
| | General | Cardiometabolic risk factors | body fat % | NR |
| | | Cardiometabolic risk factors | Body mass | NR |
| | | Cardiometabolic risk factors | Body mass index (BMI) | NR |
| | | Cardiometabolic risk factors | Fat-free mass | NR |
| | | Cardiometabolic risk factors | visceral adipose tissue | NR |
| | | Cardiometabolic risk factors | Waist Circunference | NR |
| | Physical Functioning | Cardiometabolic risk factors | VO2max | NR |
| Casas et al., 2018 | Endocrine | Adiposity | Glucose tolerance | NR |
| | General | Body adiposity | Body mass index (BMI) | NR |
| | | Adiposity | Abdominal adiposity | NR |
| | | Adiposity | Body composition | NR |
| | | Adiposity | Body mass index (BMI) | NR |
| | | Adiposity | Fat % | NR |
| | | Adiposity | fat mass | NR |
| | | Adiposity | Fat-free mass | NR |
| | | motor performance | Metabolic parameters | NR |
| | Physical Functioning | Muscle fitness | Medicine ball launch performance | NR |
| | | General motor performance | Vertical jump performance | NR |

| Study (Year) | Domain | Generic outcome (as stated by the authors) | Outcome | Outcome measure |
| --- | --- | --- | --- | --- |
| | | motor performance | strength | NR |
| Cesa et al., 2014 | Blood and Lymphatic System | Cardiometabolic risk factors | Triglycerides | NR |
| | Cardiac | Cardiometabolic risk factors | Diastolic blood pressure | NR |
| | | Cardiometabolic risk factors | Systolic blood pressure | NR |
| | General | Cardiometabolic risk factors | Body mass index (BMI) | NR |
| Chen et al., 2021 | Blood and Lymphatic System | Physical Indexes and Cardiovascular Risk Factors | High density lipoprotein (HDL) | NR |
| | | Physical Indexes and Cardiovascular Risk Factors | Low density lipoprotein (LDL) | NR |
| | | Physical Indexes and Cardiovascular Risk Factors | Total cholesterol | NR |
| | | Physical Indexes and Cardiovascular Risk Factors | Triglycerides | NR |
| | Endocrine | Physical Indexes and Cardiovascular Risk Factors | Insulin | NR |
| | | Physical Indexes and Cardiovascular Risk Factors | Insulin resistance | NR |
| | General | Physical Indexes and Cardiovascular Risk Factors | body fat % | NR |
| | | Physical Indexes and Cardiovascular Risk Factors | Body mass index (BMI) | NR |
| | Physical Functioning | Physical Indexes and Cardiovascular Risk Factors | VO2peak | NR |

| Study (Year) | Domain | Generic outcome (as stated by the authors) | Outcome | Outcome measure |
|---|---|---|---|---|
| Cheng et al., 2022 | Vascular | PWV, IMT and FMD. | Flow-mediated dilation (FMD) | NR |
| | | PWV, IMT and FMD. | Intima-media thickness | NR |
| | | PWV, IMT and FMD. | Pulse wave velocity | NR |
| Choe et al., 2022 | General | Changes in BMI | Body mass index (BMI) | NR |
| | | Changes in BMI | Body weight | NR |
| Clemente et al., 2022 | Blood and Lymphatic System | Health-related outcomes | High density lipoprotein (HDL) | NR |
| | | Health-related outcomes | Number of erythrocytes | NR |
| | | Health-related outcomes | Total cholesterol | NR |
| | | Health-related outcomes | Triglycerides | NR |
| | Cardiac | Health-related outcomes | Mean arterial blood pressure | NR |
| | | Health-related outcomes | Parasympathetic activity | NR |
| | | Health-related outcomes | Resting and maximal heart rate | NR |
| | | Health-related outcomes | Sympathetic activity | NR |
| | | Health-related outcomes | Systolic blood pressure | NR |
| | Endocrine | Health-related outcomes | C-reactive protein | NR |
| | | Health-related outcomes | Metabolic syndrome | NR |
| | General | Body composition | Body fat percentage | NR |
| | | Body composition | Body mass index (BMI) | NR |
| | | Body composition | Height | NR |

| Study (Year) | Domain | Generic outcome (as stated by the authors) | Outcome | Outcome measure |
|---|---|---|---|---|
| | | Body composition | Waist circunference | NR |
| | | Body composition | Weight | NR |
| | Physical Functioning | Physical fitness | Agility | NR |
| | | Physical fitness | Balance | NR |
| | | Physical fitness | Cardiorespiratory fitness | NR |
| | | Physical fitness | Explosive power | NR |
| | | Physical fitness | Flexibility | NR |
| | | Physical fitness | Jump ability | NR |
| | | Physical fitness | Maximal power output (POmax) | NR |
| | | Physical fitness | VO2peak | NR |
| | Vascular | Health-related outcomes | Vascular conductance | NR |
| | | Health-related outcomes | Vascular resistance | NR |
| Collins et al., 2018 | General | Obesity (weight status) | Body fat mass (% and kg) | NR |
| | | Obesity (weight status) | Body fat-free mass (kg) | NR |
| | | Obesity (weight status) | Body mass index (BMI) | BMI (kg/m2); BMI percentile; BMIzscore |
| | | Obesity (weight status) | Body weight | NR |
| | | Obesity (weight status) | Lean body mass | NR |
| | | Obesity (weight status) | Skinfold thickness (mm) | NR |
| | | Obesity (weight status) | Waist circumference (cm) | NR |

| Study (Year) | Domain | Generic outcome (as stated by the authors) | Outcome | Outcome measure |
|---|---|---|---|---|
| Collins et al., 2019 | Emotional Functioning/Wellbeing | "THE SELF (The term ‘the self’ is used in this paper to capture a range of specific terms that, while separate, are related (e.g. self-esteem, self-efficacy, self-perceptions)." | Body attractiveness | Questionnaires |
| | | "THE SELF (The term ‘the self’ is used in this paper to capture a range of specific terms that, while separate, are related (e.g. self-esteem, self-efficacy, self-perceptions)." | Exercise self-efficacy | Questionnaires |
| | | "THE SELF (The term ‘the self’ is used in this paper to capture a range of specific terms that, while separate, are related (e.g. self-esteem, self-efficacy, self-perceptions)." | Global self-steem | Questionnaires |
| | | "THE SELF (The term ‘the self’ is used in this paper to capture a range of specific terms that, while separate, are related | Physical condition | Questionnaires |

| Study (Year) | Domain | Generic outcome (as stated by the authors) | Outcome | Outcome measure |
|---|---|---|---|---|
| | | (e.g. self-esteem, self-efficacy, self-perceptions)." | | |
| | | "THE SELF (The term ‘the self’ is used in this paper to capture a range of specific terms that, while separate, are related (e.g. self-esteem, self-efficacy, self-perceptions)." | Physical self-worth | Questionnaires |
| | | "THE SELF (The term ‘the self’ is used in this paper to capture a range of specific terms that, while separate, are related (e.g. self-esteem, self-efficacy, self-perceptions)." | Physical strength | Questionnaires |
| | | "THE SELF (The term ‘the self’ is used in this paper to capture a range of specific terms that, while separate, are related (e.g. self-esteem, self-efficacy, self-perceptions)." | Resistance training beliefs | Questionnaires |

| Study (Year) | Domain | Generic outcome (as stated by the authors) | Outcome | Outcome measure |
|---|---|---|---|---|
| | | "THE SELF (The term 'the self' is used in this paper to capture a range of specific terms that, while separate, are related (e.g. self-esteem, self-efficacy, self-perceptions)." | Resistance training confidence | Questionnaires |
| | | "THE SELF (The term 'the self' is used in this paper to capture a range of specific terms that, while separate, are related (e.g. self-esteem, self-efficacy, self-perceptions)." | RT self-efficacy | Questionnaires |
| | | "THE SELF (The term 'the self' is used in this paper to capture a range of specific terms that, while separate, are related (e.g. self-esteem, self-efficacy, self-perceptions)." | sport competence | Questionnaires |
| Colquitt et al., 2016 | Adverse Events | | Adverse events | NR |
| | Blood and Lymphatic System | | High density lipoprotein (HDL) | NR |
| | | | Triglycerides | NR |

| Study (Year) | Domain | Generic outcome (as stated by the authors) | Outcome | Outcome measure |
|---|---|---|---|---|
| | Endocrine | | Insulin levels | NR |
| | | | Insulin resistance | NR |
| | General | BMI variable | Body Mass Index (BMI) | NR |
| | | BMI variable | Waist circunference | NR |
| | | BMI variable | Body weight | NR |
| | | BMI variable | Visceral fat | NR |
| | | BMI variable | Abodominal subcutaneous fat | NR |
| | Global Quality of Life | | Health-related QOL | NR |
| | Metabolism and Nutrition | | Sugar intake | NR |
| | | | Fast food intake | NR |
| | Physical Functioning | | Behavior change | NR |
| | Social Functioning | | Parent-child relationship | NR |
| Costa et al., 2020 | Blood and Lymphatic System | TC (total cholesterol) and LDL levels. | Low density lipoprotein (LDL) | NR |
| | | TC (total cholesterol) and LDL levels. | Total cholesterol | NR |
| da Rosa-Santos et al., 2019 | Blood and Lymphatic System | CRF (Cardiometabolic Risk Factors) | High density lipoprotein (HDL) | NR |
| | | CRF (Cardiometabolic Risk Factors) | Low density lipoprotein (LDL) | NR |
| | | CRF (Cardiometabolic Risk Factors) | Triglycerides | NR |

| Study (Year) | Domain | Generic outcome (as stated by the authors) | Outcome | Outcome measure |
|---|---|---|---|---|
| | Cardiac | CRF (Cardiometabolic Risk Factors) | Diastolic blood pressure | NR |
| | | CRF (Cardiometabolic Risk Factors) | Systolic blood pressure | NR |
| | Emotional Functioning/Wellbeing | CRF (Cardiometabolic Risk Factors) | Quality of life | NR |
| | Endocrine | CRF (Cardiometabolic Risk Factors) | Butyrylcholinesterase activity (BChE) | NR |
| | General | CRF (Cardiometabolic Risk Factors) | Adiposity | NR |
| | | CRF (Cardiometabolic Risk Factors) | Body mass | NR |
| | | CRF (Cardiometabolic Risk Factors) | Body mass index (BMI) | NR |
| | | CRF (Cardiometabolic Risk Factors) | Fat-free mass | NR |
| | | CRF (Cardiometabolic Risk Factors) | Waist and hip circumference | NR |
| | | CRF (Cardiometabolic Risk Factors) | Waist circumference | NR |
| | Physical Functioning | CRF (Cardiometabolic Risk Factors) | Body flexibility | NR |
| | | CRF (Cardiometabolic Risk Factors) | Muscle endurance | NR |
| | | CRF (Cardiometabolic Risk Factors) | Physical activity level | NR |

| Study (Year) | Domain | Generic outcome (as stated by the authors) | Outcome | Outcome measure |
|---|---|---|---|---|
| | Vascular | CRF (Cardiometabolic Risk Factors) | Endothelium-dependent vascular dilatation | NR |
| Delgado-Floody et al., 2019 | General | body compositions | Body mass | NR |
| | | body compositions | Body mass index (BMI) | NR |
| | | body compositions | Lean body mass | NR |
| | | body compositions | Skinfold thickness | NR |
| | | body compositions | Sum of skinfolds | NR |
| | | body compositions | trunk fat % | NR |
| | | body compositions | Waist circumference | NR |
| | Physical Functioning | cardiorespiratory fitness | VO2max | NR |
| | | cardiorespiratory fitness | VO2max/CRF | YYIET= Yo-Yo intermittent endurance test |
| Deshira et al., 2022 | Cardiac | Blood pressure | Blood pressure | PACER score |
| | | Blood pressure | Diastolic blood pressure | NR |
| | | Blood pressure | Systolic blood pressure | NR |
| | General | Health outcomes (physical) | Body mass index (BMI) | NR |
| | | Health outcomes (physical) | Waist circumference | NR |
| | | Health outcomes (physical) | Weight redution | NR |
| Dias et al., 2018 | Metabolism and Nutrition | Knowledge and/or behavioral change of diet | Portions of fruit, vegetables and water | Fruit, vegetables and water portions intake by 24- hour dietary recall |
| | Physical Functioning | Knowledge and/or behavioral change of physical activity | Time spent in moderate to vigorous physical activity | Minutes of moderate to vigorous physical exercise by accelerometry (5 consecutive days) |
| Dias et al., 2015 | General | Body composition | Body fat-free mass | NR |

| **Study (Year)** | **Domain** | **Generic outcome (as stated by the authors)** | **Outcome** | **Outcome measure** |
|---|---|---|---|---|
| | | Body composition | Body weight | NR |
| | Physical Functioning | Cardiorespiratory Fitness | Peak oxygen consumption (VO2 peak) | Measured by VO2 peak |
| | Vascular | Vascular function | Flow-mediated dilation (FMD) | Measured by FMD peak (%) or FMD area under the curve (AUC) (%s) or FMD with glyceryl trinitrate (GTN) (%) or Resting brachial artery (BA) diameter |
| Dias et al., 2021 | Cardiac | Heart Rate Variability | HF | NR |
| | | Heart Rate Variability | LF | NR |
| | | Heart Rate Variability | LF/HF | NR |
| | | Heart Rate Variability | Mean RR | NR |
| | | Heart Rate Variability | RMSSD | NR |
| | | Heart Rate Variability | SD1 | NR |
| | | Heart Rate Variability | SD2 | NR |
| | | Heart Rate Variability | SDNN | NR |
| | General | Anthropometric | Body Fat | NR |
| | | Anthropometric | Body mass index (BMI) | NR |
| Dobbins et al., 2013 | Blood and Lymphatic System | Lipid profile | Total cholesterol | Blood samples were taken for some studies after fasting, and for others with no fasting. |
| | Cardiac | Blood pressure | Mean systolic and diastolic blood pressure | Measured either manually using a mercury sphygmomanometer or via a Dinamap machine. The timing of when blood pressure was measured differed significantly across studies with timing varying between 5 and 15 min following active periods during school hours. |

| Study (Year) | Domain | Generic outcome (as stated by the authors) | Outcome | Outcome measure |
|---|---|---|---|---|
| | | Pulse rate | Resting heart rate (beats/minute) | This outcome was measured by trained professionals during school time, during seated rest. |
| | | Blood pressure | Systolic blood pressure | Measured either manually using a mercury sphygmomanometer or via a Dinamap machine. The timing of when blood pressure was measured differed significantly across studies with timing varying between 5 and 15 min following active periods during school hours. |
| | General | BMI | Body mass index (BMI) | ($kg/m^2$)This outcome was measured by trained health professionals using calibrated scales. |
| | Physical Functioning | Maximal oxygen consumption | Maximal oxygen consumption (VO2max) | This outcome was measured in different ways by trained professionals. In some instances pulse rate recovery was used as a proxy for VO2max and in other instances actual maximal oxygen uptake was measured. |
| | | Rate of moderate to vigorous physical activity (MVPA) (per cent of sample engaged in MVPA) | Rates of moderate to vigorous physical activity during the school day | Rate assessed either through self-report or through the use of accelerometers (or both). The rate was calculated by dividing the number of students engaged in MVPA by the total number of students allocated to either the intervention or control group |
| | | Duration of physical activity (time spent engaged in MVPA) | Time spent in moderate to vigorous physical activity | Time was measured as the total minutes per hour or week spent engaged in MVPA generally through self-report, although some studies collected these data via accelerometers |
| | | Television viewing (time spent watching TV) | Time spent watching television | Measured by self-report or parental report as the minutes per hour or week spent watching television |
| Eckstein et al., 2022 | Blood and Lymphatic System | High-density lipoprotein cholesterol | High density lipoprotein (HDL) | NR |
| | | Low-density lipoprotein cholesterol | Low density lipoprotein (LDL) | NR |

| Study (Year) | Domain | Generic outcome (as stated by the authors) | Outcome | Outcome measure |
|---|---|---|---|---|
| | | Total cholesterol | Total cholesterol | NR |
| | | Triglycerides | Triglycerides | NR |
| | Cardiac | Cardiovascular risk factors | Diastolic blood pressure | NR |
| | | Cardiac autonomic function | Heart rate variability | NR |
| | | Cardiovascular risk factors | Mean systolic and dastolic blood pressure | NR |
| | | Cardiovascular risk factors | Systolic blood pressure | NR |
| | Endocrine | Myokines | Brain-derived neurotrophic factor (BDNF) | ng/mL |
| | | Insuline values | C-reactive protein | mg/L |
| | | Blood glucose | Fasting glucose | NR |
| | | Blood glucose | HbA1c | NR |
| | | Insuline values | Insulin | NR |
| | | Myokines | Irisin | ng/mL |
| | | Myokines | Myostatin | NR |
| | | Serum oxidative stress biomarkers | Plasma malondialdehyde | mmol/mL |
| | | Myokines | Serum interleukin-15 | NR |
| | | Serum oxidative stress biomarkers | Superoxide dismutase | U/mL |
| | General | Body composition | Body fat mass | NR |
| | | Body composition | Body fat mass (%) | NR |
| | | Body composition | Body fat-free mass | NR |

| Study (Year) | Domain | Generic outcome (as stated by the authors) | Outcome | Outcome measure |
|---|---|---|---|---|
| | | Anthropometric parameters | Body height | NR |
| | | Anthropometric parameters | Body mass | kg |
| | | Anthropometric parameters | Body mass index (BMI) | NR |
| | | Anthropometric parameters | Body weight | NR |
| | | Lean body mass | Lean body mass | NR |
| | | Regional fat | Regional fat | NR |
| | | Body composition | Trunk fat | NR |
| | | Anthropometric parameters | Waist circunference | NR |
| | Musculoskeletal and Connective Tissue | Body composition | Regional bone density | NR |
| | | Body composition | Total bone density | NR |
| | Physical Functioning | Physical fitness variables | Balance | NR |
| | | Physical fitness variables | Cardiorespiratory endurance | NR |
| | | Physical fitness variables | Cardiorespiratory fitness | NR |
| | | Physical fitness variables | Flexibility | Sit and reach |
| | | Cardiorrespiratory fitness | Gas exchange threshold | NR |
| | | Physical fitness variables | Maximal muscle strength | NR |
| | | Cardiorrespiratory fitness | Maximum speed | km/h |
| | | Physical fitness variables | Muscle endurance | reps |
| | | Physical fitness variables | Muscle strength | NR |
| | | Physical fitness variables | Peak muscle power | Newton (N) |

| Study (Year) | Domain | Generic outcome (as stated by the authors) | Outcome | Outcome measure |
|---|---|---|---|---|
| | | Cardiorrespiratory fitness | Peak oxygen consumption (VO2 peak) | NR |
| | | Physical fitness variables | Power | Surgent vertical jump |
| | Vascular | Cardiovascular risk factors | Arterial stiffness | Right brachial-ankle pulse wave velocity (m/s-1) |
| Elber et al., 2019 | Blood and Lymphatic System | cardiovascular disease | Cholesterol | NR |
| | Cardiac | cardiovascular disease | Systolic blood pressure | NR |
| | General | obesity | body fat mass | NR |
| | | obesity | Body mass index (BMI) | NR |
| | Physical Functioning | cardiovascular disease | VO2 max | NR |
| Estéves-González et al., 2022 | Cardiac | Heart Rate Variability | HF | NR |
| | | Heart Rate Variability | LF | NR |
| | | Heart Rate Variability | LF/HF | NR |
| | | Heart Rate Variability | Mean RR | NR |
| | | Heart Rate Variability | PNN50 | NR |
| | | Heart Rate Variability | RMSSD | NR |
| | | Heart Rate Variability | SDNN | NR |
| Farah et al., 2012 | Cardiac | Blood pressure change | Diastolic blood pressure | NR |
| | | Blood pressure change | Mean systolic and diastolic blood pressure | NR |
| | | Blood pressure change | Systolic blood pressure | NR |
| | General | Body mass | Body weight | NR |

| Study (Year) | Domain | Generic outcome (as stated by the authors) | Outcome | Outcome measure |
|---|---|---|---|---|
| Feng et al., 2017 | General | Anthropometric | Body mass index (BMI) | NR |
| Frerichs et al., 2016 | General | Weight-related outcomes (obesity) | Body mass index (BMI) | BMI (kg/m2) or BMI z-scores (objective measurement of weight and height via calibrated scale and stadiometer) |
| | | Weight-related outcomes (obesity) | Body weight | Objective measurement of weight via calibrated scale |
| | | Weight-related outcomes (obesity) | Waist circumference | NR |
| | Metabolism and Nutrition | Diet behavior (obesity-related lifestyle behaviours) | Self-report of food servings | Self-reported questionnaire measures: fruit Servings, vegetable servings, sugar sweetened beverage servings, number of snacks/desserts per day |
| | Physical Functioning | Physical activity behavior (obesity-related lifestyle behaviours) | Time spent in moderate to vigorous physical activity | Measured by accelerometer or questionnaire |
| García-Hermoso et al., 2014 | Endocrine | Insulin resistance markers | Fasting glucose | Fasting glucose concentration was determined by enzymatic methods or using a glucose oxidase technique by using a radioimmunoassay (RIA) kit |
| | | Insulin resistance markers | Fasting insulin | Fasting insulin levels were determined using a chemiluminescent assay, RIA, or by an Immulite analyzer |
| | General | Percentage of body fat | Body fat mass (%) | NR |
| García-Hermoso et al., 2015 | General | Body composition | Body fat mass (kg and %) | NR |
| | | Body composition | Body fat-free mass | NR |
| | | Body composition | Body mass index (BMI) | NR |
| | | Body composition | Body weight | NR |
| | | Body composition | Waist circumference | NR |

| Study (Year) | Domain | Generic outcome (as stated by the authors) | Outcome | Outcome measure |
|---|---|---|---|---|
| García-Hermoso et al., 2016-a | Blood and Lymphatic System | Cardiometabolic risk factor (Lipids and lipoprotein) | High density lipoprotein (HDL) | The evaluations were made in the morning after 12 h of overnight fasting |
| | | Cardiometabolic risk factor (Lipids and lipoprotein) | Low density lipoprotein (LDL) | The evaluations were made in the morning after 12 h of overnight fasting |
| | | Cardiometabolic risk factor (Lipids and lipoprotein) | Total cholesterol | The evaluations were made in the morning after 12 h of overnight fasting |
| | | Cardiometabolic risk factor (Lipids and lipoprotein) | Triglycerides | The evaluations were made in the morning after 12 h of overnight fasting |
| | Cardiac | Cardiometabolic risk factor (blood pressure) | Diastolic blood pressure | Various methods were used to determine blood pressure |
| | | Cardiometabolic risk factor (blood pressure) | Systolic blood pressure | Various methods were used to determine blood pressure |
| | Endocrine | Cardiometabolic risk factor (Insulin resistance) | Fasting glucose | The evaluations were made in the morning after 12 h of overnight fasting |
| | | Cardiometabolic risk factor (Insulin resistance) | Fasting insulin | The evaluations were made in the morning after 12 h of overnight fasting |
| | | Cardiometabolic risk factor (Insulin resistance) | HOMA-index (Homeostatic Model Assessment for Insulin Resistance) | The evaluations were made in the morning after 12 h of overnight fasting |
| | General | Body composition | Body fat mass (% or kg) | Measured by bioelectrical impedance or dual-energy x-ray absorptiometry |
| | | Body composition | Body mass index (BMI) | NR |
| | | Body composition | Body weight | Measure was assessed using a medical scale |
| | | Body composition | Waist circumference | Measured using the standard protocol |

| **Study (Year)** | **Domain** | **Generic outcome (as stated by the authors)** | **Outcome** | **Outcome measure** |
|---|---|---|---|---|
| García-Hermoso et al., 2016-b | Blood and Lymphatic System | Lipids and lipoprotein | High density lipoprotein (HDL) | NR |
| | | Lipids and lipoprotein | Low density lipoprotein (LDL) | NR |
| | | Lipids and lipoprotein | Total cholesterol | NR |
| | | Lipids and lipoprotein | Triglycerides | NR |
| | Cardiac | Blood pressure | Systolic blood pressure | NR |
| | Endocrine | Adipokines | Adiponectin | NR |
| | | Insulin resistance | Fasting glucose | NR |
| | | Insulin resistance | Fasting insulin | NR |
| | | Insulin resistance | HOMA-index (Homeostatic Model Assessment for Insulin Resistance) | NR |
| | | Adipokines | Leptin | NR |
| | General | Body composition | Body fat mass (kg and %) | NR |
| | | Body composition | Body mass index (BMI) | Body mass index (kg/m2) |
| | | Body composition | Body weight | NR |
| | | Body composition | Lean body mass | NR |
| | | Body composition | Subcutaneous adipose tissue (cm) | NR |
| | | Body composition | Visceral adipose tissue (cm) | NR |

| Study (Year) | Domain | Generic outcome (as stated by the authors) | Outcome | Outcome measure |
|---|---|---|---|---|
| | | Body composition | Waist circumference (cm) | NR |
| García-Hermoso et al (2016-c) | Blood and Lymphatic System | Level of serum C-reactive protein | Serum C-reactive protein | All the techniques employed to determine CRP used ultrasensitive methods: nephelometry, latex-enhanced immunoturbidimetric assay, immunoradiometry assay, and highly sensitive enzyme-linked immunosorbent assay. |
| | Endocrine | Fasting insulin | Fasting insulin | NR |
| | General | Body composition (weight and body fat) | Body fat mass (%) | NR |
| | | Body composition (weight and body fat) | Body weight | NR |
| García-Hermoso et al., 2017-a | Endocrine | Adipokines | Adiponectin | NR |
| | | Adipokines | Leptin | NR |
| | | Adipokines | Resistin levels | NR |
| | | Adipokines | Visfatin levels | NR |
| García-Hermoso et al., 2017-b | Vascular | Carotid Arterial Wall Thickness | Intima-media thickness (IMT) | Measured the carotid IMT by high-resolution B mode ultrasound at the carotid artery |
| Garcia-Hermoso et al., 2020-a | General | Adiposity | Body composition | DXA, MRI, bioeletrical impedance analysis |
| | | Adiposity | skinfold tickness | NR |
| | Physical Functioning | Cardiorespiratory Fitness | Cardiorespiratory fitness | maximal or submaximal tests on a cycle ergometer or treadmill with spirometry |
| Garcia-Hermoso et al., 2020-b | General | Physical Fitness | Body Fat | NR |
| | | Physical Fitness | Body mass index (BMI) | calculated as weight in kilograms divided by height in meters squared |
| | | Physical Fitness | skinfold tickness | NR |

| Study (Year) | Domain | Generic outcome (as stated by the authors) | Outcome | Outcome measure |
|---|---|---|---|---|
| | | Physical Fitness | Waist Circunference | NR |
| | Physical Functioning | Physical Fitness | Cardiorespiratory fitness | NR |
| | | Fundamental Motor Skills | Fundamental motor skills | locomotor and object control skills |
| García-Hermoso et al., 2019 | Cardiac | Blood Pressure | Diastolic blood pressure | NR |
| | | Blood Pressure | Systolic blood pressure | NR |
| | General | Anthropometric | Body fat | NR |
| | | Anthropometric | Body mass index (BMI) | NR |
| | | Anthropometric | Obesity prevalence | NR |
| | | Anthropometric | Skinfold thickness | NR |
| | | Anthropometric | Waist Circumference | NR |
| | | Anthropometric | zBMI | NR |
| | Physical Functioning | Cardiorespiratory Fitness | Cardiorespiratory fitness | shuttle run test; 10x20m test |
| | | Lower-body muscular strength | Lower-body muscular strength | squat jump |
| | | Speed-agility | Speed-agility | 10mx4 shuttle run test; obstacle course test; 20m test; 6m test |
| González-Ruiz et al., 2017 | Endocrine | Liver enzymes | Alanine aminotransferase (ALT) | Measured using commercially available test kits |
| | | Liver enzymes | Aspartate aminotransferase (AST) | Measured using commercially available test kits |
| | | Liver enzymes | Gamma-glutamyl transferase (GGT) | Measured using commercially available test kits |

| Study (Year) | Domain | Generic outcome (as stated by the authors) | Outcome | Outcome measure |
|---|---|---|---|---|
| | | Intrahepatic fat | Intrahepatic fat | Measured with proton magnetic resonance spectra acquired in the same proton magnetic resonance spectroscopic system with a body matrix coil and a spine matrix |
| | General | Abdominal fat | Subcutaneous adipose tissue | Measured with magnetic resonance imaging or by ultrasound examination |
| | | Abdominal fat | Visceral adipose tissue | Measured with magnetic resonance imaging or by ultrasound examination |
| Gonzalez-Suarez et al., 2009 | General | Management of childhood obesity | Body fat mass (%) | Measured by bioelectric impedance, dual energy absorptiometry, or by the sum of skinfolds |
| | | Management of childhood obesity | Body mass index (BMI) | NR |
| | | Management of childhood obesity | Skinfold thickness | Triceps and subescapular skinfold thickness (mm) |
| | | Management of childhood obesity | Waist circumference | NR |
| | | Management of childhood obesity | Waist-to-hip ratio | NR |
| Guerra et al., 2013 | General | Body mass index (BMI) | Body mass index (BMI) | Kg/m² |
| Guerra et al., 2014 | Cardiac | Blood pressure | Diastolic blood pressure | NR |
| | | Blood pressure | Systolic blood pressure | NR |
| | General | Body mass index (BMI) | Body mass index (BMI) | NR |
| | | Body weight | Body weight | NR |
| Hale et al., 2023 | Emotional Functioning/Wellbeing | Psychological well-being and ill-being of children | body image (steem, dissatisfaction) | Questionnaires and surveys, buit it is not described, only the acronyms. |

| Study (Year) | Domain | Generic outcome (as stated by the authors) | Outcome | Outcome measure |
|---|---|---|---|---|
| | | Psychological well-being and ill-being of children | Quality of life | Questionnaires and surveys, buit it is not described, only the acronyms. |
| | | Psychological well-being and ill-being of children | self-perception | Questionnaires and surveys, buit it is not described, only the acronyms. |
| | | Psychological well-being and ill-being of children | self-steem | Questionnaires and surveys, buit it is not described, only the acronyms. |
| | Psychiatric | Psychological well-being and ill-being of children | Anxiety | Questionnaires and surveys, buit it is not described, only the acronyms. |
| | | Psychological well-being and ill-being of children | Depression | Questionnaires and surveys, buit it is not described, only the acronyms. |
| | | Psychological well-being and ill-being of children | Poor well-being | Questionnaires and surveys, buit it is not described, only the acronyms. |
| | | Psychological well-being and ill-being of children | Psychological ill-being | Questionnaires and surveys, buit it is not described, only the acronyms. |
| | | Psychological well-being and ill-being of children | Stress | Questionnaires and surveys, buit it is not described, only the acronyms. |
| Hamel et al., 2011 | General | Health outcomes | Body fat mass (%) | % measured by DEXA |
| | | Health outcomes | Body mass index (BMI) | NR |
| | | Health outcomes | Body weight | NR |
| | | Health outcomes | Waist circumference | NR |
| | Physical Functioning | Physical activity | Days of moderate-vigorous physical activity per week | self report |

| Study (Year) | Domain | Generic outcome (as stated by the authors) | Outcome | Outcome measure |
|---|---|---|---|---|
| | | Physical activity | Time spent in moderate to vigorous physical activity | Minutes of physical activity moderate-vigorous per week self-report or measured by accelerometry |
| Han et al., 2019 | Blood and Lymphatic System | | High density lipoprotein (HDL) | NR |
| | | | Low density lipoprotein (LDL) | NR |
| | | | Total cholesterol | NR |
| | | | Triglycerides | NR |
| | Cardiac | | Systolic blood pressure | NR |
| | | | Diastolic blood pressure | NR |
| | Endocrine | | Adiponectin | NR |
| | | | C-reactive protein | NR |
| | | | Fasting glucose | NR |
| | | | IL-6 | NR |
| | | | serum insulin | NR |
| | | | TNF-α | NR |
| | General | | Body mass index (BMI) | NR |
| | | | Fat % | NR |
| | | | Fat-free mass | NR |
| | | | Waist Circunference | NR |
| | | | weight | NR |
| Harris et al., 2009 | General | Body composition | Body fat mass (% or kg) | NR |

| Study (Year) | Domain | Generic outcome (as stated by the authors) | Outcome | Outcome measure |
|---|---|---|---|---|
| | | Body composition | Body fat mass (%) | NR |
| | | Body composition | Body mass index (BMI) | NR |
| | | Body composition | Lean body mass | NR |
| | | Body composition | Skinfold thickness | Triceps, subescapular and sum (mm) |
| | | Body composition | Waist circumference | NR |
| | | Body composition | Waist-to-hip ratio | NR |
| Hejazi et al., 2022 | Blood and Lymphatic System | HDL-C | High density lipoprotein (HDL) | NR |
| | | LDL-C | Low density lipoprotein (LDL) | NR |
| | | Triglycerides | Triglycerides | NR |
| | Endocrine | Adiponectin | Adiponectin | Adiponectin levels |
| | | C-Reactive Protein | C-reactive protein | NR |
| | | glucose metabolism | Fasting glucose | NR |
| | | glucose metabolism | HOMA-index (Homeostatic Model Assessment for Insulin Resistance) | NR |
| | | IL-6 | IL-6 | NR |
| | | glucose metabolism | Insulin | NR |
| | | Leptin | Leptin | NR |
| | Physical Functioning | VO2 max | VO2 max | NR |

| Study (Year) | Domain | Generic outcome (as stated by the authors) | Outcome | Outcome measure |
|---|---|---|---|---|
| Hernandez-Martin et al., 2021 | General | body composition | Body mass index (BMI) | Stadiometer,<br>Bioimpedance, DXA, Scale,<br>stadiometer, Skinfold calliper, Skinfold calliper |
| | | body composition | Fat mass | Stadiometer,<br>Bioimpedance, DXA, Scale,<br>stadiometer, Skinfold calliper, Skinfold calliper |
| | | body composition | Lean body mass | Stadiometer,<br>Bioimpedance, DXA, Scale,<br>stadiometer, Skinfold calliper, Skinfold calliper |
| | Musculoskeletal and Connective Tissue | body composition | Bone mineral content | Stadiometer,<br>Bioimpedance, DXA, Scale,<br>stadiometer, Skinfold calliper, Skinfold calliper |
| Ho et al., 2013 | Blood and Lymphatic System | Metabolic | High density lipoprotein (HDL) | NR |
| | | Metabolic | Low density lipoprotein (LDL) | NR |
| | | Metabolic | Triglycerides | NR |
| | Endocrine | Metabolic | Fasting glucose | NR |
| | | Metabolic | Fasting insulin | NR |
| | General | Weight change | Body mass index (BMI) | NR |
| | | Weight change | Body fat mass (%) | NR |
| | | Weight change | Lean body mass | NR |

| Study (Year) | Domain | Generic outcome (as stated by the authors) | Outcome | Outcome measure |
|---|---|---|---|---|
| Jago et al., 2004 | Physical Functioning | Physical activity | Time spent in moderate to vigorous physical activity | Self-reported sweating; minutes of moderate to vigorous intensity physical activity from heart rate monitoring (50%/60% heart rate reserve); total self-resported physical activity (questionnaire); travel to school questionnaire to record self report mode of transportation; SOFIT to assess minutes of moderate intensity physical activity in PE lesson; SOPLAY to asses minutes of moderate intensity physical activity out of PE lesson. |
| Jull and Chen., 2013 | General | Weight loss | Body mass index (BMI) | BMI-SDS or z-BMI |
| | | Weight loss | Percentage overweight | [percent overweight = (effective BMI/BMI 50th percentile) –1] |
| Jung et al., 2018 | General | Visceral Fat | Visceral Fat | Computerized Tomography<br>Magnetic Resonance Imaging (MRI)<br>Dual energy x-ray absorptiometry (DEXA)<br>Ultrasound |
| Jurado-Castro et al., 2020 | Physical Functioning | Time spent engaged in MVPA | Minutes of MVPA | differents protocols for acceletometer use. |
| | General | BMI | zBMI | NR |
| Kelley and Kelley., 2008 | Blood and Lymphatic System | High-density lipoprotein cholesterol (HDL-C) | High density lipoprotein (HDL) | The assessment of lipids and lipoproteins took place in the morning after |
| | | Non–high-density lipoprotein cholesterol (non–HDL-C) | Non-High-density lipoproteins (non–HDL-C) | calculated as total cholesterol (TC) minus HDL-C. The assessment of lipids and lipoproteins took place in the morning after |
| | | Total cholesterol (TC) | Total cholesterol | The assessment of lipids and lipoproteins took place in the morning after |
| | General | Percent body fat | Body fat mass (%) | NR |
| | | Body weight | Body weight | NR |

| Study (Year) | Domain | Generic outcome (as stated by the authors) | Outcome | Outcome measure |
|---|---|---|---|---|
| | Physical Functioning | cardiorespiratory capacity | Maximal oxygen consumption (VO2max) | (mL/kg/min) |
| Kelley et al., 2019 | Blood and Lymphatic System | | Fasting glucose | NR |
| | | | HbA1c | NR |
| | | | High density lipoprotein (HDL) | NR |
| | | | Low density lipoprotein (LDL) | NR |
| | | | Non-fasting glucose | NR |
| | | | Non-High-density lipoproteins (non–HDL-C) | NR |
| | | | Total cholesterol | NR |
| | | | Total cholesterol to HDL ratio | NR |
| | | | Triglycerides | NR |
| | Cardiac | | Diastolic blood pressure | NR |
| | | | Systolic blood pressure | NR |
| | Endocrine | | Fasting insulin | NR |
| | | | Non-fasting insulin | NR |
| | General | Adiposity | Body fat % | NR |
| | | Adiposity | Body mass index (BMI) | NR |
| | | | body weight | NR |
| | | Adiposity | Fat mass | NR |

| Study (Year) | Domain | Generic outcome (as stated by the authors) | Outcome | Outcome measure |
|---|---|---|---|---|
| | | | lean body mass | NR |
| | | | Waist circumference | NR |
| | | | waist/hip ratio | NR |
| | Metabolism and Nutrition | | Dietary intake | NR |
| | Physical Functioning | | Maximal strength | NR |
| | | | VO2max | NR |
| | | | Energy expenditure | NR |
| | | | Physical activity level | NR |
| Kelley et al., 2021 | Blood and Lymphatic System | Lipid and lipoprotein outcomes | High density lipoprotein (HDL) | NR (assessed with blood draw) |
| | | Lipid and lipoprotein outcomes | Low density lipoprotein (LDL) | NR (assessed with blood draw) |
| | | Lipid and lipoprotein outcomes | Total cholesterol | NR (assessed with blood draw) |
| | | Lipid and lipoprotein outcomes | Triglycerides | NR (assessed with blood draw) |
| | Cardiac | Blood pressure outcomes | Resting diastolic blood pressure (DBP) | Various types of sphygmomanometers |
| | | Blood pressure outcomes | Resting systolic blood pressure (SBP) | Various types of sphygmomanometers |
| | Endocrine | Diabetes-related outcomes | Fasting glucose | NR (assessed with blood draw) |
| | | Diabetes-related outcomes | Fasting insulin | NR (assessed with blood draw) |
| | General | Adiposity outcomes | Body fat mass | Various methods |
| | | Adiposity outcomes | Body fat mass (%) | Various methods |

| Study (Year) | Domain | Generic outcome (as stated by the authors) | Outcome | Outcome measure |
|---|---|---|---|---|
| | | Fat-free mass | Body fat-free mass | Various methods |
| | | Adiposity outcomes | Body mass index (BMI) | Measured from height and body weight |
| | | Body weight | Body weight | Various types of scales |
| | | Adiposity outcomes | Waist circumference | Tape measure |
| | Physical Functioning | Maximum oxygen consumption | Maximal oxygen consumption (VO2max) | Various methods |
| Kemp et al., 2021 | General | Cardiovascular Disease Risk | Alcohol and cigarette use | NR |
| | | Cardiovascular Disease Risk | Body mass index (BMI) | NR |
| | | Cardiovascular Disease Risk | Parents' alcohol use | NR |
| | | Cardiovascular Disease Risk | Parents' BMI | NR |
| | Metabolism and Nutrition | Cardiovascular Disease Risk | Fruit and vegetable intake | NR |
| | Physical Functioning | Cardiovascular Disease Risk | Physical activity level | NR |
| | Psychiatric | Cardiovascular Disease Risk | Depression | NR |
| Kim et al., 2017 | General | Body weight | Body weight | NR |
| Kropski et al., 2008 | General | Obesity | Body fat mass (%) | NR |
| | | Obesity | Body mass index (BMI) | NR |
| | | Obesity | Obesity/overweight prevalence | NR |
| | | Obesity | Skinfold thickness | Triceps skinfold thickness (mm) |
| Laframboise et al., 2011 | Endocrine | Adiponectin levels | Adiponectin | NR |
| | General | Adiposity | Body fat mass (%) | NR |

| **Study (Year)** | **Domain** | **Generic outcome (as stated by the authors)** | **Outcome** | **Outcome measure** |
|---|---|---|---|---|
| | | Adiposity | Body mass index (BMI) | BMI SD-score |
| | | Adiposity | Body weight | NR |
| | | Adiposity | Skinfold thickness | NR |
| | | Adiposity | Trunk and visceral fat composition | NR |
| | | Adiposity | Waist circumference | NR |
| | | Adiposity | Waist-to-hip ratio | NR |
| Lee et al., 2020 | Blood and Lymphatic System | Cardiometabolic factors | High density lipoprotein (HDL) | NR |
| | | Cardiometabolic factors | Low density lipoprotein (LDL) | NR |
| | | Cardiometabolic factors | Total cholesterol | NR |
| | | Cardiometabolic factors | Triglycerides | NR |
| | Cardiac | Blood Pressure | Diastolic blood pressure | NR |
| | | Blood Pressure | Systolic blood pressure | NR |
| | Endocrine | Inflammatory Markers | C-reactive protein | NR |
| | | Inflammatory Markers | Fasting glucose | NR |
| | | Inflammatory Markers | IL-6 | NR |
| | | Inflammatory Markers | Insulin | NR |
| | | Inflammatory Markers | Insulin resistance | HOMA-IR |
| | | Inflammatory Markers | TNF-alfa | NR |
| | General | Body fat | Body mass index (BMI) | NR |
| | | Body fat | Fat-free mass | NR |

| Study (Year) | Domain | Generic outcome (as stated by the authors) | Outcome | Outcome measure |
|---|---|---|---|---|
| | | Body fat | Waist Circumference | NR |
| | Physical Functioning | Physical Fitness | VO2peak | NR |
| Li and Chen et al., 2021 | Blood and Lymphatic System | CRF (Cardiometabolic Risk Factors) | High density lipoprotein (HDL) | NR |
| | | CRF (Cardiometabolic Risk Factors) | Low density lipoprotein (LDL) | NR |
| | | CRF (Cardiometabolic Risk Factors) | Total Cholesterol (TC) | NR |
| | | CRF (Cardiometabolic Risk Factors) | Triglyceride (TG) | NR |
| | Endocrine | CRF (Cardiometabolic Risk Factors) | HOMA-index (Homeostatic Model Assessment for Insulin Resistance) | NR |
| | General | CRF (Cardiometabolic Risk Factors) | Body mass index (BMI) | NR |
| | Physical Functioning | CRF (Cardiometabolic Risk Factors) | Peak oxygen consumption (VO2 peak) | NR |
| Li et al., 2022 | Blood and Lymphatic System | Changes in BMI | High density lipoprotein (HDL) | NR |
| | | Changes in BMI | Low density lipoprotein (LDL) | NR |
| | | Changes in BMI | Total cholesterol | NR |
| | | Changes in BMI | Triglycerides | NR |
| | Cardiac | Changes in BMI | Blood Pressure | NR |

| Study (Year) | Domain | Generic outcome (as stated by the authors) | Outcome | Outcome measure |
|---|---|---|---|---|
| | Endocrine | Changes in BMI | Fasting glucose | NR |
| | General | Changes in BMI | Body mass index (BMI) | NR |
| Littlewood et al., 2020 | Cardiac | Cardiometabolic factors | Diastolic blood pressure | NR |
| | | Cardiometabolic factors | Systolic blood pressure | NR |
| | Emotional Functioning/Wellbeing | Behavioural | Screen time | NR |
| | Endocrine | Cardiometabolic factors | Fasting insulin | NR |
| | | Cardiometabolic factors | HbA1c | NR |
| | | Cardiometabolic factors | Insulin resistance | HOMA-IR |
| | General | Anthropometric | Body fat % | NR |
| | | Anthropometric | Body mass index (BMI) | NR |
| | | Anthropometric | Fat mass | NR |
| | | Anthropometric | Waist circumference | NR |
| | Physical Functioning | Cardiometabolic factors | Physical activity levels | steps/day; IPAQ; accelerometer |
| | | Cardiometabolic factors | VO2max | NR |
| | Psychiatric | Psychological | HR-QOL | NR |
| | | Psychological | overall psychological profile | NR |
| Liu et al., 2020 | Physical Functioning | Fundamental Motor Skills | Fundamental motor skills | Test of Gross Motor Development, balance (test children's balance); agility (reaction times); physical perception (Youth Physical Self Youth Physical Self and Pictorial Scale of Perceived Competence) |
| | | Physical Fitness | Physical fitness | NR |

| Study (Year) | Domain | Generic outcome (as stated by the authors) | Outcome | Outcome measure |
|---|---|---|---|---|
| Lopes et al., 2019 | Blood and Lymphatic System | blood concentration of HDL-C | High density lipoprotein (HDL) | NR |
| Martin-Smith et al., 2020 | General | Body Composition | Body composition | |
| | Physical Functioning | Cardiorespiratory Fitness | VO2max | Direct and indirect methods |
| Mclean et al., 2003 | General | Weight management | Body mass index (BMI) | NR |
| | | Weight management | Body weight | NR |
| Mead et al., 2017 | Adverse Events | | Morbidity | NR |
| | | | Adverse events | NR |
| | Blood and Lymphatic System | | High density lipoprotein (HDL) | NR |
| | | | Low density lipoprotein (LDL) | NR |
| | | | Total cholesterol | NR |
| | | | Triglycerides | NR |
| | Cardiac | | Systolic blood pressure | NR |
| | | | Diastolic blood pressure | NR |
| | | | Heart rate change | NR |
| | Delivery of Care | | Quality of care | NR |
| | Emotional Functioning/Wellbeing | | Self-perception | NR |
| | | | Self-steem | NR |
| | | | Self-worth (self-perception) | NR |
| | | | Mood | NR |

| Study (Year) | Domain | Generic outcome (as stated by the authors) | Outcome | Outcome measure |
|---|---|---|---|---|
| | | | Physical activity enjoyment | NR |
| | Endocrine | | Fasting insulin | NR |
| | | | Insulin | NR |
| | | | Adiponectin | NR |
| | | | Fasting glucose | OGTT |
| | | | Glucose | OGTT |
| | General | Changes in BMI | Body Mass Index (BMI) | DXA and pedometers |
| | | Changes in BMI | Body fat | DXA and pedometers |
| | | Changes in BMI | Body weight | DXA and pedometers |
| | | Changes in BMI | Waist circumference | DXA and pedometers |
| | | Changes in BMI | Overweight prevalence | DXA and pedometers |
| | | Changes in BMI | Fat-free mass | DXA and pedometers |
| | | Changes in BMI | Parents outcomes | DXA and pedometers |
| | | Changes in BMI | Skinfold thickness | NR |
| | Global Quality of Life | | Health-related QoL | NR |
| | Metabolism and Nutrition | | Dietary behavior | NR |
| | Physical Functioning | | Behavior change | NR |
| | | | Screen time | NR |
| | | | Physical activity level | NR |
| | Social Functioning | | Parental acceptance | NR |

| Study (Year) | Domain | Generic outcome (as stated by the authors) | Outcome | Outcome measure |
|---|---|---|---|---|
| Mears and Jago., 2016 | Physical Functioning | Moderate-to-vigorous physical activity | Time spent in moderate to vigorous physical activity | Measured by accelerometers, heart rate (HR) monitor, self-report |
| Medina-Blanco et al., 2011 | Physical Functioning | Physical activity | Time spent in physical activity | Measure by accelerometer and pedometer (minutes, days and minutes per day) |
| Mei et al., 2016 | General | BMI | Body mass index (BMI) | $kg/m^2$ |
| Méndez-Hernández et al. (2022) | Blood and Lymphatic System | Blood markers | Glucose | NR |
| | | Blood markers | Total cholesterol | NR |
| | | Blood markers | Triglycerides | NR |
| | Cardiac | Cardiometabolic Factors | Diastolic blood pressure | NR |
| | | Cardiometabolic Factors | Systolic blood pressure | NR |
| | Endocrine | Blood markers | Adiponectin | NR |
| | | Blood markers | Insulin | NR |
| | | Blood markers | Insulin resistance | HOMA-IR |
| | | Blood markers | Leptin | NR |
| | General | Anthropometric characteristcs | Body fat % | NR |
| | | Anthropometric characteristcs | Body mass index (BMI) | NR |
| | | Anthropometric characteristcs | Fat-free mass | NR |
| | | Anthropometric characteristcs | Waist circumference | NR |

| Study (Year) | Domain | Generic outcome (as stated by the authors) | Outcome | Outcome measure |
|---|---|---|---|---|
| Menjie et al., 2022 | Blood and Lymphatic System | Outcome indexes: body shape indicators, CRF indicators and cardiovascular disease metabolic risk | High density lipoprotein (HDL) | NR |
| | | Outcome indexes: body shape indicators, CRF indicators and cardiovascular disease metabolic risk | Low density lipoprotein (LDL) | NR |
| | | Outcome indexes: body shape indicators, CRF indicators and cardiovascular disease metabolic risk | Total cholesterol | NR |
| | | Outcome indexes: body shape indicators, CRF indicators and cardiovascular disease metabolic risk | Triglycerides | NR |
| | Cardiac | Outcome indexes: body shape indicators, CRF indicators and cardiovascular disease metabolic risk | Diastolic blood pressure | NR |
| | | Outcome indexes: body shape indicators, CRF indicators and | Systolic blood pressure | NR |

| Study (Year) | Domain | Generic outcome (as stated by the authors) | Outcome | Outcome measure |
|---|---|---|---|---|
| | | cardiovascular disease metabolic risk | | |
| | | Outcome indexes: body shape indicators, CRF indicators and cardiovascular disease metabolic risk | HRmax | NR |
| | General | Outcome indexes: body shape indicators, CRF indicators and cardiovascular disease metabolic risk | Body mass index (BMI) | NR |
| | | Outcome indexes: body shape indicators, CRF indicators and cardiovascular disease metabolic risk | Percentace body fat | NR |
| | | Outcome indexes: body shape indicators, CRF indicators and cardiovascular disease metabolic risk | Waist circumference | NR |
| | Physical Functioning | Outcome indexes: body shape indicators, CRF indicators and cardiovascular disease metabolic risk | VO2 | NR |

| Study (Year) | Domain | Generic outcome (as stated by the authors) | Outcome | Outcome measure |
|---|---|---|---|---|
| Mijalkovi et al., 2022 | Cardiac | Cardiorespiratory Fitness | Blood Pressure | Automatic arterial blood pressure monitor (ABPM), Shuttle run test (SRT) |
| | | Cardiorespiratory Fitness | Global isovolumetric relaxation time | Comprehensive transthoracic echocardiography (CTE) |
| | | Cardiorespiratory Fitness | Heart rate | Step test (ST) |
| | | Cardiorespiratory Fitness | Heart rate reserve | Shuttle Run Test (SRT) |
| | | Cardiorespiratory Fitness | Left ventricular systolic ejection fraction | Comprehensive transthoracic echocardiography (CTE) |
| | | Cardiorespiratory Fitness | Rest heart rate | Shuttle Run Test (SRT) |
| | Physical Functioning | Cardiorespiratory Fitness | Excess post-exercise oxygen consumption (EPOC) | Treadmill protocol (TRE) |
| | | Cardiorespiratory Fitness | VO2max | 6 min walk test (6MWT) |
| | | Cardiorespiratory Fitness | VO2peak | Treadmill protocol (TRE) |
| Millard-Staffor et al., 2013 | Blood and Lymphatic System | Metabolic risk factor | Apolipoprotein A-1 | NR |
| | | Metabolic risk factor | Apolipoprotein B | NR |
| | | Cardiovascular risk factor | High density lipoprotein (HDL) | NR |
| | | Cardiovascular risk factor | Low density lipoprotein (LDL) | NR |
| | | Cardiovascular risk factor | Serum C-reative protein | NR |
| | | Cardiovascular risk factor | Total cholesterol | NR |
| | | Cardiovascular risk factor | Triglycerides | NR |

| Study (Year) | Domain | Generic outcome (as stated by the authors) | Outcome | Outcome measure |
|---|---|---|---|---|
| | | Metabolic risk factor | Tumor Necrosis Factor-alpha | NR |
| | Cardiac | Blood pressure | Diastolic blood pressure | NR |
| | | Blood pressure | Systolic blood pressure | NR |
| | Cognitive Functioning | Mental/Emotional | Cognitive function | NR |
| | Endocrine | Metabolic risk factor | Adiponectin | NR |
| | | Metabolic risk factor | Fasting glucose | NR |
| | | Metabolic risk factor | Fasting insulin | NR |
| | | Metabolic risk factor | HOMA-index (Homeostatic Model Assessment for Insulin Resistance) | NR |
| | | Metabolic risk factor | IL-6 | NR |
| | | Metabolic risk factor | Insulin sensitivity | NR |
| | | Metabolic risk factor | Insulin-like Growth Factor 1 | NR |
| | | Metabolic risk factor | Leptin | NR |
| | General | Adiposity | Body fat mass (% or kg) | NR |
| | | Adiposity | Body fat-free mass | NR |
| | | Musculoskeletal health | Body fat-free mass | NR |
| | | Adiposity | Body mass index (BMI) | NR |
| | | Adiposity | Visceral adipose tissue | NR |
| | | Adiposity | Waist circumference | NR |

| Study (Year) | Domain | Generic outcome (as stated by the authors) | Outcome | Outcome measure |
|---|---|---|---|---|
| | Musculoskeletal and Connective Tissue | Musculoskeletal health | Bone density | NR |
| | | Musculoskeletal health | Bone mineral content | NR |
| | Physical Functioning | Aerobic fitness | Aerobic fitness test performance | Measured by distance run test or heart response (during/post) or VO2max direct-Bike or VO2max direct-Treadmill or VO2max estimate-Lab test or Walk test |
| | | Muscular strength | Maximum dynamic strength | Measured by 1RM - Lower Body or 1RM - Upper body |
| | | Muscular strength | Maximum isometric strength | Measured by handgrip dynamometer |
| | | Muscular strength | Muscle endurance | Measured by Curl-up or Sit-up or Push-up or Weighted Ball Throw |
| | | Muscular strength | Standing Broad Jump | NR |
| | Psychiatric | Mental/Emotional | Anxiety | NR |
| | | Mental/Emotional | Depression | NR |
| Nagle et al., 2013 | General | Obesity | Body fat mass (%) | NR |
| | | Obesity | Body mass index (BMI) | BMI, BMI-z score |
| | | Obesity | Body weight | NR |
| | | Obesity | Waist circumference | NR |
| Oliveira et al., 2017 | General | Body composition | Body fat mass (%) | Measured by dual energy X-ray absorptiometry or bioelectrical impedance analysis |
| | | Body composition | Body mass index (BMI) | NR |
| | | Body composition | Lean body mass | NR |
| | | Body composition | Waist circumference | Measured by measuring tape |
| | Physical Functioning | Neuromuscular fitness | Agility test | NR |

| **Study (Year)** | **Domain** | **Generic outcome (as stated by the authors)** | **Outcome** | **Outcome measure** |
|---|---|---|---|---|
| | | Flexibility | Flexibility | Sit-and-reach test |
| | | Cardiorespiratory endurance | Maximal power output (POmax) | Measured during maximal cardiopulmonary tests, on a cycle ergometer or on a treadmill |
| | | Muscle strength | Maximum isometric strength | Handgrip muscle strength |
| | | Cardiorespiratory endurance | Peak oxygen consumption (VO2 peak) | Measured during maximal cardiopulmonary tests, on a cycle ergometer or on a treadmill |
| Oosterhoff et al., 2016 | Cardiac | Blood pressure | Diastolic blood pressure | NR |
| | | Blood pressure | Systolic blood pressure | NR |
| | General | Body mass index | Body mass index (BMI) | BMI or, when BMI was not reported, BMI z-scores (i.e. BMI standardized for age and sex) |
| Paes et al., 2015 | Blood and Lymphatic System | Lipid profile | High density lipoprotein (HDL) | NR |
| | | Lipid profile | Low density lipoprotein (LDL) | NR |
| | | Inflammatory status | Proinflammatory cytokines | e.g. proinflammatory adipokine interleukin-6; oxidative metabolites of myeloperoxidase; serum leptin; adiponectin |
| | | Lipid profile | Triglycerides | NR |
| | Cardiac | Hemodynamic Response | Cardiac output | NR |
| | | Cardiometabolic Risk | Cardiometabolic risk | NR |
| | | Hemodynamic Response | Sympathetic Nervous Activity | NR |
| | | Hemodynamic Response | Systolic blood pressure | NR |
| | Physical Functioning | Cardiorespiratory Fitness | Cardiorespiratory fitness | NR |

| Study (Year) | Domain | Generic outcome (as stated by the authors) | Outcome | Outcome measure |
|---|---|---|---|---|
| | | Energy expenditure | Energy expenditure in physical activity | NR |
| | Vascular | Endothelial Dysfunction | Endothelial dysfunction | e.g. endothelial progenitor cells; endothelial vasodilator capacity |
| Parikh and Stratton et al., 2011 | General | Adiposity | Abdominal fat | NR |
| | | Adiposity | Body fat mass (%) | Measured by energy x-ray absorptiometry (DEXA), sum of skinfolds, adiposity equations or hydrostatic weighing |
| | | Adiposity | Body mass index (BMI) | NR |
| | | Adiposity | Waist circumference | NR |
| | Physical Functioning | Cardiorespiratory fitness | Maximal oxygen consumption (VO2max) | Measured by maximal tests, indirect calorimetry, time until exhaustion or endurance time, or a change in exercise HR |
| | | Cardiorespiratory fitness | Peak oxygen consumption (VO2 peak) | Measured by maximal tests, indirect calorimetry, time until exhaustion or endurance time, or a change in exercise HR |
| Recchia et al., 2023 | Psychiatric | depressive symptoms | Depressive symptoms | validated depression rating scale |
| Ribeiro et al., 2022 | Endocrine | Insuline sensitivity | Insulin sensitivity | NR |
| | General | Body composition | Body fat | kg |
| | | Body composition | Body mass index (BMI) | kg/m² |
| | | Body composition | Lean body mass | kg |
| | | Body composition | Waist circumference | cm |
| | Physical Functioning | Cardiorrespiratory fitness | Cardiorespiratory fitness | NR |
| | | Muscle strength | Muscle strength | NR |
| | | Cardiorrespiratory fitness | VO2 max | NR |
| | | Cardiorrespiratory fitness | VO2 peak | NR |

| Study (Year) | Domain | Generic outcome (as stated by the authors) | Outcome | Outcome measure |
|---|---|---|---|---|
| Ruotsalainen et al., 2015 | Emotional functioning/wellbeing | Psychological symptoms | Attitudes about appearance | Measured by questionnaires |
| | | Psychological symptoms | Eating disorder or problematic eating attitudes | Measured by questionnaires |
| | | Psychological symptoms | Physical self-perception or body satisfaction | Measured by questionnaires |
| | | Psychological symptoms | Self-esteem | Measured by questionnaires |
| | General | Body mass index (BMI) | Body mass index (BMI) | BMI kg/m²; BMI z-scores |
| | Global Quality of Life | Psychological symptoms | Quality of life | Measured by questionnaires |
| | Physical Functioning | Physical activity | Physical activity level | Measured objectively using an accelerometer or self-reported |
| | | Physical activity | Physical activity playing active and non-active videogames | Measured by questionnaires |
| | | Physical activity | Sedentary activities | Sedentary activity self-reported |
| | Psychiatric | Psychological symptoms | Depression | Measured by questionnaires |
| Saavedra et al., 2011 | Physical Functioning | Aerobic fitness | Peak oxygen consumption (VO2 peak) | Determined by a maximal or submaximal effort test on a treadmill or cycloergometer, except for one study which gave no information on the method of evaluation |
| Sbruzzi et al., 2013 | Blood and Lymphatic System | Lipid profile | High density lipoprotein (HDL) | NR |
| | | Lipid profile | Total cholesterol (mg/dL) | NR |
| | Cardiac | Blood pressure | Diastolic blood pressure | NR |

| Study (Year) | Domain | Generic outcome (as stated by the authors) | Outcome | Outcome measure |
|---|---|---|---|---|
| | | Blood pressure | Systolic blood pressure | NR |
| | General | Anthropometric | Body mass index (BMI) | BMI kg/m² or BMI z-scores |
| | | Body composition | Body weight | NR |
| | | Anthropometric | Waist circumference (cm) | NR |
| Schranz et al., 2013 | Emotional Functioning/Wellbeing | Psychological well-being | Self-concept | Perception of physical ability and appearance |
| | | Psychological well-being | Self-efficacy | Confidence to complete a given task |
| | | Psychological well-being | Self-esteem | NR |
| | General | Body composition | Body fat mass (% and kg) | Body composition was assessed using two common methodologies by the included studies: anthropometric measures such as girths and skinfolds and dual energy X-ray absorptiometry |
| | | Body composition | Body mass index (BMI) | NR |
| | | Body composition | Lean body mass | Body composition was assessed using two common methodologies by the included studies: anthropometric measures such as girths and skinfolds and dual energy X-ray absorptiometry |
| | | Body composition | Waist circumference | Body composition was assessed using two common methodologies by the included studies: anthropometric measures such as girths and skinfolds and dual energy X-ray absorptiometry |
| | Global Quality of Life | Psychological well-being | Quality of life | Measured by Pediatric Quality of Life Inventory [PedsQL], |
| | Physical Functioning | Strength | Maximum dynamic strength | 1RM for upper and lower body |
| | | Strength | Maximum isometric strength | Isometric knee extension or hand grip strength |

| Study (Year) | Domain | Generic outcome (as stated by the authors) | Outcome | Outcome measure |
|---|---|---|---|---|
| Schwartz et al., 2017 | Blood and Lymphatic System | Metabolic parameters | High density lipoprotein (HDL) | NR |
| | | Metabolic parameters | Low density lipoprotein (LDL) | NR |
| | | Metabolic parameters | Total cholesterol | NR |
| | | Metabolic parameters | Triglycerides | NR |
| | Cardiac | Metabolic parameters | Diastolic blood pressure | NR |
| | | Metabolic parameters | Mean systolic and diastolic blood pressure | NR |
| | | Metabolic parameters | Systolic blood pressure | NR |
| | Endocrine | Appetite regulating hormones | Adiponectin | NR |
| | | Metabolic parameters | Fasting glucose | glucose concentration |
| | | Metabolic parameters | Fasting insulin | insulin concentration |
| | | Appetite regulating hormones | Leptin | NR |
| | | Appetite regulating hormones | Serum AgRP, NPY and α-MSH | NR |
| | General | Body composition | Body fat mass (%) | Body composition was assessed whether by using skin-folds or air-displacement plethysmography or biphotonic absorptiometry or bio-electrical impedance analyses |
| | | Body composition | Body fat-free mass | Body composition was assessed whether by using skin-folds or air-displacement plethysmography or biphotonic absorptiometry or bio-electrical impedance analyses |

| **Study (Year)** | **Domain** | **Generic outcome (as stated by the authors)** | **Outcome** | **Outcome measure** |
|---|---|---|---|---|
| | | Body composition | Body weight | Body composition was assessed whether by using skin-folds or air-displacement plethysmography or biphotonic absorptiometry or bio-electrical impedance analyses |
| | Metabolism and Nutrition | Energy and macronutrient intake | Dietary intake | Self-reported dietary records to assess total energy intake |
| | | Energy and macronutrient intake | Macronutrient intake | Self-reported dietary records to assess protein intake, fat intake and carbohydrate intake |
| Seral-Cortes et al., 2021 | General | Body composition | Body mass index (BMI) | NR |
| Shin et al., 2019 | General | Changes in measured BMI or body weight | Body mass index (BMI) | kg/m² |
| | | Changes in measured BMI or body weight | Body weight | kg |
| | Metabolism and Nutrition | Sugar‑sweetened beverage intake | Dietary behavior | Two items regarding consumption of SSBs; Schools Physical Activity and Nutrition Survey (SPANS); California Health Interview Survey (CHIS) questions |
| | Physical Functioning | Screen time | Sedentary activities | Adolescent Sedentary Activity Questionnaire (ASAQ), California Health Interview Survey (CHIS) questions, Physical Activity Questionnaire for Adolescents (PAQ‑A) |
| | | Physical activity | Time spent in physical activity, Days of physical activity per week | Actigraph accelerometers (counts/min), Physical activity days per week, Average daily activity counts (counts/min); |
| Silva et al., 2014 | Cardiac | Physical fitness | Maximum heart rate (beats/minute) | NR |
| | General | Body composition | Abdominal fat | Measured by dual X-ray absorptiometry (DEXA) |

| Study (Year) | Domain | Generic outcome (as stated by the authors) | Outcome | Outcome measure |
|---|---|---|---|---|
| | | Body composition | Body fat mass (%) | Measured by electrical bioimpedance or using dual X-ray absorptiometry (DEXA) |
| | | Body composition | Body fat-free mass | Measured by electrical bioimpedance or using dual X-ray absorptiometry (DEXA) |
| | | Body composition | Body mass index (BMI) | Measured by z-score de BMI or por el método de suavizado "Latent Moderated Structural" (LMS) or percentil de IMC a través de software; |
| | | Body composition | Body weight | NR |
| | | Body composition | Skinfold thickness | Subescapular and suprailic skinfolds |
| | | Body composition | Waist circumference | NR |
| | Metabolism and Nutrition | Food and Diet | Dietary behavior | Behaviors related to healthy eating; social and emotional aspects related to obesity behavior (Eating Disorder Examination" (EDE-Q) questionnaire for the psychopathological evaluation of eating disorders; and) diagnostic interview "Eating Disorders" (IDed-IV); |
| | | Food and Diet | Dietary intake | Total energy intake; total carbohydrates intake; total fats intake; total sugar intake (measured by questionnaire) |
| | | Physical fitness | Cardiovascular efficiency | NR |
| | | Physical fitness | Global and physical motor abilities | NR |

Physical Functioning

| Study (Year) | Domain | Generic outcome (as stated by the authors) | Outcome | Outcome measure |
|---|---|---|---|---|
| | | Physical fitness | Maximal oxygen consumption (VO2max) | The following tests were used to assess the CF: a) test in cycle ergometer with gas analysis; b) test on a treadmill with gas analysis and determination of VO2max. with incremental workload (speed and incline) until exhaustion; c) application of the test "Course Navette" with modifications; d) analysis of the efficiency of cardiovascular system (VO2 / FC); e) assessment of the basal metabolic rate; f) submaximal field test; g) EUROFIT test battery; h) test up and down steps (3 minutes); i) motor skills test; j) Balke test, and k) walk for 6 minutes. |
| | | Physical fitness | Peak oxygen consumption (VO2 peak) | The following tests were used to assess the CF: a) test in cycle ergometer with gas analysis; b) test on a treadmill with gas analysis and determination of VO2max. with incremental workload (speed and incline) until exhaustion; c) application of the test "Course Navette" with modifications; d) analysis of the efficiency of cardiovascular system (VO2 / FC); e) assessment of the basal metabolic rate; f) submaximal field test; g) EUROFIT test battery; h) test up and down steps (3 minutes); i) motor skills test; j) Balke test, and k) walk for 6 minutes. |
| | | Physical activity | Healthy habits related with PA practice | Measured by “The Family Habit Inventory” and “Child Behaviour Checklist” |
| | | Physical activity | Physical activity level | e.g. PA in free time; AP practice moderate and intense; usual PA practice; time dedicated to the practice of PA (measured by questionnaire and acelerometer and pedometer) |
| | | Physical activity | Sedentary activities | Time devoted to sedentary activities and television (measured by questionnaire and acelerometer and pedometer) |

| Study (Year) | Domain | Generic outcome (as stated by the authors) | Outcome | Outcome measure |
|---|---|---|---|---|
| Sims et al., 2015 | Physical Functioning | Physical activity level | Time spent in moderate to vigorous physical activity | Objective measurement or validated self-report measure (accelerometers; questionnaire) |
| | | Physical activity level | Time spent in physical activity | Objective measurement or validated self-report measure (accelerometers; questionnaire) |
| Sirico et al., 2018 | Blood and Lymphatic System | Inflammatory marker level | Plasminogen activator inhibitor-1 | NR |
| | | Inflammatory marker level | Serum C-reative protein | NR |
| | | Inflammatory marker level | Tumor Necrosis Factor-alpha | NR |
| | Endocrine | Inflammatory marker level | Adiponectin | NR |
| | | Inflammatory marker level | IL-6 | NR |
| | | Inflammatory marker level | Leptin | NR |
| | | Inflammatory marker level | Resistin levels | NR |
| Snethen et al., 2016 | General | Weight loss | Body weight | NR |
| Solera-Martinez et al., 2021 | Blood and Lymphatic System | Cardiometabolic | Glucose | NR |
| | | Cardiometabolic | High density lipoprotein (HDL) | NR |
| | | Cardiometabolic | Low density lipoprotein (LDL) | NR |
| | | Cardiometabolic | Non-High-density lipoproteins (non–HDL-C) | NR |
| | | Cardiometabolic | Total cholesterol | NR |
| | | Cardiometabolic | Triglycerides | NR |

| Study (Year) | Domain | Generic outcome (as stated by the authors) | Outcome | Outcome measure |
|---|---|---|---|---|
| | Cardiac | Blood Pressure | Diastolic blood pressure | NR |
| | | Blood Pressure | Systolic blood pressure | NR |
| | General | Body Composition | Body fat | NR |
| | | Body Composition | Body mass index (BMI) | NR |
| | Physical Functioning | Cardiorespiratory Fitness | VO2peak | NR |
| Spadaccini et al., 2022 | Blood and Lymphatic System | | Lipid profile | NR |
| | Cardiac | | Blood Pressure | NR |
| | | | Heart rate recovery | NR |
| | Emotional Functioning/Wellbeing | | Psycological disorders | NR |
| | | | Psycological well-being | NR |
| | Endocrine | | Adiponectin | NR |
| | | | C-reactive protein | NR |
| | | | Glycaemic profile | NR |
| | | | Leptin | NR |
| | | | Resistin levels | NR |
| | General | | Body circunferences | NR |
| | | | Body composition | NR |
| | | | body weight | NR |
| | | | Height | NR |
| | | | Parent report | NR |
| | | | skinfolds | NR |

| Study (Year) | Domain | Generic outcome (as stated by the authors) | Outcome | Outcome measure |
|---|---|---|---|---|
| | Global Quality of Life | | Quality of life | NR |
| | Metabolism and Nutrition | | Eating desorders | NR |
| | | | Food desire | NR |
| | | | Food intake | NR |
| | Physical Functioning | | Balance | NR |
| | | | fatigue | NR |
| | | | Muscle strength | NR |
| | | | physical performance | NR |
| George et al. (2022) | General | Body fat percentage | Body fat mass (%) | NR |
| | | BMI | Body mass index (BMI) | NR |
| | | Weight status | Body weight | NR |
| | | Waist circunference | Waist circunference | NR |
| | Metabolism and Nutrition | Sugar‑sweetened beverage intake | Dietary behavior | NR |
| | | Fruit and vegetable intake | Portions of fruit, vegetables and water | NR |
| | Physical Functioning | Physical activity | Physical activity level | NR |
| | | Sedentary activities | Sedentary activities | NR |
| Staniford et al., 2011 | General | Weight status/adiposity | Body fat mass (%) | NR |
| | | Weight status/adiposity | Body mass index (BMI) | BMI or BMI-SDS or percent BMI |
| | | Weight status/adiposity | Body weight | NR |
| | | Weight status/adiposity | Percent overweight | NR |
| | | Weight status/adiposity | Skinfold thickness | NR |

| Study (Year) | Domain | Generic outcome (as stated by the authors) | Outcome | Outcome measure |
|---|---|---|---|---|
| | | Weight status/adiposity | Waist circumference | NR |
| Steele et al., 2021 | General | Body composition | Fat mass | kg |
| | | Body composition | Fat-free mass | kg |
| Stoner et al., 2016 | Blood and Lymphatic System | Cardiometabolic risk factors | High density lipoprotein (HDL) | NR |
| | | Cardiometabolic risk factors | Low density lipoprotein (LDL) | NR |
| | | Cardiometabolic risk factors | Total cholesterol | NR |
| | | Cardiometabolic risk factors | Triglycerides | NR |
| | Cardiac | Cardiometabolic risk factors | Systolic blood pressure | NR |
| | Endocrine | Cardiometabolic risk factors | Area under the curve for glucose | NR |
| | | Cardiometabolic risk factors | Fasting glucose | NR |
| | | Cardiometabolic risk factors | Fasting insulin | NR |
| | | Cardiometabolic risk factors | HOMA-index (Homeostatic Model Assessment for Insulin Resistance) | NR |
| | General | Body composition | Body fat mass (%) | Measured by DXA or skinfolds or hydrostatic |
| | | Anthropometric | Body mass index (BMI) | BMI (kg/m²) |
| | | Body composition | Lean body mass | Measured by DXA or skinfolds or hydrostatic |
| Thivel et al., 2018 | Blood and Lymphatic System | Cardiometabolic factors | Glucose | NR |
| | | Cardiometabolic factors | High density lipoprotein (HDL) | NR |

| Study (Year) | Domain | Generic outcome (as stated by the authors) | Outcome | Outcome measure |
|---|---|---|---|---|
| | | Cardiometabolic factors | Low density lipoprotein (LDL) | NR |
| | | Cardiometabolic factors | Total cholesterol | NR |
| | | Cardiometabolic factors | Triglycerides | NR |
| | Cardiac | Blood Pressure | Diastolic blood pressure | NR |
| | | Blood Pressure | Systolic blood pressure | NR |
| | Endocrine | Metabolic profile | Adiponectin | NR |
| | | Metabolic profile | HbA1c | NR |
| | | Metabolic profile | Insulin | NR |
| | | Metabolic profile | Insulin resistance | HOMA-IR |
| | | Metabolic profile | Leptin | NR |
| | | Metabolic profile | NO2- | NR |
| | General | Anthropometric characteristics and Body composition | Body fat % | Dual X-ray Absorptiometry was used in 2 studies . Bio-impedance analysis in 7 studies. Skinfold thickness measurement in 3 studies. Air displacement plethysmography (Bod Pod) is one study. Both BIA and skinfold thickness measurement were used. |
| | | Anthropometric characteristics and Body composition | Body Mass | Dual X-ray Absorptiometry was used in 2 studies . Bio-impedance analysis in 7 studies. Skinfold thickness measurement in 3 studies. Air displacement plethysmography (Bod Pod) is one study. Both BIA and skinfold thickness measurement were used. |
| | | Anthropometric characteristics and Body composition | Body mass index (BMI) | Dual X-ray Absorptiometry was used in 2 studies . Bio-impedance analysis in 7 studies. Skinfold thickness measurement in 3 studies. Air displacement |

| Study (Year) | Domain | Generic outcome (as stated by the authors) | Outcome | Outcome measure |
|---|---|---|---|---|
| | | | | plethysmography (Bod Pod) is one study. Both BIA and skinfold thickness measurement were used. |
| | | Anthropometric characteristics and Body composition | Fat-free mass | Dual X-ray Absorptiometry was used in 2 studies . Bio-impedance analysis in 7 studies. Skinfold thickness measurement in 3 studies. Air displacement plethysmography (Bod Pod) is one study. Both BIA and skinfold thickness measurement were used. |
| | | Anthropometric characteristics and Body composition | Waist Circumf. | Dual X-ray Absorptiometry was used in 2 studies . Bio-impedance analysis in 7 studies. Skinfold thickness measurement in 3 studies. Air displacement plethysmography (Bod Pod) is one study. Both BIA and skinfold thickness measurement were used. |
| | | Anthropometric characteristics and Body composition | zBMI | Dual X-ray Absorptiometry was used in 2 studies . Bio-impedance analysis in 7 studies. Skinfold thickness measurement in 3 studies. Air displacement plethysmography (Bod Pod) is one study. Both BIA and skinfold thickness measurement were used. |
| | Metabolism and Nutrition | Energy intake | Energy intake | Dietary recall (4-day questionnaire) |
| | Physical Functioning | Other fitness parameters | Countermovement jump | Countermov Jump |
| | | Other fitness parameters | Maximal Leg Strenght | NR |
| | | Other fitness parameters | Squat Jump | Squat Jump |
| | | Aerobic fitness | CHO oxidation | 14 studies used an incremental maximal test performed in laboratory with measurement of maximal oxygen uptake (VO2max). YoYo intermittent field test was used in one study. Astrand cycling test was used in one study. A 6 minutes maximal running test was used in 2 studies. |

| Study (Year) | Domain | Generic outcome (as stated by the authors) | Outcome | Outcome measure |
|---|---|---|---|---|
| | | Aerobic fitness | HRmax | 13 studies used an incremental maximal test performed in laboratory with measurement of maximal oxygen uptake (VO2max). YoYo intermittent field test was used in one study. Astrand cycling test was used in one study. A 6 minutes maximal running test was used in 2 studies. |
| | | Aerobic fitness | Max Aerob Sp | 16 studies used an incremental maximal test performed in laboratory with measurement of maximal oxygen uptake (VO2max). YoYo intermittent field test was used in one study. Astrand cycling test was used in one study. A 6 minutes maximal running test was used in 2 studies. |
| | | Aerobic fitness | RER | 15 studies used an incremental maximal test performed in laboratory with measurement of maximal oxygen uptake (VO2max). YoYo intermittent field test was used in one study. Astrand cycling test was used in one study. A 6 minutes maximal running test was used in 2 studies. |
| | | Aerobic fitness | VO2max | 11 studies used an incremental maximal test performed in laboratory with measurement of maximal oxygen uptake (VO2max). YoYo intermittent field test was used in one study. Astrand cycling test was used in one study. A 6 minutes maximal running test was used in 2 studies. |
| | | Aerobic fitness | VO2peak | 12 studies used an incremental maximal test performed in laboratory with measurement of maximal oxygen uptake (VO2max). YoYo intermittent field test was used in one study. Astrand cycling test was used in one study. A 6 minutes maximal running test was used in 2 studies. |
| | | Aerobic fitness | ΔHRR1 | 17 studies used an incremental maximal test performed in laboratory with measurement of maximal oxygen uptake (VO2max). YoYo intermittent field test was used in one |

| Study (Year) | Domain | Generic outcome (as stated by the authors) | Outcome | Outcome measure |
|---|---|---|---|---|
| | | | | study. Astrand cycling test was used in one study. A 6 minutes maximal running test was used in 2 studies. |
| | | Aerobic fitness | ΔHRR2 | 18 studies used an incremental maximal test performed in laboratory with measurement of maximal oxygen uptake (VO2max). YoYo intermittent field test was used in one study. Astrand cycling test was used in one study. A 6 minutes maximal running test was used in 2 studies. |
| | Vascular | Endothelial Function | Endothelial function | Flow-mediated dilatation |
| Upton et al., 2014 | General | Weight change | Body mass index (BMI) | BMI; BMI percentile; BMI z-score/SDS |
| | | Weight change | Body weight | NR |
| Valeriani et al., 2021 | General | Weight status | Body mass index (BMI) | NR |
| | | Weight status | zBMI | NR |
| van Wijnen et al., 2009 | Cognitive Functioning | Psychosocial | Cognitive competence (self-perception) | Measured by Harter-Perceived Competence Scale for Children (Harter, 1982) |
| | Delivery of Care | Psychosocial | Unhealthy weight control behaviours (self-perception) | List of nine unhealthy and five health weight control methods: respondents indicate use of each method over past month (yes/no) |
| | Emotional functioning/wellbeing | Psychosocial | Athletic competence | Measured by Perceived Competence Scale for Children |
| | | Psychosocial | Binge eating (self-perception) | One question: ‘In the past month, have you ever eaten so much food in a short period of time that you would be embarrassed if other saw you (binge eating)? |
| | | Psychosocial | Body image (self-perception) | Satisfaction with body image measure by adapted body shape perception scale; Body image perception measure by Figural Rating Scale (4th–7th grades; Stunkard et al., 1983) & Schematic Figures for kindergarten–3rd grade (Collins, 1991) |

| Study (Year) | Domain | Generic outcome (as stated by the authors) | Outcome | Outcome measure |
|---|---|---|---|---|
| | | Psychosocial | Healthy attitudes (self-perception) | e.g. Dietary restraint, Disordered eating |
| | | Psychosocial | Media internalization (self-perception) | Four questions assessing internalization of media ideals for appearance/body shape (Heinberg et al., 1995; Neumark-Sztainer et al., 2000) |
| | | Psychosocial | Self-acceptance (self-perception) | Measured by Perceived Competence Scale for Children (Harter, 1982) |
| | | Psychosocial | Self-esteem (self-perception) | NR |
| | | Psychosocial | Self-worth (self-perception) | Global or general self-worth measured by Harter-Perceived Competence Scale for Children |
| | Social Functioning | Psychosocial | Aggression (self-perception) | Peer-rated aggression measure by fifteen questions about the behaviour of classmates including aggressive behaviour items, popularity items and prosocial items (Eron et al., 1971; Walder et al., 1961); Observed physical and verbal aggression measure by direct observation during free play, using the protocol by Joy et al. (1986); Parent report of aggressive and delinquent behaviour measure by Parent report form of the Child Behaviour Checklist (Achenbach, 1991); Perceptions of a mean and scary world |
| | | Psychosocial | Social acceptance (self-perception) | Measured by Perceived Competence Scale for Children (Harter, 1982) |
| Vasconcellos et al., 2014 | Blood and Lymphatic System | Inflammatory profile | Fibrinogen level | NR |
| | | Biochemical markers | High density lipoprotein (HDL) | NR |
| | | Biochemical markers | Low density lipoprotein (LDL) | NR |

| Study (Year) | Domain | Generic outcome (as stated by the authors) | Outcome | Outcome measure |
|---|---|---|---|---|
| | | Inflammatory profile | Serum C-reative protein | NR |
| | | Biochemical markers | Total cholesterol | NR |
| | Cardiac | Hemodynamic | Diastolic blood pressure | NR |
| | | Hemodynamic | Resting heart rate (beats/minute) and Maximum heart rate (beats/minute) | NR |
| | | Hemodynamic | Systolic blood pressure | NR |
| | Endocrine | Inflammatory profile | Adiponectin | NR |
| | | Biochemical markers | Fasting glucose | NR |
| | | Biochemical markers | Fasting insulin | NR |
| | | Inflammatory profile | IL-6 | NR |
| | General | Body composition | Body fat mass (%) | NR |
| | | Body composition | Body mass index (BMI) | NR |
| | | Body composition | Waist circumference | NR |
| | Physical Functioning | Cardiorespiratory fitness | Cardiorespiratory fitness | NR |
| | | Muscle strength | Muscle strength | NR |
| | Vascular | Endothelial function | Flow-mediated dilation (FMD) | NR |
| Verjans-Janssen et al., 2018 | General | BMI | Body mass index (BMI) | BMI measured by z-score or percentile for age and gender |

| Study (Year) | Domain | Generic outcome (as stated by the authors) | Outcome | Outcome measure |
|---|---|---|---|---|
| | Metabolism and Nutrition | Nutrition behavior | Dietary intake | Fruit intake (exchanges/day or portions/day); Vegetable intake (exchanges/day or portions/day); Dairy intake (exchanges/day); Fats and oils intake (exchanges/day); Meat intake (exchanges/day); Grains intake (exchanges/day); Sweets and beverages intake (exchanges/day); Sugar-sweetened beverages intake (portions/day); Soda intake (portions/day); Chocolate and candy intake (portions/day); Snack intake (portions/day); Water consumption (servings/day); Saturated fat intake (mg/weekday); Total fat (g/day); Cholesterol (g/day); Protein (g/day); Carbohydrate (g/day); Fiber (g/day) |
| | Physical Functioning | Sedentary behavior | Sedentary activities | Sitting (h/day); TV watching (h/day); Computer and video games (h/day); Sedentary behavior (hours of screen viewing/day); TV viewing (frequency of TV viewing while getting ready for school; Time spent sedentary (min/day); TV viewing week day / weekend (% children watching TV 2 hours or less per week day / weekend) |
| | | Physical activity | Time spent in physical activity | MVPA (min/day); Outdoor play (h/day); Physical education (h/week); Supervised sports or dancing (h/week); Time spent in outdoor activities (h/week); Time spent in extracurricular sports courses (h/week); PA behavior (PA behavior compared to other children of same age and sex); Team sports; Light PA (min/day); Leisure-time MVPA (h/week); PA (% Children performing daily PA); Active > 60 min (days/week); Active games at lunchtime (% children performing active games at lunchtime); Being outside yesterday (% children being outside 2 hours or more after school yesterday); Being outside weekend day (% children being outside 2 hours or more on a weekend day); Jogging/running frequency (% children with improved |

| Study (Year) | Domain | Generic outcome (as stated by the authors) | Outcome | Outcome measure |
|---|---|---|---|---|
| | | | | frequency); Walking frequency (% children with improved frequency); Ball playing (% children improved frequency); Walking or riding bicycles to school (% children improved frequency) |
| Vissers et al., 2016 | General | Visceral adipose tissue | Visceral adipose tissue (VAT) | VAT assessed by medical imaging (computed tomography, magnetic ressonance imaging, ultrasound) |
| Voskuil et al., 2017 | General | Body composition | Body fat mass (%) | Bioelectrical impedance analysis, dual-energy x-ray absorptiometry and estimation of % bpdy fat from skin fold measurements |
| | | Body composition | Body mass index (BMI) | NR |
| | Physical Functioning | Accelerometer-measured physical activity | Physical activity level | Accelerometer-measured moderate-to-vigorous physical activity |
| Wang et al., 2022 | General | Visceral Fat | visceral adipose tissue | NR |
| Whitlock et al., 2010 | General | Weight change | Body mass index (BMI) | Change in BMI from baseline as the preferred measure of weight change when it was available. If BMI change was unavailable and could not be calculated or obtained from the author, we used change in BMI standard deviation score (SDS) as our second choice and change in percent overweight as the third choice. |
| Wolfa et al., 2019 | General | Obesity | Body mass index (BMI) | NR |
| | | Obesity | Waist Circunference | NR |
| Xu et al., 2022 | General | weight loss | body fat mass | NR |
| | | weight loss | Body mass index (BMI) | NR |
| | | weight loss | body weight | NR |
| Yukel et al., 2020 | Cardiac | Cardiovascular risk | Cardiovascular risk | An automated oscillograph |

| Study (Year) | Domain | Generic outcome (as stated by the authors) | Outcome | Outcome measure |
|---|---|---|---|---|
| | Emotional Functioning/Wellbeing | Healthy lifestyle | Healthy lifestyle | A self-administered non-quantitative FFQ |
| | | Quality of life | Quality of life | Health questionnaire |
| | General | Body fat percentage | Body fat percentage | Percentage of body fat was calculated from triceps and subscapular skinfolds measurements |
| | | Obesity | Body mass index (BMI) | kg/m² - Tanita BC 418MA Segmental Body Composition Analyzer |
| | | Obesity | Height | Seca Leicester Height Measure stadiometer, stadiometer, portable stadiometer, standing stadiometer (Seca model 720, Germany). |
| | | Obesity | Skinfold | mm - Harpenden calipers (HSK BI, British Indicators) |
| | | Obesity | Waist circunference | A measuring tape between the lower rib and the iliac-crest; Flexible tape |
| | | Obesity | Weight | SecaTM 899 digital scales, portable digital scale, lever scale |
| | Metabolism and Nutrition | Daily fruit and vegetable consumption | Daily fruit and vegetable consumption | A Day in the Life Questionnaire |
| | | Diet | Diet | Self-completed questionnaires |
| | Physical Functioning | Physical Fitness | Aerobic fitness | 20m shuttle run |
| | | Physical Fitness | Cardiorespiratory fitness | Monark ergometer bike (W/kg), 20m shuttle run |
| | | Physical Fitness | Peak power | calibrated friction loaded ergometer. |
| | | Physical Activity | Physical activity level | Actigraph, pedometer, questionare, accelerometer, CLASS questionare |
| | | Physical Fitness | Physical fitness | 1 min of sit‑ups, 1 min of push‑ups, a measured vertical jump, a measured long jump, a stand-and‑reach test, a timed 30‑m sprint, 20m shuttle run, 1 mile run, sit and reach, curl-ups, accelerometer |

| Study (Year) | Domain | Generic outcome (as stated by the authors) | Outcome | Outcome measure |
|---|---|---|---|---|
| | | Sedentary behaviour | Sedentary behaviour | Accelerometer |
| Zabatiero et al., 2018 | General | Sedentary behaviour | Body mass index (BMI) | NR |
| | Global Quality of Life | Sedentary behaviour | Health-related QOL | Pedriatric QOL Inventory |
| | Physical Functioning | Sedentary behaviour | sedentary time | accelerometry and inclinometry |
| | | Sedentary behaviour | time spent in MVPA | accelerometry and inclinometry |
| Zhao et al., 2022-a | Endocrine | Inflammatory Response | C-reactive protein | NR |
| | | Inflammatory Response | IL-6 | NR |
| | | Inflammatory Response | TNF-α | NR |
| Zhao et al., 2022-b | Blood and Lymphatic System | Cardiometabolic Factors | Glucose | NR |
| | | Cardiometabolic Factors | High density lipoprotein (HDL) | NR |
| | | Cardiometabolic factors | Low density lipoprotein (LDL) | NR |
| | | Cardiometabolic Factors | Non-High-density lipoproteins (non–HDL-C) | NR |
| | | Inflammatory Markers | TNF-alfa | NR |
| | | Cardiometabolic Factors | Total cholesterol | NR |
| | | Cardiometabolic Factors | Triglycerides | NR |
| | Endocine | Inflammatory Markers | C-reactive protein | NR |
| | | Cardiometabolic Factors | Adiponectin | NR |
| | | Cardiometabolic Factors | Chemerin | NR |
| | | Cardiometabolic Factors | IGF-1 | NR |

| Study (Year) | Domain | Generic outcome (as stated by the authors) | Outcome | Outcome measure |
|---|---|---|---|---|
| | | Cardiometabolic Factors | IGFBP-3 | NR |
| | | Inflammatory Markers | IL-6 | NR |
| | | Cardiometabolic Factors | Insulin resistance | HOMA-IR |
| | | Cardiometabolic Factors | Leptin | NR |
| | General | Anthropometric | Biceps circumference | NR |
| | | Anthropometric | Body mass index (BMI) | NR |
| | | Anthropometric | Body weight | NR |
| | | Anthropometric | Fat mass | NR |
| | | Anthropometric | Forearm circumference | NR |
| | | Anthropometric | Hip circumference | NR |
| | | Anthropometric | Limbs circumference | NR |
| | | Anthropometric | thigh circumference | NR |
| | | Anthropometric | torso circumference | NR |
| | | Anthropometric | Waist circumference | NR |
| | | Anthropometric | Waist circumference | NR |
| | | Anthropometric | waist/hip ratio | NR |
| Zhu et al., 2021 | General | obesity | Body fat percentage | NR |
| | | obesity | Body mass index (BMI) | NR |
| | | obesity | Body weight | NR |
| | | obesity | Waist circumference | NR |
| Zouhal et al., 2020 | Cardiac | Physical performance | HRmax | NR |

| Study (Year) | Domain | Generic outcome (as stated by the authors) | Outcome | Outcome measure |
|---|---|---|---|---|
| | General | Anthropometric characteristcs | Body fat % | NR |
| | | Anthropometric characteristcs | Body mass index (BMI) | NR |
| | | Anthropometric characteristcs | Body weight | NR |
| | | Anthropometric characteristcs | Fat-free mass | NR |
| | Physical Functioning | Physical performance | Lactate threshold | NR |
| | | Physical performance | Maximal strength | NR |
| | | Physical performance | Mean power | NR |
| | | Physical performance | Peak power | NR |
| | | Physical performance | Running economy | NR |
| | | Physical performance | Sprint performance | NR |
| | | Physical performance | Ventilatory threshold | NR |
| | | Physical performance | VO2max | NR |
| Liu et al., 2024 | Cardiac | Blood pressure | Blood pressure | NR |
| | General | Body composition | BMI | NR |
| | | Body composition | BMI-z | NR |
| | | Body composition | Fat mass | NR |
| | | Body composition | Body fat % | NR |
| | Physical Functioning | Cardiorespiratory fitness | VO2max | NR |
| | | | Muscular strenght | NR |

| **Study (Year)** | **Domain** | **Generic outcome (as stated by the authors)** | **Outcome** | **Outcome measure** |
|---|---|---|---|---|
| Dos Santos et al., 2021 | Blood and Lymphatic System | | Total cholesterol | NR |
| | | | Low density lipoprotein (LDL) | NR |
| | Cardiac | | Resting heart rate and Maximum heart rate | NR |
| | Endocrine | | Fasting blood glucose | NR |
| | Physical Functioning | | Time spent in physical activity | NR |
| | | | Daily sedentary time | NR |
| García-Hermoso et al., 2020-c | Cardiac | | Systolic blood pressure | NR |
| | | | Diastolic blood pressure | NR |
| | General | | Body mass index (BMI) | NR |
| | | | Waist circumference | NR |
| | | | Body fat | NR |
| | | | Fat-free mass | NR |
| | Physical Functioning | | Agility | NR |
| Da Silva et al., 2020 | Endocrine | Plasma adipokines | Adiponectin | NR |
| | | Plasma adipokines | Leptin | NR |
| | | Plasma adipokines | Adipocyte fat (aFABP) | NR |
| Comeras-Chueca et al., 2021 | General | Health-related physical fitness | Body mass index (BMI) | dual-energy x-ray absorpiometry or bioeletrical impedance |
| | | Health-related physical fitness | Body weight | dual-energy x-ray absorpiometry or bioeletrical impedance |

| Study (Year) | Domain | Generic outcome (as stated by the authors) | Outcome | Outcome measure |
|---|---|---|---|---|
| | | Health-related physical fitness | Body fat % | dual-energy x-ray absorpiometry or bioeletrical impedance |
| | | Health-related physical fitness | Free-fat mass | dual-energy x-ray absorpiometry or bioeletrical impedance |
| | | Health-related physical fitness | Waist circumference | dual-energy x-ray absorpiometry or bioeletrical impedance |
| | Physical Functioning | Health-related physical fitness | Cardiorespiratory fitness | 20-m shuttle run test, 3-min step test or submaximal test with a cycle ergometer |
| | | Health-related physical fitness | Muscular fitness | 20-m shuttle run test, 3-min step test or submaximal test with a cycle ergometer |
| | | Motor competence | Motor competence | Bruininks-Oseretsky Test and Movement Assessment Battery for Children Test-Second Edition |
| Liu et al., 2020 | Blood and Lymphatic System | Cardiometabolic risk factors | Total cholesterol | NR |
| | | Cardiometabolic risk factors | High density lipoprotein (HDL) | NR |
| | | Cardiometabolic risk factors | Low density lipoprotein (LDL) | NR |
| | Cardiac | Cardiometabolic risk factors | Systolic blood pressure | NR |
| | | Cardiometabolic risk factors | Diastolic blood pressure | NR |
| | Endocrine | Cardiometabolic risk factors | Fasting blood glucose | NR |
| | | Cardiometabolic risk factors | Fasting insulin | NR |
| | | Cardiometabolic risk factors | HOMA-index (Homeostatic Model Assessment for Insulin Resistance) | NR |

| Study (Year) | Domain | Generic outcome (as stated by the authors) | Outcome | Outcome measure |
|---|---|---|---|---|
| | General | Cardiometabolic risk factors | Body mass index (BMI) | NR |
| | | Cardiometabolic risk factors | Body weight | NR |
| | | Cardiometabolic risk factors | Waist circumference | NR |
| | Physical Functioning | Cardiometabolic risk factors | VO2peak | NR |

**eTable 4. Clinical trials included.**

| Trials_1st_author | Trial_DOI | Trial_year | Participants_sample_size |
|---|---|---|---|
| CL Davis | 10.1001/2012.jama.10762 | 2012 | 222 |
| S L Gortmaker | 10.1001/archpedi.153.4.409 | 1999 | 1295 |
| Nader PR | 10.1001/archpedi.153.7.695 | 1999 | 3714 |
| BA Denninson | 10.1001/archpedi.158.2.170 | 2004 | 77 |
| R P Treviño | 10.1001/archpedi.158.9.911 | 2004 | 1419 |
| Aaron L Carrel | 10.1001/archpedi.159.10.963 | 2005 | 50 |
| Karen J Coleman | 10.1001/archpedi.159.3.217 | 2005 | 896 |
| DR Young | 10.1001/archpedi.160.12.1255 | 2006 | 221 |
| Kevin Patrick | 10.1001/archpedi.160.2.128 | 2006 | 878 |
| A L Carrel | 10.1001/archpedi.161.6.561 | 2007 | 53 |
| D M Janicke | 10.1001/archpedi.162.12.1119 | 2008 | 93 |
| DL Paineau | 10.1001/archpediatrics.2007.2 | 2008 | 477 |
| DL Weintraub | 10.1001/archpediatrics.2007.43 | 2008 | 21 |
| Epstein | 10.1001/archpediatrics.2007.45 | 2008 | 70 |
| AS Sighn | 10.1001/archpediatrics.2009.2 | 2009 | 1108 |
| Robert C. Klesges | 10.1001/archpediatrics.2010.196 | 2010 | 303 |
| Thomas N Robinson | 10.1001/archpediatrics.2010.197 | 2010 | 261 |
| Nicole P M Ezendam | 10.1001/archpediatrics.2011.204 | 2012 | 883 |
| Elsie M Taveras | 10.1001/archpediatrics.2011.44 | 2011 | 457 |
| Gianni Bocca | 10.1001/archpediatrics.2012.1638 | 2012 | 75 |
| Lubans, D.R | 10.1001/archpediatrics.2012.41 | 2012 | 357 |
| Ingul | 10.1001/arquipediatria.2010.158 | 2010 | 10 |
| Bocca | 10.1001/arquipediatria.2012.1638 | 2012 | 75 |
| Killen, J.D | 10.1001/jama.1988.03410120074030 | 1988 | 1447 |
| R V Luepker | 10.1001/jama.1996.03530340032026 | 1996 | 5106 |
| Davis | 10.1001/jama.2015.16174. | 2012 | 205 |
| Barkin | 10.1001/jama.2018.9128 | 2018 | 610 |
| T N Robinson | 10.1001/jama.282.16.1561 | 1999 | 225 |
| M Savoye | 10.1001/jama.297.24.2697 | 2007 | 209 |
| Denise E Wilfley | 10.1001/jama.298.14.1661 | 2007 | 204 |
| Sigal | 10.1001/jamapediatria.2014.1392 | 2014 | 304 |
| Madsen | 10.1001/jamapediatrics.2013.1071 | 2013 | 156 |
| Jess Haines | 10.1001/jamapediatrics.2013.2356 | 2013 | 121 |
| SG Trost | 10.1001/jamapediatrics.2013.3436 | 2014 | 75 |
| Robert G Santos | 10.1001/jamapediatrics.2013.3688 | 2014 | 687 |

| Trials_1st_author | Trial_DOI | Trial_year | Participants_sample_size |
|---|---|---|---|
| Ronald J. Sigal | 10.1001/jamapediatrics.2014.1392 | 2014 | 304 |
| O van der Baan-Slootweg | 10.1001/jamapediatrics.2014.521 | 2014 | 90 |
| E M Taveras | 10.1001/jamapediatrics.2015.0182 | 2015 | 549 |
| E M Taveras | 10.1001/jamapediatrics.2017.1325 | 2017 | 721 |
| D Wilfley | 10.1001/jamapediatrics.2017.2960 | 2017 | 172 |
| DS Buchan | 10.1002/ajhb.21166 | 2011 | 57 |
| A Dalziell | 10.1002/berj.3514 | 2019 | 143 |
| Levine | 10.1002/eat.1091 | 2001 | 24 |
| Donnelly JE | 10.1002/j.1550-8528.1996.tb00541.x | 1996 | 338 |
| RM Daly | 10.1002/jbmr.2688 | 2016 | 727 |
| AE Staiano | 10.1002/oby.20210 | 2017 | 33 |
| Fitzgibbon | 10.1002/oby.20269. | 2017 | 147 |
| Amanda E Staiano | 10.1002/oby.20282 | 2013 | 54 |
| JA Wright | 10.1002/oby.20388 | 2013 | 50 |
| P Obert | 10.1002/oby.20495 | 2013 | 28 |
| CE Krafft | 10.1002/oby.20518 | 2014 | 43 |
| A G Alexander | 10.1002/oby.20557 | 2014 | 749 |
| Haines | 10.1002/oby.21314 | 2016 | 112 |
| Mazurak | 10.1002/oby.21355 | 2016 | 87 |
| Butte | 10.1002/oby.21929 | 2017 | 549 |
| CY Anderson | 10.1002/oby.21967 | 2017 | 103 |
| Soltero | 10.1002/oby.22300 | 2018 | 136 |
| Da Silva | 10.1002/ppul.21502 | 2012 | 76 |
| P Mucci | 10.1002/ppul.22646 | 2013 | 18 |
| Elloumi | 10.1002/pri.470 | 2011 | 28 |
| Vandongen, R | 10.1006/pmed.1995.1003 | 1995 | 1240 |
| TL McKenzie | 10.1006/pmed.1996.0074 | 1996 | 5.106 |
| Manois Y | 10.1006/pmed.1998.0388 | 2002 | 1046 |
| Hamelink-Basteen | 10.1007/BF03086988 | 2008 | 426 |
| Lazzer | 10.1007/BF03346399 | 2008 | 19 |
| Hasselstrom et al | 10.1007/s00223-008-9166-x | 2008 | 243 |
| Y Ning | 10.1007/s00394-013-0612-9 | 2014 | 145 |
| C Nourry | 10.1007/s00421-005-1341-4 | 2005 | 18 |
| Hara | 10.1007/s00421-005-1374-8 | 2005 | 21 |
| F X Gamelin | 10.1007/s00421-008-0955-8 | 2009 | 22 |
| G Racil | 10.1007/s00421-013-2689-5 | 2013 | 34 |

| Trials_1st_author | Trial_DOI | Trial_year | Participants_sample_size |
|---|---|---|---|
| L P Bharath | 10.1007/s00421-018-3898-8 | 2018 | 40 |
| Sung | 10.1007/s00421-018-4051-4 | 2019 | 40 |
| Eliakim, A | 10.1007/s00431-002-0980-2 | 2002 | 202 |
| S Amaro | 10.1007/s00431-006-0153-9 | 2006 | 241 |
| Maciste H Macias-Cervantes | 10.1007/s00431-008-0907-7 | 2009 | 76 |
| Thivel | 10.1007/s00431-011-1466-x | 2011 | 457 |
| J Woo | 10.1007/s00431-011-1518-2 | 2012 | 39 |
| P Delgado-Floody | 10.1007/s00431-018-3149-3 | 2018 | 197 |
| D Coknaz | 10.1007/s00431-019-03457-x | 2019 | 106 |
| Wunram | 10.1007/s00787-017-1071-2 | 2018 | 64 |
| JE Chavarro | 10.1007/s10552-005-0404-5 | 2005 | 508 |
| Alaimo | 10.1007/s10900-015-0005-5 | 2015 | 410 |
| Messiah | 10.1007/s10900-017-0393-9 | 2018 | 2261 |
| Sharma | 10.1007/s10935-015-0395-2 | 2015 | 57 |
| Ketelhut | 10.1007/s11332-018-0463-0. | 2018 | 160 |
| De la CruzMunoz | 10.1007/s11695-012-0730-0 | 2013 | 71 |
| Campos | 10.1007/s12020-012-9613-3 | 2012 | 40 |
| Huang | 10.1007/s12020-012-9808-7 | 2013 | 65 |
| Amanda L Penko | 10.1007/s12160-010-9164-x | 2010 | 24 |
| Noe C Crespo | 10.1007/s12160-011-9332-7 | 2012 | 808 |
| M Kalavainen | 10.1007/s12519-011-0324-2 | 2011 | 70 |
| James J Annesi | 10.1007/s12529-013-9361-7 | 2013 | 2308 |
| K A Dias | 10.1007/s40279-017-0777-0 | 2018 | 99 |
| N Andre | 10.1007/s40519-015-0219-7 | 2015 | 16 |
| S Lazzer | 10.1007/s40618-016-0551-4 | 2017 | 30 |
| Norris | 10.1016/0022-3999(92)90114-H | 1992 | 147 |
| Epstein | 10.1016/0306-4603(94)90038-8 | 1994 | 44 |
| J F Lisón | 10.1016/j.acap.2012.03.003 | 2012 | 110 |
| Falbe J | 10.1016/j.acap.2015.02.004 | 2015 | 55 |
| LS Webber | 10.1016/j.amepre.2007.11.018 | 2008 | 1.712 |
| P A Estabrooks | 10.1016/j.amepre.2008.09.024 | 2009 | 135 |
| Dianne Neumark-Sztainer | 10.1016/j.amepre.2010.07.017 | 2010 | 356 |
| Tom Baranowski | 10.1016/j.amepre.2010.09.029 | 2011 | 133 |
| Emilie Bonsergent | 10.1016/j.amepre.2012.09.055 | 2013 | 3538 |
| Olvera | 10.1016/j.amepre.2012.11.018 | 2013 | 122 |
| Castro | 10.1016/j.amepre.2012.11.024 | 2013 | 95 |

| Trials_1st_author | Trial_DOI | Trial_year | Participants_sample_size |
|---|---|---|---|
| Boudreau | 10.1016/j.amepre.2012.11.026 | 2013 | 41 |
| Deborah L Dewar | 10.1016/j.amepre.2013.04.014 | 2013 | 357 |
| Bernadette M Melnyk | 10.1016/j.amepre.2013.05.013 | 2013 | 779 |
| Z J Cao | 10.1016/j.amepre.2014.12.014 | 2015 | 2446 |
| Mendoza | 10.1016/j.amepre.2015.09.017 | 2016 | 160 |
| Soltero | 10.1016/j.amepre.2018.07.034 | 2019 | 68 |
| Prado | 10.1016/j.amepre.2020.06.010 | 2020 | 280 |
| Ounis | 10.1016/j.ando.2009.03.003 | 2009 | 27 |
| CW Kim | 10.1016/j.anr.2015.07.006 | 2016 | 42 |
| OK Ham | 10.1016/j.anr.2016.03.003 | 2016 | 75 |
| Frenn M | 10.1016/j.apnr.2004.04.003 | 2005 | 103 |
| Diane Berry | 10.1016/j.apnr.2006.01.007 | 2007 | 80 |
| MM Covell | 10.1016/j.apnr.2006.12.004 | 2008 | 62 |
| Lakes | 10.1016/j.appdev.2004.04.002 | 2004 | 209 |
| Helen M Hendy | 10.1016/j.appet.2011.01.024 | 2011 | 382 |
| J Carnier | 10.1016/j.appet.2013.05.018 | 2013 | 26 |
| P Warshburger | 10.1016/j.appet.2016.04.007 | 2016 | 685 |
| MacKelvie | 10.1016/j.bone.2003.12.017 | 2004 | 64 |
| RC Nogueira | 10.1016/j.bone.2014.08.006 | 2014 | 151 |
| Benson | 10.1016/j.cct.2006.11.004 | 2008 | 66 |
| Bishop | 10.1016/j.childyouth.2018.01.037 | 2018 | 122 |
| Garcıa-Morales | 10.1016/j.clinthera.2006.05.008 | 2006 | 46 |
| T Reinehr | 10.1016/j.clnu.2009.12.010 | 2010 | 66 |
| N Vernadakis | 10.1016/j.compedu.2015.01.001 | 2015 | 66 |
| Benavides | 10.1016/j.ctcp.2008.12.004 | 2009 | 14 |
| Dreimane | 10.1016/j.diabres.2006.05.017 | 2007 | 264 |
| K Casazza | 10.1016/j.eatbeh.2006.01.007 | 2007 | 5 |
| Craeynest | 10.1016/j.eatbeh.2007.03.002 | 2008 | 38 |
| L H Epstein | 10.1016/j.eatbeh.2015.02.001 | 2015 | 24 |
| Vissers, D | 10.1016/j.eclnm.2008.05.002 | 2008 | 76 |
| Daniel Klein | 10.1016/j.eclnm.2010.03.002 | 2010 | 1050 |
| Espinoza-Silva | 10.1016/j.endinu.2019.05.005 | 2019 | 274 |
| Esteve Llargués | 10.1016/j.endonu.2012.03.002 | 2012 | 2010 |
| B Horsak | 10.1016/j.gaitpost.2019.02.032 | 2019 | 51 |
| K Wright | 10.1016/j.ijnurstu.2012.09.004 | 2013 | 251 |
| Jae-Young Jeon | 10.1016/j.imr.2013.10.001 | 2013 | 15 |

| Trials_1st_author | Trial_DOI | Trial_year | Participants_sample_size |
|---|---|---|---|
| K Watts | 10.1016/j.jacc.2004.01.032 | 2004 | 19 |
| A A Meyer | 10.1016/j.jacc.2006.07.035 | 2006 | 67 |
| N J Farpour-Lambert | 10.1016/j.jacc.2009.08.030 | 2009 | 44 |
| Sanches | 10.1016/j.jacl.2014.02.007 | 2014 | 79 |
| Savoye | 10.1016/j.jada.2004.12.009 | 2005 | 25 |
| R G Díaz | 10.1016/j.jada.2009.10.042 | 2010 | 76 |
| J N Davis | 10.1016/j.jada.2011.05.009 | 2011 | 104 |
| Marks J.T | 10.1016/j.jadohealth.2005.11.002 | 2006 | 319 |
| Haerens L | 10.1016/j.jadohealth.2006.09.028 | 2007 | 2840 |
| Chehab | 10.1016/j.jadohealth.2006.12.009 | 2007 | 46 |
| BH Knöpfli | 10.1016/j.jadohealth.2007.08.015 | 2008 | 128 |
| AC Doyle | 10.1016/j.jadohealth.2008.01.011 | 2008 | 80 |
| Jyu-Lin Chen | 10.1016/j.jadohealth.2010.11.243 | 2011 | 54 |
| Robin Whittemore | 10.1016/j.jadohealth.2012.07.014 | 2013 | 384 |
| Johnston | 10.1016/j.jadohealth.2013.04.001 | 2007 | 71 |
| J L Chen | 10.1016/j.jadohealth.2018.08.022 | 2019 | 40 |
| D Inoue | 10.1016/j.jdiacomp.2014.11.002 | 2015 | 45 |
| Toulabi | 10.1016/j.jfma.2011.05.007 | 2012 | 152 |
| Leach RA, Yates JM | 10.1016/j.jmpt.2008.06.003 | 2008 | 22 |
| Weigel | 10.1016/j.jneb.2007.07.009 | 2008 | 37 |
| Hammons | 10.1016/j.jneb.2013.01.023 | 2013 | 73 |
| G L George | 10.1016/j.jneb.2015.12.010 | 2016 | 126 |
| CW Kim | 10.1016/j.jnutbio.2010.10.001 | 2010 | 25 |
| Edson Dos Santos Farias | 10.1016/j.jped.2014.06.004 | 2015 | 386 |
| K Watts | 10.1016/j.jpeds.2004.02.027 | 2004 | 28 |
| A S Kelly | 10.1016/j.jpeds.2004.08.004 | 2004 | 25 |
| P Balagopal | 10.1016/j.jpeds.2004.11.033 | 2005 | 21 |
| ML Fitzgibbon | 10.1016/j.jpeds.2004.12.019 | 2005 | 362 |
| Aguilar | 10.1016/j.jpeds.2009.12.046 | 2010 | 1044 |
| Okely AD | 10.1016/j.jpeds.2010.03.028 | 2010 | 165 |
| Nemet | 10.1016/j.jpeds.2010.10.040 | 2011 | 725 |
| Shelley Kirk | 10.1016/j.jpeds.2012.01.041 | 2012 | 102 |
| Finkelstein | 10.1016/j.jpeds.2013.01.009 | 2012 | 285 |
| SE Nourse | 10.1016/j.jpeds.2015.06.015 | 2015 | 20 |
| Van Biljon | 10.1016/j.jpeds.2018.07.067 | 2018 | 109 |
| M Harrison | 10.1016/j.jsams.2006.06.012 | 2006 | 293 |

| Trials_1st_author | Trial_DOI | Trial_year | Participants_sample_size |
|---|---|---|---|
| T M Johnson | 10.1016/j.jsams.2015.05.002 | 2016 | 36 |
| D Sheehan | 10.1016/j.jshs.2013.02.002 | 2013 | 64 |
| Z Gao | 10.1016/j.jshs.2018.12.001 | 2019 | 65 |
| G P Nassis | 10.1016/j.metabol.2005.05.013 | 2005 | 19 |
| Aaron S. Kelly | 10.1016/j.metabol.2007.03.009 | 2007 | 19 |
| D A Caranti | 10.1016/j.metabol.2007.05.004 | 2007 | 83 |
| K C Shih | 10.1016/j.metabol.2009.06.035 | 2010 | 106 |
| Whyte | 10.1016/j.metabol.2010.01.002 | 2010 | 27 |
| Bluher | 10.1016/j.metabol.2016.11.015 | 2017 | 20 |
| M Laguna | 10.1016/j.numecd.2012.10.002 | 2013 | 437 |
| M Siergrist | 10.1016/j.nut.2012.12.011 | 2013 | 402 |
| M Yackobovitch Gavan | 10.1016/j.nutres.2009.05.007 | 2009 | 162 |
| E Moslehi | 10.1016/j.obmed.2019.100104 | 2019 | 30 |
| YS Danielsen | 10.1016/j.orcp.2012.06.003 | 2011 | 49 |
| CB Ingul | 10.1016/j.pcad.2018.01.012 | 2018 | 99 |
| A C Seabra | 10.1016/j.pcad.2020.07.007 | 2020 | 40 |
| Haerens L | 10.1016/j.pec.2007.01.003 | 2007 | 281 |
| Haerens L | 10.1016/j.pec.2009.03.020 | 2009 | 1171 |
| B M Melnyk | 10.1016/j.pedhc.2007.02.009 | 2007 | 23 |
| J L Chen | 10.1016/j.pedn.2015.01.026 | 2015 | 70 |
| Schroeder K | 10.1016/j.pedn.2017.07.004 | 2017 | 521 |
| A Bruñó | 10.1016/j.pedn.2018.04.008 | 2018 | 52 |
| Dâmaso | 10.1016/j.peptídeos.2011.04.025 | 2011 | 86 |
| De Carvalho-Ferreira | 10.1016/j.peptídeos.2014.11.010 | 2015 | 75 |
| Carnier | 10.1016/j.physbeh.2011.08.014 | 2012 | 83 |
| D Thivel | 10.1016/j.physbeh.2014.06.013 | 2014 | 26 |
| GS Goldfield | 10.1016/j.physbeh.2018.04.026 | 2018 | 228 |
| MR Da Silva | 10.1016/j.physbeh.2019.112728 | 2020 | 50 |
| L M Barnett | 10.1016/j.pmedr.2015.08.007 | 2015 | 95 |
| S. A. Costigan | 10.1016/j.pmedr.2015.11.001 | 2015 | 65 |
| Burkart S | 10.1016/j.pmedr.2017.08.002 | 2017 | 76 |
| Alhassan S | 10.1016/j.pmedr.2018.05.009 | 2018 | 32 |
| D R Lubans | 10.1016/j.psychsport.2010.06.009 | 2010 | 108 |
| Lucas | 10.1016/j.puhe.2018.02.014 | 2018 | 232 |
| E M D'Agostino | 10.1016/j.puhe.2018.02.025 | 2018 | 1546 |
| DN Ardoy | 10.1016/j.recesp.2011.01.009 | 2011 | 67 |

| Trials_1st_author | Trial_DOI | Trial_year | Participants_sample_size |
|---|---|---|---|
| Elloumi M | 10.1016/j.scispo.2007.09.001 | 2007 | 20 |
| Zehsaz | 10.1016/j.scispo.2016.07.007 | 2017 | 32 |
| A Hamila | 10.1016/j.scispo.2018.03.076 | 2018 | 31 |
| D Alonso-Fernández | 10.1016/j.scispo.2019.04.001 | 2019 | 26 |
| De la CruzMunoz | 10.1016/j.soard.2010.06.004 | 2010 | 78 |
| Lohman | 10.1016/j.ypmed.2003.08.004 | 2003 | 1367 |
| Jago R | 10.1016/j.ypmed.2005.12.010 | 2006 | 473 |
| K Reed | 10.1016/j.ypmed.2008.02.020 | 2008 | 237 |
| McManus AM | 10.1016/j.ypmed.2008.06 | 2008 | 132 |
| McManus | 10.1016/j.ypmed.2008.06.001 | 2008 | 210 |
| Lubans DR | 10.1016/j.ypmed.2009. 12.003. | 2010 | 67 |
| LR Peralta | 10.1016/j.ypmed.2009.04.007 | 2009 | 32 |
| JE Donnelly | 10.1016/j.ypmed.2009.07.022 | 2009 | 1490 |
| SP Schinke | 10.1016/j.ypmed.2009.08.001 | 2099 | 916 |
| Lubans | 10.1016/j.ypmed.2009.12.003 | 2010 | 108 |
| Leanne M Mauriello | 10.1016/j.ypmed.2010.08.004 | 2010 | 1800 |
| David R Lubans | 10.1016/j.ypmed.2011.01.009 | 2011 | 100 |
| N Eather | 10.1016/j.ypmed.2012.10.019 | 2013 | 213 |
| M Heidemann | 10.1016/j.ypmed.2013.04.015 | 2013 | 717 |
| A Lana | 10.1016/j.ypmed.2013.11.015 | 2014 | 2001 |
| Davis | 10.1016/j.ypmed.2016.05.018 | 2016 | 1898 |
| MJ Babic | 10.1016/j.ypmed.2016.07.014 | 2016 | 322 |
| C Braet | 10.1016/S0005-7894(00)80004-0 | 2000 | 109 |
| Kirschenbaum | 10.1016/S0005-7894(84)80051-9 | 1984 | 40 |
| L H Epstein | 10.1016/S0005-7894(85)80002-2 | 1985 | 41 |
| Israel | 10.1016/S0005-7894(85)80043-5 | 1985 | 33 |
| L H Epstein | 10.1016/S0005-7894(85)80046-0 | 1985 | 19 |
| Epstein | 10.1016/S0005-7894(87)80016-3 | 1987 | 41 |
| HA McKay | 10.1016/S0022-3476(00)70095-3 | 2000 | 145 |
| L H Epstein | 10.1016/s0022-3476(85)80506-0 | 1985 | 23 |
| J S Harrell | 10.1016/s0022-3476(96)70332-3 | 1996 | 1274 |
| B Gutin | 10.1016/s0022-3476(97)70280-4 | 1997 | 35 |
| Burke | 10.1016/S0022-3476(98)70315-4 | 1998 | 720 |
| E van Beurden | 10.1016/S0091-7435(02)00044-0 | 2003 | 1.045 |
| D Neumark-Sztainer | 10.1016/S0091-7435(03)00057-4 | 2003 | 201 |
| Sallis JF | 10.1016/s0749-3797(02)00646-3 | 2003 | 24 |

| Trials_1st_author | Trial_DOI | Trial_year | Participants_sample_size |
|---|---|---|---|
| Yan-Ping Li | 10.1016/S0895-3988(10)60050-5 | 2010 | 4700 |
| McMurray RG | 10.1016/s1054-139x(02)00348-8 | 2002 | 1140 |
| L Velázquez-López | 10.1016/S1575-0922(09)73311-X | 2009 | 40 |
| P Lau | 10.1016/S1728-869X(10)60008-1 | 2010 | 18 |
| Elaine Rush | 10.1017/S0007114511003151 | 2012 | 1348 |
| E Rush | 10.1017/S0007114513002316 | 2012 | 926 |
| Camilla T Damsgaard | 10.1017/S0007114514003043 | 2014 | 834 |
| Masquio | 10.1017/S0007114515001129 | 2015 | 69 |
| S M Wilksch | 10.1017/S003329171400289X | 2015 | 1316 |
| Senediak - | 10.1017/S0141347300012027 | 1985 | 45 |
| Graf C | 10.1017/s1047951105000594 | 2005 | 1678 |
| Verstraete | 10.1017/S1368980007223900 | 2006 | 764 |
| R Sichieri | 10.1017/S1368980008002644 | 2009 | 927 |
| Freia De Bock | 10.1017/S136898001100200X | 2012 | 18 |
| Valerie De Coen | 10.1017/S1368980012000687 | 2012 | 1102 |
| Jelalian and Mehlenbeck | 10.1023/A:1014179811950 | 2002 | 16 |
| Winett R.A. | 10.1023%2FA%3A1023233416859 | 1999 | 180 |
| T Gunnarsdottir | 10.1027/1901-2276/a000024 | 2014 | 16 |
| R DeStefano | 10.1034/j.1399-5448.2000.010202.x | 2000 | 15 |
| Epstein | 10.1037//0022-006x.49.5.674 | 1981 | 76 |
| L H Epstein | 10.1037//0022-006x.52.3.429 | 1984 | 53 |
| Epstein | 10.1037//0022-006x.55.1.91 | 1987 | 76 |
| Epstein LH | 10.1037//0278-6133.13.5.373 | 1994 | 36 |
| Epstein | 10.1037/0022-006X.54.3.400 | 1986 | 41 |
| Epstein | 10.1037/0278-6133.23.4.371 | 2004 | 63 |
| CL Davis | 10.1037/a0021766 | 2011 | 161 |
| L Fang | 10.1037/a0030925 | 2013 | 108 |
| H F Byrnes | 10.1037/adb0000442 | 2019 | 411 |
| G S Goldfield | 10.1037/ccp0000038 | 2015 | 304 |
| Amanda S Lochrie | 10.1037/cpp0000020 | 2013 | 130 |
| Shinn | 10.1037/cpp0000204 | 2017 | 43 |
| C D Jensen | 10.1037/cpp0000268 | 2019 | 47 |
| Olive | 10.1037/edu0000338 | 2019 | 821 |
| SR Chen | 10.1037/hea0000390 | 2016 | 50 |
| Olvera N | 10.1038/ oby.2009.439 | 2010 | 31 |
| Singhal, N | 10.1038/ejcn.2009.150 | 2010 | 209 |

| Trials_1st_author | Trial_DOI | Trial_year | Participants_sample_size |
| --- | --- | --- | --- |
| Coppins | 10.1038/ejcn.2011.43 | 2011 | 65 |
| A C Benson | 10.1038/ijo.2008.5 | 2008 | 78 |
| A Sanigorski | 10.1038/ijo.2008.79 | 2008 | 1807 |
| C Simon | 10.1038/ijo.2008.99 | 2008 | 954 |
| C Marcus | 10.1038/ijo.2009.38 | 2009 | 3135 |
| P J Morgan | 10.1038/ijo.2010.151 | 2011 | 71 |
| M Kalavainen | 10.1038/ijo.2011.1 | 2007 | 70 |
| H Crocker | 10.1038/ijo.2011.182 | 2011 | 72 |
| E Makni | 10.1038/ijo.2011.257 | 2012 | 131 |
| B Nguyen | 10.1038/ijo.2012.74 | 2013 | 151 |
| L A Daniels | 10.1038/ijo.2012.96 | 2012 | 698 |
| Alberga | 10.1038/ijo.2015.133 | 2015 | 304 |
| H Zamrazilova | 10.1038/ijo.2015.167 | 2015 | 184 |
| Koot B | 10.1038/ijo.2015.175 | 2016 | 42 |
| J Hay | 10.1038/ijo.2015.241 | 2016 | 106 |
| Hollis | 10.1038/ijo.2016.107 | 2016 | 1150 |
| D C Berry | 10.1038/nutd.2013.42 | 2014 | 358 |
| B Gutin | 10.1038/oby.2000.3 | 2000 | 79 |
| Epstein | 10.1038/oby.2001.103 | 2001 | 89 |
| Saelens | 10.1038/oby.2002.4 | 2002 | 44 |
| J Harvey | 10.1038/oby.2003.87 | 2003 | 37 |
| MA White | 10.1038/oby.2004.132 | 2004 | 57 |
| S Lazzer | 10.1038/oby.2004.30 | 2004 | 26 |
| M Golan | 10.1038/oby.2004.45 | 2004 | 60 |
| Kirschenbaum | 10.1038/oby.2005.187 | 2005 | 83 |
| K Resnicow | 10.1038/oby.2005.212 | 2005 | 147 |
| Yin Z | 10.1038/oby.2005.267 | 2005 | 601 |
| Goran, M.I | 10.1038/oby.2005.86 | 2005 | 209 |
| SA Spiegel | 10.1038/oby.2006.11 | 2006 | 1013 |
| D A Williamson | 10.1038/oby.2006.140 | 2006 | 57 |
| Susan J Rodearmel | 10.1038/oby.2006.158 | 2006 | 105 |
| Fitzgibbon | 10.1038/oby.2006.186 | 2006 | 401 |
| CL Davis | 10.1038/oby.2006.232 | 2006 | 100 |
| L Haerens | 10.1038/oby.2006.98 | 2006 | 294 |
| P Barbeu | 10.1038/oby.2007.247 | 2007 | 201 |
| G Fullerton | 10.1038/oby.2007.306 | 2007 | 86 |

| Trials_1st_author | Trial_DOI | Trial_year | Participants_sample_size |
|---|---|---|---|
| E S Kim | 10.1038/oby.2007.360 | 2007 | 26 |
| J Davis | 10.1038/oby.2009.19 | 2009 | 54 |
| C A Johnston | 10.1038/oby.2009.241 | 2010 | 60 |
| D Foschini | 10.1038/oby.2009.247 | 2010 | 32 |
| G J Van Der Heijden | 10.1038/oby.2009.274 | 2010 | 15 |
| Deanna M Hoelscher | 10.1038/oby.2009.430 | 2010 | 1107 |
| P M Sacher | 10.1038/oby.2009.433 | 2010 | 116 |
| Jayne A. Fulkerson | 10.1038/oby.2009.434 | 2010 | 44 |
| Debra Haire-Joshu | 10.1038/oby.2009.435 | 2010 | 296 |
| J Gittelsohn | 10.1038/oby.2009.436 | 2010 | 117 |
| Olvera | 10.1038/oby.2009.439 | 2010 | 46 |
| PB Balagopal | 10.1038/oby.2009.498 | 2010 | 8 |
| K Boutelle | 10.1038/oby.2010.238. | 2011 | 80 |
| Marian L Fitzgibbon | 10.1038/oby.2010.314 | 2010 | 618 |
| Simone A French | 10.1038/oby.2010.328 | 2011 | 90 |
| L Greening | 10.1038/oby.2010.329 | 2011 | 450 |
| L Greening | 10.1038/oby.2010.329 | 2011 | 450 |
| Hasson | 10.1038/oby.2010.343 | 2012 | 100 |
| Lori J Stark | 10.1038/oby.2010.87 | 2011 | 18 |
| Donald A Williamson | 10.1038/oby.2012.60 | 2012 | 2060 |
| Mary Story | 10.1038/oby.2012.89 | 2012 | 454 |
| C Visuthranukul | 10.1038/pr.2015.142 | 2015 | 70 |
| I Lousa | 10.1038/pr.2018.15 | 2018 | 26 |
| YC Learmonth | 10.1038/s41366-018-0300-1 | 2019 | 1.009 |
| Davis | 10.1038/s41366-019-0482-1 | 2019 | 175 |
| S Vantieghen | 10.1038/s41390-018-0047-3 | 2018 | 48 |
| R G Duft | 10.1038/s41598-020-73943-y | 2020 | 37 |
| Kafatos A | 10.1038/sj.ejcn.1602216 | 2005 | 541 |
| Edwards | 10.1038/sj.ejcn.1602353 | 2006 | 37 |
| Vignolo | 10.1038/sj.ejcn.1602819 | 2007 | 31 |
| MA Ferguson | 10.1038/sj.ijo.0800968 | 2010 | 70 |
| Gately, P | 10.1038/sj.ijo.0801405 | 2000 | 217 |
| Warshburger | 10.1038/sj.ijo.0801708 | 2001 | 197 |
| Dao | 10.1038/sj.ijo.0802535 | 2004 | 55 |
| Dao | 10.1038/sj.ijo.0802542 | 2004 | 55 |
| M F Rolland-Cachera | 10.1038/sj.ijo.0802605 | 2004 | 99 |

| Trials_1st_author | Trial_DOI | Trial_year | Participants_sample_size |
|---|---|---|---|
| J Kain | 10.1038/sj.ijo.0802611 | 2004 | 3086 |
| S Lazzer | 10.1038/sj.ijo.0802845 | 2005 | 27 |
| U Korsten-Reck | 10.1038/sj.ijo.0802875 | 2005 | 496 |
| S Lazzer | 10.1038/sj.ijo.0802977 | 2005 | 26 |
| Jelalian | 10.1038/sj.ijo.0803069 | 2006 | 76 |
| McCallum - | 10.1038/sj.ijo.0803509 | 2007 | 163 |
| M Kalavainen | 10.1038/sj.ijo.0803628 | 2012 | 70 |
| Reinehr | 10.1038/sj.ijo.0803637 | 2007 | 131 |
| V Martínez Vizcaíno | 10.1038/sj.ijo.0803738 | 2008 | 1044 |
| J Salmon | 10.1038/sj.ijo.0803805 | 2008 | 311 |
| Davis | 10.1038%2Foby.2009.19 | 2009 | 54 |
| A E Tjønna | 10.1042/CS20080249 | 2009 | 54 |
| Nova | 10.1046/j.1467-0658.2001.00135.x | 2001 | 186 |
| C Morissey | 10.1055/a-0577-4280 | 2018 | 29 |
| C Morrissey | 10.1055/a-0639-0726 | 2018 | 29 |
| A Keller | 10.1055/s-0028-1105883 | 2006 | 183 |
| D M Prado | 10.1055/s-0029-1233486 | 2009 | 38 |
| DM Prado | 10.1055/s-0030-1267158 | 2010 | 33 |
| C Ackel-D'Elia | 10.1055/s-0033-1345128 | 2014 | 72 |
| F Vasconcelos | 10.1055/s-0034-1398654 | 2015 | 25 |
| Eliakim A | 10.1055/s-2000-3779 | 2000 | 38 |
| G Baquet | 10.1055/s-2001-14343 | 2001 | 551 |
| A M McManus | 10.1055/s-2005-837438 | 2004 | 25 |
| FM Impellizzeri | 10.1055/s-2005-865839 | 2006 | 29 |
| HEALTHY Study Gp 2010 | 10.1056/NEJMoa1001933 | 2010 | 2267 |
| Nichols | 10.1067/mpd.2001.116698 | 2001 | 67 |
| Barbeau P | 10.1067/mpd.2002.127497 | 2002 | 74 |
| Zask | 10.1071/he12010 | 2019 | 560 |
| Manios | 10.1079/BJN2002672 | 2002 | 1046 |
| Golan M | 10.1079/bjn20061757 | 2006 | 37 |
| Stratton G | 10.1080/001401300750003961 | 2000 | 47 |
| Campaigne BN | 10.1080/00913847.1985.11708949 | 1985 | 40 |
| M Kargarfard | 10.1080/00913847.2016.1200442 | 2016 | 20 |
| Farzad Zehsaz | 10.1080/00913847.2016.1248223 | 2016 | 32 |
| Winter | 10.1080/02568543.2011.580211 | 2011 | 405 |
| Ingle | 10.1080/02640410500457117 | 2006 | 47 |

| Trials_1st_author | Trial_DOI | Trial_year | Participants_sample_size |
|---|---|---|---|
| Graf | 10.1080/02640410801930176 | 2008 | 615 |
| P R Hansen | 10.1080/02640414.2013.792951 | 2013 | 20 |
| Dâmaso | 10.1080/02640414.2014.900692 | 2014 | 116 |
| Dâmaso | 10.1080/02640414.2014.900692 | 2014 | 139 |
| J Jarani | 10.1080/02640414.2015.1031161 | 2016 | 760 |
| D Lambrick | 10.1080/02640414.2015.1048521 | 2015 | 55 |
| Fabrício Vasconcellos | 10.1080/02640414.2015.1064150 | 2016 | 20 |
| JE Nunes | 10.1080/02640414.2015.1080384 | 2015 | 25 |
| A C Seabra | 10.1080/02640414.2016.1140219 | 2016 | 88 |
| Wendell Arthur Lopes | 10.1080/02640414.2016.1142107 | 2016 | 33 |
| Birnbaum | 10.1080/02640414.2016.1166390 | 2017 | 1293 |
| S Crouter | 10.1080/02640414.2016.1209305 | 2016 | 30 |
| MC Gallota | 10.1080/02640414.2016.1225969 | 2017 | 230 |
| S. A. Costigan | 10.1080/02640414.2017.1356026 | 2018 | 65 |
| Brand | 10.1080/02640414.2020.1725384 | 2020 | 35 |
| Brasil | 10.1080/02640414.2020.1792189 | 2020 | 35 |
| Ignico | 10.1080/02701367.1995.10607659 | 1997 | 28 |
| Rimmer | 10.1080/02701367.1997.10608868 | 1997 | 25 |
| Flanagan | 10.1080/02701367.2002.10609029 | 2002 | 58 |
| Faigenbaum | 10.1080/02701367.2002.10609041 | 2002 | 55 |
| Hopper, C.A | 10.1080/02701367.2005.10599275 | 2005 | 238 |
| Dos Santos cunha | 10.1080/02701367.2014.982782 | 2015 | 18 |
| M A Perez-Sousa | 10.1080/02701367.2019.1645939 | 2020 | 170 |
| Vasconcellos | 10.1080/02701367.2019.1711007 | 2021 | 13 |
| Roya Kelishadi | 10.1080/07315724.2009.10719792 | 2009 | 120 |
| Serbescu | 10.1080/08035250600599719. | 2006 | 370 |
| Stergioulas A | 10.1080/08035259850156986 | 1998 | 28 |
| Araujo-Soares V | 10.1080/08870440802040707 | 2009 | 291 |
| Silva | 10.1080/09603123.2019.1612041 | 2020 | 33 |
| Son | 10.1080/10641963.2017.1288742 | 2017 | 40 |
| Won-Mok Son | 10.1080/10641963.2017.1288742 | 2017 | 40 |
| McKinney | 10.1080/10852352.2014.881180 | 2014 | 159 |
| Vardar | 10.1080/13813455.2017.1369998 | 2018 | 12 |
| Brown | 10.1080/14635240.2016.1157511 | 2016 | 3290 |
| K Togashi | 10.1080/15438620903423924 | 2010 | 33 |
| Tian | 10.1080/17408989.2015.1072509 | 2007 | 110 |

| Trials_1st_author | Trial_DOI | Trial_year | Participants_sample_size |
|---|---|---|---|
| VP Lopes | 10.1080/17408989.2017.1341474 | 2017 | 60 |
| F Lucertini | 10.1080/17461391.2012.746732 | 20136 | 101 |
| Bendiksen | 10.1080/17461391.2014.884168 | 2014 | 59 |
| P W C Lau | 10.1080/17461391.2014.933880 | 2015 | 48 |
| CA Johnston | 10.1080/17477160701305864 | 2007 | 71 |
| Nowicka | 10.1080/17477160701379810 | 2007 | 44 |
| Alhassan | 10.1080/17477160701520108 | 2017 | 32 |
| Bernard Gutin | 10.1080/17477160801896457 | 2008 | 206 |
| Spruijt-Metz | 10.1080/17477160802113415 | 2008 | 459 |
| L Van Vlierberghe | 10.1080/17477160802220533 | 2009 | 66 |
| R K Evans | 10.1080/17477160802314997 | 2009 | 168 |
| Palmer S | 10.1080/19325037.2005.10608164 | 2005 | 223 |
| Aperman‐Itzhak | 10.1080/19325037.2018.1486755 | 2018 | 396 |
| M S Sothern | 10.1080/713794562 | 2000 | 56 |
| Sothern | 10.1080/713794562 | 2000 | 56 |
| Davis C. L. | 10.1080%2F02701367.2007.10599450 | 2007 | 94 |
| Nidhi, R | 10.1089/acm.2011.0868 | 2013 | 72 |
| Yli-Piipari | 10.1089/acm.2017.0130 | 2018 | 22 |
| Slusser | 10.1089/chi.2011.0060 | 2012 | 160 |
| Z Yin | 10.1089/chi.2011.0085 | 2012 | 574 |
| B Adam Dennis | 10.1089/chi.2011.0092 | 2013 | 112 |
| K Wright | 10.1089/chi.2012.0045 | 2012 | 251 |
| Bender | 10.1089/chi.2012.0118 | 2013 | 43 |
| Yin | 10.1089/chi.2012.0125. | 2012 | 423 |
| M Ramon-Krauel | 10.1089/chi.2013.0022 | 2013 | 17 |
| Davis | 10.1089/chi.2013.0053 | 2013 | 210 |
| D Steven | 10.1089/chi.2013.0107 | 2014 | 72 |
| L Epstein | 10.1089/chi.2013.0123 | 2014 | 101 |
| K Love-Osborne | 10.1089/chi.2013.0165 | 2014 | 82 |
| KN Doughty | 10.1089/chi.2014.0004 | 2015 | 12 |
| Parra-Medina | 10.1089/chi.2014.0120 | 2015 | 118 |
| Madsen | 10.1089/chi.2015.0002 | 2015 | 879 |
| B Saelens | 10.1089/chi.2016.0233 | 2017 | 29 |
| Tripicchio | 10.1089/chi.2017.0021 | 2017 | 64 |
| N Chuensiri | 10.1089/chi.2017.0024 | 2018 | 48 |
| S Armstrong | 10.1089/chi.2017.0089 | 2018 | 101 |

| Trials_1st_author | Trial_DOI | Trial_year | Participants_sample_size |
|---|---|---|---|
| Sharma | 10.1089/chi.2018.0010 | 2019 | 672 |
| AE Maloney | 10.1089/g4h.2011.0009 | 2012 | 64 |
| A Christison | 10.1089/g4h.2015.0097 | 2016 | 84 |
| E P Wickham | 10.1089/met.2008.0038 | 2009 | 165 |
| Hobkrik | 10.1089/met.2011.0050 | 2016 | 75 |
| Mo-Suwan L | 10.1093/ajcn/68.5.1006 | 1998 | 292 |
| Ferguson | 10.1093/ajcn/69.6.1130 | 1999 | 43 |
| B Gutin | 10.1093/ajcn/75.5.818 | 2002 | 80 |
| Benjamin Caballero | 10.1093/ajcn/78.5.1030 | 2003 | 1704 |
| Reinehr | 10.1093/ajcn/84.3.490 | 2006 | 240 |
| M Tsiros | 10.1093/ajcn/87.5.1134 | 2008 | 47 |
| M J Aguilar-Cordero | 10.1093/ajh/hpz174 | 2020 | 98 |
| Nabkasorn | 10.1093/eurpub/cki159 | 2006 | 59 |
| Angelopoulos | 10.1093/eurpub/ckp004 | 2009 | 646 |
| Leeuwen | 10.1093/fampra/cmy061 | 2018 | 154 |
| J M Warren | 10.1093/heapro/dag402 | 2003 | 213 |
| Salmon | 10.1093/heapro/dah502 | 2008 | 293 |
| Leen Haerens | 10.1093/her/cyl115 | 2006 | 2840 |
| Kristjan Thor Magnusson | 10.1093/her/cys049 | 2012 | 321 |
| Israel | 10.1093/jpepsy/19.6.737 | 1994 | 36 |
| C Braet | 10.1093/jpepsy/jsh054 | 2004 | 122 |
| Petty | 10.1093/jpepsy/jsp007 | 2009 | 207 |
| Goldfield GS | 10.1093/jpepsy/jss084 | 2012 | 26 |
| A M Davis | 10.1093/jpepsy/jst005 | 2013 | 58 |
| Olvera | 10.1093/jpepsy/jst041 | 2013 | 99 |
| B Saelens | 10.1093/jpepsy/jst054 | 2013 | 89 |
| Kerri N Boutelle | 10.1093/jpepsy/jst142 | 2014 | 44 |
| Lori J Stark | 10.1093/jpepsy/jsu059 | 2014 | 33 |
| U Njardvik | 10.1093/jpepsy/jsy055 | 2018 | 90 |
| Jyu-Lin Chen | 10.1093/pubmed/fdp105 | 2010 | 67 |
| CF Williams | 10.1093/tbm/ibz015 | 2019 | 175 |
| Chen | 10.1093%2Fpubmed%2Ffdp105 | 2010 | 67 |
| J Dove | 10.1096/fasebj.22.2_supplement.793 | 2008 | 42 |
| J Wong | 10.1096/fasebj.29.1_supplement.747.18 | 2015 | 38 |
| M S Treuth | 10.1097/00005768-199807000-00017 | 1998 | 11 |
| M S Treuth | 10.1097/00005768-199812000-00013 | 1998 | 20 |

| Trials_1st_author | Trial_DOI | Trial_year | Participants_sample_size |
|---|---|---|---|
| S Owens | 10.1097/00005768-199901000-00022 | 1999 | 74 |
| Humphries | 10.1097/00005768-200209000-00005 | 2002 | 78 |
| Kang H | 10.1097/00005768-200212000-00010 | 2002 | 80 |
| Lorraine B Robbins | 10.1097/00006199-200605000-00007 | 2006 | 77 |
| Roth | 10.1097/00006842-198707000-00004 | 1987 | 2441 |
| Tock | 10.1097/01.meg.0000243872.86949.95 | 2006 | 73 |
| T Reinehr | 10.1097/01.mpg.0000235752.29735.31 | 2006 | 119 |
| Harrell, T.K | 10.1097/01.smj.0000182499.59715.07 | 2005 | 205 |
| Graziano | 10.1097/DBP.0000000000000499 | 2017 | 16 |
| Paul Branscum | 10.1097/FCH.0b013e31826d7607 | 2013 | 71 |
| Kilanowsk | 10.1097/FCH.0b013e31829d277e | 2013 | 59 |
| De Piano | 10.1097/MEG.0b013e3283346df2 | 2010 | 50 |
| A de Piano | 10.1097/MEG.0b013e32835793ac | 2012 | 58 |
| Wong | 10.1097/MPG.0000000000001088 | 2016 | 877 |
| H Gronbaek | 10.1097/MPG.0b013e31822cdedf | 2012 | 117 |
| Klish | 10.1097/MPG.0b013e3182318b39 | 2012 | 1714 |
| K Casazza | 10.1097/MPG.0b013e31823df207 | 2012 | 26 |
| De Vries | 10.1111/apa.12880 | 2015 | 161 |
| P Danielsson | 10.1111/apa.13360 | 2016 | 589 |
| Donath | 10.1111/cch.12232 | 2015 | 41 |
| Latorre-Román | 10.1111/cch.12550 | 2018 | 111 |
| A Fleischman | 10.1111/cob.12166 | 2016 | 40 |
| N Farpour-Lambert | 10.1111/cob.12335 | 2019 | 74 |
| D C L Masquio | 10.1111/ijcp.12573 | 2014 | 60 |
| Norman | 10.1111/ijpo.12013 | 2016 | 106 |
| Gatto | 10.1111/ijpo.12102 | 2017 | 319 |
| AE Staiano | 10.1111/ijpo.12117 | 2017 | 38 |
| Sadeghi | 10.1111/ijpo.12135 | 2019 | 782 |
| Hull | 10.1111/ijpo.12197 | 2018 | 272 |
| C Mameli | 10.1111/ijpo.12201 | 2018 | 30 |
| A E Staiano | 10.1111/ijpo.12438 | 2018 | 46 |
| A P Vidmar | 10.1111/ijpo.12464 | 2018 | 35 |
| Lopez-Alarcon | 10.1111/ijpo.12499 | 2019 | 366 |
| N Sherwood | 10.1111/ijpo.12523 | 2018 | 421 |

| Trials_1st_author | Trial_DOI | Trial_year | Participants_sample_size |
|---|---|---|---|
| Brito | 10.1111/ijpo.240 | 2014 | 15 |
| Mendelson | 10.1111/ijpo.255 | 2015 | 20 |
| Fraser | 10.1111/j.1099-0860.2010.00329.x | 2010 | 325 |
| Hills AP, Parker AW | 10.1111/j.1365-2214.1988.tb00592.x | 1988 | 20 |
| Jiang | 10.1111/j.1365-2214.2007.00738.x | 2007 | 2452 |
| Huang | 10.1111/j.1365-2214.2010.01173.x | 2011 | 85 |
| TM O'Connor | 10.1111/j.1365-2214.2011.01344.x | 2013 | 40 |
| H J Kim | 10.1111/j.1365-2265.2007.03058.x | 2008 | 17 |
| Kelishadi R | 10.1111/j.1365-2265.2008.03220.x | 2008 | 100 |
| Sacher | 10.1111/j.1365-277x.2004.00578.x | 2005 | 11 |
| Jiang J | 10.1111/j.1365–2214.2007.00738.x. | 2007 | 2425 |
| McCallum - | 10.1111/j.1440-1754.2005.00689.x | 2005 | 163 |
| Shelton | 10.1111/j.1440-1754.2007.01150.x | 2007 | 43 |
| Torigoe | 10.1111/j.1442-200x.1997.tb03551.x | 1997 | 23 |
| C Chang | 10.1111/j.1467-789X.2007.00455.x | 2008 | 49 |
| AS Singh | 10.1111/j.1467-789X.2008.00475.x | 2009 | 520 |
| Duffy | 10.1111/j.1469-7610.1993.tb01107.x | 1993 | 27 |
| AC Sollerhed | 10.1111/j.1600-0838.2007.00636.x | 2008 | 132 |
| Gallota M C | 10.1111/j.1600-0838.2009.01009.x | 2011 | 44 |
| Resaland | 10.1111/j.1600-0838.2009.01028.x | 2011 | 256 |
| O Faude | 10.1111/j.1600-0838.2009.01087.x | 2010 | 22 |
| D. S. Buchan | 10.1111/j.1600-0838.2011.01303.x | 2011 | 57 |
| M Siegrist | 10.1111/j.1600-0838.2011.01387.x | 2011 | 724 |
| I Ericsson | 10.1111/j.1600-0838.2012.01458.x | 2014 | 220 |
| Reybrouck | 10.1111/j.1651-2227.1990.tb11336.x | 1990 | 14 |
| Danielzik S | 10.1111/j.1651-2227.2007.00165.x | 2007 | 344 |
| N Lazaar | 10.1111/j.1651-2227.2007.00426.x | 2007 | 425 |
| Hagstromer | 10.1111/j.1651-2227.2008.01116.x | 2009 | 31 |
| Elloumi | 10.1111/j.1651-2227.2009.01365.x | 2009 | 21 |
| Ben Ounis | 10.1111/j.1651-2227.2010.01920.x | 2010 | 32 |
| BK McFarlin | 10.1111/j.1740-8709.2011.00398.x | 2013 | 221 |
| A Satoh | 10.1111/j.1742-7924.2007.00077.x | 2007 | 81 |
| Speroni | 10.1111/j.1744-6155.2008.00149.x | 2008 | 32 |
| Grey, M | 10.1111/j.1746-1561.2004.tb06595.x | 2004 | 41 |
| Katz, D.L | 10.1111/j.1746-1561.2010.00553.x | 2011 | 1180 |
| Herrick | 10.1111/j.1746-1561.2012.00722.x | 2012 | 100 |

| Trials_1st_author | Trial_DOI | Trial_year | Participants_sample_size |
|---|---|---|---|
| J F Sallis | 10.1111/j.1749-6632.1993.tb18844.x | 1993 | 549 |
| M T de Mello | 10.1111/j.1751-7176.2010.00388.x | 2011 | 30 |
| B K Weeks | 10.1111/j.2047-6310.2011.00026.x | 2012 | 99 |
| SM Willi | 10.1111/j.2047-6310.2011.00042.x | 2012 | 4363 |
| J de Niet | 10.1111/j.2047-6310.2012.00048.x | 2012 | 141 |
| TL Wagener | 10.1111/j.2047-6310.2012.00065.x | 2012 | 41 |
| S Marild | 10.1111/j.2047-6310.2012.00105.x | 2013 | 265 |
| B Q Farah | 10.1111/j.2047-6310.2012.00145.x | 2013 | 43 |
| JP Elder | 10.1111/j.2047-6310.2013.00164.x | 2013 | 541 |
| McCormack S | 10.1111/j.2047-6310.2013.00180.x | 2014 | 18 |
| L. Elizondo-Montemayor | 10.1111/jhn.12070 | 2013 | 96 |
| J Chen | 10.1111/jhn.12446 | 2017 | 36 |
| E M Y Wong | 10.1111/jocn.12098 | 2013 | 185 |
| Schuna | 10.1111/josh.12005 | 2013 | 116 |
| Craig A Johnston | 10.1111/josh.12013 | 2013 | 835 |
| R Sacchetti | 10.1111/josh.12076 | 2013 | 438 |
| SE Erfle | 10.1111/josh.12217 | 2015 | 10.206 |
| Parsons | 10.1111/jspn.12235 | 2019 | 11 |
| Mohammed | 10.1111/pedi.13203 | 2021 | 40 |
| Thompson W M | 10.1111/phn.12009 | 2013 | 41 |
| Kilanowsk | 10.1111/phn.12175 | 2015 | 138 |
| L B Christiansen | 10.1111/sms.12088 | 2013 | 1348 |
| A C Seabra | 10.1111/sms.12268 | 2014 | 20 |
| Krustrup | 10.1111/sms.12277 | 2014 | 97 |
| CT Rexen | 10.1111/sms.12293 | 2015 | 1.247 |
| N Cvetković | 10.1111/sms.13241 | 2018 | 42 |
| Hacke | 10.1111/sms.13390 | 2019 | 135 |
| H Zhang | 10.1111/sms.13803 | 2021 | 59 |
| Martinez-Viscaiano | 10.1111/sms.14113 | 2022 | 562 |
| Gallotta, M. | 10.1111/sms.2011.21.issue-2 | 2011 | 21 |
| Geria | 10.1111/telefone.12379 | 2018 | 101 |
| J Marshal | 10.1123/apaq.14.3.222 | 1997 | 406 |
| K J Lee | 10.1123/ijsnem.20.4.275 | 2010 | 18 |
| JA Reed | 10.1123/jpah.10.2.185 | 2013 | 470 |
| HJG Silva | 10.1123/jpah.2013-0199 | 2016 | 132 |
| M Schmidt | 10.1123/jsep.2015-0069 | 2015 | 181 |

| Trials_1st_author | Trial_DOI | Trial_year | Participants_sample_size |
|---|---|---|---|
| Siegel | 10.1123/pes.1.2.145 | 1989 | 50 |
| Faigenbaum | 10.1123/pes.17.3.237 | 2005 | 43 |
| Kaufman C | 10.1123/pes.19.1.82 | 2007 | 20 |
| Heyman E | 10.1123/pes.19.4.408 | 2007 | 16 |
| Henaghan J | 10.1123/pes.20.2.169 | 2008 | 46 |
| H Youssef | 10.1123/pes.2014-0008 | 2015 | 39 |
| W L Prado | 10.1123/pes.2015-0018 | 2015 | 43 |
| Horner | 10.1123/pes.2015-0067 | 2015 | 81 |
| RA Jones | 10.1123/pes.2015-0116 | 2015 | 37 |
| P Farinatti | 10.1123/pes.2015-0191 | 2016 | 44 |
| O Rey | 10.1123/pes.2016-0105 | 2017 | 24 |
| Tan | 10.1123/pes.2016-0107 | 2017 | 104 |
| A Wong | 10.1123/pes.2017-0198 | 2018 | 30 |
| Angus A. Leahy | 10.1123/pes.2018-0039 | 2019 | 68 |
| Martin-Smith | 10.1123/pes.2018-0155 | 2017 | 56 |
| A Paahoo | 10.1123/pes.2020-0138 | 2021 | 30 |
| S Tan | 10.1123/pes.22.3.477 | 2010 | 60 |
| Faigenbaum | 10.1123/pes.23.4.573 | 2011 | 40 |
| Jones | 10.1123/pes.23.4.600 | 2011 | 97 |
| Faigenbaum | 10.1123/pes.5.4.339 | 1993 | 24 |
| R Shephard | 10.1123/pes.6.1.75 | 1994 | 546 |
| Blessing DL | 10.1123/pes.7.2.192 | 1995 | 25 |
| JX Jiang | 10.1136/adc.2005.071753 | 2005 | 68 |
| R R Kipping | 10.1136/adc.2007.116970 | 2008 | 679 |
| Robertson | 10.1136/adc.2008.139162 | 2008 | 27 |
| Reinehr T | 10.1136/adc.2008.143594 | 2009 | 152 |
| A L Ford | 10.1136/adc.2009.165340 | 2010 | 106 |
| Schwingshandl J | 10.1136/adc.81.5.426 | 1999 | 30 |
| R Sung | 10.1136/adc.86.6.407 | 2002 | 82 |
| A Kokkvoll | 10.1136/archdischild-2014-307107 | 2015 | 97 |
| Bhave | 10.1136/archdischild-2015-308673 | 2016 | 491 |
| B Benestad | 10.1136/archdischild-2015-309813 | 2017 | 94 |
| W Robertson | 10.1136/archdischild-2016-311514 | 2017 | 105 |
| P Obert | 10.1136/bjsm.2007.039941 | 2009 | 50 |
| LB Andersen | 10.1136/bjsports-2011-090333 | 2015 | 39 |
| N Schranz | 10.1136/bjsports-2013-092209 | 2014 | 56 |

| Trials_1st_author | Trial_DOI | Trial_year | Participants_sample_size |
|---|---|---|---|
| May Grydeland | 10.1136/bjsports-2013-092284 | 2014 | 1324 |
| Martínez-Vizcaíno | 10.1136/bjsports-2018-099655 | 2018 | 741 |
| HS Hansen | 10.1136/bmj.303.6804.682 | 1991 | 132 |
| P Sahota | 10.1136/bmj.323.7320.1027 | 2001 | 634 |
| Sahota, P | 10.1136/bmj.323.7320.1029 | 2001 | 613 |
| James | 10.1136/bmj.38077.458438.ee | 2004 | 644 |
| John J Reilly | 10.1136/bmj.38979.623773.55 | 2006 | 545 |
| J James | 10.1136/bmj.39342.571806.55 | 2007 | 644 |
| Wake | 10.1136/bmj.b3308 | 2009 | 258 |
| S Kriemler | 10.1136/bmj.c785 | 2010 | 540 |
| J J Puder | 10.1136/bmj.d6195 | 2011 | 652 |
| M Wake | 10.1136/bmj.f3092 | 2013 | 118 |
| RR Kipping | 10.1136/bmj.g3256 | 2014 | 2221 |
| K M Wyatt | 10.1136/bmjopen-2010-000026 | 2011 | 398 |
| Cui | 10.1136/bmjopen-2011-000721 | 2012 | 682 |
| Adab | 10.1136/bmjopen-2013-004579 | 2014 | 574 |
| E Llargues | 10.1136/jech.2009.102319 | 2011 | 704 |
| Rudolf | 10.1136%2Fadc.2005.089896 | 2006 | 94 |
| Rowland D | 10.1136%2Fadc.88.1.8 | 2003 | 21 |
| Welsman JR | 10.1136%2Fbjsm.31.2.139 | 1997 | 96 |
| McNarry | 10.1139/apnm-2015-0051 | 2015 | 26 |
| Tan | 10.1139/apnm-2015-0174 | 2016 | 46 |
| G Racil | 10.1139/apnm-2015-0384 | 2016 | 68 |
| Alberga | 10.1139/apnm-2015-0413 | 2016 | 304 |
| B E Starkoff | 10.1139/apnm-2016-0240 | 2014 | 27 |
| Goldfield | 10.1139/apnm-2016-0386 | 2016 | 304 |
| L Plavsic | 10.1139/apnm-2019-0137 | 2020 | 44 |
| M Miguet | 10.1139/apnm-2019-0160 | 2020 | 43 |
| S Lee | 10.1139/apnm-2019-0993 | 2020 | 60 |
| S A Rasooli | 10.1139/apnm-2020-0171 | 2021 | 40 |
| Stergioulas AT | 10.1139/h06-020 | 2006 | 38 |
| Adamo KB | 10.1139/H10-078 | 2010 | 26 |
| Doyle-Baker | 10.1139/h11-042 | 2011 | 27 |
| Carine Gueugnon | 10.1139/h2012-045 | 2012 | 47 |
| S J Lee | 10.1152/ajpendo.00285.2013 | 2013 | 44 |
| NC Moller | 10.1152/japplphysiol.00316.2010 | 2012 | 12 |

| Trials_1st_author | Trial_DOI | Trial_year | Participants_sample_size |
|---|---|---|---|
| SK Rosenkranz | 10.1152/japplphysiol.00663.2011 | 2012 | 16 |
| Tsang | 10.1155/2010/672751 | 2010 | 20 |
| Cheryl A Howe | 10.1155/2011/358581 | 2011 | 106 |
| Maria Waling | 10.1155/2012/913965 | 2012 | 93 |
| A R Dâmaso | 10.1155/2013/541032 | 2013 | 97 |
| Kong | 10.1155/2013/575016 | 2013 | 60 |
| Juliana Kain | 10.1155/2014/618293 | 2014 | 1949 |
| Keating | 10.1155/2014/834865 | 2014 | 38 |
| Foster | 10.1155/2016/2609504 | 2016 | 60 |
| Kong | 10.1155/2016/4073618 | 2016 | 10 |
| Towey | 10.1155%2F2011%2F619643 | 2011 | 272 |
| T G Park | 10.1159/000104137 | 2007 | 44 |
| Munsch S | 10.1159/000129659 | 2008 | 56 |
| Carnier | 10.1159/000139148 | 2018 | 20 |
| S Adam | 10.1159/000234415 | 2009 | 162 |
| S Shalitin | 10.1159/000245931 | 2009 | 162 |
| O B Ounis | 10.1159/000275888 | 2010 | 28 |
| Susanne Brandstetter | 10.1159/000336255 | 2012 | 2154 |
| Da Silva | 10.1159/000345840 | 2012 | 84 |
| Ouerghi | 10.1159/000471882 | 2017 | 18 |
| K S Woo | 10.1161/01.CIR.0000126599.47470.BE | 2004 | 82 |
| Ribeiro | 10.1161/01.CIR.0000161959.04675.5A | 2005 | 39 |
| Rocchini | 10.1161/01.hyp.10.3.267 | 1987 | 50 |
| Walther C | 10.1161/CIRCUL | 2009 | 182 |
| Pate RR | 10.1161/circulationaha.106.177052 | 2006 | 1539 |
| Walther | 10.1161/CIRCULATIONAHA.109.865808 | 2009 | 182 |
| O Saygun | 10.11613/BM.2015.013 | 2011 | 40 |
| Rice | 10.1177/0009922807306168 | 2008 | 68 |
| Kong | 10.1177/0009922810370364 | 2010 | 28 |
| Shari L Barkin | 10.1177/0009922810379039 | 2011 | 72 |
| MK Bean | 10.1177/0009922810393497 | 2011 | 186 |
| Amy Christison | 10.1177/0009922811429480 | 2012 | 48 |
| A Murphy | 10.1177/0009922814528038 | 2015 | 13 |
| Shannon M Looney | 10.1177/0009922814541803 | 2014 | 22 |

| Trials_1st_author | Trial_DOI | Trial_year | Participants_sample_size |
|---|---|---|---|
| E R Wald | 10.1177/0009922817733703 | 2018 | 73 |
| C Pesce | 10.1177/0017896912444176 | 2013 | 125 |
| Coleman | 10.1177/0145721710377360 | 2010 | 62 |
| Shaib | 10.1177/0145721712446635 | 2012 | 15 |
| Blakely Brown | 10.1177/0145721712465343 | 2013 | 64 |
| Camila Tc de Lira | 10.1177/0260106017720350 | 2017 | 107 |
| P H Boer | 10.1177/0269215513498609 | 2013 | 32 |
| Amy Bohnert | 10.1177/0272431612466174 | 2012 | 76 |
| Natale | 10.1177/0890117116661156 | 2017 | 1211 |
| M Ming Hao | 10.1177/1010539519848275 | 2019 | 229 |
| Shore | 10.1177/1054773813485240 | 2014 | 92 |
| Hawthorne | 10.1177/1059840510391669 | 2011 | 1293 |
| Smith | 10.1177/1059840512472708 | 2013 | 160 |
| Kilanowski | 10.1177/1059840513506999 | 2014 | 64 |
| T Boyle-Holmes | 10.1177/1090198109343895 | 2010 | 1.464 |
| Anderson | 10.1177/1090198114547813 | 2015 | 62 |
| M R Stolley | 10.1177/109019819702400204 | 1997 | 55 |
| D Mayorga-Vega | 10.1177/1356336X15599010 | 2016 | 111 |
| M Piéron | 10.1177/1356336X9600200204 | 1996 | 7.721 |
| A M Davis | 10.1177/1357633X15586642 | 2016 | 103 |
| A J Wilson | 10.1177/1359105311434050 | 2012 | 43 |
| P Nowicka | 10.1177/1403494809344444 | 2009 | 76 |
| Karacebey | 10.1177/14732300090370 | 2009 | 40 |
| Kursat Karacabey | 10.1177/147323000903700523 | 2009 | 40 |
| Ruby A Natale | 10.1177/1524839914523429 | 2014 | 307 |
| Cruz | 10.1177/1524839916629974 | 2016 | 655 |
| Hatfield | 10.1177/1524839916642714 | 2017 | 101 |
| Maria Bryant | 10.1177/1740774511424766 | 2011 | 70 |
| Amanda E Staiano | 10.1177/193229681200600412 | 2012 | 31 |
| K Patrick | 10.1177/193229681300700322 | 2013 | 101 |
| DC Wilks | 10.1177/2047487312465691 | 2014 | 429 |
| Shaibi | 10.1177/2150131910377909 | 2010 | 102 |
| Lynch | 10.1177/2150131916644888 | 2016 | 51 |
| Hildemar Dos Santos | 10.1177/2333794X16644139 | 2016 | 55 |
| Josephine Ho | 10.1177/2333794X16669014 | 2016 | 99 |
| ML Kolsgaard | 10.1186/1471-2431-11-47 | 2011 | 230 |

| Trials_1st_author | Trial_DOI | Trial_year | Participants_sample_size |
|---|---|---|---|
| Fisher | 10.1186/1471-2431-11-97 | 2011 | 64 |
| J H Park | 10.1186/1471-2431-12-111 | 2012 | 29 |
| H Klakk | 10.1186/1471-2431-13-170 | 2013 | 632 |
| D Nemet | 10.1186/1471-2431-13-45 | 2013 | 342 |
| L Velazquez | 10.1186/1471-2431-14-175 | 2014 | 49 |
| N M Harder-Lauridsen | 10.1186/1471-2431-14-273 | 2014 | 38 |
| Klijn | 10.1186/1471-2431-7-19 | 2007 | 15 |
| Richard R Rosenkranz | 10.1186/1471-2458-10-81 | 2010 | 76 |
| Teresa Shamah Levy | 10.1186/1471-2458-12-152 | 2010 | 1020 |
| Sigmund | 10.1186/1471-2458-12-570 | 2012 | 176 |
| Duncan S Buchan | 10.1186/1471-2458-13-498 | 2013 | 89 |
| Stuart J Fairclough | 10.1186/1471-2458-13-626 | 2013 | 318 |
| Xiao-Hui Li | 10.1186/1471-2458-14-1282 | 2014 | 921 |
| A P S Kong | 10.1186/1471-2458-14-180 | 2014 | 104 |
| Abbey Alkon | 10.1186/1471-2458-14-215 | 2014 | 689 |
| Carla Habib-Mourad | 10.1186/1471-2458-14-940 | 2014 | 374 |
| A J Daley | 10.1186/1471-2458-5-113 | 2005 | 81 |
| Weigensberg | 10.1186/1472-6882-14-28 | 2014 | 35 |
| J L Rosado | 10.1186/1475-2891-7-28 | 2008 | 147 |
| Alwis et al | 10.1186/1476-5918-7-8 | 2008 | 42 |
| M Grydeland | 10.1186/1479-5868-10-17 | 2013 | 96 |
| L Foley | 10.1186/1479-5868-11-46 | 2014 | 322 |
| Jones | 10.1186/1479-5868-5-42 | 2008 | 606 |
| C Ni Mhurchu | 10.1186/1479-5868-5-8 | 2008 | 20 |
| D A Dzewaltowski | 10.1186/1479-5868-7-90 | 2010 | 273 |
| R Maddison | 10.1186/1479-5868-9-54 | 2012 | 322 |
| K J Coleman | 10.1186/1479-5868-9-80 | 2012 | 827 |
| Jago 26 | 10.1186/1479-5868-9-83 | 2012 | 793 |
| DA Gentille | 10.1186/1741-7015-7-49 | 2009 | 1201 |
| D Christie | 10.1186/1745-6215-12-242 | 2011 | 200 |
| S A Boodai | 10.1186/1745-6215-15-234 | 2014 | 82 |
| Tarro | 10.1186/1745-6215-15-58 | 2014 | 1939 |
| Telles | 10.1186/1753-2000-7-37 | 2014 | 98 |
| Kamal | 10.1186/1758-5996-4-27 | 2012 | 93 |
| Linnea Johansson | 10.1186/s12887-020-02338-9 | 2020 | 28 |
| M Cao | 10.1186/s12887-021-03079-z | 2022 | 36 |

| Trials_1st_author | Trial_DOI | Trial_year | Participants_sample_size |
|---|---|---|---|
| J Wang | 10.1186/s12889-015-2535-2 | 2015 | 90 |
| E Waters | 10.1186/s12889-017-4625-9 | 2018 | 2965 |
| N Ahmad | 10.1186/s12937-018-0379-1 | 2018 | 134 |
| Paula Alves Monteiro | 10.1186/s12944-015-0152-9 | 2015 | 32 |
| MF Smout | 10.1186/s12955-021-01907-5 | 2022 | 100 |
| R Maddison | 10.1186/s12966-014-0111-2 | 2014 | 251 |
| Jago 27 | 10.1186/s12966-014-0114-z | 2014 | 600 |
| Susana Andrade | 10.1186/s12966-014-0153-5 | 2014 | 1440 |
| Martinez-Vizcaino | 10.1186/s12966-014-0154-4 | 2014 | 712 |
| Thompson | 10.1186/s12966-015-0199-z | 2015 | 400 |
| Jago R | 10.1186/s12966-015-0289-y | 2015 | 508 |
| DR Lubans | 10.1186/s12966-016-0420-8 | 2016 | 361 |
| Wylie-Rosett | 10.1186/s12966-017-0639-z | 2018 | 365 |
| GA Ten Hoor | 10.1186/s12966-018-0727-8 | 2018 | 695 |
| R Maddison | 10.1186/s12966-12014-10111-12962 | 2014 | 251 |
| Otterbach | 10.1186/s40608-018-0188-2 | 2018 | 94 |
| Tanas | 10.1186%2F1471-2431-7-33 | 2007 | 190 |
| Monteiro PA | 10.1186%2Fs12944-015-0152-9 | 2015 | 48 |
| Wilson | 10.1207/s15324796abm2404_07 | 2002 | 53 |
| L H Epstein | 10.1207/s15324796abm3003_4 | 2005 | 41 |
| Balagopal | 10.1210/jc.2004-2427 | 2005 | 8 |
| Rosenbaum, M. | 10.1210/jc.2006-1516 | 2007 | 79 |
| L M Bell | 10.1210/jc.2007-0779 | 2007 | 14 |
| Garibay-Nieto | 10.1210/jc.2016-2701 | 2017 | 83 |
| G Q Shaibi | 10.1249/01.mss.0000227304.88406.0f | 2006 | 22 |
| Ahamed | 10.1249/01.mss.0000241654.45500.8e | 2007 | 287 |
| Zwinkels | 10.1249/01.mss.0000518945.27391.2b | 2017 | 68 |
| Shaibi | 10.1249/01.mss.000227304.88406 | 2006 | 22 |
| KE Cohen | 10.1249/MSS.0000000000000452 | 2015 | 460 |
| Sim | 10.1249/MSS.0000000000000687 | 2015 | 10 |
| Kristina Roth | 10.1249/MSS.0000000000000703 | 2015 | 709 |
| Dias | 10.1249/MSS.0000000000000705 | 2015 | 44 |
| Koutsandréou | 10.1249/MSS.0000000000000869 | 2016 | 71 |
| L H Naylor | 10.1249/MSS.0b013e318182a9e0 | 2008 | 23 |

| Trials_1st_author | Trial_DOI | Trial_year | Participants_sample_size |
|---|---|---|---|
| J Davis | 10.1249/MSS.0b013e31819b6aea | 2009 | 41 |
| G J Van Der Heijden | 10.1249/MSS.0b013e3181df16d9 | 2010 | 12 |
| DP Cliff | 10.1249/MSS.0b013e3181e741e8 | 2011 | 402 |
| R Jago | 10.1249/MSS.0b013e31820c9797 | 2011 | 4063 |
| Aburto | 10.1249/mss.0b013e318217ebec. | 2011 | 699 |
| J N Davis | 10.1249/MSS.0b013e31821f5d4e | 2011 | 38 |
| C Rynders | 10.1249/MSS.0b013e31823cef5e | 2014 | 16 |
| A Bugge | 10.1249/MSS.0b013e31824bd579 | 2012 | 696 |
| B Löfgren | 10.1249/MSS.0b013e31827c0889 | 2013 | 232 |
| R D Telford | 10.1249/MSS.0b013e318293b1ee | 2013 | 708 |
| Davis JN | 10.1249%2FMSS.0b013e31821f5d4e | 2012 | 26 |
| Pitson & Wallace | 10.1258/jhsrp.2010.010076 | 2011 | 47 |
| Schiel | 10.1258/jtt.2007.070504 | 2008 | 140 |
| Gerosa-Neto | 10.12965/jer.1632770.385 | 2016 | 32 |
| Ghahramani | 10.13005/bbra/2318 | 2016 | 40 |
| F. Cao | 10.13481/j.1671-587x.2012.03.005 | 2012 | 40 |
| Li | 10.13481/j.1671-587x.20140536 | 2014 | 40 |
| Q. Chen | 10.13481/j.1671-587x.20150537 | 2015 | 30 |
| Danielle Hollar | 10.1353/hpu.0.0304 | 2010 | 3769 |
| Linden C | 10.1359/jbmr.060304 | 2006 | 99 |
| BK Weeks | 10.1359/jbmr.080226 | 2008 | 99 |
| A C Araujo | 10.1371/journal.pone.0042747 | 2012 | 30 |
| D B Cunha | 10.1371/journal.pone.0057498 | 2013 | 574 |
| Meng | 10.1371/journal.pone.0077971 | 2013 | 8301 |
| U Meyer | 10.1371/journal.pone.0087929 | 2014 | 289 |
| Gisela Nyberg | 10.1371/journal.pone.0116876 | 2015 | 243 |
| S Gerards | 10.1371/journal.pone.0122240 | 2015 | 86 |
| A A Abraham | 10.1371/journal.pone.0125673 | 2015 | 48 |
| M Simons | 10.1371/journal.pone.0126023 | 2014 | 260 |
| Fisher | 10.1371/journal.pone.0138853 | 2015 | 28 |
| Xu F | 10.1371/journal.pone.0141421 | 2015 | 1182 |
| Crouter | 10.1371/journal.pone.0141584. | 2015 | 36 |
| N Serra-Paya | 10.1371/journal.pone.0144502 | 2015 | 113 |
| H Truby | 10.1371/journal.pone.0151787 | 2016 | 87 |
| K Traberg | 10.1371/journal.pone.0157182 | 216 | 115 |
| Tarp | 10.1371/journal.pone.0158087 | 2016 | 632 |

| Trials_1st_author | Trial_DOI | Trial_year | Participants_sample_size |
|---|---|---|---|
| Kong | 10.1371/journal.pone.0158589 | 2016 | 31 |
| L Zhen | 10.1371/journal.pone.0235951 | 2010 | 60 |
| YG Seo | 10.1371/journal.pone.0245875 | 2021 | 242 |
| Mu Taiyang | 10.1371/journal.pone.0271845 | 2019 | 2995 |
| Blau, H | 10.1378/chest.121.4.1117 | 2002 | 13 |
| Eirini Bathrellou | 10.14310/horm.2002.1267 | 2010 | 42 |
| I H Park | 10.14352/jkaie.2015.19.1.99 | 2015 | 30 |
| Johnston | 10.14485/HBPR.4.5.5 | 2017 | 198 |
| Hailu K. Alemayehu | 10.14814/phy2.13888 | 2018 | 16 |
| Wissal Abassi | 10.1515/hmbci-2020-0031 | 2020 | 43 |
| Hogg J | 10.1515/jpem-2012-0027 | 2012 | 61 |
| I Partsalaki | 10.1515/jpem-2012-0131 | 2012 | 21 |
| V Calcaterra | 10.1515/jpem-2012-0157 | 2013 | 22 |
| Kelly | 10.1515/jpem-2014-0470 | 2015 | 26 |
| Liu | 10.1515/jpem-2017-0431 | 2018 | 50 |
| J Kim | 10.1515/jpem-2019-0327 | 2020 | 48 |
| B Falk | 10.1515/jpem.2002.15.5.597 | 2002 | 30 |
| Gillis | 10.1515/jpem.2007.20.2.197 | 2007 | 27 |
| Eliakim | 10.1515/jpem.2007.20.6.711 | 2007 | 54 |
| D Nemet | 10.1515/JPEM.2008.21.5.461 | 2008 | 22 |
| Rohrer | 10.1515/JPEM.2008.21.9.837 | 2008 | 22 |
| Hyun-Wook Chae | 10.1515/jpem.2010.168 | 2010 | 38 |
| O T Hardy | 10.1515/jpem.2011.386 | 2012 | 27 |
| G Baquet | 10.1519/13813.1 | 2004 | 100 |
| C C W Yu | 10.1519/14994.1 | 2005 | 82 |
| Faigenbaum | 10.1519/1533-4287(1996)010<0109: TEOSTA>2.3.CO;2 | 1996 | 24 |
| Chaouachi | 10.1519/JSC.0000000000000305 | 2014 | 30 |
| Faigenbaum | 10.1519/JSC.0000000000000812. | 2015 | 41 |
| GG Nobre | 10.1519/JSC.0000000000001684 | 2017 | 59 |
| BHM Branco | 10.1519/JSC.0000000000002877 | 2018 | 18 |
| M R McGuigan | 10.1519/jsc.0b013e3181876a56 | 2009 | 48 |
| M Sgro | 10.1519/JSC.0b013e3181910746 | 2009 | 31 |
| Dorgo | 10.1519/JSC.0b013e3181b8d42a | 2009 | 192 |
| A Velez | 10.1519/JSC.0b013e3181cc230a | 2010 | 28 |
| G Baquet | 10.1519/JSC.0b013e3181d1575a | 2010 | 77 |
| B Sperlich | 10.1519/JSC.0b013e3181d67c38 | 2011 | 19 |

| Trials_1st_author | Trial_DOI | Trial_year | Participants_sample_size |
|---|---|---|---|
| O Sandbakk | 10.1519/JSC.0b013e3182752f08 | 2013 | 15 |
| Shing | 10.1519/JSC.0b013e31827e1644 | 2013 | 7 |
| Campos | 10.1519/JSC.0b013e3182a996df | 2014 | 42 |
| Kotzamanidis | 10.1519/R-16194.1 | 2006 | 30 |
| Lopez-Alarcon | 10.1530/EC-19-0461 | 2020 | 45 |
| Debar | 10.1542/peds. 2011-0863 | 2012 | 208 |
| J S Harrell | 10.1542/peds.102.2.371 | 1998 | 422 |
| Mitchell | 10.1542/peds.109.5.e73 | 2002 | 81 |
| MacKelvie KJ | 10.1542/peds.112.6.e447 | 2003 | 64 |
| D Nemet | 10.1542/peds.2004-2172 | 2005 | 46 |
| CB Ebbeling | 10.1542/peds.2005-0983 | 2006 | 103 |
| Goldfield et al | 10.1542/peds.2005-3052 | 2006 | 30 |
| Daley | 10.1542/peds.2006-1285 | 2006 | 81 |
| Golley | 10.1542/peds.2006-1746 | 2007 | 111 |
| Susan J Rodearmel | 10.1542/peds.2006-2927 | 2007 | 192 |
| S Stock | 10.1542/peds.2006-3003 | 2007 | 383 |
| C A Johnston | 10.1542/peds.2006-3321 | 2007 | 60 |
| Gary D Foster | 10.1542/peds.2007-1365 | 2008 | 1349 |
| Hughes | 10.1542/peds.2007-1786 | 2008 | 134 |
| M A Kalarchian | 10.1542/peds.2008-3727 | 2009 | 192 |
| Maureen M Black | 10.1542/peds.2009-1832 | 2010 | 235 |
| M Savoye | 10.1542/peds.2010-0697 | 2012 | 105 |
| LL DeBar | 10.1542/peds.2011-0863 | 2012 | 208 |
| Lloyd-Richardson | 10.1542/peds.2011-3283 | 2012 | 118 |
| Barkin | 10.1542/peds.2011-3762 | 2012 | 106 |
| Teresa Quattrin | 10.1542/peds.2012-0701 | 2012 | 105 |
| A M Davoli | 10.1542/peds.2013-1738 | 2013 | 372 |
| Naiman A Khan | 10.1542/peds.2013-2246 | 2014 | 220 |
| Vander Ploeg | 10.1542/peds.2013-2383 | 2008 | 1157 |
| Dzewaltowski | 10.1542/peds.2013-2621 | 2014 | 2211 |
| Hillman | 10.1542/peds.2013-3219 | 2014 | 296 |
| Jordan J Smith | 10.1542/peds.2014-1012 | 2014 | 361 |
| L Bruyndonckx | 10.1542/peds.2014-1577 | 2015 | 61 |
| A H Pakpour | 10.1542/peds.2014-1987 | 2015 | 357 |
| RW Taylor | 10.1542/peds.2015-0595 | 2015 | 206 |
| Bogart | 10.1542/peds.2015-2493 | 2016 | 2439 |

| Trials_1st_author | Trial_DOI | Trial_year | Participants_sample_size |
|---|---|---|---|
| I Vajda | 10.1556/APhysiol.94.2007.3.4 | 2007 | 21 |
| VA Kovacs | 10.1556/APhysiol.96.2009.3.7 | 2009 | 51 |
| Al Saif | 10.1589/jpts.27.1697 | 2015 | 40 |
| Lee | 10.1589/jpts.27.1903 | 2015 | 30 |
| S S Lee | 10.1589/jpts.27.3063 | 2015 | 20 |
| ASR Alves | 10.1590/0001-3765201920181264 | 2019 | 40 |
| B Antunes | 10.1590/2359-3997000000095 | 2015 | 25 |
| GE Milano | 10.1590/S0004-27302013000700006 | 2013 | 75 |
| Stella | 10.1590/S0100-879X2005001100017 | 2005 | 40 |
| Silva | 10.1590/S0101-32892014000200002 | 2014 | 36 |
| JGB Alves | 10.1590/s0102-311x2008001400020 | 2008 | 78 |
| B Antunes | 10.1590/S0103-05822013000300015 | 2013 | 34 |
| Leite | 10.1590/S1413-35552009005000009 | 2009 | 64 |
| A R Dâmaso | 10.1590/S1517-86922006000500008 | 2006 | 28 |
| MR Chehuen | 10.1590/S1517-86922011000400003 | 2012 | 205 |
| Saeid Fazelifar | 10.1590/s1517-86922013000500010 | 2013 | 24 |
| Martinez | 10.1590/S1980-6574201600040022 | 2016 | 94 |
| T Cohen | 10.17269/CJPH.107.5470 | 2016 | 78 |
| Muth | 10.18043/ncm.69.6.432 | 2008 | 75 |
| Berry | 10.1891/1540-4153.9.4.186 | 2011 | 56 |
| Russell R Pate | 10.2105/AJPH.2004.045807 | 2005 | 2744 |
| De Heer | 10.2105/ajph.2011.300177 | 2011 | 804 |
| Hendrik D. de Heer | 10.2105/AJPH.2011.300177 | 2011 | 901 |
| Richard D Telford | 10.2105/AJPH.2011.300220 | 2012 | 620 |
| French | 10.2105/AJPH.2018.304696 | 2018 | 534 |
| JF Sallis | 10.2105/AJPH.87.8.1328 | 1997 | 955 |
| Ewart | 10.2105/ajph.88.6.949 | 1998 | 88 |
| Colchico | 10.2105/ajph.90.6.977 | 2000 | 30 |
| French | 10.2105/ajph.94.9.1507 | 2004 | 750 |
| Bayne-Smith | 10.2105/ajph.94.9.1538 | 2004 | 442 |
| M Bayne-Smith | 10.2105/ajph.94.9.1538 | 2004 | 442 |
| Pate | 10.2105%2FAJPH.2004.045807 | 2005 | 2744 |
| Safdie | 10.21149/spm.v55s3.5138 | 2013 | 830 |
| Kellam | 10.21149/spm.v55s3.5142 | 2013 | 62 |
| G Fiorilli | 10.2147/DMSO.S122110 | 2017 | 41 |
| Huang | 10.2147/DMSO.S223514 | 2019 | 21 |

| Trials_1st_author | Trial_DOI | Trial_year | Participants_sample_size |
|---|---|---|---|
| A G Militão | 10.2147/DMSO.S52166 | 2013 | 34 |
| Karner-Rezek | 10.2147/IJGM.S40187 | 2013 | 28 |
| Falk | 10.2165/00007256-199622030-00004 | 1996 | 32 |
| J Mota | 10.21767/2471-8173.100010 | 2016 | 17 |
| A Direito | 10.2196/jmir.4568 | 2015 | 34 |
| D Cvejic | 10.22190/FUPES1703437C | 2017 | 178 |
| Cvetković | 10.22190/FUPES180604039M | 2018 | 42 |
| WL Do Prado | 10.2223/JPED.1889 | 2009 | 688 |
| R G McMurray | 10.2310/6650.2007.06031 | 2007 | 58 |
| S J Lee | 10.2337/db12-0214 | 2012 | 45 |
| KC Copeland | 10.2337/dc12-2534 | 2013 | 699 |
| M Savoye | 10.2337/dc13-1571 | 2014 | 75 |
| Leandro | 10.23736/S0022-4707.20.11648-7 | 2021 | 41 |
| Procopio, A. M | 10.23860/thesis-procopio-andrew-2014 | 2014 | 6 |
| MA Paschoal | 10.24220/2318-0897v27n3a4196 | 2018 | 15 |
| D Mayorga-Vega | 10.2466/06.10.25.PMS.115.6.984-996 | 2012 | 75 |
| Zervas | 10.2466/pms.1991.72.3c.1215 | 1991 | 18 |
| Jung | 10.26582/k.50.1.2 | 2018 | 20 |
| Bielec | 10.3109/01460862.2013.777818 | 2013 | 243 |
| L Larsen | 10.3109/02813432.2015.1067511 | 2015 | 80 |
| Emily C-S Murphy | 10.3109/17477160902846187 | 2009 | 35 |
| S Berntsen | 10.3109/17477160902957166 | 2010 | 60 |
| B E Saelens | 10.3109/17477166.2010.482157 | 2010 | 29 |
| S W Wafa | 10.3109/17477166.2011.566340 | 2011 | 107 |
| W Jansen | 10.3109/17477166.2011.575151 | 2011 | 2622 |
| Lillegaard | 10.3109/17518429709167353 | 1997 | 91 |
| Maurice Duggins | 10.3122/jabfm.2010.03.080266 | 2010 | 83 |
| Violet Siwik | 10.3122/jabfm.2013.02.120118 | 2013 | 35 |
| Djaafar | 10.31838/ijpr/2019.11.02.043 | 2019 | 98 |
| JL Ramirez | 10.3305/nh.2012.27.3.5725 | 2012 | 84 |
| M Bacardí-Gascon | 10.3305/nh.2012.27.3.5756 | 2012 | 532 |
| F Vásquez | 10.3305/nh.2013.28.2.6159 | 2013 | 111 |
| Ausias Cebolla i Martí | 10.3305/nh.2015.31.2.7929 | 2014 | 42 |
| Julian Hamilton-Shield | 10.3310/hta18470 | 2014 | 36 |
| Kim | 10.33549/fisiolres.932997 | 2016 | 28 |

| Trials_1st_author | Trial_DOI | Trial_year | Participants_sample_size |
|---|---|---|---|
| P Zhao | 10.3389/fendo.2022.972954 | 2013 | 45 |
| Gallotta | 10.3389/fnhum.2015.00577 | 2015 | 78 |
| C Pesce | 10.3389/fpsyg.2016.00349 | 2016 | 920 |
| Xiang | 10.3389/fpsyg.2019.01385 | 2019 | 44 |
| Jana Markert | 10.3390/ijerph111010327 | 2014 | 289 |
| R L Lee | 10.3390/ijerph14101178 | 2017 | 115 |
| Ye | 10.3390/ijerph16214080 | 2019 | 81 |
| Garza | 10.3390/ijerph16245133 | 2019 | 71 |
| Roh | 10.3390/ijerph17072505 | 2020 | 20 |
| Romero-Perez | 10.3390/ijerph17134655 | 2020 | 105 |
| Y Liang | 10.3390/ijerph17217984 | 2020 | 87 |
| J McGann | 10.3390/ijerph191911914 | 2019 | 54 |
| Walid El Ansari | 10.3390/ijerph7041649 | 2010 | 160 |
| Piziak | 10.3390/ijerph9041319 | 2012 | 413 |
| Rafaela Rosário | 10.3390/ijerph9041355 | 2012 | 464 |
| Meng Cao | 10.3390/jcm11185436 | 2022 | 40 |
| S Xiong | 10.3390/jcm8040469 | 2019 | 60 |
| Y G Seo | 10.3390/nu11010137 | 2019 | 103 |
| P Warshburger | 10.3390/nu11092053 | 2019 | 266 |
| H J Yu | 10.3390/nu12010194 | 2020 | 171 |
| AE Rigamonti | 10.3390/nu12010208 | 2020 | 644 |
| Špela Bogataj | 10.3390/nu13010238 | 2021 | 48 |
| Ming-Xiao Sun | 10.3760/cma.j.issn.0366-6999.2011.03.001 | 2011 | 51 |
| Alberga | 10.3810/psm.2013.09.2028 | 2013 | 19 |
| Ozgur Sevinc | 10.3906/sag-1009-1179 | 2011 | 6847 |
| SB Murer | 10.3945/ajcn.110.002212 | 2011 | 203 |
| M Ralph | 10.3945/ajcn.110.009142 | 2011 | 322 |
| Mirza | 10.3945/ajcn.112.042630 | 2013 | 113 |
| RW Taylor | 10.3945/ajcn.2007.25749 | 2008 | 389 |
| M Waling | 10.3945/jn.110.125435 | 2010 | 83 |
| Xiuming, Z | 10.3969/j.issn.1004-0188.2014.04.023 | 2014 | 52 |
| Kim YH & Yang YO | 10.4040/jkan.2005.35.5.858 | 2005 | 27 |
| Y H Lee | 10.4070/kcj.2010.40.4.179 | 2010 | 54 |
| DS Buchan | 10.4081/jphr.2012.e24 | 2012 | 41 |
| S Suh | 10.4093/dmj.2011.35.4.418 | 2011 | 38 |

| Trials_1st_author | Trial_DOI | Trial_year | Participants_sample_size |
|---|---|---|---|
| Regaieg | 10.4103/2230-8210.122619 | 2013 | 28 |
| Maahs | 10.4158/ep.12.1.18 | 2006 | 40 |
| CW Kim | 10.4162/nrp.2020.14.3.262 | 2020 | 66 |
| Vivian | 10.4172/2165-7904.1000191 | 2013 | 67 |
| Da Silva | 10.4187/respcare.01307 | 2012 | 35 |
| M Mendelson | 10.4187/respcare.02948 | 2014 | 18 |
| DY Seo | 10.4196/kjpp.2012.16.3.175 | 2012 | 20 |
| Dae Yun Seo | 10.4196/kjpp.2012.16.3.175 | 2012 | 20 |
| F. Duman | 10.4274/jcrpe.2284 | 2016 | 50 |
| J S Rosa | 10.4274/jcrpe.v3i3.23 | 2011 | 66 |
| Pate RP | 10.4278/0890-1171-17.3.171 | 2003 | 436 |
| Abroms L.C | 10.4278/0890-1171-19.1.28 | 2004 | 70 |
| Matvienko, O., & Ahrabi-Fard, I. | 10.4278/ajhp.08050146 | 2010 | 70 |
| Romero | 10.4278/ajhp.090123-ARB-24 | 2012 | 81 |
| Cronk | 10.4278/ajhp.091222-quan-396 | 2011 | 54 |
| Feng | 10.4278/ajhp.140801-QUAN-384 | 2016 | 555 |
| Fredericks | 10.4314/sajrs.v28i1.25929 | 2006 | 58 |
| L. S. Eddy Ives | 10.4321/S1139-76322012000300005 | 2012 | 174 |
| Ceballos-Gurrola | 10.47197/retos.v38i38.77003 | 2020 | 62 |
| Laura L Bellows | 10.5014/ajot.2013.005777 | 2013 | 201 |
| CCW Yu | 10.5409/wjcp.v5.i3.293 | 2016 | 38 |
| R Martin | 10.5604/20831862.1173644 | 2015 | 49 |
| G Racil | 10.5604/20831862.1198633 | 2016 | 47 |
| B Ghorbanian | 10.5812/ijem.8178 | 2013 | 30 |
| Pahoo | 10.5812/ijp.99760 | 2020 | 45 |
| Arlinghaus | 10.5888/pcd14.170130 | 2017 | 189 |
| Saygin | 10.5897/AJPP11.114 | 2011 | 39 |
| LRA Lima | 10.5935/2359-4802.20170032 | 2017 | 45 |
| Olvera | 10.5993/ajhb.34.2.2 | 2010 | 37 |
| D Thompson | 10.5993/AJHB.40.4.9 | 2016 | 80 |
| SL Silva | 10.6063/motricidade.9(1).2459 | 2013 | 38 |
| Q Tang | 10.6133/apjcn.2014.23.3.02 | 2014 | 20 |
| A Lai | 10.7150/ijbs.4918 | 2013 | 88 |
| AL Ricci-Vitor | 10.7322/jhgd.119257. | 2016 | 15 |
| J. Kim | 10.7570/kjo.2011.20.3.138 | 2011 | 45 |
| Starkoff | 10.7575/aiac.ijkss.v.2n.3p.1 | 2014 | 27 |

| Trials_1st_author | Trial_DOI | Trial_year | Participants_sample_size |
|---|---|---|---|
| Xie | 10.7717/peerj.3286 | 2017 | 58 |
| Subramanian | 10.7860/JCDR/2015/14881.6818 | 2015 | 439 |
| Koubaa | 10.9790/3013-32103137 | 2013 | 29 |
| Xiuming | ANF | | 70 |
| Jago R | ANF | 2003 | 46 |
| Ozgur, H., & Cakmakci, E. | ANF | 2011 | 40 |
| VanDongen R | ANF | 1995 | 1147 |
| Guijun | ANF | 2009 | 47 |
| Liangmei | ANF | 2015 | 24 |
| Mingwei | ANF | 2015 | 41 |
| Ting | ANF | 2015 | 50 |
| Xinhui | ANF | 2016 | 28 |
| Chunyan | ANF | 2016 | 25 |
| Yuan | ANF | 2017 | 64 |
| Dongqi | ANF | 2018 | 68 |
| Mu Qin | ANF | 2020 | |
| Huang | ANF | 2011 | |
| M E Errickson | NR | 2014 | 24 |
| St. Elizabeth's Medical Center | NR | 2005 | 30 |
| K D Brownell | NR | 1983 | 42 |
| Icahn School of Medicine at Mount Sinai | NR | 2008 | 77 |
| Adsiz | NR | 2012 | 60 |
| Manjunath | NR | 2001 | 34 |
| Manjunath | NR | 2004 | 60 |
| Katz | NR | 2010 | 1106 |
| Faigenbaum | NR | 2006 | 118 |
| Viciana | NR | 2013 | 75 |
| Benson | NR | 2008 | 78 |
| Yoshimoto | NR | 2016 | 52 |
| Lau | NR | 2004 | 38 |
| Takai | NR | 2013 | 94 |
| Bush | NR | 1989 | 431 |
| Stephens | NR | 1998 | 90 |
| Rocchini | NR | 1987 | 50 |
| Wong | NR | 2008 | 24 |

| Trials_1st_author | Trial_DOI | Trial_year | Participants_sample_size |
|---|---|---|---|
| Burke V | NR | 1998 | 471 |
| Connolly P, McKenzie TL | NR | 1995 | 56 |
| Brown MK | NR | 1981 | 38 |
| Campaigne BN | NR | 1982 | 19 |
| Sanders, K | NR | 2014 | 24 |
| L Mo-Suwan | NR | 1998 | 170 |
| T Baranowski | NR | 2003 | 31 |
| M Story | NR | 2003 | 53 |
| BM Beech | NR | 2003 | 30 |
| TN Robinson | NR | 2003 | 61 |
| BM Beech | NR | 2003 | 30 |
| Kain | NR | 2004 | 1636 |
| James | NR | 2004 | 574 |
| Paineau | NR | 2008 | 471 |
| Vizcaino | NR | 2008 | 530 |
| Haerens | NR | 2006 | 709 |
| Haerens | NR | 2006 | 557 |
| Y M Kim | NR | 2010 | 20 |
| Dietrich and Widhalm | NR | 2004 | 59 |
| PC Wong | NR | 2008 | 24 |
| AFNJ Brandão | NR | 2015 | 25 |
| JK Song | NR | 2012 | 22 |
| A Faigenbaum | NR | 2006 | 118 |
| G Jurak | NR | 2008 | 328 |
| PC Wong | NR | 2008 | 12 |
| A V Biljon | NR | 2012 | 31 |
| B L Rooney | NR | 2005 | 87 |
| JK Song | NR | 2012 | 22 |
| E Moslehi | NR | 2019 | 30 |
| O Saygın | NR | 2011 | 39 |
| Brandao | NR | 2015 | 27 |
| Lee | NR | 2012 | 44 |
| Shultz | NR | 2014 | 3 |
| Rocchini | NR | 1988 | 72 |
| Hayashi | NR | 1987 | 18 |
| Sijie | NR | 2012 | 60 |

| Trials_1st_author | Trial_DOI | Trial_year | Participants_sample_size |
|---|---|---|---|
| Wong | NR | 2008 | 24 |
| CCW Yu | NR | 2005 | 82 |
| Cao F | NR | 2012 | 40 |
| Q Chen | NR | 2015 | 60 |
| HL Cheng | NR | 2012 | 60 |
| BL Rooney | NR | 2005 | 59 |
| D Silva | NR | 2012 | 14 |
| JK Song | NR | 2012 | 22 |
| LM Boddy | NR | 2010 | 16 |
| Boer | NR | 2014 | 46 |
| Y Chansavang | NR | 2015 | 18 |
| Horton | NR | 2013 | 361 |
| Cao F | NR | 2012 | 40 |
| Cheng | NR | 2012 | 60 |
| Song | NR | 2012 | 22 |
| Wong | NR | 2008 | 24 |
| Jung | NR | 2016 | 23 |
| Tsang | NR | 2009 | 20 |
| Kim | NR | 2010 | 20 |
| Takai | NR | 2013 | 94 |
| Yoshimoto | NR | 2016 | 52 |
| Yetgin | NR | 2018 | 16 |
| Cao | NR | 2012 | 40 |
| Juliana Pizzi | NR | 2017 | 54 |
| Pizzi | NR | 2017 | 54 |
| Zorba | NR | 2011 | 40 |
| IBF Dias | NR | 2013 | 44 |
| AR Dâmaso | NR | 2014 | 146 |
| Ulices Alvirde-García | NR | 2012 | 1224 |
| Eloisa Colín Ramírez | NR | 2009 | 619 |
| Gonzalez-Heredia | NR | 2014 | 60 |
| Rodrıguez-Moran | NR | 2014 | 115 |
| Rosas-Nexticapa | NR | 2017 | 121 |
| Mohammadi | NR | 2011 | 1300 |
| Peng | NR | 2015 | 121 |
| Roshan | NR | 2011 | 24 |

| Trials_1st_author | Trial_DOI | Trial_year | Participants_sample_size |
|---|---|---|---|
| Talakoub | NR | 2012 | 72 |
| Zhang | NR | 2021 | 138 |
| K J Kim | NR | 2012 | 30 |
| Pizzi | NR | 2017 | 54 |
| Van Biljon A | NR | 2012 | 31 |
| Mahgoub, M. S. E., and Aly, S | NR | 2015 | 30 |
| Cheunsiri | NR | 2018 | 48 |
| Haghshenas | NR | 2019 | 100 |
| R Flores | PMCID: PMC1382101 | 1995 | 110 |
| M S Sothern | PMID: 10432772 | 1999 | 87 |
| L H Epstein | PMID: 10965646 | 2000 | 67 |
| Tom Baranowski | PMID: 12713209 | 2003 | 35 |
| Bettina M Beech | PMID: 12713210 | 2003 | 60 |
| Mary Story | PMID: 12713211 | 2003 | 54 |
| Thomas N Robinson | PMID: 12713212 | 2003 | 61 |
| T J Yin | PMID: 15977132 | 2005 | 140 |
| Rooney | PMID: 16138517 | 2005 | 87 |
| Liu AL | PMID: 17458137 | 2007 | 753 |
| S H Huang | PMID: 18265541 | 2007 | 120 |
| P C Wong | PMID: 18461212 | 2008 | 24 |
| Gentile, D.A | PMID: 19765270 | 2009 | 1323 |
| Ming-Xiao Sun | PMID: 21362327 | 2011 | 93 |
| Iversen | PMID: 21886292 | 2011 | 119 |
| J W Farris | PMID: 22163176 | 2011 | 25 |
| E Zorba | PMID: 22212270 | 2011 | 40 |
| Epstein | PMID: 2232019 | 1990 | 76 |
| C D Legantis | PMID: 22648470 | 2012 | 48 |
| J K Song | PMID: 22976739 | 2012 | 22 |
| Cecile Rausch Herscovici | PMID: 24096971 | 2013 | 405 |
| Tsang, T | PMID: 24150562 | 2009 | 30 |
| Shultz, S | PMID: 25435766 | 2014 | 11 |
| Morrissey C | PMID: 29710370 | 2018 | 29 |
| Bonney E | PMID: 31568378 | 2019 | 52 |
| Rocchini AP | PMID: 3288957 | 1988 | 45 |
| Irandoust K | PMID: 33176381 | 2021 | 59 |
| Becque MD | PMID: 3357722 | 1988 | 22 |

| Trials_1st_author | Trial_DOI | Trial_year | Participants_sample_size |
|---|---|---|---|
| Savage MP | PMID: 3702647 | 1986 | 48 |
| Mellin | PMID: 3819254 | 1987 | 66 |
| Linder CW | PMID: 6353127 | 1983 | 50 |
| C E Flodmark | PMID: 8474806 | 1993 | 1774 |
| Baranowski T | PMID:12713209 | 2003 | 35 |

**eTable 5. Characteristics of 138 reviews included.**

| Study | Country | Design | Participants | Included studies (n) | Objective | Setting | Who receive the intervention | Intervention(s) |
|---|---|---|---|---|---|---|---|---|
| Bento et al., 2021 | Spain | SR | F and M; 6-12 years; Overweight/ Obesity; Other Health conditions = NR. n = NR | 14 RCTs | To evaluate the utility of a HIIT program integrated into high school PEC on physical fitness, PA, and motivation for exercise | NR | Adolescents | HIIT, MIIT,Control group, HIIE, LIIE, P+HIIT, Football. 6-12 weeks. |
| Bento et al., 2022 | Spain | SR | F and M; 10-19 years; Normal weight + overweight + obesity; Other conditions = NR; n=NR. | 14 RCTs | In an aIn an attempt to broaden knowledge about evidence that relates to the variables mentioned above, this systematic review was based on the following question: What are the effects of a HIIT program integrated into high school PEC (students aged 10–19 years) on physical fitness, PA, and motivation for exercise? The aim of this review was (a) to evaluate the utility of a HIIT program integrated into high school PEC on physical fitness, PA, and motivation for exercise and (b) to evaluate the effects of protocols and periodization on previous outcomes. | School | Adolescents | Aerobic 8 weeks; HIIT 7-12 weeks; MIIT 12 weeks; LIIE 12 weeks. |

| Study | Country | Design | Participants | Included studies (n) | Objective | Setting | Who receive the intervention | Intervention(s) |
|---|---|---|---|---|---|---|---|---|
| Burns et al., 2019 | United States of America | SRMA | F and M; 12-18 years; Normal weight + obesity + overweight; Other conditions: NR; n = 311. | 9 RCTs | The purpose of this study was to examine the pooled effect of resistance training interventions on insulin sensitivity in children and adolescents. | NR | Adolescents | Resistance training 12-16 weeks; Resistance training + nutritional education 16 weeks. |
| Collins et al., 2019 | United Kingdom | SRMA | F and M; 5-18 years; Normal weight + Overweight + obesity; Other conditions NR; n = 256. | 11 RCTs | argued that there has been limited research on the effects of RT on the psychological well-being of children and adoles- cents, and 14 years later, the position appears to have changed little [19]. The current review will therefore be the first to examine the isolated effect of RT on 'the self'. Hence, the purpose of this review was to systematically examine the effect of RT interventions on 'the self' in youth. | School and Community | Adolescents | Control time NR; Strength training 8-24 weeks. |
| da Rosa-Santos et al., 2019 | Brazil | SR | F and M; 12-19 years; Overweight; Other conditions = NR; n = 20 | 17 RCTs | Therefore, the purpose of this systematic review was to examine the published literature on the influence and morphofunctional adaptations related to CRF in response to resistance exercise among adolescents, in order to identify its main benefits and responses | NR | Adolescents | Combined resistance and Aerobics 10 weeks; Resistance 12 weeks |

| Study | Country | Design | Participants | Included studies (n) | Objective | Setting | Who receive the intervention | Intervention(s) |
|---|---|---|---|---|---|---|---|---|
| | | | | | related to decreasing the CRF in this age range. | | | |
| Da Silva et al., 2020 | Brazil | SR | F and M; adolescents; age: NR; Overweight + Obesity; Other conditions: NR; n = 169 | 11 RCTs and n-RCTs | to gather the latest information about the effects of different protocols of HIIT on adipokine plasma concentrations. | NR | Adolescents | HIIT: 12-26 weeks; Ergometer rowling, cycle ergometer, outdoor all-out shots. |
| Farah et al., 2012 | Brazil | SR | F and M; 10-17y; Overweight/obesity; Other health condition= NR; n=760 | 8 RCTs | To systematically review the results of studies that analyzed the effects of physical training on the blood pressure of obese adolescents | NR | Adolescents | PA (aerobic and combined exercises); 1 – 6 days/wk, 40 – 90'/day, 55 - 95% HRmax and 55 – 75% 1RM, 8 to 48 weeks |
| Martin-Smith et al., 2020 | United Kingdom | SRMA | F and M; 10,7 – 17 years; Obesity and Overweight; Other conditions: NR; n = NR | 18 RCTS | The aim of this systematic review and meta-analysis was to primarily investigate if a statistically significant difference existed in the effect size of HIIT on CRF when comparing studies that used: indirect calorimetry (laboratory) vs. predicted (field) measures of CRF, MICE or No Exercise as the control group, and unfit vs. fit or obese vs. healthy weight adolescents. | NR | Adolescents | HIIT/Sprint interval training 5-15 weeks. |
| Ribeiro et al., 2022 | Portugal | SRMA | F and M; 10,9 – 17,7 years; obesity; other | 21 RCTs | This review aimed to summarize the findings and conclusions | NR | Adolescents | Isolated Resistance training (RT) program 4-48 weeks; |

| Study | Country | Design | Participants | Included studies (n) | Objective | Setting | Who receive the intervention | Intervention(s) |
|---|---|---|---|---|---|---|---|---|
| | | | conditions: NR; n = NR. | | reported in the literature on the effect of RT in adolescents. | | | |
| Ruotsalainen et al., 2015 | Finland | SR | F and M; 10-18y; Overweight/obesity; Other health conditions= No; n= 1321 | 14 RCTs | To examine the effects of physical activity and exercise interventions on body mass index, subsequent physical activity and psychological symptoms for overweight and obese adolescents | Home; Home and community; Schools; Hospitals | Adolescents | PA (supervised exercise); Others (behavioral management skills; material and information support; social support); PA + Others; PA + Diet (Dietary regimen or restriction); 8 to 24 weeks |
| Shin et al., 2019 | Korea | SRMA | F and M; 10-19 years; Normal weight + obesity+ overweight; Other conditions: NR. n = 1472. | 11 RCTS | Therefore, the current study aims to (1) systematically review mobile phone interventions among adolescents, (2) statistically analyze intervention effects, and (3) provide evidence that interventions were optimized to promote physical he | Hong kong, Australia. | Adolescents | Use of mobile phone: personalized SMS 24 weeks."switch-off 4 healthy minds" 6month24weeks. Use of mobile phone 3mont12weeks. Apps for improving fitness 8 weeks. And others. |

| Study | Country | Design | Participants | Included studies (n) | Objective | Setting | Who receive the intervention | Intervention(s) |
|---|---|---|---|---|---|---|---|---|
| Stoner et al., 2016 | New Zealand | SRMA | F and M; 10-19y; Overweight/obesity; Other health conditions = prehypertension; n= 556 | 13 RCTs | To estimate whether exercise intervention meaningfully improves body composition and cardiometabolic risk factors in overweight and obese adolescents; and discuss the implications of the findings in terms of primary healthcare provision and public health policy, using New Zealand as an exemplar context | School; Hospital; University | Adolescents | PA (aerobic, resistance and combined exercises); Educational (nutritional and PA behavior changes); PA + Educational; 2-3 days/wk, 60 – 90'/day, 8 to 36 weeks |
| Vasconcellos et al., 2014 | Brazil | SR | F and M; 12-17y; Overweight/obesity; Other health conditions = NR; n=1635 | 24 RCTs, non-RCTs and pre-post test studies | To systematically review the effect of PA interventions on body composition, physical fitness components, hemodynamic variables, biochemical markers, endothelial function, and low-grade inflammation in overweight and obese adolescents | NR | Adolescents | PA (aerobic, resistance and combined exercises; sports; dance); PA + Diet or Educational; 2 – 5 days/wk, 16 -90'/day, 7 to 52 weeks |
| Wolfa et al., 2019 | Brazil | SR | F and M; 10-17 years; Normal weight + overweight + obesity; Other conditions: NR. N = NR. | 12 RCTs | to check the efficacy of educational interventions based on orientation about physical activities and nutrition for adolescent students. | School | Adolescents | Educational intervention 10-20 weeks; educational + nutritional counseling; Educational + physical activity 12-104 weeks. |

| Study | Country | Design | Participants | Included studies (n) | Objective | Setting | Who receive the intervention | Intervention(s) |
|---|---|---|---|---|---|---|---|---|
| Zhao et al., 2022-a | China | SRMA | F and M, 10-20 years; Obesity; Other conditions: NR; n = 307. | 15 RCTS | To systematically assess the effects of ECWD, exercise alone, diet alone, and no intervention on body composition and serum biochemical markers in adolescents with obesity. | NR | Adolescents | Exercise Only(aerobic) 6-24 weeks; Exercise training 4-52 weeks; Diet 4 weeks- 1year. |
| Zhao et at., 2022-b | China | SRMA | F and M; 13-18 years; Obesity; other conditions: NR; n = NR. | 14 RCTs | to compare the effects of different training modalities on the inflammatory response level in adolescents with obesity | NR | Adolescents | Aerobic training 6-24 weeks; Aerobic training + resistance training; 8-24 weeks; Resistance training 8-22 weeks; HIIT 12 WEEKS; |
| Zhu et al., 2021 | China | SRMA | F and m; 7-16 years; Obesity and overweight; other conditions = no. n= 488. | 11 RCTs | Perform a meta-analysis to quantitatively evaluate the effects and safety of HIIT in the treatment of adolescent obesity, to provide a theoretical basis for the treatment old adolescent obesity | NR | Adolescents | HIIT for 6-12 weeks; Blank control 6-12 weeks; Dietary intervention 12 weeks; Walking 12 weeks; |
| Al-Khudairy et al., 2017 | United Kingdom | SRMA | F and M; 12-17 years; Obesity + overweight; Other conditions: NR ; n = 4781. | 44 RCTs | To assess the effects of diet, physical activity and behavioural interventions for the treatment of overweight or obese adolescents aged 12 to 17 years. | Schools, community, home, research clinics and hospitals | Adolescents and parents | Nutritional + Physcial Activity + Behavioral components 26-104 weeks; usual care 26-104 weeks. |

| Study | Country | Design | Participants | Included studies (n) | Objective | Setting | Who receive the intervention | Intervention(s) |
|---|---|---|---|---|---|---|---|---|
| Boff et al,. 2017 | Brazil | SR | F and M; 10-19y; Overweight/obesity; Other health condition= NR; n= 2099 | 26 RCTs | To determine what factors contribute to the efficacy of non-drug treatments designed to promote weight loss in overweight and obese adolescents | Clinics; Homes; Schools | Adolescents and parents | Educational (PA and/or nutrition education); PA (aerobic and resistance exercises; games; yoga); 10 weeks to 24 months |
| Silva et al., 2014 | Spain | SR | F and M; 12-19y; Overweight/obesity; Other health conditions= NR; n= 2501 | 23 RCTs and non-RCTs | To realize one systematic review the effectiveness of intervention programs which were based on physical activity and dietary control of overweight and/or obese children and adolescents | School | Adolescents and parents | Educational (material and counseling about healthy eating, increased PA levels and reduction of screen time); 2 - 5 days/wk, 45 -90'/day, 12 weeks to 5 years |
| Chen et al., 2021 | China | SRMA | F and M; 8-10 (mean); Overweight + Obesity; Other condition: NR. N = 504. | 12 RCTs | To analyze the effects of aerobic exercise and resistance exercise on physical indexes and cardiovascular risk factors in school-age children with obesity or overweight. | NR | Children | Aerobic Exercise 8-12 weeks; Resistance Exercise 12 weeks; Combined Exercises (AE+ RE) 16 weeks; Aerobic Exercise + Diet 16 weeks; Control 8-16 weeks. |
| Dias et al., 2018 | Brazil | SR | F and M; 8-15y; Overweight/obesity; Other health condition= NR; n= 2484 | 6 RCTs | To identify in the literature the efficacy of serious games to improve knowledge for and/or behavioral changes among overweight or obese children | School; Home; Medical center | Children | Educational (games about diet and exercise); 9-25 sessions, 25-40'/session, 1-6 months |

| Study | Country | Design | Participants | Included studies (n) | Objective | Setting | Who receive the intervention | Intervention(s) |
|---|---|---|---|---|---|---|---|---|
| Garcia-Hermoso et al., 2020-a | Spain | SRMA | F and M; 5-12 years; normal weight + overweight + obesity. other condition: NR. n = 48185. | 66 RCTs | to examine the association of quality- and quantity-based Physical Education interventions with health-related physical fitness and Fundamental Motor Skills in children and adolescents. | NR | Children | Fitness infusion 28-39 weeks; Teaching strategies; |
| Hale et al., 2023 | United Kingdom | SR | F and M; 6-11 years; Normal weight + Overweight + Obesity; Other condition = NR; n = 1392. | 23 RCTs | Thus, taking previous research into account, the aim of the current review was to (1) provide an update on the literature of PA interventions for children under 11 years old, (2) identify any gaps in knowledge and research in this area, (3) summarize the key intervention components (e.g., PA type and education) and discuss effectiveness in enhancing the psychological well-being and ill-being of children. | School | Children | Aerobics 9-35 weeks; Aerobics + football 20-26 Weeks; Aerobics + sports and fitness skills 26 weeks; |
| Liu, W et al., 2020 | China | SR | F and M; 6-12 years; overweight + obesity; other conditions: NR; N= NR. | 10 RCTs | review to summarize the empirical findings regarding the impact of Active Video Games on healthy children’s Fundamental Motor Skills and physical fitness. | School and lab | Children | Active video game(Nintendo Wii and Xbox Kinect, and one study developed their own purpose-built AVG)s 6-12 weeks; |

| Study | Country | Design | Participants | Included studies (n) | Objective | Setting | Who receive the intervention | Intervention(s) |
|---|---|---|---|---|---|---|---|---|
| Mijalkovic et al., 2022 | Serbia | SR | F and M; 6-10 years; Obesity; Other conditions: NR. n = 2810. | 13 RCTs | The aim of this study was to conduct a systematic review of the school-based exercise programs for promoting cardiorespiratory fitness in overweight and obese children aged 6 to 10. | School | Children | AL - Additional lessons, 26 weeks (60min). Kids4Fit-Multidisciplinary weight reduction program 12-156weeks (2 x week; 2 x weeks (20min). |
| Saavedra et al., 2011 | Spain | SRMA | F and M; school age; Overweight/obesity; Other health conditions= NR; n= 349 | 9 RCTs and non-RCTs | To assess the effectiveness of diverse interventions in aerobic fitness adjusted for weight in obese children | NR | Children | PA (aerobics, resistance and combined exercises; sports); 3-5 days/wk, 29-90'/day, 8 to 36 weeks |
| Seral-Cortes et al., 2021 | Spain | SRMA | F and M; 6-12 years; normal weight + obesity; other conditions: diabetes Mellitus II; n = NR. | 41 RCTs | to identify the effectiveness in terms of body composition parameters in a generation of articles to prevent obesity and T2DM in children. | School and community center. Medical center; | Children | PA, D and BS, 14-168 Weeks; PA and BD for 24-192; PA and D 12-96 weeks; D and BS for 72 weeks; D 6-144 weeks; PA 6-144 weeks; BS 24-96weeks. |
| Snethen et al., 2016 | United States of America | SRMA | F and M; 6-15y; Overweight/obesity; Other health condition= NR; n= 2317 | 16 RCTs and non-RCTs | To examine studies related to overweight children and to determine which weight loss interventions were effective versus no intervention in promoting weight loss | NR | Children | PA; Diet; PA + Diet; Educational; Diet + Educational; Others (behavioral). *There is no additional information on interventions |

| Study | Country | Design | Participants | Included studies (n) | Objective | Setting | Who receive the intervention | Intervention(s) |
|---|---|---|---|---|---|---|---|---|
| Solera-Martinez et al., 2021 | Spain | SRMA | F and M; 5-12 years; Obesity and overweight; Other conditions: NR. N = 512. | 11 RCTS | synthesize evidence regarding the effectiveness of HIIT interventions on improving cardiovascular risk factors and cardiorespiratory fitness (CRF) in children from 5 to 12 years old. | Cycling and Shuttle runs. | Children | HIIT 6-12 weeks. |
| Aceves-Martins et al., 2022 | United Kingdom | SRMA | F and M; 8.8 -15.8 years; Obesity + Overweight. Other conditions: Diabetes Mellitus. N = 2302. | 4 RCTs; 25 non RCTS | In this study, we aimed to systematically review experimental studies assessing lifestyle, environmental, behavioral, pharmacologic, or surgical interventions to treat obesity in Mexican children and adolescents (<18 years old). | - Public secondary school in a municipality in the urban area; - Public primary care unit within a secondary care hospital; - Pediatric obesity clinic at a public hospital; - Family medicine unit of a public hospital; - Public elementary school; - Public children's hospital; - Clinical nutrition research unit of a public hospital; - Public family medicine unit; - Pediatric clinic of a third-level hospital; - Private school | Children and adolescents | Nutritional Component: 4-52 weeks; PA component 4-52 weeks; Psychological or behavioral component: 2-12 weeks, And others. |

| Study | Country | Design | Participants | Included studies (n) | Objective | Setting | Who receive the intervention | Intervention(s) |
|---|---|---|---|---|---|---|---|---|
| Álvarez-Bueno et al., 2017 | Spain | SRMA | F and M; 4-14y; normal weight + Overweight/obesity; Other health condition= NR; n= 5527 | 36 RCTs and non-RCTs | To assess the effect of physical activity interventions on children's and adolescents' cognition and metacognition | Curricular school time; Out of school; Summer camp | Children and adolescents | PA (physical education lessons; active breaks; PA extracurricular activities); 4 to 54 weeks |
| Andrade et al., 2019 | Brazil | SRMA | F and M; 6-19 years; Overweight and Obesity; Other condition = NR. n = 336. | 9 RCT | Thus, the objective of the study was to verify the psycho- logical effects of exergames for children and adolescents with overweight or obesity through a systematic review and meta-analysis of experimental studies. To the best of our knowledge, this is the first study that summarizes and per- forms a meta-analysis specifically on psychological effects of exergames on children and adolescents with overweight or obesity. | Laboratory, laboratory and school, community Center, | Children and adolescents | Control 6-20 weeks; Exergames 6-20 weeks; Competitive games; Cooperative games; Several games, Wii, Xbox, Dance Revolution, ranging from 30 - 60 min of activity for 10 weeks, 3 times per week; Exergame + physical activity 1 time per week, for 10 weeks. |

| Study | Country | Design | Participants | Included studies (n) | Objective | Setting | Who receive the intervention | Intervention(s) |
|---|---|---|---|---|---|---|---|---|
| Birch et al., 2019 | United Kingdom | SR | F and M; 4-19 years; Obesity; other conditions = yes. n = 8 to 203. | 98 RCTs | This paper aims to establish the minimum change in BMI-SDS needed to effect improvements in adiposity markers of children and adolescents with obesity. This is the first of a series of three papers reporting on the find- ings from studies identified in a large systematic review (n=90 studies; searched up to May 2017) and focuses on the evidence in relation to adiposity (percentage body fat); the others relating to metabolic and cardiovascular health | Clinic (Hospital) Academic Institution; Community | Children and adolescents | Exercise + diet 12-24 weeks |
| Bondyra-Wisniewska et al., 2021 | Poland | SR | F and M; 6-18 years; Obesity and Overweight; Other conditions: NR. N = 1587. | 23 RCTS | The aim was to identify common components of lifestyle interventions leading to weight loss and improvement of selected cardiometabolic parameters | NR | Children and adolescents | Nutritional Intervention 4weeks-2years; Physical Activity 10 weeks – 2 years. |
| Brown et al., 2015 | United Kingdom | SRMA | F and M; School age; Normal weight + overweight/obesity; Other health conditions= type 2 diabetes and metabolic syndrome; n= 11527 | 29 RCTs, non-RCTs and pre-post test studies | To assess the effectiveness of diet and physical activity interventions to prevent or treat obesity in South Asians living in or outside of South Asia and to describe the characteristics of effective interventions | General practice; Community; Home; Hospital; School; University and workplace | Children and adolescents | PA; PA + Diet; 3 to 8 months. *There is no additional information on interventions |

| Study | Country | Design | Participants | Included studies (n) | Objective | Setting | Who receive the intervention | Intervention(s) |
|---|---|---|---|---|---|---|---|---|
| Busnatu et al., 2022 | Romania | SRMA | F and M; 6-18 years; Overweight; Other conditions: NR; n: 479. | 12 RCTs | to evaluate the effectiveness of exercise in reducing body weight, BMI, diastolic blood pressure (DBP), systolic blood pressure (SBP), HDL, LDL, and blood glucose, and waist circumference in overweight children and adolescents. | Treadmill, walking-jogging, running sprints, weight machines and free weight | Children and adolescents | Aerobic training 4-24 weeks; Strength training 12-24 weeks; Multicomponent 12 weeks. |
| Cao et al., 2019 | China | SRMA | F and M; 6-17 years; Normal weight + overweight + obesity; other conditions: NR; n = 563. | 17 RCTs | to review existing evidence of the effects of HIIT versus MICT on improving CRF in healthy children and adolescents | School and clinical; | Children and adolescents | HIIT 6-24 WEEKS; MICT 6-24 WEEKS; |
| Cao et al., 2021 | China | SRMA | F and M; 10-16 years; Overweight + obesity; other conditions: NR. N= 325. | 12 RCTs | to conduct a comparative study on the effects of HIIT and MICT on cardiometabolic risk factors (such as body composition, CRF, blood pressure and glycolipid metabolism) in children and adolescents with obesity based on RCT studies. | School and lab | Children and adolescents | HIIT 6-24 weeks; MICT: moderate-intensity continuous training 6-24 weeks; |
| Casas et al., 2018 | Argentina | SRMA | F and M; 6-18y; Normal weight + overweight/obesity; Other health condition= No; n= 1135 | 22 RCTs and non-RCTs | To evaluate the effectiveness of resistance training on the body fat percentage and motor skill performance in children and adolescents who do not participate in professional competitive sports | NR | Children and adolescents | PA (resistance exercises); 1 - 3 days/wk, 10 – 40’/day, 50-100% 1RM, 6 to 60 weeks |

| Study | Country | Design | Participants | Included studies (n) | Objective | Setting | Who receive the intervention | Intervention(s) |
|---|---|---|---|---|---|---|---|---|
| Cheng et al., 2022 | China | SRMA | F and M; 7-17 years; Overweight + Obesity; Other conditions: NR; n=661 | 26 RCTs | Thus, we aimed to evaluate the effects of exercise training intervenations on the arterial stiffness and endothelial function of obese children; PWV, IMT, and FMD were the outcomes. Subgroup analyses were conducted to assess the effects of exercise training type and frequency, the age, and BMI on arterial stifness in obese children. | NR | Children and adolescents | Whole body physically active games 8 weeks; Aerobics 8-32 weeks; Combined 8-24 weeks; Aerobics + sports 12-24 weeks; Strength 12-24 weeks; Dance revolution 12 weeks; Multiple sports 12 weeks; HIIT 12 weeks;Control NR. |
| Choe et al., 2022 | United States of America | SRMA | F and M; 4-16 Years; Normal weight + overweight + obesity; Other condition: NR; n = 768. | 19 RCTs | TO address this gap in the literature by quantitatively investigating the effect of obesity interventions for Korean children and adolescents. | School | Children and adolescents | Physical Activity + Diet 5-16 years; Physical Education + Health Education 12 weeks; Exercise 12-16 weeks. |
| Clemente et al., 2022 | Portugal | SR | F and M; 8,9-18,2; Normal weight + Obesity; other conditions: metabolic syndrome; n = NR. | 17 RCTs | The purpose of this systematic review was to analyze physical fitness and health-related markers in untrained children and youths exposed to recreational soccer (RS). The study mainly focused on comparisons with the control groups, aiming to understand the isolated effects on body | NR | Children and adolescents | Soccer 8-40/43 weeks. Training duration varied between 12 and 90 min. |

| Study | Country | Design | Participants | Included studies (n) | Objective | Setting | Who receive the intervention | Intervention(s) |
|---|---|---|---|---|---|---|---|---|
| | | | | | composition, health-related outcomes, and physical fitness. | | | |
| Collins et al., 2018 | United Kingdom | SRMA | F and M; 8-16y; Normal weight + overweight/obesity; Other health condition= NR; n= 1153 | 18 RCTs and non-RCTs | The purpose of this meta-analysis was to examine the effect of resistance training interventions on weight status in youth | School; Community | Children and adolescents | PA (resistance exercises); 2 – 6 days/wk, 10 – 90'/day, 50 – 100% 1RM, 6 to 24 weeks |
| Comeras-Chueca et al., 2021 | Spain | SRMA | F and M; 6-18 years; Overweight + Obesity; Other conditions: No; n = 1,232 | 15 RCTs and n-RCTs | to summarize and critically appraise the existing research on the effects of AVGs on health-related physical fitness and motor competence in children and adolescents with overweight and obesity and to extract conclusions from a fair comparison of the studies included. | NR | Children and adolescents | Active Video Games: 8-24 weeks; Xbox 360 with Kinect, Nintendo Wii, Sony PlayStation 2, dance mats, and interactive video game cycling. Games included Just Dance, Wii Fit, Wii Sports, Kinect Adventures, Kinect Sport, Dance Central, Dance Dance Revolution (DDR), and EyeToy. |

| Study | Country | Design | Participants | Included studies (n) | Objective | Setting | Who receive the intervention | Intervention(s) |
|---|---|---|---|---|---|---|---|---|
| Costa et al., 2020 | Brazil | SRMA | F and M; 7-17 years; Normal weight + overweight + obesity; Other conditions: NR; n = 698; | 18 RCTs | The aim of the present study was to conduct a systematic review and meta-analysis evaluating the effects of super- vised exercise training on TC and LDL adaptations in children and adolescents compared to a control group. Additionally, we aimed to know the possible moderators of exercise effect in this population on TC and LDL levels. | NR | Children and adolescents | Control 8-12 weeks; HIIT 12 weeks; Endurance, mixed practices and strength 10-24 weeks; Rope interval 6-8 weeks; Endurance bike: 8 weeks; Mixed Sports and Activities 10 weeks; Combined 16 weeks and others. |
| Delgado-Floody et al., 2019 | Chile | SR | F and M; 9-16 years; Overweight/Obesity; Other Health conditions = NR. n = 136. | 6 RCTs | The aim of this systematic review was to critically analyze the feasibility of including HIIT-based programs in PE classes to improve the body compositions and cardiorespiratory fitness of overweight and obese students. | NR | Children and adolescents | HIIE, LIIE, CON, MIIT, P+ HIIT, HIIT. 6 Weeks; 6-26 weeks, 12 weeks. |
| Deshira et al., 2022 | United States of America | SR | F and M; 6-15 years; Overweight/Obesity; Other conditions = NR. n = NR. | 15 RCTs and non RCTs | To present a comprehensive systematic review of the effects of park-based inter- ventions on health outcomes among youth, defined as children and adolescents. | Park and Camp | Children and adolescents | Fit2Play; Health lifestyle fitness. 6-43 WEEKS. |

| Study | Country | Design | Participants | Included studies (n) | Objective | Setting | Who receive the intervention | Intervention(s) |
|---|---|---|---|---|---|---|---|---|
| Dias et al., 2015 | Australia | SRMA | F and M; 7-15.8y; Overweight/obesity; Other health condition= NR; n= 219 | 6 RCTs | To summarize the effect of exercise training on flow-mediated dilation in overweight and obese children and adolescents as well as investigate the role of cardiorespiratory fitness on effects observed | NR | Children and adolescents | PA (combined exercise): 2 – 5 days/wk, 30 – 75'/day, 50 – 80% VO2peak and 55% to 70% 1RM (only one study specified the intensity of the resistance exercises), 6 to 12 weeks |
| Dias et al., 2021 | Brazil | SRMA | F and M; 5-18 years; Obesity; Other condition: NR. N = 398. | 19 RCTS | the study objective here was to evaluate the influence of chronic exposure to exercise on the HRV of children and adolescents affected by obesity and to specify exercise as an enhancer for combating potential autonomic dysfunctions caused by obesity. | NR | Children and adolescents | Resistance training 12 weeks; Sport 12 weeks. |
| Dos Santos et al., 2021 | Brazil | SR | F and M; 6-19 years; Overweight + Obesity; Other condition = No; n = 335 | 37 RCTs and n-RCTs | to verify the amount of time spent at MVPA (primary outcome), by children and adolescents, in two different situations: 1) During dance classes, and 2) Before and after dance interventions | NR | Children and adolescents | Dance: 16 weeks - 2 years; African jazz, hip-hop, step, rumba, salsa, ballet, cheerleading, modern, line dance. |

| Study | Country | Design | Participants | Included studies (n) | Objective | Setting | Who receive the intervention | Intervention(s) |
|---|---|---|---|---|---|---|---|---|
| Eberl et al., 2019 | Germany | SR | F and M; 8-12 years; Normal weight + overweight; Other conditions = NO. n = NR. | 17 RCT and 3 non RCT | Collate the empirical evidence on the health effects of recreational football training as an intervention in healthy people of any age with regard to the prevention of obesity, hypertension, diabetes mellitus, and cardiovascular disease. | NR | Children and adolescents | Football group 13-26 weeks; NUT, Nutrition intervention group NR; Control 13-26 weeks. |
| Eckstein et al., 2022 | Germany | SR | F and M; 10-15.2 years; Normal weight + obesity; Other conditions: DM1, metabolic disease. N = 472. | 11 RCTs | The aim of this systematic review was to investigate how individuals with metabolic diseases respond to combat sports and if they are feasible, safe, and applicable. | After school activity program at an indoor gym; Training session were conducted at the university and PE at school; hospital setting. | Children and adolescents | Taekwondo for 16 weeks; judo 60min judo sessions; kung fu 24 weeks; tai chi 24 weeks. |
| Estéves-González et al., 2022 | Spain | SRMA | F and M; Age: NR; Obesity; Other conditions; Diabetes Mellitus 1; n = 252. | 10 RCTS | Consequently, the present study aimed to perform a systematic review and quantitative synthesis of all available studies that reported the effects of physical training programs on HRV, as a measure of sympathovagal balance, in children and adolescents with chronic diseases. | NR | Children and adolescents | Aerobic training: 6-72weeks; Resistance training 12 weeks; Combined aerobic and resistance training 6 weeks. |

| Study | Country | Design | Participants | Included studies (n) | Objective | Setting | Who receive the intervention | Intervention(s) |
|---|---|---|---|---|---|---|---|---|
| Feng et al., 2017 | China | SRMA | F and M; 6-19y; Normal weight + overweight/obesity; Other health condition= NR; n= 72620 | 76 RCTs and non-RCTs | To assess the effectiveness of school-based interventions for childhood obesity conducted in mainland China | School | Children and adolescents | PA (increased PA intensity and duration within and out of school); Educational (lessons, workshops, materials about health at school); Diet (modification of school lunches); Others (weight management/ school policies / psychological counseling / infrastructure support); 3 months to 4 years |
| Frerichs et al., 2016 | United States of America | SR | F and M; 10-18y; Normal weight + overweight/obesity; Other health condition= NR; n=14278 | 18 RCTs or quasi-experimental | To identify child and youth participatory interventions in the peer-reviewed literature in order to characterize the approaches and examine their impact on obesity and obesity-related lifestyle behaviors | School; Home; Community | Children and adolescents | Educational (PA and diet educational material; environmental and behavioral changes); 5 weeks to 3 years |
| García-Hermoso et al., 2014 | Spain | SRMA | F and M; 6-18y; Overweight/obesity; Other health | 9 RCTs | To examine the evidence for the effectiveness of aerobic exercise interventions on reducing insulin | NR | Children and adolescents | PA (sports; walking/running; ergometer-based; jump rope); 3 – 6 |

| Study | Country | Design | Participants | Included studies (n) | Objective | Setting | Who receive the intervention | Intervention(s) |
|---|---|---|---|---|---|---|---|---|
| | | | condition = NR; n=367 | | resistance markers in obese children and/or adolescents | | | days/wk, 30 – 90/day, 66-75% VO2max, 6 to 36 weeks |
| García-Hermoso et al., 2015 | Chile | SRMA | F and M; 6-18y; Overweight/obesity; Other health condition= NR; n=365 | 9 RCTs | To determine the effectiveness of aerobic plus resistance exercise interventions on body composition related to variables in overweight and obese youth | NR | Children and adolescents | PA (aerobic and combined exercises; sports); 2 – 4 days/wk, 45 – 90'/day, 55-65% VO2max, 8 to 24 weeks |
| García-Hermoso et al., 2016-a | Chile | SRMA | F and M; 6-17y; Overweight/obesity; Other health condition= NR; n=274 | 9 RCTs | To determine the effectiveness of high intensity interval training interventions on cardio-metabolic risk factors and aerobic capacity in overweight and obese youth, in comparison with other forms of exercise | NR | Children and adolescents | PA (aerobic exercises); 2 – 4 days/wk, 18 – 45'/day, 80 – 100% VO2max, 4 to 24 weeks |
| García-Hermoso et al., 2016-b | Chile | SRMA | F and M; 11-18y; Overweight/obesity; Other health condition= NR; n=555 | 12 RCT and non-RCT | To determine if the combination of aerobic and resistance exercise is superior to aerobic exercise alone for the health of obese children and adolescents | NR | Children and adolescents | PA (aerobic, resistance and combined exercises); 2-3 days/wk, 20 - 60'/day, 10 to 48 weeks |
| García-Hermoso et al., 2016-c | Chile | SRMA | F and M; 6.7 - 17.1y; Overweight/obesity; Other health condition= NR; n=427 | 9 RCTs | To examine the evidence for the effectiveness of physical exercise interventions on modifying the levels of serum C-reactive protein in children and | NR | Children and adolescents | PA (resistance and aerobic exercises; sports); 2 – 6 days/wk, 20 – 90'/day, 55 - 85% VO2max, 6 to 24 weeks |

| Study | Country | Design | Participants | Included studies (n) | Objective | Setting | Who receive the intervention | Intervention(s) |
|---|---|---|---|---|---|---|---|---|
| | | | | | adolescents with excess of weight | | | |
| García-Hermoso et al., 2017-a | Chile | SRMA | F and M; 6-17y; Overweight/obesity; Other health condition= NR; n=347 | 14 RCTs | To determine the effectiveness of exercise interventions on adipokines in pediatric obesity | NR | Children and adolescents | PA (combined exercises); 2 – 5 days/wk, 10-90'/day; 60-110% VO2peak, 6 to 24 weeks |
| García-Hermoso et al., 2017-b | Chile | SRMA | F and M; 6-18y; Overweight/obesity; Other health condition = NR; n=303 | 6 RCTs | To expand on the current body of literature by providing a quantitative estimate of the change in carotid intima-media thickness following exercise training as well as to provide an exploratory analysis of potential moderators associated with the variation in response to an exercise training intervention in overweight and obese youth | NR | Children and adolescents | PA (aerobic, resistance and combined exercises); 3 days/wk, 60–70'/day, 60–75% VO2peak and 60% 1RM, 14 to 24 weeks |
| Garcia-Hermoso et al., 2020-b | Spain | SRMA | F and m; 7-18 years; overweight + obesity; other conditions: NR; N = 1790. | 23 RCTs | to determine the minimum change in cardiorespiratory fitness (CRF) required to reduce adiposity (percent body fat) in exercise programs for overweight and obese youth. percent body fat in exercise programs in overweight and obese children and adolescents. | NR | Children and adolescents | Aerobic 8-36 weeks; Strength 12-22 weeks; Concurrent 10-22 weeks; |

| Study | Country | Design | Participants | Included studies (n) | Objective | Setting | Who receive the intervention | Intervention(s) |
|---|---|---|---|---|---|---|---|---|
| González-Ruiz et al., 2017 | Chile | SRMA | F and M; 6-18y; Overweight/obesity; Other health condition= NR; n= 1231 | 14 RCTs and non-RCTs | To determine the effectiveness of exercise interventions on abdominal fat and liver enzymes in overweight and obese youth | NR | Children and adolescents | PA (aerobic and resistance exercises); PA + Educational (nutrition/behavior education); 2- 5 days/wk, 20’–60’/day, 10 to 96 weeks |
| Guerra et al., 2013 | Brazil | SRMA | F and M; 6-16y; Normal weight + overweight/obesity; Other health condition= NR; n= NR | 12 RCTs | To evaluate the effectiveness of interventions that promote physical activity in the school environment and reduced overweight and obesity in children and adolescents | School | Children and adolescents | PA (sports; multiple exercises): 75 - 270’/wk, moderate intensity; Educational (PA and nutrition education); 2 weeks to 48 months |
| Han et al., 2019 | China | SRMA | F and M; 7-17 years; overweight + obesity; other conditions: NR; n = 623. | 11 RCTs | to conduct a meta-analysis of the studies on the role PA plays in modulating critical proinflammatory cytokine levels associated with overweight or obesity in children and adolescents to explore the effectiveness of exercise intervention within this population. | NR | Children and adolescents | Physical Activity (soccer, aerobic training, resistance training, combined training, after-school exercise, dance, jump roping, stationary cycling) 5-26 weeks. |
| Hejazi et al., 2022 | Iran | SRMA | F and M; 10,5-16,6y; Overweight/Obesity; Other Health conditions = NR. n = 468. | 14 RCTs | To examine the effects of PE, including different exercise methods, on adiponectin, insulin resistance indices, and | NR | Children and adolescents | Combined training; aerobic training; Interval training, P+HIIT. |

| Study | Country | Design | Participants | Included studies (n) | Objective | Setting | Who receive the intervention | Intervention(s) |
|---|---|---|---|---|---|---|---|---|
| | | | | | inflammatory factors in obese and overweight children. | | | 12 weeks; 8-39 weeks. |
| Hernandez -Martin et al., 2021 | Spain | SRMA | F and M; 9-12 years; Normal weight + obesity; Other condition = NR; n = 1643. | 14 RCT | Therefore, the aim of this systematic review and meta-analysis was to investigate and analyze the effects of football practice on body composition—fat-free mass, fat mass, and bone mineral content (BMC)—in children. | NR | Children and adolescents | Control 20-24 weeks; Football Training 12-24 weeks; |
| Jago et al., 2004 | United States of America | SR | F and M; 5-18y; Normal weight + overweight/obesity; Other health condition= NR; n= 919 | 9 RCTs and non-RCTs | To examine the effectiveness of non curricular interventions for promoting physical activity | School; Camp; Clubs | Children and adolescents | PA (increased PA during school breaks, travel to school and others extracurricular time): 4 weeks to 5 months |
| Jung et al., 2018 | United States of America | SRMA | F and M; 6 – 19 years; Obesity; other conditions: NAFDL, metabolic Syndrome; n = nr. | 12 RCTs and 22 non-RCTs. | this meta-analysis was aimed to examine the overall effects of exercise intervention on visceral fat in obese children and adolescents, and to propose the most effective exercise regimens to reduce visceral fat. | NR | Children and adolescents | Aerobic training 12-52 weeks; Resistance Training 12-21 weeks; |

| Study | Country | Design | Participants | Included studies (n) | Objective | Setting | Who receive the intervention | Intervention(s) |
|---|---|---|---|---|---|---|---|---|
| Jurado-Castro et al., 2020 | Spain | SRMA | F and M; 9 ± 1.57 years; Obesity; Other condition: NR. N = 478. | 10 RCTs | the aim of the present study is to evaluate the effect of current interventions with a PA component on body mass index (BMI) and time spent engaged in MVPA via accelerometry measurement, among school-age children with obesity to establish better practices that promote adherence to a more active lifestyle. | NR | Children and adolescents | Multicomponent intervention 12-104 weeks; Active video game 24 weeks; |
| Kelley and Kelley., 2008 | United States of America | SRMA | F and M; 8-16y; Normal weight + overweight/obesity; Other health condition= type 1 diabetes; n= 404 | 12 RCTs | To examine the effects of aerobic exercise on non–high-density lipoprotein cholesterol in children and adolescents | NR | Children and adolescents | PA (aerobic exercises): 3 – 5 days/wk, 20 – 135'/day, 44 - 90% VO2max, 5 to 16 weeks |
| Kelley et al., 2019 | United States of America | SRMA | F and M; 2-18years; Obesity and Overweight Other conditions: NR; N = 2792. | 57 RCTS | Determine both the effects and hierarchy of effectiveness for exercise interventions (aerobic, strength training or both) on selected measures of adiposity (body mass index (BMI) in kg/m2 , fat mass and per cent body fat) in overweight and obese children and adolescents. | NR | Children and adolescents | Aerobic exercises 6-36 weeks; Strength training 6-36 weeks. Combined aerobic and strength 6-36 weeks. |

| Study | Country | Design | Participants | Included studies (n) | Objective | Setting | Who receive the intervention | Intervention(s) |
|---|---|---|---|---|---|---|---|---|
| Kelley et al., 2021 | United States of America | SRMA | F and M; 7-19 years; Obesity; Other condition: hyperlipidemia, hypertension or prehypertension, hyperinsulinemia and insulin resistance; n =NR | 39 RCTs | To examine the effects of exercise on cardiovascular disease (CVD) risk factors in children and adolescents with obesity. | Schools | Children and adolescents | Aerobic, strenght training, both and usual care 6-24 weeks. |
| Kim et al., 2017 | United States of America | SRMA | F and M; 8-16y; Normal weight + overweight/obesity; Other health condition= NR; n=1777 | 18 RCTs and non-RCTs | To evaluate the overall impact of sport-based interventions on body weight among children and adolescents | NR | Children and adolescents | PA (recreational sports, martial arts, aerobic and resistance exercises); PA + Diet (diet prescription); 4 weeks to 2 years |
| Laframbois e et al., 2011 | Canada | SR | F and M; 6.5-16y; Normal weight + overweight/obesity; Other health condition= type 1 diabetes; n=2521 | 10 RCTs | To determine the quality of current evidence forming the relationship between aerobic physical activity and adiposity changes in school-aged children and youth | NR | Children and adolescents | PA (aerobic exercises); 1 – 5 days/wk, 30 – 90’/day, 50-90% FCmáx, 8 weeks to 8 months |
| Lee et al., 2020 | Republic of Korea | SRMA | F and M; 7-18 years; Obesity and overweight; Other conditions: NR; n = NR. | 27 RCTS | Therefore, the purpose of this metanalysis was to investigate the effects of exercise interventions on childhood and adolescence overweight and obesity depending on exercise type. | NR | Children and adolescents | Aerobic exercise 8-54weeks; Resistance exercise 22-24 weeks; Combined aerobic training 8-22weeks. |

| Study | Country | Design | Participants | Included studies (n) | Objective | Setting | Who receive the intervention | Intervention(s) |
|---|---|---|---|---|---|---|---|---|
| Li and Chen et al., 2021 | China | SRMA | F and M; 8-17 years; Obesity; other condition = no. n = 704. | 19 RCTs | (1) To explore the effect of different exercise modalities in the treatment of CRF in obese adolescents with sedentary behavior (sub-analysis with different exercise modalities and intervention duration. (2) To explore the optimal exercise modality for obese adolescents with different abnormal physiological parameters and provide reasonable exercise prescription. | NR | Children and adolescents | MIIT for 12-20 weeks; Continuous aerobic 10-12 weeks; HIIT 8-12 weeks; Combined (aerobic + strength) 8-12 weeks; Strength 8 weeks; Yoga 8 weeks; Soccer 12 weeks; Rope jumping 12 weeks; Control 8-20 weeks. |
| Li et al., 2022 | China | SRMA | F and M; 6-18 years; Overweight + obesity; Other conditions: NR; n = 915. | 8 RCTS | to evaluate the effectiveness of lifestyle intervention for the treatment of overweight/obesity in children in China | NR | Children and adolescents | Diet + physical activity 6-52 weeks; Physical activity + health education 8-24 weeks; Physical activity 12 weeks; Control 6-52 weeks. |
| Littlewood et al., 2020 | Australia | SR | F and M; 2-17 years; Obesity; Other conditions: NR; n = 6629 | 6 RCTS | The aim of this review is to identify and evaluate global studies reporting interventions to prevent or treat childhood and/or adolescent overweight/obesity in the unique population of Māori and Pacific Islander peoples. | Home Environment | Children and adolescents | Switch for 20 weeks. |

| Study | Country | Design | Participants | Included studies (n) | Objective | Setting | Who receive the intervention | Intervention(s) |
|---|---|---|---|---|---|---|---|---|
| Liu et al., 2020 | China | SRMA | F and M; 10-16 years; Obesity; Other conditions: NR; n = 309 | 9 RCTs | to compare the effectiveness of HIIT and MICT in reducing the abovementioned cardiometabolic risk factors and determine which HIIT modality is effective and time-efficient in managing the abovementioned risks. | NR | Children and adolescents | HIIT: 4-12 weeks. MICT: 4-12 weeks |
| Liu et al., 2024 | United Kingdom | SRMA | F and M; 5-18 years; Overweight + Obesity + Normal weight; Other conditions: No; n = 577 | 77 RCTs | to examine the extent of process evaluation reporting in school-based HIIT studies. | School | Children and adolescents | HIIT: 6-12 weeks; Running, games, resistance training, sprint cycling. |
| Lopes et al., 2019 | Brazil | SRMA | F and M; 7-19 years; Normal weight + overweight + obesity; Other conditions = NR; n = 1199. | 25 RCTs | Therefore, the purpose of this study was to systematically review the literature published during the last 10 years that evaluated the effect of different modalities of physical exercises on HDL-C concentrations in children and adolescents. | NR | Children and adolescents | Control group + nutritional Guidance 12 weeks; Control group 12 weeks; Continuous aerobic 48 weeks; HIIT + nutritional 12 weeks; Continuous Aerobic 12 weeks; Nutritional + exercise 12 weeks and others. |
| Mears and Jago., 2016 | United Kingdom | SRMA | F and M; 5-18y; Normal weight + overweight/obesity; Other health | 15 RCTs and non-RCTs | To examine the effectiveness of after-school interventions at increasing MVPA levels in children and adolescents | School; Home | Children and adolescents | PA (general sports; games; jumping rope; dance; aerobic exercises; active commuting); PA + |

| Study | Country | Design | Participants | Included studies (n) | Objective | Setting | Who receive the intervention | Intervention(s) |
|---|---|---|---|---|---|---|---|---|
| | | | conditions= NR; n= 3547 | | | | | Diet; 2 - 7 days/wk, 60 – 135’/day |
| Mei et al., 2016 | China | SRMA | F and M; 6-18y; Normal weight + overweight/obesity; Other health conditions= NR; n= 22381 | 18 RCTs | Understand whether long-term school-based randomized clinical trials PA interventions could benefit primary school children’s growth | School | Children and adolescents | PA (increased PA level in and outside school); PA + Educational (health education); 2–5 days/wk, 20 – 100’/wk, 1 month to 6 years |
| Méndez-Hernández et al., 2022 | Mexico | SRMA | F and M; 5-19years; Obesity and overweight; other conditions: NR; n= 1834. | 27 RCTS | The objective of this study was to perform a systematic meta-analysis to determine the effect of strength training on body fat in children and/or adolescents with overweight or obesity. | Only home-based reported | Children and adolescents | Strength training 5-52 weeks. |
| Menjie et al., 2022 | China | SRMA | F and M; 5-19 years; Obesity Other conditions – NR; n = 2995. | 47 RCTs | Given the above, this study will systematically evaluate the effectiveness and safety of HIIT for children and adolescents, expecting to provide a scientific basis for the promotion of HIIT in children and adolescents. | NR | Children and adolescents | Control with diet 12 weeks; Control without physical activity 12-16 weeks; Control or usual training/activities 8-48 weeks; Aerobics 12-16 weeks; Diet + HIIT 12 weeks; HIIT 12 weeks; Equipment exercise 8 weeks; Judo + aerobics 12 |

| Study | Country | Design | Participants | Included studies (n) | Objective | Setting | Who receive the intervention | Intervention(s) |
|---|---|---|---|---|---|---|---|---|
| | | | | | | | | weeks; Aerobic training nr; Plyometrics 12 weeks; Physical Activity 6 weeks and others. |
| Millard-Staffor et al., 2013 | United States of America | SR | F and M; 8.9 - 16.9y; Overweight/obesity; Other health condition= NR; n= 1578 | 33 RCTs | To systematically examine and summarize: a decade of peer-reviewed literature describing the association between selected components of physical fitness (i.e., aerobic capacity, muscular strength and muscular endurance) on health in school-aged youth who are overweight and obese; and whether improvements in fitness are associated with beneficial changes in measures of chronic disease including adiposity, cardiovascular or metabolic health risk factors | NR | Children and adolescents | PA (sports; games; tai chi chuan; yoga; aerobic, resistance and combined exercises); PA + Diet (restrictions); PA + Educational (lifestyle related to nutrition, physical activity or behavior change); PA + Diet + Educational; 80 – 420'/wk, 6 to 36 weeks |
| Nagle et al., 2013 | United States of America | SR | F and M; 9.9-16.9y; Overweight/obesity; Other health condition= NR; n= NR | 4 RCTs and pre-post test studies | To identify evidence-based strategies associated with effective healthcare interventions for the prevention and treatment of childhood obesity in Latin America | Health clinics; Medical offices | Children and adolescents | Educational (multidisciplinary intervention consisting of therapy sessions and counseling about diet and PA); Educational + PA (aerobic and |

| Study | Country | Design | Participants | Included studies (n) | Objective | Setting | Who receive the intervention | Intervention(s) |
|---|---|---|---|---|---|---|---|---|
| | | | | | | | | recreational exercises); Diet (individualized diets); Diet + PA; 3 to 12 months |
| Oliveira et al., 2017 | Portugal | SRMA | F and M; 8-17y; Overweight/obesity; Other health conditions = NR; n= 141 | 6 RCTs and non-RCTs | Systematically summarize experimental and quasi-experimental studies which reported the effects of group sports on health-related physical fitness of overweight/obese youth when compared to absence of an intervention or to standard interventions | NR | Children and adolescents | PA (sports); 2 – 4 days/wk, 45 - 90 min/day, 50 - 80% FCMax, 3 to 6 months |
| Paes et al., 2015 | Brazil | SR | F and M; 8-17y; Normal weight + overweight/obesity; Other health conditions= NR; n= NR | 39 RCTs | To review the current literature regarding the effects of exercise on several metabolic variables of childhood obesity | NR | Children and adolescents | PA (aerobic, resistance, combined and recreational activities); 4 to 13 weeks |
| Parikh and Stratton et al., 2011 | United Kingdom | SR | F and M; 5-18y; Normal weight + overweight/obesity; Other health conditions= NR; n= 18210 | 25 RCTs and pre-post test studies | To evaluate and highlight the effect of intensity on fatness and cardiorespiratory fitness | NR | Children and adolescents | PA (aerobic exercises); medium to high intensity, 7 weeks to 10 months |

| Study | Country | Design | Participants | Included studies (n) | Objective | Setting | Who receive the intervention | Intervention(s) |
|---|---|---|---|---|---|---|---|---|
| Recchia et al., 2023 | China | SRMA | F and M; 8-19 years and 9-15 years(obesity group); Other conditions = diabetes; n = 2441. | 17 RCTs 4 NRCTs. | With this systematic review and meta-analysis, we aimed to gather a wider range of studies to provide critical insight into the association between physical activity interventions and de- pressive symptoms in children and adolescents. Further- more, we aimed to identify possible participant- and trial- related characteristics that might moderate the overall treatment effect. | NR | Children and adolescents | Aerobic training for 4-144 weeks. |
| Schranz et al., 2013 | Australia | SRMA | F and M; 6-18y; Overweight/obesity; Other health conditions= NR; n= NR | 40 RCTs, non-RCTs and uncontrolled trials | To determine the effect of resistance training on the strength, body composition and psychosocial status of overweight and/or obese children and/or adolescents | NR | Children and adolescents | PA (resistance and combined exercises); PA + Educational (nutrition and exercise education); PA + Others (behavioral therapy); 3–5 days/wk, 50-92%1RM and 60-80% VO2ma, 6 to 52 weeks |
| Schwartz et al., 2017 | France | SRMA | F and M; 5-18y; Overweight/obesity; Other health conditions= NR; n= 549 | 16 RCTs and uncontrolled trials | To determine if structured physical activity interventions affect energy and macronutrient intake in obese and overweight children and adolescents | NR | Children and adolescents | PA (aerobic, resistance and combined exercises); 2 – 3 days/wk, 16 – 60'/day, 10 weeks to 12 months |

| Study | Country | Design | Participants | Included studies (n) | Objective | Setting | Who receive the intervention | Intervention(s) |
|---|---|---|---|---|---|---|---|---|
| Sirico et al., 2018 | Italy | SRMA | F and M; 18y or younger; Overweight/obesity; Other health conditions= NR; n= 250 | 7 RCTs | To investigate the effectiveness of physical exercise in reducing the systemic inflammation related to obesity in children | NR | Children and adolescents | PA (aerobic and combined exercises); 3 days/wk, 20 – 60'/day; 6 to 20 weeks |
| Steele et al., 2021 | United Kingdom | SRMA | F and M; 7 – 16 yeas; Normal weight + obesity. Other conditions: NR; N = NR. | 54 RCTs | Purpose: To conduct a systematic review and multilevel meta-analysis of the current literature as to the effects of interval training (IT) vs moderate intensity continuous training (MICT) on measures of body composition, both on a whole-body and regional level. | NR | Children and adolescents | MICT = moderate intensity continuous training 4-12 weeks; IT = interval training 4-12 weeks; |
| Thivel et al., 2018 | France | SRMA | F and M; 8-18years; Obesity and Overweight; Other conditions: NR. N = NR. | 15 RCTS | The primary aim of the present article was thus to determine from a systematic review and meta-analysis whether HIIT improves body composition and aerobic fitness, and decreases cardiovascular and metabolic risk parameters in overweight and obese children and adolescents. | NR | Children and adolescents | HIIT 4-24 weeks; |
| Valeriani et al., 2021 | Italy | SR | F and M; 0-18 years; Obesity and overweight; other | 10 RCTs | Therefore, to clarify the possible role of exergames in improving weight status among | Home-based Scholl-based clinics | Children and adolescents | Exergames 6-40weeks; |

| Study | Country | Design | Participants | Included studies (n) | Objective | Setting | Who receive the intervention | Intervention(s) |
|---|---|---|---|---|---|---|---|---|
| | | | conditions: NR; n = NR. | | overweight/obese children and adolescents, | | | |
| van Wijnen et al., 2009 | Netherlands | SR | F and M; 5-18y; Overweight/obesity; Other health conditions = NR; n= 2502 | 7 RCTs and non-RCTs | To gain insight in the extent to which psychosocial effects of obesity prevention programmes have been studied, to give an overview of the methods used to measure the particular psychosocial aspects and – if possible – to quantify the effects found | School | Children and adolescents | Educational (behavioral change about diet and PA level); Diet (change in snacks); PA (resistance exercises); 16 weeks to 3 years |
| Vissers et al., 2016 | Belgium | SRMA | F and M; 7-19y; Overweight/obesity; Other health conditions = NAFLD; n= 782 | 13 RCTs and non-RCTs | To evaluate the effect of supervised lifestyle interventions (diet, exercise, or a combination of both) on visceral adipose tissue in overweight and obese children and adolescents | NR | Children and adolescents | Diet (food/calorie restriction); PA (aerobic, resistance and combined exercises):55 – 75% VO2peak, 60% 1RM, 120 – 200'/wk; Diet + PA; 3 months to 1 year |
| Voskuil et al., 2017 | United States of America | SR | F; 6-18y; Normal weight + overweight/obesity; Other health conditions= NR; n= 14120 | 15 RCTs | To evaluate the evidence for PA intervention effects on accelerometer-measured PA, body mass index, and percent body fat among girls | NR | Children and adolescents | Educational (PA and/or diet counseling/information); PA (exercise or sports); Educational + PA; 2 – 5 days/wk, 20 – 90'/day, 12 weeks to 2 years |

| Study | Country | Design | Participants | Included studies (n) | Objective | Setting | Who receive the intervention | Intervention(s) |
|---|---|---|---|---|---|---|---|---|
| Wang et al., 2022 | China | SRMA | F and M; 6-18 years; Overweight + obesity; Other conditions: NR; n = 1134. | 15 RCTs | To evaluate and compare the effect of exercise interventions (AE, RE, CE, and HIIT) on visceral fat in overweight or obese young individuals | NR | Children and adolescents | Aerobic exercise 12-43 years; Resistance Exercise 12-43; Combined Exercise (aerobic + resistance) 12-43 years; HIIT 12 weeks; |
| Whitlock et al., 2010 | United States of America | SRMA | F and M; 4-18y; Overweight/obesity; Other health conditions= NR; n= 2807 | 15 RCTs and non-RCTs | To examine the benefits and harms of behavioral and pharmacologic weight-management interventions for overweight and obese children and adolescents | Health primary care | Children and adolescents | Educational (PA and/or diet counseling/information); Pharmacological (drug administration); Educational + Pharmacological; 24 weeks to 1 year |
| Xu et al., 2022 | China | SRMA | F and M; 9.3-17 years; Overweight/Obesity; Other conditions = NO. n = 2.599. | 46 RCTs | To evaluate the overall impact of the effect of different doses of exercise on weight loss in overweight/obese children, and provide a scientific basis for the study of the quantity– effect relationship between exercise and weight loss. | NR | Children and adolescents | Aerobic training; Resistance training; Combined Training; HIIT training. 20-54 weeks; 20-50 weeks; 30-90 weeks; 6-52 weeks. |

| Study | Country | Design | Participants | Included studies (n) | Objective | Setting | Who receive the intervention | Intervention(s) |
|---|---|---|---|---|---|---|---|---|
| Yuksel et al., 2020 | Turkey | SR | F and M; 4-15 years; Normal weight + obesity. Other conditions: NR. n = 6000. | 19 RCTs | The purpose of this review is to systematically examine the possibility of school‑based interventions on promoting physical activity and physical fitness as well as preventing obesity | School | Children and adolescents | Lifestyle intervention for 1 year. Intervention 52-260 weeks;Child health weight 10 weeks; Multicomponent intervention 86 weeks. School based PA intervention 12-260 weeks; Comprehensive school health intervention 208 weeks. |
| Zouhal et al., 2020 | France | SR | F and M; 5-90 years; Obesity + overweight; Other conditions: NR; n = 692. | 116 RCTs | This systematic review aim summarizes recent findings on the effects of physical training on anthropometric characteristics (e.g., body weight, BF and body mass index [BMI]), physical performances (e.g., endurance, sprint, strength and power) and physiological capacities (e.g., oxygen uptake, heart rate and lactate threshold) in individuals with overweight and obesity. | NR | Children and adolescents | Aerobic endurance 8-56 weeks; HIIT: 5-21 weeks; Resistance and Strength training: 6-56 weeks; |

| Study | Country | Design | Participants | Included studies (n) | Objective | Setting | Who receive the intervention | Intervention(s) |
|---|---|---|---|---|---|---|---|---|
| Albornoz-Guerrero et al., 2021 | Chile | SR | F and M. 6-12 years; Overweight + obesity other conditions: NR. n = 4434. | 29 RCTs | is to analyze the characteristics of multicomponent interventions aimed at reducing overweight and obesity in children in territories with an extremely cold climate. | health centers, educational centers, university centers, community centers. | Children and parents | Physical activity and nutrition 26-52weeks; Education 26-52 weeks; Behavioral and education 26-52weeks; physical activity 26-52 weeks. Behavioral and nutrition 2-52 weeks; physical activity and behavioural 2-52 weeks; Physical activity and behavioral and education and nutrition 26-52 weeks; physical activity and behavioural and nutrition 26-52 weeks. |
| Andrade et al., 2018 | United States of America | SR | F and M; 6-12y; Overweight/Obesity; Other health conditions = NR; n = 10.365 | 6 RCTs and 4 non-RCT | To examine the frameworks used within school-based intervention programs that shower improvements in obesity-related outcomes (eg, improvement in dietary behaviors and anthropometric measurements) among Hispanic children in the United States and Mexico. | School | Children and parents | Nutritional education, Physical education, Physical activity, modified cafeteria offering, nutrition/lifestyle e/curriculum; 43 – 156 weeks; 96 weeks. |

| Study | Country | Design | Participants | Included studies (n) | Objective | Setting | Who receive the intervention | Intervention(s) |
|---|---|---|---|---|---|---|---|---|
| Braaksma et al., 2018 | Netherlands | SR | F and M; 6-12y; Normal weight + overweight/obesity; Other health condition= NR; n= 7071 | 23 RCTs | To examine the characteristics of physical activity interventions and the effects on cardiorespiratory fitness in healthy children based on treatment theory | School; Home; Research facility; Combination of school and community | Children and parents | PA (aerobic exercises);<br>1 – 5 days/wk; 40 – 80'/day, 80–100% of maximal aerobic speed or 50–80% VO2peak, 6 to 104 weeks |
| Cesa et al., 2014 | Brazil | SRMA | F and M; 6-12y; Normal weight + overweight/obesity; Other health condition= NR; n= 10748 | 11 RCTs | To assess the effects of physical activity interventions in preventing cardiovascular risk factors in childhood | School; Out of school | Children and parents | PA (increased time in PA physical education classes and/or extracurricular activities); 2 - 6 days/wk, 20 – 110'/day; 6 months to 3 years |
| Colquitt et al., 2016 | United Kingdom | SRMA | F and M; 0-6 years; Obesity + overweight; Other conditions: NR; n = 923. | 7 RCTs | To assess the effects of diet, physical activity, and behavioural interventions for the treatment of overweight or obesity in preschool children up to the age of 6 years. | Outpatient setting, primary care, community, home and obesity reasearch clinic | Children and parents | Nutritional + Physcial Activity + Behavioral components 26-156 weeks; usual care 16-156 weeks. |
| García-Hermoso et al., 2020-c | Spain | SRMA | F and M; 1-6 years; Overweight + Obesity + Normal weight; Other conditions: No; n = 7,843 | 19 RCTs | to evaluate the effects of exercise interventions on health-related physical fitness and blood pressure in preschoolers. | NR | Children and parents | PA internventions: 10-16 weeks; Active lifestyle, quick walking, slow running, jumping, rope skipping, |

| Study | Country | Design | Participants | Included studies (n) | Objective | Setting | Who receive the intervention | Intervention(s) |
|---|---|---|---|---|---|---|---|---|
| | | | | | | | | semi-squating and slow crawling. |
| Jull and Chen., 2013 | New Zealand | SRMA | F and M; 6-14y; Overweight/obesity; Other health conditions = NR; n=266 | 4 RCTs | To assess the effectiveness of weight loss interventions that compared a parent-only condition with a parent-child condition in overweight and obese children | University clinics; Community offices; Psychiatric outpatient clinic | Children and parents | Educational (PA; diet; behavioral skills): 10 – 120'/day, 10 weeks to 6 months |
| Mead et al., 2017 | United Kingdom | SRMA | F and M; 6-11 years; Obesity + overweight; Other conditions: asthma, type 2 diabetes, metabolic syndrome, depression, anxiety and fatty liver diagnoses ; n = 8461. | 70 RCTs | To assess the effects of diet, physical activity and behavioural interventions (behaviour‐changing interventions) for the treatment of overweight or obese children aged 6 to 11 years. | Primary care, secondary care, university reasearch clinics, community, home and schools | Children and parents | Nutritional + Physcial Activity + Behavioral components 2-104 weeks; usual care 2-104 weeks. |
| Medina-Blanco et al., 2011 | Mexico | SR | F and M; 5-12y; Normal weight + overweight/obesity; Other health conditions= NR; n= 1263 | 7 RCTs and non-RCTs | To assess physical activity promotion programs in school children from 6 to 12 years old | NR | Children and parents | Educational (lessons about healthy eating and increased PA); PA (increased intensity and time of PA); PA + educational; 2-3 days/wk, 45 – 180'/wk |

| Study | Country | Design | Participants | Included studies (n) | Objective | Setting | Who receive the intervention | Intervention(s) |
|---|---|---|---|---|---|---|---|---|
| Oosterhoff et al., 2016 | Netherlands | SRMA | F and M; 4-12y; Normal weight + overweight/obesity; Other health conditions= (pre)hypertension; n= 72934 | 85 RCTs | To review the current evidence on the impact of school-based lifestyle randomized control trials on children's BMI and blood pressure with the use of multilevel meta-analysis techniques | School | Children and parents | Educational (materials and/or lectures about dietary and activity behaviors); PA (increase of physical education load, reduction sedentary time) + Educational; Diet (changes in foods provided by school) + Educational; PA + Diet; PA+ Diet + Educational; 1.5 to 96 months |
| Sbruzzi et al., 2013 | Brazil | SRMA | F and M; 6-12y; Normal weight + overweight/obesity;Other health conditions= hypertension, dyslipidemia; n= 23617 | 26 RCTs | To systematically review educational interventions, including behavioral modification, nutrition and physical activity as compared to usual care or no intervention, for prevention or treatment of childhood obesity | NR | Children and parents | Educational (multicomponent interventions designed to promote behavior changes to prevent or treat obesity); 6 to 48 months |
| Zabatiero et al., 2018 | Australia | SRMA | F and M; 6-12 years; overweight + obesity; Other conditions: NR; n = 1252. | 9 RCTs | to assess the effectiveness of interventions aimed at reducing objectively-measured sedentary behaviour (i.e., measured by accelerometry or inclinometry) amongst people with overweight or obesity. | School | Children and parents | School-based intervention 20 weeks; Behavior + physical activity 34 weeks; Sedentary beharior + physical activity + diet 26 |

| Study | Country | Design | Participants | Included studies (n) | Objective | Setting | Who receive the intervention | Intervention(s) |
|---|---|---|---|---|---|---|---|---|
| | | | | | | | | weeks; control 2-=34 weeks. |
| Azevedo et al., 2023 | United Kingdom | SRMA | F and M; 3-16 years; Normal weight + overweight + obesity; Other condition = NR; n = 2.352. | 16 RCT | Therefore, this narrative systematic review and meta-analysis aimed to examine the effectiveness of e-health interventions for the treatment of overweight or obesity in children (0–17 years old). The secondary aim was to explore subgroup differences according to age, weight status, duration, behaviors targeted, income, and study quality on intervention outcomes and provide a narrative synthesis to gather knowledge of the components of e-health interventions to guide future research and implementation. | Home | Children, Adolescents and Parents | Control/ usual care 24-48 weeks; Educational contend/ counseling 24-48 weeks. |
| Brown et al., 2019 | United Kingdom | SRMA | F and M; 0-18 years; Obesity + overweight; Other conditions: No ; n = NR. | 153 RCTs | To determine the effectiveness of a range of interventions that include diet or physical activity components, or both, designed to prevent obesity in children. | Home, schools, childcare, healthcare, community | Children, Adolescents and Parents | Physical Activity + Nutritional components 52-104 weeks; usual care 52-104 weeks. |

| Study | Country | Design | Participants | Included studies (n) | Objective | Setting | Who receive the intervention | Intervention(s) |
|---|---|---|---|---|---|---|---|---|
| Dobbins et al., 2013 | Canada | SR | F and M; 6-18y; Normal weight + overweight/obesity; Other health condition= No; n=36593 | 44 RCTs | To summarize the evidence of the effectiveness of school-based interventions in promoting physical activity and fitness in children and adolescents | School; Home; Community | Children, Adolescents and Parents | Educational (material and counseling about self-esteem, nutrition, physical activity and motivational strategies); Educational + PA (increased PA in physical education classes and at home); 12 weeks to 6 years |
| Godin et al., 2015 | Canada | SR | F and M; Elementary and secondary school age; Normal weight + overweight /obesity; Other health condition= NR; n=NR | 15 RCTs and non-RCTs | To identify school-based programs that have been developed to prevent obesity and the determinants of obesity (physical activity and healthy eating) among First Nations, Inuit and Métis (FNIM) youth in Canada | Elementary school in rural and remote communities or reserves | Children, Adolescents and Parents | Educational (diet; diabetes knowledge; healthy eating; PA) |
| Gonzalez-Suarez et al., 2009 | Spain | SRMA | F and M; 11+ y; Normal weight + overweight/obesity; Other health condition = NR; n=NR | 22 RCTs and non-RCTs | To evaluate the evidence for the effectiveness of school-based programs in the management of childhood obesity in terms of BMI, percentage of body fat, waist girth, triceps skinfold, and waist–hip ratio | School | Children, Adolescents and Parents | Educational (lessons about changes in nutrition, PA and hours spent in TV viewing) + PA (increased time and intensity of physical education classes); 6 months to 2 years |

| Study | Country | Design | Participants | Included studies (n) | Objective | Setting | Who receive the intervention | Intervention(s) |
|---|---|---|---|---|---|---|---|---|
| Guerra et al., 2014 | Brazil | SRMA | F and M; 6-18y; Normal weight + overweight/obesity; Other health condition= No; n=35690 | 38 RCTs | To assess the effect of school based physical activity and nutritional education interventions by randomized controlled studies on the reduction of BMI among children and adolescents | School | Children, Adolescents and Parents | Educational (Diet and PA lessons/support; school food service changes); PA (dance classes; reduction screen time); Educational + PA; 8 weeks to 6 years |
| Hamel et al., 2011 | Ireland | SR | F and M; 8-18y; Normal weight + overweight/obesity; Other health condition= NR; n=6226 | 14 RCTs | To identify, retrieve and synthesize the existing scientific literature on using computer and web-based interventions to increase preadolescent and adolescent PA alone or in combination with a dietary intervention | School; Home; Camp/troop | Children, Adolescents and Parents | Educational (computer and web-based interventions to increase PA alone or in combination with a dietary intervention); 2 weeks to 2 years |
| Harris et al., 2009 | Canada | SRMA | F and M; 5-18y; Normal weight + overweight/obesity; Other health condition= NR; n= 18141 | 18 RCTs and non-RCTs | To determine whether school-based physical activity interventions improve children's body composition, as measured by BMI | School | Children, Adolescents and Parents | PA (increased PA and exercise levels); PA + Educational (nutrition or health education); 2 – 5 days/week, 10 – 90'/day, 6 months to 3 years |
| Ho et al., 2013 | Australia | SRMA | F and M; 5-18y; Overweight/obesity; Other health condition= NR; n= 879 | 15 RCTs | Systematically assess the effects of diet-only interventions compared with diet plus exercise interventions on both weight loss and metabolic risk reduction | School; Hospital; Community | Children, Adolescents and Parents | Diet (caloric/food restriction); PA (aerobic, resistance and combined |

| Study | Country | Design | Participants | Included studies (n) | Objective | Setting | Who receive the intervention | Intervention(s) |
|---|---|---|---|---|---|---|---|---|
| | | | | | | | | exercises); PA + Diet; 6 weeks to 6 months |
| Kemp et al., 2021 | United Kingdom | SR | F and M;9-16 years; Normal weight + overweight; other conditions = NR. n = 1727 | 7 RCTs | to evaluate the effectiveness of family-based eHealth interventions in CVD risk reduction. | Community-based setting; Usual Care; | Children, Adolescents and Parents | Family-based e-health 8-12 weeks; control 8-12weeks; |
| Kropski et al., 2008 | United States of America | SR | F and M; 4-14y; Normal weight + overweight/obesity; Other health conditions= NR; n=43130 | 14 RCTs and non-RCTs | To provide a focused evaluation of the quality and results of long-term school-based obesity prevention programs and to offer guidance for future investigations into this intervention modality | School; School and home | Children, Adolescents and Parents | Educational (lesson about reducing carbonated beverages and benefits of a healthy diet); PA (increase intensity in physical education lessons; reduction of screen time; family games); PA + Diet (reduction of energy from fat; changes in school lunches); 3 days/wk, 6 months to 4 years |
| Luckner et al., 2012 | United Kingdom | SRMA | F and M; 6-18y; Normal weight + overweight/obesity; Other health conditions= NR; n=41419 | 103 RCTs or non-RCTs | To evaluate interventions that promote healthy weight (defined as a reduction in BMI or %BF) in general populations (unselected by weight) from Western countries in a comprehensive metaanalysis | School; Out of school | Children, Adolescents and Parents | PA (reduction of screen time); Educational (health lifestyle); Diet; Educational + PA; Educational + Diet; |

| Study | Country | Design | Participants | Included studies (n) | Objective | Setting | Who receive the intervention | Intervention(s) |
|---|---|---|---|---|---|---|---|---|
| | | | | | | | | PA + Educational + Diet; 1 to 90 months |
| Mclean et al., 2003 | United Kingdom | SR | F and M; 6-16y; Overweight/obesity; Other health condition= type 2 diabetes; n=763 | 21 RCTs | To identify trials evaluating family involvement in weight control, weight maintenance and weight-loss interventions targeting food intake and/or physical activity | Home | Children, Adolescents and Parents | Others (family based interventions changing food intake and physical activity); 9 to 104 weeks |
| Sims et al., 2015 | United Kingdom | SRMA | F and M; 5-18y; Overweight/obesity; Other health conditions= NR; n= 8687 | 14 RCTs and non-RCTs | To explore the effect of interventions on maintained whole-day childhood PA, including studies that measured physical activity level with either accelerometers or questionnaire | School; community | Children, Adolescents and Parents | Educational (healthy eating and PA lessons/counseling; behavior modification); PA (increased time in physical education classes); PA + Educational; 4 weeks to 3 years |
| Spadaccini et al., 2022 | Italy | SR | F and M; 6-19 years; Obesity; other conditions: NR; n = 5510. | 36 RCTs | to systematically review the currently available literature on non-pharmacological and non-surgical inpatient programs aimed at weight loss, to clarify the efficacy and the characteristics of these interventions. | Health clinics | Children, Adolescents and Parents | Dietary intervention + physical Activity intervention + psychotherapy. 5,4-43 weeks. |

| Study | Country | Design | Participants | Included studies (n) | Objective | Setting | Who receive the intervention | Intervention(s) |
|---|---|---|---|---|---|---|---|---|
| Staniford et al., 2011 | United Kingdom | SR | F and M; 5-18y; Overweight/obesity; Other health conditions = NR; n= NR | 61 RCTs and non-RCTs | To provide an update of the evidence, illustrating the efficacy of childhood obesity treatment, considering whether treatment fidelity has been measured and/or reported and whether this related to the treatment effect size | NR | Children, Adolescents and Parents | Others (multiple health behaviors including nutrition, PA and/or sedentary behaviors coupled with behavioral change/modification strategies and incorporated family involvement); 10 to 52 weeks |
| Upton et al., 2014 | United Kingdom | SR | F and M; 4-16y; Overweight/obesity; Other health conditions= NR; n= 1002 | 10 RCTs and non-RCTs | To present the available evidence from United Kingdom family-based childhood obesity interventions | Community | Children, Adolescents and Parents | Others (Family-based intervention for weight-management); 12 weeks to 1 year |
| Verjans-Janssen et al., 2018 | Netherlands | SR | F and M; 4-12y; Normal weight + overweight/obesity; Other health conditions = NR; n=39101 | 25 RCTs, quasi-experimental and pre-post test studies | To study the effectiveness of primary school-based physical activity, sedentary behavior and nutrition interventions with direct parental involvement on children's BMI or BMI z-score, physical activity, sedentary behavior and nutrition behavior and categorize intervention components into targeted socio-cognitive determinants and environmental types using the | School | Children, Adolescents and Parents | Others (changes to the school's policies or physical and social environment or budget for implementing PA and diet activities); 10 weeks to 4 years |

| Study | Country | Design | Participants | Included studies (n) | Objective | Setting | Who receive the intervention | Intervention(s) |
|---|---|---|---|---|---|---|---|---|
| | | | | | Environmental Research framework for weight Gain prevention | | | |

Interventions are classified as follows: PA (physical activity); Diet; Educational; Pharmacological; Others; Combinations (e.g. PA + diet). BMI = Body mass index; HRmax = maximum heart rate; NAFLD = nonalcoholic fatty liver disease; NR = not reported; RM= repetition maximum; $VO_{2peak}$= peak oxygen consumption; $VO_{2max}$= maximal oxygen consumption; Combined exercises= aerobic + resistance.

### eTable 6. Frequency of all unique outcomes and domains.

| Domains | Cumulative outcome frequency, N | Subdomains | Outcomes | Frequency, N | Frequency, % (reviews) |
|---|---|---|---|---|---|
| Adverse Events | 6 | | Adverse events | 4 | 2.9 |
| | | | Morbidity | 2 | 1.4 |
| Blood and Lymphatic System | 118 | lipid profile | High-density lipoproteins (HDL) | 31 | 22.5 |
| | | | Total Cholesterol | 28 | 20.3 |
| | | | Low-density lipoproteins (LDL) | 26 | 18.8 |
| | | | Triglycerides | 25 | 18.1 |
| | | | Non-High-density lipoproteins (non–HDL-C) | 3 | 2.2 |
| | | | Lipid profile | 2 | 1.4 |
| | | | Total cholesterol to HDL ratio | 1 | 0.7 |
| | | | Number of erythrocytes | 1 | 0.7 |
| | | | Plasma malondialdehyde | 1 | 0.7 |
| Cardiac | 94 | blood pressure | Systolic blood pressure (SBP) | 31 | 22.5 |
| | | | Diastolic blood pressure (DBP) | 27 | 19.6 |
| | | | Blood pressure | 8 | 5.8 |
| | | | Mean systolic and diastolic blood pressure | 4 | 2.9 |
| | | | Mean arterial blood pressure | 1 | 0.7 |
| | | | Maximum Heart Rate (HRmax) | 5 | 3.6 |
| | | | Resting and maximal heart rate | 5 | 3.6 |
| | | | Heart rate variability | 4 | 2.9 |
| | | | Heart rate recovery | 2 | 1.4 |
| | | | Mean Respiratory Rate | 2 | 1.4 |
| | | | Sympathetic activity | 2 | 1.4 |
| | | | Cardiac output | 1 | 0.7 |
| | | | Parasympathetic activity | 1 | 0.7 |
| | | | Ventilatory threshold | 1 | 0.7 |
| Cognitive Functioning | 8 | | Cognitive competence (self-perception) | 1 | 0.7 |
| | | | Cognitive flexibility | 1 | 0.7 |
| | | | Food knowledge | 1 | 0.7 |
| | | | Metacognition | 1 | 0.7 |
| | | | Non-executive cognitive function | 1 | 0.7 |
| | | | Physical activity knowledge | 1 | 0.7 |

| Domains | Cumulative outcome frequency, N | Subdomains | Outcomes | Frequency, N | Frequency, % (reviews) |
|---|---|---|---|---|---|
| | | | Selective attention-inhibition | 1 | 0.7 |
| | | | Working memory | 1 | 0.7 |
| Delivery of care | 4 | | Participants' views of the intervention | 1 | 0.7 |
| | | | Peer support | 1 | 0.7 |
| | | | Quality of care | 1 | 0.7 |
| | | | Unhealthy weight control behaviours (self-perception) | 1 | 0.7 |
| Emotional Functioning/ Wellbeing | 37 | | Self-esteem | 9 | 6.5 |
| | | | Self-efficacy | 4 | 2.9 |
| | | | Self-perception | 4 | 2.9 |
| | | | Self-worth (self-perception) | 3 | 2.2 |
| | | | Intrinsic motivation | 2 | 1.4 |
| | | | Mood | 2 | 1.4 |
| | | | PA enjoyment | 2 | 1.4 |
| | | | Alcohol and cigarette use | 1 | 0.7 |
| | | | Attitudes about appearance | 1 | 0.7 |
| | | | Body image (self-perception) | 1 | 0.7 |
| | | | Media internalization (self-perception) | 1 | 0.7 |
| | | | Parents' alcohol use | 1 | 0.7 |
| | | | Physical self-perception or body satisfaction | 1 | 0.7 |
| | | | Positive expectations | 1 | 0.7 |
| | | | Psychological attraction to exergame | 1 | 0.7 |
| | | | Psychological well-being | 1 | 0.7 |
| | | | Self-acceptance (self-perception) | 1 | 0.7 |
| | | | Social-emotional development | 1 | 0.7 |
| Endocrine | 117 | glucose metabolism | Fasting blood glucose | 13 | 9.4 |
| | | | Fasting insulin | 9 | 6.5 |
| | | | Glucose | 8 | 5.8 |
| | | | Homeostatic Model Assessment-Insulin Resistance (HOMA-IR) | 7 | 5.1 |
| | | | Insulin resistance | 7 | 5.1 |
| | | | Insulin levels | 4 | 2.9 |
| | | | Glycated Haemoglobin (HbA1c) | 3 | 2.2 |
| | | | Insulin sensitivity | 2 | 1.4 |

| Domains | Cumulative outcome frequency, N | Subdomains | Outcomes | Frequency, N | Frequency, % (reviews) |
|---|---|---|---|---|---|
| | | | Insulin-like Growth Factor (IGF-1) | 2 | 1.4 |
| | | | Blood insulin | 1 | 0.7 |
| | | | Insulin-like Growth Factor Binding Protein-3 (IGFBP-3) | 1 | 0.7 |
| | | | Non-fasting glucose | 1 | 0.7 |
| | | | Non-fasting insulin | 1 | 0.7 |
| | | | Serum insulin | 1 | 0.7 |
| | | | Adiponectin | 9 | 6.5 |
| | | | Leptin levels | 9 | 6.5 |
| | | | C-reactive protein | 6 | 4.3 |
| | | | Interleukin-6 (IL-6) | 6 | 4.3 |
| | | | Tumor Necrosis Factor (TNF-α) | 5 | 3.6 |
| | | | Resistin levels | 3 | 2.2 |
| | | | Adipocyte fat (aFABP) | 1 | 0.7 |
| | | | Alanine aminotransferase (ALT) | 1 | 0.7 |
| | | | Apolipoprotein A-1 | 1 | 0.7 |
| | | | Apolipoprotein B | 1 | 0.7 |
| | | | Aspartate aminotransferase (AST) | 1 | 0.7 |
| | | | Brain-derived neurotrophic factor (BDNF) | 1 | 0.7 |
| | | | Chemerin | 1 | 0.7 |
| | | | Fibrinogen level | 1 | 0.7 |
| | | | Intrahepatic fat | 1 | 07 |
| | | | Irisin | 1 | 0.7 |
| | | | Lactate threshold | 1 | 0.7 |
| | | | Metabolic syndrome | 1 | 0.7 |
| | | | Myostatin | 1 | 0.7 |
| | | | Plasminogen activator inhibitor-1 | 1 | 0.7 |
| | | | Proinflammatory cytokines | 1 | 0.7 |
| | | | Serum AgRP, NPY and α-MSH | 1 | 0.7 |
| | | | Serum interleukin-15 | 1 | 0.7 |
| | | | Superoxide dismutase | 1 | 0.7 |
| | | | Visfatin levels | 1 | 0.7 |
| General | 255 | body composition | Body Mass Index (BMI) | 72 | 52.2 |
| | | | Body weight | 39 | 28.3 |

| Domains | Cumulative outcome frequency, N | Subdomains | Outcomes | Frequency, N | Frequency, % (reviews) |
|---|---|---|---|---|---|
| | | | Body fat | 34 | 24.6 |
| | | | Body Fat-free mass | 30 | 21.7 |
| | | | Skinfold thickness | 16 | 11.6 |
| | | | Waist Circumference | 16 | 11.6 |
| | | | Regional fat | 13 | 9.4 |
| | | | Body mass | 8 | 5.8 |
| | | | Height | 6 | 4.3 |
| | | | Obesity/Overweight prevalence | 6 | 4.3 |
| | | | Waist-to-hip ratio | 5 | 3.6 |
| | | | Hip circumference | 3 | 2.2 |
| | | | Subcutaneous adipose tissue | 3 | 2.2 |
| | | | Parents' BMI | 2 | 1.4 |
| | | | Body circumferences | 1 | 0.7 |
| | | | Waist-to-height ratio | 1 | 0.7 |
| Global Quality of Life | 13 | | Quality of Life | 7 | 5.1 |
| | | | Health-related Quality of Life | 6 | 4.3 |
| Metabolism and Nutrition | 28 | food intake | Food intake | 9 | 6.5 |
| | | | Fruit and vegetable intake | 2 | 1.4 |
| | | | Portions of fruit, vegetables, and water | 2 | 1.4 |
| | | | Dietary behavior | 9 | 6.5 |
| | | | Eating disorders | 3 | 2.2 |
| | | | Food desire | 1 | 0.7 |
| | | | Gamma-glutamyl transferase (GGT) | 1 | 0.7 |
| | | | Self-report of food servings | 1 | 0.7 |
| Musculoskeletal and Connective Tissue | 4 | | Total bone density | 2 | 1.4 |
| | | | Bone mineral content | 1 | 0.7 |
| | | | Regional bone density | 1 | 0.7 |
| Perceived Health Status | 5 | | Perceived Athletic competence | 2 | 1.4 |
| | | | Perceived Physical condition | 2 | 1.4 |
| | | | Healthy attitudes (self-perception) | 1 | 0.7 |
| Physical Functioning | 128 | | Time spent in physical activity | 17 | 12.3 |
| | | | Muscle strength | 14 | 10.1 |
| | | | Physical activity level | 12 | 8.7 |
| | | | Sedentary behavior | 12 | 8.7 |

| Domains | Cumulative outcome frequency, N | Subdomains | Outcomes | Frequency, N | Frequency, % (reviews) |
|---|---|---|---|---|---|
| | | | Cardiorespiratory Fitness | 11 | 8.0 |
| | | | Maximal Oxygen Uptake (VO2max) | 8 | 5.8 |
| | | | Peak Oxygen Uptake (VO2peak) | 7 | 5.1 |
| | | | Agility | 6 | 4.3 |
| | | | Healthy lifestyle | 5 | 3.6 |
| | | | Flexibility | 4 | 2.9 |
| | | | Aerobic fitness | 3 | 2.2 |
| | | | Balance | 3 | 2.2 |
| | | | Jump ability | 3 | 2.2 |
| | | | Peak muscle power | 3 | 2.2 |
| | | | Energy expenditure in physical activity | 2 | 1.4 |
| | | | Maximal power output | 2 | 1.4 |
| | | | Muscle endurance | 2 | 1.4 |
| | | | Muscle Power | 2 | 1.4 |
| | | | Physical activity behavior | 2 | 1.4 |
| | | | Physical performance | 2 | 1.4 |
| | | | Explosive power | 1 | 0.7 |
| | | | Fatigue | 1 | 0.7 |
| | | | Maximum speed | 1 | 0.7 |
| | | | Mean power | 1 | 0.7 |
| | | | Motor competence | 1 | 0.7 |
| | | | Physical activity playing active and non-active videogames | 1 | 0.7 |
| | | | Running economy | 1 | 0.7 |
| | | | Sprint performance | 1 | 0.7 |
| Psychiatriac | 11 | | Depression | 7 | 5.1 |
| | | | Anxiety | 4 | 2.9 |
| Social Functioning | 10 | | Relationship with parents | 3 | 2,2 |
| | | | Social acceptance | 2 | 1.4 |
| | | | Aggression (self-perception) | 1 | 0.7 |
| | | | Parent acceptance | 1 | 0.7 |
| | | | Parenting skill and relationships | 1 | 0.7 |
| | | | Social stress | 1 | 0.7 |
| | | | Strength and difficulties perceived by parents | 1 | 0.7 |

| Domains | Cumulative outcome frequency, N | Subdomains | Outcomes | Frequency, N | Frequency, % (reviews) |
|---|---|---|---|---|---|
| Vascular | 7 | | Flow-mediated dilation (FMD) | 2 | 1.4 |
| | | | Arterial stiffness | 1 | 0.7 |
| | | | Endothelial dysfunction | 1 | 0.7 |
| | | | Intima-media thickness (IMT) | 1 | 0.7 |
| | | | Vascular conductance | 1 | 0.7 |
| | | | Vascular resistance | 1 | 0.7 |
